# Consensus meta-analysis of genome-wide association studies for Alzheimer’s disease and related dementia

**DOI:** 10.1101/2025.10.20.25338060

**Authors:** ADGC, Bonn, CHARGE, EADB, Celine Bellenguez, EADI, FinnGen, GERAD, GR@ACE/DEGESCO, PGC-ALZ

## Abstract

To better characterize the genetic architecture underlying Alzheimer’s disease (AD) and related dementia (ADRD), we performed a meta-analysis of European ancestry genome-wide association studies in 128,681 cases or proxy cases of ADRD and 849,833 (proxy) controls. We identified 91 genetic loci associated with ADRD risk, including 16 which are novel, and 56 which are specifically detected in clinically diagnosed AD cases. We also provide a list of 18 loci (15 novel) requiring further external validation. A polygenic score combining the effect of the ADRD loci except *APOE* was primarily associated with AD rather than non-AD pathology. Individuals in the 10^th^ decile of the score had a 2-fold increased risk to present with Braak neurofibrillary tangles stage above 5 and moderate/severe neuritic amyloid plaque pathology at death compared to individuals in the median score group.

## Main text

More than 90 genetic loci are associated with ADRD. Most of them were identified through large-scale genome-wide association studies (GWAS) performed in European ancestry samples by, among others, the International Genomics of Alzheimer’s Project (IGAP), the European Alzheimer & Dementia Biobank (EADB), and the Psychiatric Genomics Consortium – AD Working Group (PGC-ALZ)^1–7^. Although the latest GWASs on ADRD partly overlapped, they used different imputation panels or analytical approaches, and some results were discordant across studies. They all included, but to different extents, large biobank samples using ICD codes to identify Alzheimer’s Disease (AD) cases, proxy ADRD cases (i.e. individuals reporting at least one parent or sibling with dementia) or both. This strategy increases power to identify AD loci, but it can also blur the distinction between AD and non-AD dementia signals as the AD phenotype definition is less specific in those samples. To better characterize the genetic architecture and pathophysiology underlying AD and ADRD, the three consortia joined their efforts to perform a consensus meta-analysis of ADRD GWAS across all their samples of European ancestry, the UK Biobank (UKBB), and FinnGen. To further delineate the impact of known genomic loci on AD vs ADRD, sensitivity analyses were performed by excluding proxy or large biobank samples.

The meta-analysis included 72,721 AD cases, 55,960 proxy ADRD cases, 614,267 controls and 235,566 proxy controls from 52 studies, corresponding to a rough effective sample size of 230,631 (Supplementary Information, Supplementary Table 1). Following quality control, associations were considered for 20,045,120 variants (Supplementary Figures 1 and 2). We identified 91 genome-wide significant loci (P ≤ 5×10^-8^, defined as “Tier 1”); sixteen of the loci – *EIF4G3, PTPRC, MGAT5, PPP2R3A, ADGRL3, FAM193B, TMEM184A, DOCK4, IPMK, UBFD1, VMAC, VAV1, LRRC25, CEP89, LILRB1/LILRB4* and *SRC* - were novel in European ancestry samples at the time of analysis although *ADGRL3* was recently identified in a trans-ethnic GWAS^8^ (Figure 1, Supplementary Tables 2 and 3, Supplementary Figures 3-55). Following a stepwise conditional analysis, we identified 25 independent secondary signals in 16 loci (Figure 1, Supplementary Table 3, Supplementary Figures 3-55). This is the first time an independent association signal has been detected in the *CD33*, *HLA*, *PICALM* and *RHOH* loci. We also detected additional independent signals in *ABCA7*, *BIN1*, *PTK2B/CLU*, *NCK2* and *PLCG2*^4–6,9^ (Figure 1). While some main and secondary signals are likely linked to the same gene, e.g. the low-frequency variants in *SORL1* and *ABCA7*, some may be linked to two different genes in the same locus. Of note, in addition to the 91 Tier 1 loci, we detected 5 loci where the significance dropped after conditional analyses (P > 1×10 ^-7^) within or across loci, suggesting a slight inflation of the unconditional analysis (Figure 1, Supplementary Tables 2, 3 and 4). Those loci require external validation and were thus classified as “Tier 2”. Three were known (*SEC61G/EGFR*, *SPPL2A/USP8/USP50*, *KAT8/BCKDK*) and two were novel in European ancestry samples (*TRIB1* and *AXIN1)*, although *TRIB1* was identified in a recent trans-ethnic GWAS^8^. Finally, four loci identified in the two previous largest ADRD GWAS meta-analyses on European ancestry samples^5,6^ were not genome-wide significant in the EADB-IGAP-PGC meta-analysis: *HAVCR2*, *SLC2A4RG/LIME1*, *FOXF1* and *NTN5* (Supplementary Table 5, Supplementary Figure 56).

**Figure 1:**
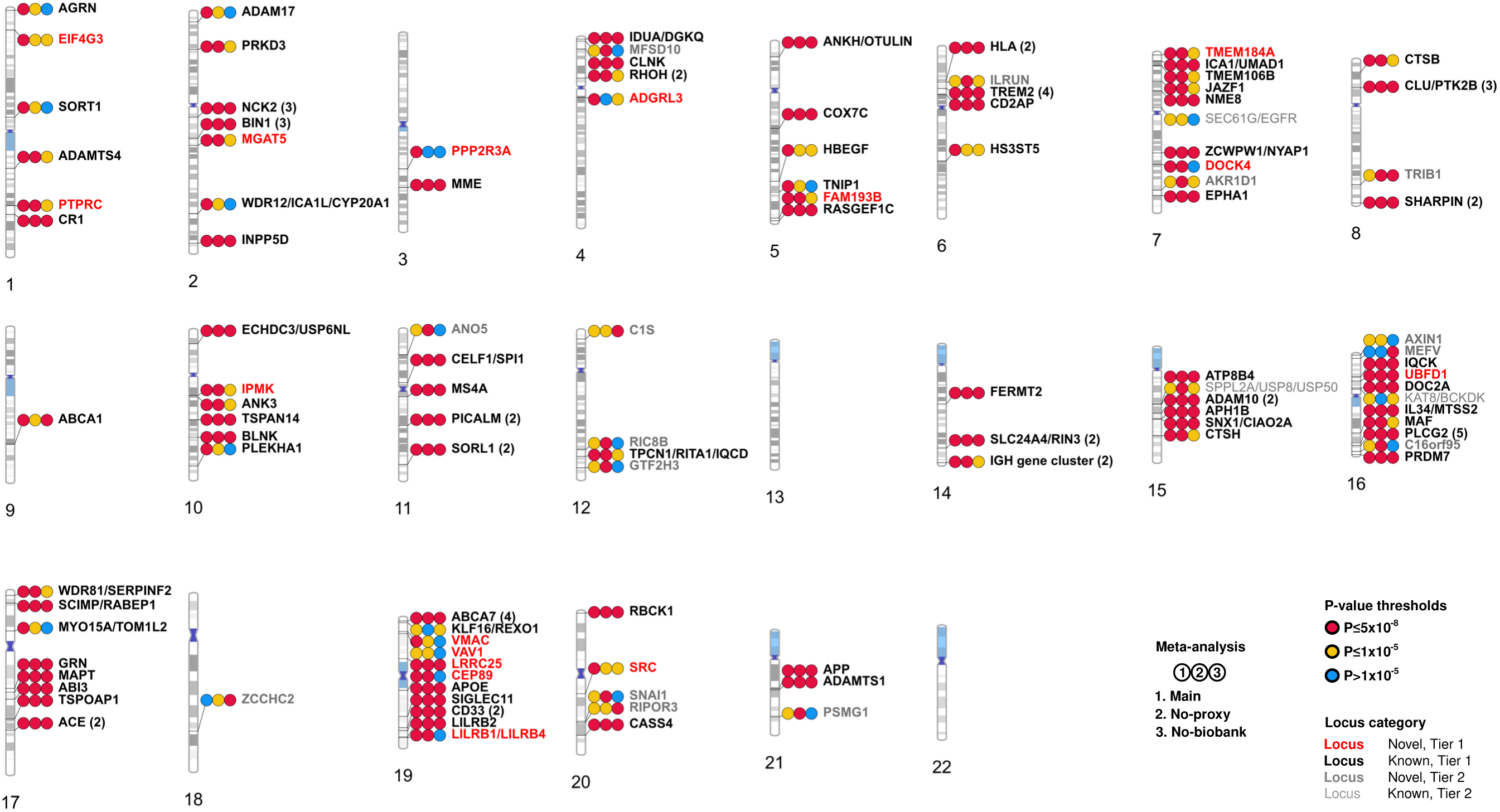
Ideogram of the Tier 1 and 2 loci. Tier 1 loci are considered genuine genome-wide significant signals, with P ≤ 5×10^-8^ in the unconditional analysis and P ≤ 1×10^-7^ in conditional analyses within or across loci. Tier 2 loci are genome-wide significant signals requiring further external validation because i) P was above 1×10^-7^ after conditional analyses or ii) they were genome-wide significant only in the sensitivity no-proxy and no-biobank meta-analyses. For each locus, the figure shows the P-value categories for the association with ADRD or AD risk in the main, no-proxy and no-biobank meta-analyses: P≤5×10^-8^, P≤1×10^-5^, P>1×10^-5^ (two-sided raw P-values). We considered the P-value of the lead variant when the locus was detected in the meta-analysis and otherwise the minimum P-value across all index variants of the locus. Novel Tier 1 loci are in bold red, known Tier 1 loci in bold black, known Tier 2 loci in grey, and novel Tier 2 loci in bold grey. Numbers in parentheses refer to the number of genome-wide significant independent signals in the locus according to the main meta-analysis.

We assessed the robustness of the signals according to the AD diagnosis quality by excluding proxy ADRD samples or large biobank samples from the meta-analysis. Overall, the genetic correlation among the three AD phenotypes thus defined was above 0.97 (Supplementary Table 6). Among the 91 Tier 1 loci, 75 (82.4%) were genome-wide significant in the no-proxy meta-analysis, and 56 (61.5%) in the no-biobank meta-analysis focusing on clinically diagnosed AD cases (Figure 1, Supplementary Tables 2 and 7). Ten of the 75 loci detected in both the main and no-proxy meta-analyses were novel: *PTPRC*, *MGAT5*, *FAM193B*, *TMEM184A*, *DOCK4*, *IPMK*, *UBFD1*, *LRRC25*, *CEP89* and *LILRB1/LILRB4*. The *UBFD1* and *LRRC25* novel loci were also genome-wide significant in the no-biobank meta-analysis, while 12 known loci were genome-wide significant for the first time after focusing on clinically diagnosed AD cases (*NCK2, MME, ANKH/OTULIN, ABCA1, BLNK, IGH, ATP8B4, SNX1/CIAO2A, IL34/MTSS2, SIGLEC11, RBCK1* and *APP*). We additionally checked for putative signals only detected in the sensitivity meta-analyses: nine loci were only identified in the no-proxy meta-analysis, while four loci were only identified in the no-biobank meta-analysis (Figure 1, Supplementary Tables 2 and 7, Supplementary Figures 57-61). All these 13 loci were novel and classified as “Tier 2”. The Supplementary Information further describes the GWAS results and provides putative secondary signals at a more lenient significance threshold of 1×10^-5^ (Supplementary Table 3).

Among the index variants of the main or secondary Tier 1 signals, three were missense variants with a REVEL score above 0.25 (in the *MME*, *TREM2* and *ABCA7* loci); only seven index variants were rare, with minor allele frequency or MAF below 1%, in the *SORT1*, *NCK2*, *ADGRL3*, *TREM2*, *PLCG2* and *ABCA7* loci (Supplementary Tables 2 and 3). We further assessed the impact on AD risk of rare variants within the novel Tier 1 loci using a previous gene-based analysis of our samples’ sequencing data. In the *SRC* gene, loss-of-function variants and missense variants with a REVEL score above 50 were jointly associated with AD risk (OR=4.23 (2.04-8.79), P=1.06×10^-4^, Supplementary Table 8) in the ADES-ADSP summary statistics^10^.

As in previous GWASs, genes enriched in ADRD or AD association signal in the main, no-proxy, and no-biobank meta-analyses were over-expressed in microglia in four different datasets spanning different brain regions (Supplementary Information, Supplementary Tables 9 and 10, Supplementary Figures 62-64). Similarly to previous GWAS results, association signals were significantly enriched in pathways related to Tau, amyloid, lipids, immunity or endosome/lysosome, in the main, no-proxy, and no-biobank meta-analyses (Supplementary Table 11). By performing a phenome-wide association study, we also linked some of the new loci (both Tiers) to pathways related to tau, lipids or immunity (Supplementary Tables 12, 13 and 14). For example, the *C16orf95* locus is associated with phosphorylated tau (pTau) levels in cerebrospinal fluid (CSF), and also with ventricular volume (Supplementary Information) ^11,12^. This latter observation supports the validity of some of the Tier 2 loci. After exclusion of *APOE*, the no-proxy ADRD phenotype was significantly (P≤3.13×10^-3^) genetically correlated with Lewy body dementia (r=0.64, 95% confidence interval (CI) [0.37, 0.92]), amyotrophic lateral sclerosis (r=0.29, 95% CI [0.18, 0.40]), Parkinson’s disease (r=0.16, 95% CI [0.06, 0.27]) and educational attainment (r=-0.12, 95% CI [-0.17, –0.07]) (Supplementary Table 6). Those results, consistent with previous studies^13,14^, were similar when considering the main and no-biobank summary statistics except for educational attainment when using the main meta-analysis results (r=-0.02, 95% CI [-0.07, 0.02]). This is in accordance with previous reports of biases observed with genome-wide summary statistics of studies including proxy cases^14–16^.

Genetic correlation with ADRD could not be assessed reliably for most of the 11 neuropathological endophenotypes (NPE) we considered due to the low study samples sizes or heritability of those traits (Supplementary Table 6). Those NPEs comprised i) three AD-related NPEs: Braak neurofibrillary tangles (NFT) stage, amyloid-β plaques and CERAD score for neuritic amyloid plaques; ii) five cerebrovascular NPEs: arteriolosclerosis, circle of Willis atherosclerosis, cerebral amyloid angiopathy (CAA), gross infarcts and microinfarcts and iii) three non-AD NPEs: limbic-predominant age-related TDP-43 encephalopathy neuropathological change (LATE-NC), Lewy bodies and hippocampal sclerosis. To better study the genetic link between ADRD and NPEs, we constructed three polygenic scores (PGS) based on the Tier 1 main and secondary signals (except *APOE*) detected in the main, no-proxy and no-biobank meta-analyses respectively (Supplementary Table 15). We then assessed their association with the 11 NPEs in the Adult Changes in Thought (ACT) cohort (N=677 including 12.9% with dementia) and in the Alzheimer Disease Centers /National Alzheimer’s Coordinating Center (ADC/NACC) dataset (N=5,808 including 82.7% with dementia) (Supplementary Table 16)^17^. The main score was significantly associated (P≤2.27×10^-3^) in ADC/NACC with the three AD-related NPEs and with LATE-NC, and the signals were in the same direction in the smaller ACT study (Figure 2a, Supplementary Table 17, Supplementary Information). None of the scores was significantly associated with the cerebrovascular NPEs, nor with hippocampal sclerosis or Lewy body (Figure 2a, Supplementary Table 17) while the genetic correlation between ADRD and Lewy body dementia was significant after exclusion of *APOE*; this may indicate common pathological pathways underlying conversion from the pathology to AD or Lewy body dementia, although we can not exclude other explanations, such as a lack of statistical power of the PGS analysis or misdiagnoses among AD and Lewy body dementia cases impacting genetic correlation. Results were similar across the main, no-proxy and no-biobank PGS (Supplementary Table 17). After adjustment on the (other) AD NPEs, only the associations with Braak stage and CERAD score remained significant in ADC/NACC (Figure 2b, Supplementary Table 17). The association with LATE-NC remained significant at the nominal level only (P=3.78×10^-2^, Supplementary Table 17); a larger sample size is required to assess if the association with LATE-NC is due or not to the frequent co-occurrence of LATE-NC and AD NPEs (Supplementary Information). There was no significant interaction (P≤2.27×10^-3^) between the PGS and the number of APOE ε4 and ε2 alleles for Braak stage and CERAD score (Supplementary Table 17). In the ADC/NACC dataset, compared to the individuals with a main PGS in the median quintile (40 to 60%), individuals in the 10^th^ decile had a risk increased by 2.05-fold (95% CI=[1.47-2.85]) and 1.96-fold (95% CI=[1.39-2.78]) for Braak stage above 5 and moderate/severe neuritic amyloid plaque pathology at death respectively, while individuals in the 1^st^ decile had a risk decreased by 0.47-fold (95% CI=[0.37-0.61]) and 0.43-fold (95% CI=[0.33-0.56]) respectively (Figure 3, Supplementary Tables 18 and 19, Supplementary Figure 67 and Supplementary Information). Compared to a model considering only age at death, sex and the number of APOE ε4 and ε2 alleles, the main PGS allowed a significant improvement (P≤2.27×10^-3^) in discrimination measured by the area under the Receiver Operating Characteristic (ROC) curve (AUC) for Braak stage and CERAD score in the ADC/NACC dataset (Supplementary Table 20). However, the variance explained by the score was low: Nagelkerke’s pseudo R-squared was 3.98% and 4.37% for Braak stage and CERAD score respectively, while variance explained on the liability scale varied between 2.43% and 3.6% for Braak stage, and between 3.32% and 4.93% for CERAD score, depending on the population prevalence considered. The discriminative power as measured by AUC was similar for the main, no-proxy and no-biobank PGS (P > 0.05, Supplementary Table 21).

**Figure 2:**
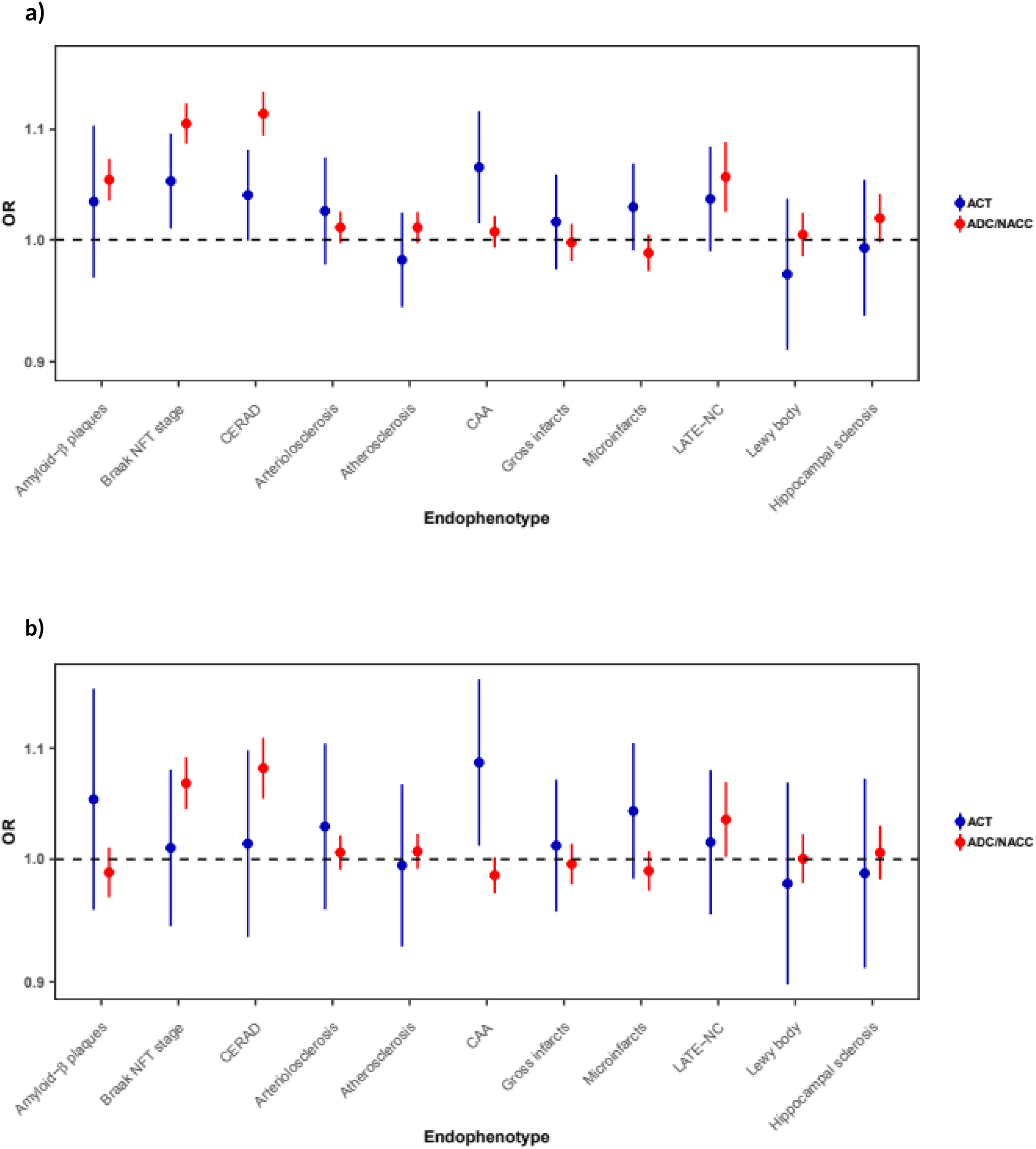
Association of the main ADRD polygenic score (PGS) with 11 neuropathology endophenotypes (NPE) in the ACT and ADC/NACC datasets. After a) minimal adjustment on age at death, sex, the number of APOE ε4 and ε2 alleles, principal components and centers and b) additional adjustment on (other) AD NPEs. OR: odds-ratio; NFT: neurofibrillary tangles; LATE-NC: limbic-predominant age-related TDP-43 encephalopathy neuropathological change. Bars indicate 95% confidence intervals.

**Figure 3:**
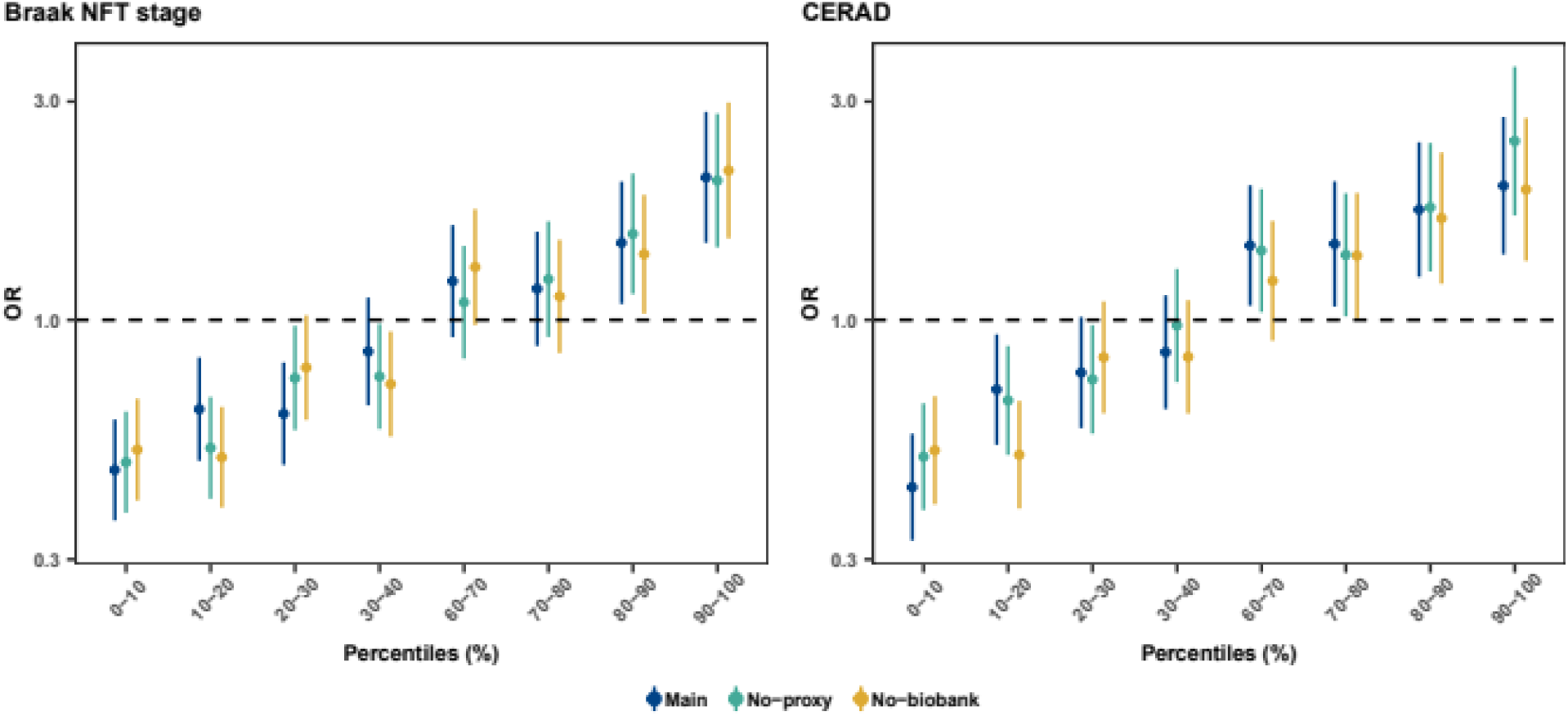
Association of the main, no-proxy and no-biobank ADRD polygenic score (PGS) deciles with Braak stage and CERAD score in the ADC/NACC dataset. The reference is the median (40%-60%) quintile. OR: odds-ratio. Bars indicate 95% confidence intervals.

In summary, this consensus meta-analysis identified 91 genetic loci associated with ADRD risk, including 16 novel loci in European ancestry samples. 56 of the loci were associated with the risk of clinically diagnosed AD. We also further characterized the impact of known loci by validating – or not – their association with ADRD and AD risk in larger samples and by identifying new secondary signals in some of them. Except for genetic correlation with education, our results were consistent across the main, no-proxy and no-biobank meta-analyses and the three PGS excluding *APOE* were primarily associated with AD rather than non-AD pathology. However, assessing the sensitivity of results of GWAS or GWAS secondary analyses (especially those based on genome-wide statistics^14^) on clinically-diagnosed cases will become increasingly important as the proportion of proxy and biobank cases included in the GWAS increases. This will be made easier by the release of the no-proxy and no-biobank summary statistics. Additionally, some ADRD loci are significantly associated with non-AD NPEs but not with AD NPEs (Supplementary Table 14)^17^. Such results are difficult to interpret though considering the low power of NPE GWASs for variants with a small effect on ADRD risk; studies using further well-characterized AD patients and neuropathological data will thus be required to better delineate the impact of each of the loci on AD pathology versus other neuropathologies. Follow-up analyses of rare damaging variants and structural variants using sequencing data and functional studies will provide further insights on the biological impact of the loci on ADRD and AD.

## Online Methods

### Samples

We analysed genotyping data from European-ancestry samples from 52 studies: 46 case-control or cohort studies, 2 family studies (NIA-LOAD and FHS) and 4 large biobanks (the UKBB, FinnGen, deCODE and HUNT). The samples are described in Supplementary Table 1 and in the Supplementary Information. In the UKBB, proxy ADRD cases included participants who reported at least one biological relative (parents and siblings) affected with dementia, either at baseline or follow up (Supplementary Information). Eleven NPEs were measured in autopsied individuals from the ACT and ADC/NACC studies (Supplementary Table 16 and Supplementary Information). The NPE definitions and harmonization approach are described in Shade et al^17^. Written informed consent was obtained from study participants or, for those with substantial cognitive impairment, from a caregiver, legal guardian, or other proxy. The appropriate review boards from the ADGC, Bonn, CHARGE, EADB, EADI, GERAD, GR@ACE/DEGESCO and PGC-ALZ reviewed and approved the study protocol. The research included researchers from each participating consortium throughout the research process.

### Quality control and imputation

Classical quality control protocols were applied to samples and autosomal variants of each study (Supplementary Information). Most of the samples were imputed with the TOPMed reference panel^18,19^; one study was imputed with the Haplotype Reference Consortium (HRC) panel^20^ while the UKBB, FinnGen and deCODE biobanks were imputed with study-specific panels (Supplementary Table 22).

### Genome-wide association studies and meta-analyses

Association of each autosomal variant with ADRD risk was tested in each study under an additive genetic model. Logistic regression was used in most studies. When necessary, relatedness was taken into account with generalized estimating equation (GEE) or logistic mixed models (Supplementary Information). Analyses were adjusted on principal components (PCs) and center/batches. In a few studies, adjustment was also performed on sex, age or both (Supplementary Table 22). In deCODE, correction for inflation of test statistics due to relatedness and population stratification was performed using the intercept estimate (1.30) from LD score regression^21^. In the UKBB-proxy analysis, effect sizes and standard errors were corrected by a factor of two^22,23^. For all studies, we filtered out duplicated variants and variants with (i) missing data on the effect size, standard error, or P*-*value, (ii) an absolute effect size above 5, (iii) an imputation quality below 0.3 (0.8 for the GenADA study). For deCODE and UKBB, data were analysed in the GRCh37 assembly, and we excluded variants for which conversion of position or alleles from the GRCh37 assembly to the GRCh38 assembly was not possible or problematic^6^. For the UKBB and the EADB core HRC study, variants with very large difference of frequency between the TOPMed reference panel and the reference panels used to perform imputation were also excluded^6^.

### Meta-analyses

Results were combined across studies with a fixed-effect meta-analysis with an inverse variance weighted approach, as implemented in METAL v2020-05-05 software^24^. In the main meta-analysis, all studies were considered, and we filtered in each study variants with an effective allele count (product of the imputation quality and the expected minimum minor allele count between the cases and the controls) less than 5^25^. After meta-analysis, we filtered out (i) variants with frequency amplitude above 0.4 (defined as the difference between the maximum and minimum frequencies across all the studies) and (ii) variants analyzed in less than 40% of the effective number of cases. In each study, the effective number of cases was defined as the raw number of cases, except in the UKBB-proxy, where it was computed by dividing the raw number of proxy cases by four^22,26^. Several sensitivity meta-analyses were conducted by:

1. excluding the UKBB-proxy study (no-proxy meta-analysis). The UKBB study was included in the meta-analysis, but only diagnosed cases were considered (Supplementary Information, Supplementary Table 1);
2. excluding the UKBB, FinnGen, deCODE and HUNT studies, corresponding to large biobanks using ICD codes to identify AD cases (no-biobank meta-analysis, Supplementary Table 1);
3. removing the per-study variant filtering on effective allele count, except in the FHS study (Supplementary Information, Supplementary Table 23).

Additionally, we assessed sensitivity of the results at the genome-wide significant lead variants in the main meta-analysis after adjustment on age and sex, and age, sex and number of *APOE*-ε4 and *APOE*-ε2 alleles (Supplementary Table 24).

### Loci definition

The loci were defined using an approach similar to the one described in Hautakangas et al.^27^. We first selected variants with a genome-wide significant (GWS) signal, and also suggestive variants (P≤1×10^-5^) located at +/- 500kb from a GWS variant, and that were analysed in at least 70% of the effective number of cases. Linkage disequilibrium (LD) between the variants was computed in the EADB-core dataset using genotype dosages with LDstore 2^28^. For variants not available in EADB-core, LD was computed in 1000 Genomes EUR samples (version 3) using emeraLD^29^ which took phase into account. Variants not available in EADB-core nor 1000 Genomes were considered as having no LD with other variants. In each chromosome, GWS variants were ordered in a candidate list according to their P-value (in ascending order) and then to their absolute effect size (in descending order). The first variant is considered as an index variant. All variants with LD (according to r^2^) >= 0.1 with this variant are excluded from the candidate list. The process is then repeated for the second variant in the updated candidate list until no remaining candidate variants remained. We then defined a clump around each index variant, made up of all suggestive variants (including GWS variants) with an LD >= 0.6 with the clump’s index variant. Each clump was assigned start and end positions according to the extreme positions of the variants in the clump, and clumps located less than 250kb apart were merged with bedtools v2.31.1 (https://bedtools.readthedocs.io). Each remaining GWS or suggestive variant was then assigned to the closest clump. Clumps’ start and end positions were updated again, and clumps located less than 250kb apart were merged with bedtools. The clumps at this final step define the loci.

In each locus, the variant with the lowest P-value (and the highest absolute effect size in case of equal P-values) was defined as the lead variant. None of the variants with missing LD in both EADB-core and 1000G was selected as a lead variant.

A locus was considered known if a variant previously associated with AD at the GWS level according to the GWAS catalog version e112_r2024-07-08 was located in this locus^30^. For that, we restricted the GWAS catalog to “MAPPED_TRAIT” equals “late-onset Alzheimers disease”, “Alzheimer disease”, “Alzheimer disease, family history of Alzheimer’s disease”, “family history of Alzheimer’s disease” or “Alzheimer disease, dementia, family history of Alzheimer’s disease”.

A gene was assigned to each lead variant: the protein-coding gene where the lead variant is located, and otherwise, the nearest protein-coding gene according to VEP release 109^31^ and considering only transcripts with the GENCODE basic tag. The locus was named according to the gene assigned to its lead variant and to the locus name in the literature for known loci.

The ideogram was generated with PhenoGram (https://ritchielab.org/software/phenogram-downloads).

### Conditional and joint analyses

To identify secondary signals independent of the lead variant signal in the loci, a stepwise conditional analysis was performed in each locus, except *APOE*, with GCTA COJO^32,33^ based on the summary statistics of the main meta-analysis, and on the LD computed in the EADB-core samples. For that, EADB-core genetic data were converted to best-guess genotype data, using a threshold on genotype probabilities of 0.8. Only variants analysed in at least 70% of the effective number of cases were considered, leading to the exclusion of the *ADGRL3* locus from those analyses. The P-value threshold for defining secondary signals was set to 1×10^-5^. Then, to check the independence of the signals across the loci, a joint analysis was performed with GCTA COJO of i) the 157 index variants of the lead and secondary signals detected by the stepwise conditional analysis (Supplementary Information) and ii) the *ADGRL3* lead variant. Those conditional and joint analyses were also performed for the no-proxy and no-biobank sensitivity meta-analyses. To assess the sensitivity of the results of the approximate stepwise conditional analysis to LD and imputation quality, we performed i) a strict stepwise conditional analysis and ii) an exact conditional analysis. The strict stepwise conditional analysis was performed on variants with imputation quality greater than 0.8 in the EADB-core dataset and analysed in at least 90% of the effective number of cases. The exact conditional analyses were performed with SNPTEST^34,35^ on the raw data of a subset of the studies: i) between the index variants of the lead and secondary signals within the same locus; ii) between the index variants of the main and secondary signals of the *KAT8/BCKDK* and *DOC2A* loci on the one hand and the *APH1B* and *SNX1/CIAO2A* loci on the other hand (Supplementary Information). The following studies were considered: EADB-core, Bonn, DemGene, EADI, GERAD, Gothenburg, STSA, TwinGene, and all the case-control ADGC studies. Exact conditional results were combined across studies with an inverse variance weighted approach, as implemented in METAL.

To compare the signals across the main, no-proxy and no-biobank meta-analyses, we performed two joint analyses with GCTA COJO. We jointly analysed all the index variants of the lead and secondary signals of i) the main and no-biobank meta-analyses on the one hand, and ii) of the main and no-proxy meta-analyses on the other hand. We only considered loci with a secondary signal in at least one of the two meta-analyses being compared, but all variants were tested jointly across those loci. Both joint analyses were performed on the summary statistics of the main meta-analysis. From each joint analysis, we excluded one index variant from each pair in LD (r^2^ > 0.75 in the EADB-core dataset, as computed by PLINK 2.0) to avoid collinearity issues. In each such pair, the index variant from the main meta-analysis was kept.

### Rare variant analysis

We extracted 65 protein-coding genes with the Gencode basic tag (version 43) located within the 16 novel Tier 1 loci, using the start and end position of the loci. 51 genes were available in the ADES-ADSP summary statistics of the comparison of the gene-based rare-variant burdens between 12,652 AD cases and 8,693 controls^10^ (Supplementary Table 8). Those samples largely overlap with the ones included in the meta-analysis. We considered a significance threshold of 9.8×10^-4^, corresponding to a Bonferroni correction for 51 tests.

### Single-cell enrichment analysis

We assessed the association between gene overexpression in specific cell types, compared to the average gene expression, and gene association with ADRD risk using the three-step process implemented in FUMA v1.6.1^36^. As input, we used the MAGMA gene level results provided as input for the pathway analysis. We tested 6 models (1 primary model and 5 sensitivity models), corresponding to the primary, common-only, no-apoe, larger window, no-proxy and no-biobank gene-level summary statistics. We used gene expression data from 6 datasets of adult human brain tissue: GSE168408_Human_Prefrontal_Cortex_level2_Adult^37^, GSE168408_Human_Prefrontal_Cortex_level1_Adult^37^, PsychENCODE_Adult^38^, DroNc_Human_Hippocampus^39^, and Allen_Human_MTG_level1 (Middle temporal gyrus) and Allen_Human_MTG_level2^40^. The FUMA three-step process is further described in the Supplementary Information.

### Pathway analyses

Pathway analyses were performed in MAGMA v1.08^41,42^, with correction for the number of variants in each gene, LD between variants and LD between genes. LD was computed from the EADB-core dataset using high-quality imputed genotypes (imputation quality above 0.8) and setting as missing genotypes with genotype probability below 0.9. The measure of pathway enrichment was the MAGMA ‘competitive’ test (in which the association statistic for genes in the pathway is compared with those for all other protein-coding genes), as recommended by De Leeuw et al.^43^. We used the ‘mean’ test statistic, which uses the sum of −log(variant P-value) across all genes. Total sample size (N) was used as sample size. The assignments of Gene Ontology (GO) terms to human genes were obtained from the “gene2go” file (downloaded from the NCBI on June 19th, 2023). “Parent” GO terms were assigned to genes by propagating annotations of “Children” terms, using the GeneOntology .obo (go-basic.obo) file downloaded on the same date from GeneOntology and the ontologyIndex R package^44^. GO terms were assigned to genes on the basis of experimental or curated evidence of a specific type, and so we excluded evidence codes IEA (electronic annotation), NAS (non-traceable author statement), and RCA (inferred from reviewed computational analysis). Pathways were downloaded from the Reactome website on July 17th, 2023. Biocarta, KEGG and Pathway Interaction Database (PID) pathways were downloaded on the same date from the Molecular Signatures Database (MSigDB v2023.1.Hs updated March 2023). Our analysis was restricted to GO terms containing between 10 and 2000 genes. No size restrictions were placed on the other gene sets, since there were fewer of them. This approach resulted in a total of 8,034 gene sets for analysis. Eight pathway analyses were run using results from 1) the main meta-analysis (“primary” model); 2) the main meta-analysis restricted to common variants (MAF>0.01) (“common-only” model); 3) the main meta-analysis after excluding the *APOE* region (44–46 Mb on chromosome 19 in GRCh38) (“no-apoe” model); 4) the main meta-analysis, but mapping variants to genes using a 35kb upstream and 10kb downstream window (“larger window” model); 5) the no-proxy meta-analysis (“no-proxy” model); 6) the no-proxy meta-analysis restricted to common variants (“common only no-proxy” model); 7) the no-biobank meta-analysis (“no-biobank” model); 6) the no-biobank meta-analysis restricted to common variants (“common only no-biobank” model).

### PheWAS

We extracted with FUMA v1.5.2^45^ all variants in LD (r^2^ > 0.75) with the index variants of the novel Tier 1 and Tier 2 loci in the EUR population from 1000 Genomes Phase 3. The index variant rs7481951 was not available in FUMA, and was replaced by rs10833712, its best tag variant (r^2^=0.827) according to TopLD^46^. We then extracted from the GWAS catalog (e112_r2024-07-08) all traits associated with those variants at the GWS level.

We also extracted the results of the frequent (MAF > 1%) index variants of the Tier 1 and Tier 2 loci in the GWAS of 11 NPEs^17^. The samples included in the neuropathological endophenotypes GWAS largely overlap with the ones included in the ADRD meta-analysis.

### Genetic correlation analyses

Using the no-proxy ADRD GWAS summary statistics, we computed with LDSC v1.0.1^21,47^ the genetic correlation between ADRD and Parkinson’s disease^48^, frontotemporal dementia (FTD)^49^, frontotemporal lobar degeneration with neuronal inclusions of the TAR DNA-binding protein 43 (FTLD-TDP)^50^, Lewy body dementia^51^ amyotrophic lateral sclerosis^52^, educational attainment^53^, stroke and its subtypes^54^ and 11 neuropathological endophenotypes^17^. For amyotrophic lateral sclerosis, FTLD-TDP, Lewy body dementia, Parkinson’s disease and the stroke phenotypes, we used the harmonized version of the summary statistics available in the GWAS catalog^55^. We used the pre-computed “eur_w_ld_chr” LD scores built from 1000 Genomes European data. The analysis was restricted to HapMap 3 variants and excluded variants in the *APOE* locus, with A/T or C/G alleles, with MAF below 1%, with duplicated rsID and indels. Correlation was considered significant if P-value was below 3.13×10^-3^, corresponding to a Bonferroni correction for 16 phenotypes for which genetic correlation could be computed. The no-proxy summary statistics were selected for this analysis to maximize power while avoiding biases that can be observed when using genome-wide summary statistics from studies including proxy cases^14–16^. However, we assessed the sensitivity of the results using the main and no-biobank ADRD summary statistics.

### Polygenic score analyses

We constructed three polygenic scores (PGS) using the Tier 1 main and secondary signals (except *APOE*) detected in the main, no-proxy and no-biobank meta-analyses respectively (Supplementary Table 15), and tested their association with the 11 NPEs in the ADC/NACC and ACT studies. Ordinal NPEs (amyloid-β plaques, CERAD score, arteriolosclerosis, atherosclerosis, CAA, LATE-NC and Lewy body) were dichotomized in two groups - none/mild vs moderate/severe -, as well as Braak NFT stage – stages 0 to 4 vs stages 5 and 6. We first considered as candidate signals the genome-wide significant lead and secondary Tier 1 signals detected in the main analysis. For each PGS (main, no-proxy and no-biobank), we then i) selected only the candidate signals that were genome-wide significant in the respective meta-analysis and ii) selected the index variant of each of those signals in the respective meta-analysis. The *LILRB2* signals detected in the no-biobank meta-analysis were different from the main and secondary signals detected in the same locus in the main meta-analysis; we thus manually included the lead variant of this signal (chr19:54267597:C:T) in the no-biobank meta-analysis in the corresponding PGS so that this signal could be taken into account. Variant chr4:993555:G:T was not available in the NPE genetic data and was replaced by chr4:973547:G:T, its best tag variant in the European TOPMed population according to TopLD (r^2^=0.869). 115, 91 and 65 variants were finally considered to compute the main, no-proxy and no-biobank PGS respectively. Those scores were computed for each individual with PLINK 2.0 using the function *score* as the weighted average of the number of risk-increasing alleles for each variant, using dosages, and were scaled to obtain the PGS^56^:

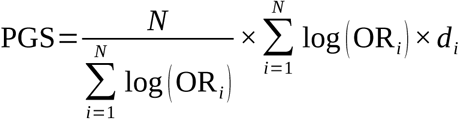

where N is the number of variants included in the score, and OR_i_ and d_i_ are respectively the odds-ratio and dosage of the risk allele of variant i. Each odds-ratio represent the impact of the variant on ADRD risk and was estimated by performing again the meta-analysis after excluding the biobanks (UKBB, FinnGen, deCODE and HUNT) and the studies overlapping with the NPE ones (ACT, ADC/NACC, ROSMAP1 and CSDC). Odds-ratio were then estimated with GCTA COJO by performing for each score separately a joint analysis of all variants included, following the same pipeline as in the conditional and joint analyses described above (Supplementary Table 15). The effect size estimated for chr4:993555:G:T was considered for the tag variant chr4:973547:G:T. Association between each binary NPE and each PGS was measured with a logistic regression in the ADC/NACC and ACT studies separately. The models were adjusted for age at death, sex, the number of APOE ε4 and ε2 alleles, 10 PCs and centers. Effect sizes were then meta-analysed across studies with METAL using a fixed-effect meta-analysis with an inverse variance weighted approach. An association was considered significant if P-value was below 2.27×10^-3^, corresponding to a Bonferroni correction for 22 tests (11 NPEs in two studies). Sensitivity analyses were conducted by additionally adjusting on i) AD diagnosis, coded in three categories (not impaired, AD/mild cognitive impairment and unknown/other dementia); ii) the three AD NPEs; iii) both AD diagnosis and the 3 AD NPEs or iv) AD NPEs and LATE-NC. For Braak stage and CERAD score, we additionally tested the interaction between the PGS and the number of APOE ε4 and APOE ε2 alleles.

The odds-ratio of the association with the PGS measures the effect of carrying one additional average risk allele. Those odds-ratio can not be compared across the three scores as the average risk is different in each of them. To allow comparisons across scores, we divided each genetic score into the quintiles and the deciles of the pooled distribution of the PGS across all individuals from the ADC/NACC and ACT studies. We then computed in ADC/NACC the association of Braak stage and CERAD score with the PGS deciles with the same model as the raw genetic scores, using the median quintile as the reference group. To allow comparison with the smaller ACT study, association results were also computed per quintiles in both ACT and ADC/NACC.

The discriminative performance of the PGS was assessed through three statistics: i) the AUC; ii) Nagelkerke’s pseudo R-squared^57^ and iii) R^2^ on the liability-scale. AUC was computed for each study separately with the *auc* function from the pROC 1.18.5 R package^58^ for the null logistic model including only age, sex, *APOE* ε4 and ε2, PCs and centers, and for the full model including additionally the PGS. AUC between the two models were then compared with the DeLong’s test^59^ as implemented in the *roc.test* function. The same pipeline was applied to compare the AUC across the models including respectively the main, no-proxy and no-biobank PGS. Nagelkerke pseudo-R^2^ was computed with the *nagelkerke* function from the R package rcompanion 2.5.0^60^. We also computed the variance explained by each PGS on the observed scale as the difference of the fraction of variance explained by a linear model under the full and the null model. It was then transformed to the liability scale using the approach from Lee et al^61^ and varying population prevalence values from 0.1 to 0.9.

## Data availability

Summary statistics of the main, no-proxy and no-biobank meta-analyses will be made available upon publication through the European Bioinformatics Institute GWAS Catalog (https://www.ebi.ac.uk/gwas/) and NIAGADS (https://dss.niagads.org/).

## Code availability

The software we used is referenced in the Online Methods and Supplementary Information, and the corresponding URLs are provided in the Supplementary Information.

## Acknowledgments

Acknowledgments and funding of each consortium are provided in the Supplementary Information.

Authorship

## Corresponding authors

Jean-Charles Lambert: Céline Bellenguez

## EADB, EADI and Bonn

Céline Bellenguez^1^, Atahualpa Castillo^2^, Najaf Amin^3^, Sven J. van der Lee^4,5,6^, Manon Muntaner^1^, Kayenat Parveen^7,8^, Fahri Küçükali^9,10^, Benjamin Grenier-Boley^1^, Sami Heikkinen^11^, Itziar de Rojas^12,13^, Maria Carolina Dalmasso^7,14^, Luca Kleineidam^7,15,16^, Oliver Peters^17,18^, Anja Schneider^19,20^, Martin Dichgans^21,22,23^, Dan Rujescu^24^, Norbert Scherbaum^25^, Jürgen Deckert^26^, Steffi Riedel-Heller^27^, Lucrezia Hausner^28^, Laura Molina Porcel^29,30^, Emrah Düzel^31,32^, Timo Grimmer^33^, Jens Wiltfang^34,35,36^, Stefanie Heilmann-Heimbach^37^, Susanne Moebus^38^, Matthias Schmid^39,40^, Thomas Tegos^41^, Nikolaos Scarmeas^42,43^, Oriol Dols-Icardo^44,45^, Fermin Moreno^46,45,47^, Jordi Pérez-Tur^48,45^, María J. Bullido^49,45,50,51^, Pau Pastor^52,53^, Raquel Sánchez-Valle^54^, Victoria Álvarez^55,56^, Mercè Boada^12,13^, Pablo García-González^12^, Raquel Puerta^12^, Pablo Mir^57,45^, Luis M. Real^58,59^, Gerard Piñol-Ripoll^60,61^, Jose María García-Alberca^62,45,59^, Eloy Rodriguez-Rodriguez^63,45^, Hilkka Soininen^64^, Alexandre de Mendonça^65^, Shima Mehrabian^66^, Jakub Hort^67^, Martin Vyhnalek^68,69^, Nicolai Sandau^70^, Jiao Luo^70^, Jesper Qvist Thomassen^70^, Yolande A.L. Pijnenburg^4,6^, Wiesje van der Flier^4,6^, Harro Seelaar^71^, Inez Ramakers^72^, Janne Papma^71^, Marc Hulsman^4,6^, Gert-Jan Biessels^71^, Caroline Graff^73,74^, Hakan Thonberg^73,74^, Abbe Ullgren^73,74^, Goran Papenberg^75^, Vilmantas Giedraitis^76^, Malin Löwenmark^76^, Lena Kilander^76^, Julie Williams^77,2^, Peter Holmans^2^, Julie Le Borgne^1^, Sagnik Palmal^1^, Aude Nicolas^1^, Philippe Amouyel^1^, Anne Boland^78^, Jean-François Deleuze^78^, Gael Nicolas^79^, Carole Dufouil^80,81^, Florence Pasquier^82^, Olivier Hanon^83^, Stéphanie Debette^84,85^, Edna Grünblatt^86,87,88^, Julius Popp^89,90,91^, Daniela Galimberti^92,93^, Beatrice Arosio^94,95^, Patrizia Mecocci^96,97^, Vincenzo Solfrizzi^98,99^, Lucilla Parnetti^100^, Alessio Squassina^101^, Lucio Tremolizzo^102,103^, Barbara Borroni^104,105^, Michael Wagner^8,19^, Benedetta Nacmias^106,107^, Marco Spallazzi^108^, Davide Seripa^109^, Innocenzo Rainero^110^, Antonio Daniele^111,112^, Paola Bossù^113^, Carlo Masullo^114^, Giacomina Rossi^115^, Frank Jessen^116,19,117^, Henne Holstege^4,6,118^, Karen Mather^119^, M. Victoria Fernandez^12^, Patrick G Kehoe^120^, Magda Tsolaki^41^, Cornelia van Duijn^121,122,123^, Ruth Frikke-Schmidt^70,124^, Roberta Ghidoni^105^, Pascual Sánchez-Juan^125,45^, Kristel Sleegers^9,10^, Martin Ingelsson^76,126,127^, Mikko Hiltunen^11^, Rebecca Sims^2^, Ole Andreassen^128,129^, Agustín Ruiz^12,13,130^, Alfredo Ramirez^7,8,131,132,19^, Jean-Charles Lambert^1^

^1^Univ. Lille, Inserm, CHU Lille, Institut Pasteur de Lille, LabEx DISTALZ - U1167-RID-AGE Facteurs de risque et déterminants moléculaires des maladies liées au vieillissement, Lille, France

^2^Centre for Neuropsychiatric Genetics and Genomics, Division of Psychological Medicine and Clinical Neuroscience, School of Medicine, Cardiff University, Cardiff, UK

^3^Nuffield Department of Population Health, University of Oxford, Oxford, UK

^4^Alzheimer Center Amsterdam, Neurology, Vrije Universiteit Amsterdam, Amsterdam UMC location VUmc, Amsterdam, The Netherlands.

^5^Department of Complex Trait Genetics, Center for Neurogenomics and Cognitive Research, Amsterdam Neuroscience, Vrije University, Amsterdam, The Netherlands.

^6^Amsterdam Neuroscience, Neurodegeneration, Amsterdam, The Netherlands.

^7^Division of Neurogenetics and Molecular Psychiatry, Department of Psychiatry and Psychotherapy, Faculty of Medicine and University Hospital Cologne, University of Cologne, Cologne, Germany

^8^Department of Cognitive Disorders and Old Age Psychiatry, University Hospital Bonn, Medical Faculty, Bonn, Germany

^9^Complex Genetics of Alzheimer’s Disease Group, VIB Center for Molecular Neurology, VIB, Antwerp, Belgium

^10^Department of Biomedical Sciences, University of Antwerp, Antwerp, Belgium

^11^Institute of Biomedicine, University of Eastern Finland, Finland

^12^Ace Alzheimer Center Barcelona, Universitat Internacional de Catalunya (UIC), Barcelona, Spain

^13^Networking Research Center on Neurodegenerative Diseases (CIBERNED), Instituto de Salud Carlos III, Madrid, Spain

^14^Estudios en Neurociencias y Sistemas Complejos (ENyS) CONICET-HEC-UNAJ

^15^Department of Neurodegeneration and Geriatric Psychiatry, University of Bonn, Bonn, Germany

^16^German Center for Neurodegenerative Diseases (DZNE Bonn), Bonn, Germany

^17^German Center for Neurodegenerative Diseases (DZNE), Berlin, Germany

^18^Charité – Universitätsmedizin Berlin, corporate member of Freie Universität Berlin, Humboldt-Universität zu Berlin, and Berlin Institute of Health, Institute of Psychiatry and Psychotherapy, Hindenburgdamm 30, 12203 Berlin, Germany

^19^German Center for Neurodegenerative Diseases (DZNE), Bonn, Germany

^20^Department of Cognitive Disorders and Old Age Psychiatry, University Hospital Bonn, Venusberg-Campus 1, 53127 Bonn, Germany

^21^Institute for Stroke and Dementia Research (ISD), University Hospital, LMU Munich, Munich, Germany.

^22^German Center for Neurodegenerative Diseases (DZNE), Munich, Germany

^23^Munich Cluster for Systems Neurology (SyNergy), Munich, Germany

^24^Martin-Luther-University Halle-Wittenberg, University Clinic and Outpatient Clinic for Psychiatry, Psychotherapy and Psychosomatics, Halle (Saale), Germany

^25^LVR-University Hospital Essen, Department of Psychiatry and Psychotherapy, Medical Faculty, University of Duisburg-Essen, Essen, Germany

^26^Department of Psychiatry, Psychosomatics and Psychotherapy, Center of Mental Health, University Hospital of Würzburg, Germany

^27^Institute of Social Medicine, Occupational Health and Public Health, University of Leipzig, 04103 Leipzig, Germany.

^28^Department of Geriatric Psychiatry, Central Institute for Mental Health Mannheim, Faculty Mannheim, University of Heidelberg, Germany

^29^Neurological Tissue Bank - Biobanc-Hospital Clinic-IDIBAPS, Barcelona, Spain

^30^Alzheimer’s disease and other cognitive disorders Unit. Neurology Department, Hospital Clinic, Barcelona, Spain

^31^German Center for Neurodegenerative Diseases (DZNE), Magdeburg, Germany

^32^Institute of Cognitive Neurology and Dementia Research (IKND), Otto-von-Guericke University, Magdeburg, Germany

^33^Center for Cognitive Disorders, Department of Psychiatry and Psychotherapy, Technical University of Munich, School of Medicine, Munich, Germany

^34^Department of Psychiatry and Psychotherapy, University Medical Center Goettingen, Goettingen, Germany

^35^German Center for Neurodegenerative Diseases (DZNE), Goettingen, Germany

^36^Medical Science Department, iBiMED, Aveiro, Portugal

^37^Institute of Human Genetics, University of Bonn, School of Medicine & University Hospital Bonn, Bonn, Germany

^38^Institute for Urban Public Health, University Hospital of University Duisburg-Essen, Essen, Germany.

^39^German Center for Neurodegenerative Diseases (DZNE, Bonn), Bonn, Germany

^40^Institute of Medical Biometry, Informatics and Epidemiology, University Hospital of Bonn, Bonn, Germany

^41^1st Department of Neurology, Medical school, Aristotle University of Thessaloniki, Thessaloniki, Makedonia, Greece

^42^Taub Institute for Research in Alzheimer’s Disease and the Aging Brain, The Gertrude H. Sergievsky Center, Department of Neurology, Columbia University, New York, NY

^43^1st Department of Neurology, Aiginition Hospital, National and Kapodistrian University of Athens, Medical School, Greece

^44^Sant Pau Memory Unit, Institut de Recerca Sant Pau, Department of Neurology, Hospital de la Santa Creu i Sant Pau, Universitat Autònoma de Barcelona, Barcelona, Spain.

^45^CIBERNED, Network Center for Biomedical Research in Neurodegenerative Diseases, National Institute of Health Carlos III, Madrid, Spain

^46^Department of Neurology. Hospital Universitario Donostia. San Sebastian, Spain

^47^Neurosciences Area. Instituto Biodonostia. San Sebastian, Spain

^48^Unitat de Genètica Molecular, Institut de Biomedicina de València-CSIC, Valencia, Spain

^49^Centro de Biología Molecular Severo Ochoa (UAM-CSIC)

^50^Instituto de Investigacion Sanitaria ‘Hospital la Paz’ (IdIPaz), Madrid, Spain

^51^Universidad Autónoma de Madrid

^52^Fundació Docència i Recerca MútuaTerrassa, Terrassa, Barcelona, Spain

^53^Memory Disorders Unit, Department of Neurology, Hospital Universitari Mutua de Terrassa, Terrassa, Barcelona, Spain

^54^Alzheimer’s disease and other cognitive disorders unit. Service of Neurology. Hospital Clínic of Barcelona. Institut d’Investigacions Biomèdiques August Pi i Sunyer, University of Barcelona, Barcelona, Spain

^55^Laboratorio de Genética. Hospital Universitario Central de Asturias, Oviedo, Spain

^56^Instituto de Investigación Sanitaria del Principado de Asturias (ISPA)

^57^Unidad de Trastornos del Movimiento, Servicio de Neurología y Neurofisiología. Instituto de Biomedicina de Sevilla (IBiS), Hospital Universitario Virgen del Rocío/CSIC/Universidad de Sevilla, Seville, Spain

^58^Unidad Clínica de Enfermedades Infecciosas y Microbiología. Hospital Universitario de Valme, Sevilla, Spain

^59^Depatamento de Especialidades Quirúrgicas, Bioquímica e Inmunología. Facultad de Medicina. Universidad de Málaga. Málaga, Spain

^60^Unitat Trastorns Cognitius, Hospital Universitari Santa Maria de Lleida, Lleida, Spain

^61^Institut de Recerca Biomedica de Lleida (IRBLLeida), Lleida, Spain

^62^Alzheimer Research Center & Memory Clinic, Andalusian Institute for Neuroscience, Málaga, Spain. ^63^Neurology Service, Marqués de Valdecilla University Hospital (University of Cantabria and IDIVAL), Santander, Spain.

^64^Institute of Clinical Medicine - Neurology, University of Eastern Finland, Finland

^65^Faculty of Medicine, University of Lisbon, Portugal

^66^Clinic of Neurology, UH “Alexandrovska”, Medical University - Sofia, Sofia, Bulgaria

^67^Memory Clinic, Department of Neurology, Charles University, Second Faculty of Medicine and Motol University Hospital, Czech Republic

^68^Memory Clinic, Department of Neurology, Charles University, 2nd Faculty of Medicine and Motol University Hospital, Czech Republic

^69^International Clinical Research Center, St. Anne’s University Hospital Brno, Brno, Czech Republic ^70^Department of Clinical Biochemistry, Copenhagen University Hospital - Rigshospitalet, Copenhagen, Denmark

^71^Amsterdam Neuroscience, Neurodegeneration, Amsterdam, The Netherlands

^72^Maastricht University, Department of Psychiatry & Neuropsychologie, Alzheimer Center Limburg, Maastricht, the Netherlands

^73^Karolinska Institutet, Center for Alzheimer Research, Department NVS, Division of Neurogeriatrics, 171 64 Stockholm Sweden

^74^Unit for Hereditary Dementias, Karolinska University Hospital-Solna, 171 64 Stockholm Sweden

^75^Aging Research Center, Department of Neurobiology, Care Sciences and Society, Karolinska Institutet and Stockholm University, Stockholm, Sweden

^76^Department of Public Health and Caring Sciences, Molecular Geriatrics, Rudbeck Laboratory, Uppsala University, Uppsala, Sweden

^77^UKDRI@ Cardiff, School of Medicine, Cardiff University, Cardiff, UK

^78^Université Paris-Saclay, CEA, Centre National de Recherche en Génomique Humaine, 91057, Evry, France

^79^Univ Rouen Normandie, Normandie Univ, Inserm U1245 and CHU Rouen, Department of Genetics and CNRMAJ, F-76000 Rouen, France

^80^Inserm, Bordeaux Population Health Research Center, UMR 1219, Univ. Bordeaux, ISPED, CIC 1401-EC, Univ Bordeaux,Bordeaux, France

^81^CHU de Bordeaux, Pole santé publique, Bordeaux, France

^82^Univ Lille Inserm 1171, CHU Clinical and Research Memory Research Centre (CMRR) of Distalz Lille France.

^83^Université de Paris, EA 4468, APHP, Hôpital Broca, Paris, France

^84^University Bordeaux, Inserm, Bordeaux Population Health Research Center, France

^85^Department of Neurology, Bordeaux University Hospital, Bordeaux, France

^86^Department of Child and Adolescent Psychiatry and Psychotherapy, University Hospital of Psychiatry Zurich, University of Zurich, Zurich, Switzerland

^87^Neuroscience Center Zurich, University of Zurich and ETH Zurich, Switzerland

^88^Zurich Center for Integrative Human Physiology, University of Zurich, Switzerland

^89^Old Age Psychiatry, Department of Psychiatry, Lausanne University Hospital, Lausanne, Switzerland ^90^Department of Geriatric Psychiatry, University Hospital of Psychiatry Zürich, Zürich, Switzerland ^91^Institute for Regenerative Medicine, University of Zürich, Switzerland

^92^Neurodegenerative Diseases Unit, Fondazione IRCCS Ca’ Granda, Ospedale Policlinico, Milan, IT

^93^Dept. of Biomedical, Surgical and Dental Sciences, University of Milan, Milan, IT

^94^Department of Clinical Sciences and Community Health, University of Milan, 20122 Milan, Italy ^95^Geriatric Unit, Fondazione IRCCS Ca’ Granda Ospedale Maggiore Policlinico, 20122 Milan, Italy ^96^Institute of Gerontology and Geriatrics,Department of Medicine and Surgery, University of Perugia, Italy

^97^Division of Clinical Geriatrics, Department of Neurobiology, Care Sciences and Society, Karolinska Institutet, Stockholm, Sweden

^98^Interdisciplinary Department of Medicine, Geriatric Medicine and Memory Unit, University of Bari “A. Moro, Bari, Italy

^99^Academic Division “C. Frugoni” & Hospital Division of Internal and Geriatric Medicine, Policlinico Hospital, Bari, Italy

^100^Centre for Memory Disturbances, Lab of Clinical Neurochemistry, Section of Neurology, University of Perugia, Perugia, Italy

^101^Department of Biomedical Sciences, Section of Neuroscience and Clinical Pharmacology, University of Cagliari, Cagliari, Italy.

^102^Neurology Unit, IRCCS “San Gerardo dei Tintori”, Monza, Italy

^103^School of Medicine and Surgery, University of Milano-Bicocca, Monza, Italy

^104^Department of Clinical and Experimental Sciences, University of Brescia, Italy

^105^Molecular Markers Laboratory, IRCCS Istituto Centro San Giovanni di Dio Fatebenefratelli, Brescia, Italy

^106^Department of Neuroscience, Psychology, Drug Research and Child Health University of Florence, Florence, Italy

^107^IRCCS Fondazione Don Carlo Gnocchi, Florence, Italy

^108^Department of Medicine and Surgery, Unit of Neurology, University-Hospital of Parma, Parma, Italy ^109^Department of Hematology and Stem Cell Transplant, Vito Fazzi Hospital, Lecce, Italy ^110^Department of Neuroscience “Rita Levi Montalcini”, University of Torino, Torino, Italy

^111^Department of Neuroscience, Università Cattolica del Sacro Cuore, Rome, Italy

^112^Neurology Unit, IRCCS Fondazione Policlinico Universitario A. Gemelli, Rome, Italy

^113^Laboratory of Experimental Neuropsychobiology, Clinical Neuroscience and Neurorehabilitation Department, IRCCS Santa Lucia Foundation, Rome, Italy

^114^Institute of Neurology, Catholic University of the Sacred Heart, Rome, Itlay

^115^Unit of Neurology V - Neuropathology, Fondazione IRCCS Istituto Neurologico Carlo Besta, Milan, Italy ^116^Department of Psychiatry and Psychotherapy, Faculty of Medicine and University Hospital Cologne, University of Cologne, Cologne, Germany

^117^Cluster of Excellence Cellular Stress Responses in Aging-associated Diseases (CECAD), University of Cologne, Cologne, Germany

^118^Genomics of Neurodegenerative Diseases and Aging, Human Genetics, Vrije Universiteit Amsterdam, Amsterdam UMC location VUmc, Amsterdam, The Netherlands.

^119^Centre for Healthy Brain Ageing, School of Psychiatry, Faculty of Medicine, University of New South Wales, Sydney, Australia

^120^Translational Health Sciences, Bristol Medical School, University of Bristol, Bristol, UK

^121^Department of Epidemiology, ErasmusMC, Rotterdam, The Netherlands

^122^Nuffield Department of Population Health, University of Oxford, Oxford, United Kingdom ^123^Centre for Artificial Intelligence in Precision Medicine, University of Oxford, United Kingdom ^124^Department of Clinical Medicine, University of Copenhagen, Copenhagen, Denmark ^125^Alzheimer’s Centre Reina Sofia-CIEN Foundation-ISCIII, Madrid, Spain

^126^Krembil Brain Institute, University Health Network, Toronto, Ontario, Canada

^127^Tanz Centre for Research in Neurodegenerative Diseases, Departments of Medicine and Laboratory Medicine & Pathobiology, University of Toronto, Toronto, Ontario, Canada

^128^NORMENT Centre, Division of Mental Health and Addiction, Oslo University Hospital, Oslo, Norway

^129^Institute of Clinical Medicine, University of Oslo, Oslo, Norway

^130^Biggs Institute for Alzheimer’s and Neurodegenerative Diseases, University of Texas Health Science Center, San Antonio, Texas, USA.

^131^Department of Psychiatry & Glenn Biggs Institute for Alzheimer’s and Neurodegenerative Diseases, San Antonio, TX, USA

^132^Cologne Excellence Cluster on Cellular Stress Responses in Aging-Associated Disease (CECAD), University of Cologne, Cologne, Germany.

## ADGC

Adam C. Naj^1,2^, Farid Rajabli^3,4^, Penelope Benchek^5^, Lincoln M.P. Shade^6^, Qi Qiao^6,7^, Nicholas Kushch^4^, Jin Sha^1^, Katrina Bazemore^1^, Congcong Zhu^8^, Wan-Ping Lee^2^, Jacob Haut^1^, Kara L. Hamilton-Nelson^4^, Nicholas R. Wheeler^5,9^, Yi Zhao^2^, John J. Farrell^8^, Michelle A. Grunin^5^, Yuk Yee Leung^2^, Pavel P. Kuksa^2^, Donghe Li^8^, Eder Lucio da Fonseca^4^, Jesse B. Mez^10^, Ellen L. Palmer^5^, Jagan Pillai^11^, Richard M. Sherva^12^, Yeunjoo E. Song^9,5^, Xiaoling Zhang^8,13^, Takeshi Ikeuchi^14^, Taha Iqbal^1^, Otto Valladares^2^, Dolly Reyes-Dumeyer^15,16^, Amanda B. Kuzma^2^, Erin Abner^17,7^, Larry D. Adams^3^, Alyssa Aguirre^18^, Marilyn S. Albert^19^, Roger L. Albin^20,21,22^, Mariet Allen^23^, Liana G. Apostolova^24,25^, Steven E. Arnold^26^, Sanjay Asthana^27,28,29^, Craig S. Atwood^27,28,29^, Sanford Auerbach^10^, Clinton T. Baldwin^8^, Robert C. Barber^30^, Lisa L. Barnes^31,32,33^, Sandra Barral^15,34,16^, Thomas G. Beach^35^, James T. Becker^36^, Gary W. Beecham^4^, Duane Beekly^37^, David A. Bennett^31,33^, John Bertelson^38^, Thomas D. Bird^39,40^, Deborah Blacker^41,42^, Bradley F. Boeve^43^, James D. Bowen^44^, Adam Boxer^45^, James Brewer^46^, Jeffrey M. Burns^47^, Joseph D. Buxbaum^48,49,50^, Nigel J. Cairns^51^, Laura B. Cantwell^2^, Chuanhai Cao^52^, Christopher S. Carlson^53^, Cynthia M. Carlsson^29,28^, Regina M. Carney^54^, Minerva M. Carrasquillo^23^, Marie-Francoise Chesselet^55^, Nathaniel A. Chin^27,28^, Helena C. Chui^56^, Jaeyoon Chung^8^, Steven A. Claas^6^, Suzanne Craft^57^, Paul K. Crane^58^, David H. Cribbs^59^, Elizabeth A. Crocco^60^, Carlos Cruchaga^61,62^, Michael L. Cuccaro^4,3^, Munro Cullum^63^, Eveleen Darby^64^, Barbara Davis^65^, Philip L. De Jager^66^, Charles DeCarli^67^, John DeToledo^68^, Malcolm Dick^69^, Dennis W. Dickson^23^, Beth A. Dombroski^2^, Rachelle S. Doody^64^, Ranjan Duara^70^, Logan C. Dumitrescu^71^, Nilüfer Ertekin-Taner^23,72^, Denis A. Evans^73^, Kelley M. Faber^74^, Thomas J. Fairchild^75^, Kenneth B. Fallon^76^, Martin R. Farlow^77^, Victoria Fernandez-Hernandez^78^, Robert P. Friedland^79^, Tatiana M. Foroud^74^, Matthew P. Frosch^80^, Brian Fulton-Howard^81^, Douglas R. Galasko^46^, Marla Gearing^82,83^, Daniel H. Geschwind^55^, Bernardino Ghetti^84^, John R. Gilbert^4,3^, Rodney C.P. Go^76^, Alison M. Goate^48^, Thomas J. Grabowski^40,85^, Neill R. Graff-Radford^23,72^, Nora E. Gray^86,87^, John H. Growdon^88^, Hakon Hakonarson^89,90^, James Hall^30^, Ronald L. Hamilton^91^, Oscar Harari^92^, John Hardy^93,94^, Elizabeth Head^95^, Victor W. Henderson^96,97^, Michelle Hernandez^68^, Timothy J. Hohman^71,98^, Lawrence S. Honig^15^, Ryan M. Huebinger^99^, Matthew J. Huentelman^100^, Bradley T. Hyman^88^, Linda S. Hynan^63,65^, Laura Ibanez^101,102^, Gail P. Jarvik^103,104^, Suman Jayadev^40^, Lee-Way Jin^105^, Kim Johnson^68^, Leigh Johnson^106^, M. Ilyas Kamboh^107,108,109^, Yuriko Katsumata^6,7^, Mindy J. Katz^110^, John S. Kauwe^111,112^, Jeffrey A. Kaye^86,87^, C. Dirk Keene^113^, Aisha Khaleeq^64^, Masataka Kikuchi^14^, Janice Knebl^106^, Neil W. Kowall^10,114^, Joel H. Kramer^115^, Walter A. Kukull^116^, Frank M. LaFerla^117^, James J. Lah^118^, Eric B. Larson^119^, Alan Lerner^5^, James B. Leverenz^11^, Allan I. Levey^118^, Andrew P. Lieberman^120^, Richard B. Lipton^110^, Mark Logue^8,121,122^, Oscar L. Lopez^36^, Kathryn L. Lunetta^13^, Constantine G. Lyketsos^123^, Douglas Mains^106,124^, Daniel C. Marson^125^, Eden R R. Martin^4,3^, Frank Martiniuk^126^, Deborah C. Mash^127^, Eliezer Masliah^46,128^, Paul Massman^64^, Arjun Masurkar^129^, Wayne C. McCormick^58^, Susan M. McCurry^130^, Ann C. McKee^10,131^, Marsel Mesulam^132,133^, Bruce L. Miller^134^, Carol A. Miller^135^, Joshua W. Miller^105^, Thomas J. Montine^136^, Edwin S. Monuki^137^, John C. Morris^51,138,139,102^, Shubhabrata Mukherjee^58^, Amanda J. Myers^140^, Trung Nguyen^141^, Thomas Obisesan^142^, Sid O’Bryant^143^, John M. Olichney^144^, Raymond Palmer^145^, Joseph E. Parisi^146^, Henry L. Paulson^20,22^, Valory Pavlik^64^, David Paydarfar^18^, Victoria Perez^68^, Elaine Peskind^147^, Ronald C. Petersen^43^, Helen Petrovitch^148^, Marsha Polk^145^, Wayne W. Poon^69^, Huntington Potter^149^, Liming Qu^2^, Mary Quiceno^150,151^, Joseph F. Quinn^86,87^, Ashok Raj^52^, Murray Raskind^147^, Eric M. Reiman^100,152,153,154^, Barry Reisberg^155,129^, Joan S. Reisch^156^, John M. Ringman^157^, Erik D. Roberson^125^, Monica Rodriguear^64^, Ekaterina Rogaeva^158^, Howard J. Rosen^45^, Roger N. Rosenberg^141^, Donald R. Royall^159^, Marwan Sabbagh^160^, A. Dessa Sadovnick^161^, Mark A. Sager^28^, Mary Sano^50^, Andrew J. Saykin^74,162^, Julie A. Schneider^31,33,163^, Lon S. Schneider^164,56^, William W. Seeley^45^, Susan H. Slifer^4^, Scott Small^15,16^, Amanda G. Smith^52^, Joshua A. Sonnen^113^, Peter St George-Hyslop^165,166^, Takiyah D. Starks^167^, Robert A. Stern^10^, Alan B. Stevens^168,169,170^, Stephen M. Strittmatter^171^, David Sultzer^172^, Russell H. Swerdlow^47^, Rudolph E. Tanzi^88^, Jeffrey L. Tilson^173^, Juan C. Troncoso^174^, Magda Tsolaki^175^, Debby W. Tsuang^39,147^, Vivianna M. Van Deerlin^2^, Linda J. Van Eldik^176,7^, Jeffery M. Vance^3,4^, Badri N. Vardarajan^15^, Robert Vassar^132,133^, Harry V. Vinters^177,157^, Jean-Paul Vonsattel^15^, Sandra Weintraub^178^, Kathleen A. Welsh-Bohmer^179,180^, Patrice L. Whitehead^4^, Ellen M. Wijsman^103,104,181^, Kirk C. Wilhelmsen^182^, Benjamin Williams^183^, Jennifer Williamson^15^, Henrik Wilms^68^, Thomas S. Wingo^118^, Thomas Wisniewski^184,185^, Randall L. Woltjer^186^, Clinton B. Wright^187^, Chuang-Kuo Wu^68^, Steven G. Younkin^23,72^, Chang-En Yu^58^, Lei Yu^31,33^, Xiongwei Zhu^188^, Brian W. Kunkle^3,4^, William S. Bush^5,9^, Akinori Miyashita^14^, Giuseppe Tosto^15,16^, Gyungah R. Jun^8,13,189^, Christiane Reitz^190,34,16^, Goldie S. Byrd^191^, David W. Fardo^192,7^, Li-San Wang^2^, Lindsay A. Farrer^12,10,189,193,13^, Jonathan L. Haines^5,9^, Richard Mayeux^15^, Margaret A. Pericak-Vance^3,4^, Gerard D. Schellenberg^2^

^1^Department of Biostatistics, Epidemiology, and Informatics, Perelman School of Medicine, University of Pennsylvania, Philadelphia, Pennsylvania, USA

^2^Penn Neurodegeneration Genomics Center, Department of Pathology and Laboratory Medicine, Perelman School of Medicine, University of Pennsylvania, Philadelphia, Pennsylvania, USA

^3^Dr. John T. Macdonald Foundation Department of Human Genetics, Miller School of Medicine, University of Miami, Miami, Florida, USA

^4^The John P. Hussman Institute for Human Genomics, University of Miami, Miami, Florida, USA ^5^Department of Population and Quantitative Health Sciences, Case Western Reserve University, Cleveland, Ohio, USA

^6^Department of Biostatistics, College of Public Health, University of Kentucky, Lexington, Kentucky, USA

^7^Sanders-Brown Center on Aging, University of Kentucky, Lexington, Kentucky, USA

^8^Department of Medicine (Biomedical Genetics), Boston University Chobanian & Avedisian School of Medicine, Boston, Massachusetts, USA

^9^Cleveland Institute for Computational Biology, Case Western Reserve University, Cleveland, Ohio, USA ^10^Department of Neurology, Boston University Chobanian & Avedisian School of Medicine, Boston, Massachusetts, USA

^11^Cleveland Clinic Lou Ruvo Center for Brain Health, Cleveland Clinic, Cleveland, Ohio, USA

^12^Department of Medicine (Biomedical Genetics),Boston University Chobanian & Avedisian School of Medicine, Boston, Massachusetts, USA

^13^Department of Biostatistics, Boston University School of Public Health, Boston, Massachusetts, USA

^14^Molecular Genetics Division, Brain Research Institute, Niigata University, Niigata, Japan

^15^Taub Institute for Research in Alzheimer’s Disease and the Aging Brain, The Gertrude H. Sergievsky Center, Department of Neurology, Columbia University, New York, New York, USA

^16^Department of Neurology, Columbia University, New York, New York, USA

^17^Department of Epidemiology and Environmental Health, College of Public Health, University of Kentucky, Lexington, Kentucky, USA

^18^Department of Neurology, Dell Medical School, University of Texas at Austin, Austin, Texas, USA

^19^Department of Neurology, Johns Hopkins University, Baltimore, Maryland, USA

^20^Department of Neurology, University of Michigan, Ann Arbor, Michigan, USA

^21^Geriatric Research, Education and Clinical Center (GRECC), VA Ann Arbor Healthcare System (VAAAHS), Ann Arbor, Michigan, USA

^22^Michigan Alzheimer’s Disease Center, University of Michigan, Ann Arbor, Michigan, USA

^23^Department of Neuroscience, Mayo Clinic, Jacksonville, Florida, USA

^24^Departments of Neurology, Radiology, and Medical and Molecular Genetics, Indiana University School of Medicine, Indianapolis, Indiana, USA

^25^Indiana Alzheimer’s Disease Research Center, Indiana University School of Medicine, Indianapolis, Indiana, USA

^26^Department of Psychiatry, Perelman School of Medicine, University of Pennsylvania, Philadelphia, Pennsylvania, USA

^27^Geriatric Research, Education and Clinical Center (GRECC), University of Wisconsin, Madison, Wisconsin, USA

^28^Department of Medicine, University of Wisconsin, Madison, Wisconsin, USA

^29^Wisconsin Alzheimer’s Disease Research Center, Madison, Wisconsin, USA

^30^Department of Pharmacology and Neuroscience, University of North Texas Health Science Center, Fort Worth, Texas, USA

^31^Department of Neurological Sciences, Rush University Medical Center, Chicago, Illinois, USA

^32^Department of Behavioral Sciences, Rush University Medical Center, Chicago, Illinois, USA

^33^Rush Alzheimer’s Disease Center, Rush University Medical Center, Chicago, Illinois, USA

^34^Gertrude H. Sergievsky Center, Columbia University, New York, New York, USA

^35^Civin Laboratory for Neuropathology, Banner Sun Health Research Institute, Phoenix, Arizona, USA

^36^Departments of Psychiatry, Neurology, and Psychology, University of Pittsburgh School of Medicine, Pittsburgh, Pennsylvania, USA

^37^National Alzheimer’s Coordinating Center, University of Washington, Seattle, Washington, USA

^38^Department of Psychiatry, University of Texas at Austin/Dell Medical School, Austin, Texas, USA

^39^VA Puget Sound Health Care System/GRECC, Seattle, Washington, USA

^40^Department of Neurology, University of Washington, Seattle, Washington, USA

^41^Department of Epidemiology, Harvard School of Public Health, Boston, Massachusetts, USA

^42^Department of Psychiatry, Massachusetts General Hospital/Harvard Medical School, Boston, Massachusetts, USA

^43^Department of Neurology, Mayo Clinic, Rochester, Minnesota, USA

^44^Swedish Medical Center, Seattle, Washington, USA

^45^Department of Neurology, University of California San Francisco, San Francisco, California, USA

^46^Department of Neurosciences, University of California San Diego, La Jolla, California, USA

^47^University of Kansas Alzheimer’s Disease Center, University of Kansas Medical Center, Kansas City, Kansas, USA

^48^Department of Genetics and Genomic Sciences, Ronald M. Loeb Center for Alzheimer’s Disease, Icahn School of Medicine at Mount Sinai, New York, New York, USA

^49^Department of Neuroscience, Icahn School of Medicine at Mount Sinai, New York, New York, USA

^50^Department of Psychiatry, Mount Sinai School of Medicine, New York, New York, USA

^51^Department of Pathology and Immunology, Washington University, St. Louis, Missouri, USA

^52^USF Health Byrd Alzheimer’s Institute, University of South Florida, Tampa, Florida, USA

^53^Fred Hutchinson Cancer Research Center, Seattle, Washington, USA

^54^Mental Health and Behavioral Science Service, Bruce W. Carter VA Medical Center, Miami, Florida, USA

^55^Neurogenetics Program, University of California Los Angeles, Los Angeles, California, USA

^56^Department of Neurology, University of Southern California, Los Angeles, California, USA

^57^Section of Gerontology and Geriatric Medicine Research, Wake Forest School of Medicine, Winston-Salem, North Carolina, USA

^58^Department of Medicine, University of Washington, Seattle, Washington, USA

^59^Department of Neurology, University of California Irvine, Irvine, California, USA

^60^Department of Psychiatry and Behavioral Sciences, Miller School of Medicine, University of Miami, Miami, Florida, USA

^61^NeuroGenomics and Informatics, Washington University, St. Louis, Missouri, USA

^62^Department of Psychiatry, Washington University in St. Louis, St Louis, Missouri, USA

^63^Department of Psychiatry, University of Texas Southwestern Medical Center, Dallas, Texas, USA

^64^Alzheimer’s Disease and Memory Disorders Center, Baylor College of Medicine, Houston, Texas, USA

^65^Department of Population and Data Sciences, University of Texas Southwestern Medical Center, Dallas, Texas, USA

^66^Center for Translational and Computational Neuroimmunology, Department of Neurology and the Taub Institute for Research in Alzheimer’s Disease and the Aging Brain, Columbia University Irving Medical Center, New York, New York, USA

^67^Department of Neurology, University of California Davis, Sacramento, California, USA

^68^Departments of Neurology, Pharmacology and Neuroscience, Texas Tech University Health Science Center, Lubbock, Texas, USA

^69^Institute for Memory Impairments and Neurological Disorders, University of California Irvine, Irvine, California, USA

^70^Wien Center for Alzheimer’s Disease and Memory Disorders, Mount Sinai Medical Center, Miami Beach, Florida, USA

^71^Vanderbilt Memory and Alzheimer’s Center, Department of Neurology, Vanderbilt University Medical Center, Nashville, Tennessee, USA

^72^Department of Neurology, Mayo Clinic, Jacksonville, Florida, USA

^73^Rush Institute for Healthy Aging, Department of Internal Medicine, Rush University Medical Center, Chicago, Illinois, USA

^74^Department of Medical and Molecular Genetics, Indiana University, Indianapolis, Indiana, USA

^75^Office of Strategy and Measurement, University of North Texas Health Science Center, Fort Worth, Texas, USA

^76^Department of Pathology, University of Alabama at Birmingham, Birmingham, Alabama, USA

^77^Department of Neurology, Indiana University, Indianapolis, Indiana, USA

^78^Department of Psychiatry and Hope Center Program on Protein Aggregation and Neurodegeneration, Washington University School of Medicine, St. Louis, Missouri, USA

^79^Department of Neurology, University of Louisville School of Medicine, Lousiville, Kentucky, USA

^80^C.S. Kubik Laboratory for Neuropathology, Massachusetts General Hospital, Charlestown, Massachusetts, USA

^81^Department of Neuroscience, Ronald M. Loeb Center for Alzheimer’s Disease, Icahn School of Medicine at Mount Sinai, New York, New York, USA

^82^Department of Pathology and Laboratory Medicine, Emory University, Atlanta, Georgia, USA

^83^Emory Alzheimer’s Disease Center, Emory University, Atlanta, Georgia, USA

^84^Department of Pathology and Laboratory Medicine, Indiana University, Indianapolis, Indiana, USA

^85^Department of Radiology, University of Washington, Seattle, Washington, USA

^86^Department of Neurology, Oregon Health and Science University, Portland, Oregon, USA

^87^Department of Neurology, Portland Veterans Affairs Medical Center, Portland, Oregon, USA

^88^Department of Neurology, Massachusetts General Hospital/Harvard Medical School, Boston, Massachusetts, USA

^89^Center for Applied Genomics, Children’s Hospital of Philadelphia, Philadelphia, Pennsylvania, USA

^90^Division of Human Genetics, Department of Pediatrics, Perelman School of Medicine, University of Pennsylvania, Philadelphia, Pennsylvania, USA

^91^Department of Pathology (Neuropathology), University of Pittsburgh, Pittsburgh, Pennsylvania, USA

^92^Department of Neurology, Ohio State University, Columbus, Ohio, USA

^93^UCL Institute of Neurology, University College London, London, England, UK

^94^Department of Molecular Neuroscience, UCL Institute of Neurology, University College London, London, England, UK

^95^Department of Pathology and Laboratory Medicine, University of California Irvine, Irvine, California, USA

^96^Department of Epidemiology and Population Health, Stanford University, Stanford, California, USA

^97^Department of Neurology and Neurological Sciences, Stanford University, Stanford, California, USA

^98^Vanderbilt Genetics Institute, Division of Genetic Medicine, Department of Medicine, Vanderbilt University Medical Center, Nashville, Tennessee, USA

^99^Department of Surgery, University of Texas Southwestern Medical Center, Dallas, Texas, USA

^100^Neurogenomics Division, Translational Genomics Research Institute, Phoenix, Arizona, USA

^101^Department of Psychiatry, Washington University School of Medicine, St. Louis, Missouri, USA

^102^Hope Center Program on Protein Aggregation and Neurodegeneration, Washington University School of Medicine, St. Louis, Missouri, USA

^103^Department of Genome Sciences, University of Washington, Seattle, Washington, USA

^104^Department of Medicine (Medical Genetics), University of Washington, Seattle, Washington, USA

^105^Department of Pathology and Laboratory Medicine, University of California Davis, Sacramento, California, USA

^106^Department of Health Behavior and Health Systems, University of North Texas Health Science Center, Fort Worth, Texas, USA

^107^Department of Psychiatry, University of Pittsburgh, Pittsburgh, Pennsylvania, USA

^108^Department of Human Genetics, University of Pittsburgh, Pittsburgh, Pennsylvania, USA

^109^Alzheimer’s Disease Research Center, University of Pittsburgh, Pittsburgh, Pennsylvania, USA.

^110^Department of Neurology, Albert Einstein College of Medicine, New York, New York, USA

^111^Department of Neuroscience, Brigham Young University, Provo, Utah, USA

^112^Department of Biology, Brigham Young University, Provo, Utah, USA

^113^Department of Laboratory Medicine and Pathology, University of Washington, Seattle, Washington, USA

^114^Department of Pathology, Boston University, Boston, Massachusetts, USA

^115^Department of Neuropsychology, University of California San Francisco, San Francisco, California, USA

^116^Department of Epidemiology, University of Washington, Seattle, Washington, USA

^117^Department of Neurobiology and Behavior, University of California Irvine, Irvine, California, USA

^118^Department of Neurology, Emory University, Atlanta, Georgia, USA

^119^Kaiser Permanente Washington Health Research Institute, Seattle, Washington, USA

^120^Department of Pathology, University of Michigan, Ann Arbor, Michigan, USA

^121^National Center for PTSD at Boston VA Healthcare System, Boston, Massachusetts, USA

^122^Department of Psychiatry, Boston University Chobanian & Avedisian School of Medicine, Boston, Massachusetts, USA

^123^Department of Psychiatry, Johns Hopkins University, Baltimore, Maryland, USA

^124^Department of Health Management and Policy, School of Public Health, University of North Texas Health Science Center, Fort Worth, Texas, USA

^125^Department of Neurology, University of Alabama at Birmingham, Birmingham, Alabama, USA

^126^Department of Medicine - Pulmonary, New York University, New York, New York, USA

^127^Department of Neurology, Miller School of Medicine, University of Miami, Miami, Florida, USA

^128^Department of Pathology, University of California San Diego, La Jolla, California, USA

^129^Department of Psychiatry, New York University, New York, New York, USA

^130^School of Nursing Northwest Research Group on Aging, University of Washington, Seattle, Washington, USA

^131^Department of Pathology, Boston University Chobanian & Avedisian School of Medicine, Boston, Massachusetts, USA

^132^Department of Pathology, Northwestern University Feinberg School of Medicine, Chicago, Illinois, USA

^133^Cognitive Neurology and Alzheimer’s Disease Center, Northwestern University Feinberg School of Medicine, Chicago, Illinois, USA

^134^Weill Institute for Neurosciences, Memory and Aging Center, University of California San Francisco, San Francisco, California, USA

^135^Department of Pathology, University of Southern California, Los Angeles, California, USA

^136^Department of Pathology, Stanford University School of Medicine, Stanford, California, USA

^137^Department of Pathology and Laboratory Medicine and Alzheimer’s Disease Research Center, University of California Irvine, Irvine, California, USA

^138^Department of Neurology, Washington University, St. Louis, Missouri, USA

^139^Department of Psychiatry, Washington University School of Medicine, St. Louis Missouri, USA

^140^Department of Cell Biology, Miller School of Medicine, University of Miami, Miami, Florida, USA

^141^Department of Neurology, University of Texas Southwestern Medical Center, Dallas, Texas, USA

^142^Department of Research Regulatory Compliance, College of Medicine, Howard Unviersity, Washington, DC, USA

^143^Institute for Translational Research, University of North Texas Health Science Center, Fort Worth, Texas, USA

^144^Center for Mind and Brain and Department of Neurology, University of California Davis, Sacramento, California, USA

^145^Department of Family and Community Medicine, University of Texas Health Science Center San Antonio, San Antonio, Texas, USA

^146^Department of Laboratory Medicine and Pathology, Mayo Clinic, Rochester, Minnesota, USA

^147^Department of Psychiatry and Behavioral Sciences, University of Washington School of Medicine, Seattle, Washington, USA

^148^Pacific Health Research & Education Institute, Veterans Affairs Pacific Islands Healthcare System, Honolulu, Hawaii, USA

^149^Department of Neurology, University of Colorado School of Medicine, Aurora, Colorado, USA

^150^Department of Internal Medicine and Geriatrics, University of North Texas Health Science Center, Fort Worth, Texas, USA

^151^Department of Medical Education, TCU/UNTHSC School of Medicine, Fort Worth, Texas

^152^Arizona Alzheimer’s Consortium, Phoenix, Arizona, USA

^153^Banner Alzheimer’s Institute, Phoenix, Arizona, USA

^154^Department of Psychiatry, University of Arizona, Phoenix, Arizona, USA

^155^Alzheimer’s Disease Center, New York University, New York, New York, USA

^156^Department of Biostatistics, O’Donnell School of Public Health, the University of Texas Southwestern Medical Center, Dallas, TX USA

^157^Department of Neurology, University of California Los Angeles, Los Angeles, California, USA

^158^Tanz Centre for Research in Neurodegenerative Disease, University of Toronto, Toronto, Ontario, Canada

^159^Departments of Psychiatry, Medicine, Family and Community Medicine, and the Glenn Biggs Institute for Alzheimer’s and Neurodegenerative Diseases, UT Health Science Center at San Antonio, San Antonio, Texas, USA

^160^Department of Neurology, Barrow Neurological Institute St. Joseph’s Hospital and Medical Center, Phoenix, Arizona, USA

^161^Department of Medical Genetics, University of British Columbia, Vancouver, Canada

^162^Department of Radiology and Imaging Sciences, Indiana University, Indianapolis, Indiana, USA

^163^Department of Pathology (Neuropathology), Rush University Medical Center, Chicago, Illinois, USA

^164^Department of Psychiatry, University of Southern California, Los Angeles, California, USA

^165^Cambridge Institute for Medical Research, University of Cambridge, Cambridge, England, UK

^166^Faculty of Medicine, Department of Medicine (Neurology), University of Toronto, Toronto, Ontario, Canada

^167^Maya Angelou Center for Health Equity, Wake Forest University School of Medicine, Winston-Salem, North Carolina, USA; Center for Outreach in Alzheimer’s, Aging and Community Health at North Carolina A&T State University, Greensboro, North Carolina, USA

^168^Center for Applied Health Research, Baylor Scott & White Health, Temple, Texas

^169^Center for Population Health and Aging, Texas A&M University Health Science Center, Lubbock Texas, USA

^170^College of Medicine, Texas A&M University Health Science Center, College Station, Texas, USA

^171^Program in Cellular Neuroscience, Neurodegeneration and Repair, Yale University School of Medicine, New Haven, Connecticut, USA

^172^Department of Psychiatry and Human Behavior, and Institute for Memory Impairments and Neurological Disorders (UCI-MIND), University of California Irvine, Irvine, California, USA

^173^Renaissance Computing Institute, University of North Carolina Chapel Hill, Chapel Hill, North Carolina, USA

^174^Department of Pathology, Johns Hopkins University, Baltimore, Maryland, USA

^175^Department of Neurology, Aristotle University of Thessaloniki, Thessaloniki, Macedonia

^176^Department of Neuroscience, College of Medicine, University of Kentucky, Lexington, Kentucky, USA ^177^Department of Pathology and Laboratory Medicine, University of California Los Angeles, Los Angeles, California, USA

^178^Department of Psychiatry and Behavioral Sciences, Northwestern University Feinberg School of Medicine, Chicago, Illinois, USA

^179^Department of Psychiatry and Behavioral Sciences, Duke University, Durham, North Carolina, USA

^180^Department of Medicine, Duke University, Durham, North Carolina, USA

^181^Department of Biostatistics, University of Washington, Seattle, Washington, USA

^182^Department of Genetics, University of North Carolina Chapel Hill, Chapel Hill, North Carolina, USA

^183^Department of Neurology, Section of Gerontology and Geriatric Medicine Research, Wake Forest School of Medicine, Winston-Salem, North Carolina, USA

^184^Department of Psychiatry, New York University Grossman School of Medicine, New York, New York, USA

^185^Center for Cognitive Neurology and Departments of Neurology and Pathology, New York University Grossman School of Medicine, New York, USA

^186^Department of Pathology, Oregon Health and Science University, Portland, Oregon, USA

^187^National Institute of Neurological Disorders and Stroke, National Institutes of Health, Bethesda, MD, USA

^188^Department of Pathology, Case Western Reserve University, Cleveland, Ohio, USA

^189^Department of Ophthalmology, Boston University Chobanian & Avedisian School of Medicine, Boston, Massachusetts, USA

^190^Department of Epidemiology, Columbia University, New York, New York, USA

^191^Social Sciences & Health Policy, Wake Forest School of Medicine, Winston-Salem, North Carolina,

^192^Department of Biostatistics, University of Kentucky, Lexington, Kentucky, USA

^193^Department of Epidemiology, Boston University School of Public Health, Boston, Massachusetts, USA

## CHARGE

Bernard Fongang^1,2,3^, Amber Yaqub^4^, Muralidharan Sargurupremraj^1,3^, Xueqiu Jian^1,3^, Aniket Mishra^5^, Joshua C Bis^6^, Monica Gireud-Goss^1^, Jayandra Jung Himali^1,7,8,9^, Habil Zare^1^, Vilmundur Guðnason^10,11^, Lenore Launer^12^, Jan Bressler^13^, Hans J. Grabe^14,15^, M. Arfan Ikram^4^, Bruce M Psaty^6,16,17^, W T. Longstreth^16,18^, Sigurdur Sigurdsson^11^, Mohsen Ghanbari^4^, Franck J. Wolters^4^, Eric Boerwinkle^13,19^, Alexa S Beiser^7,8,9^, Chloe Sarnowski^20^, Thomas H. Mosley^21^, Oscar L Lopez^22^, Cornelia van Duijn^4,23,24^, Claudia Satizabal^1,3^, M. Kamran Ikram^4,25^, Yang Qiong^9^, Carole Dufouil^5,26^, Stéphanie Debette^5,27^, Myriam Fornage^28,13^, Sudha Seshadri^1,7,29,8^

^1^The Glenn Biggs Institute for Alzheimer’s and Neurodegenerative Diseases, The University of Texas Health Science Center at San Antonio, San Antonio, Texas, USA

^2^Department of Biochemistry and Structural Biology, The University of Texas Health Science Center at San Antonio, San Antonio, Texas, USA

^3^Department of Population Health Sciences, University of Texas Health Science Center, San Antonio, TX, USA

^4^Department of Epidemiology, Erasmus MC, University Medical Center, Rotterdam, the Netherlands ^5^University of Bordeaux, Inserm, Bordeaux Population Health Research Center, UMR 1219, F-33000 Bordeaux, France

^6^Cardiovascular Health Research Unit, Department of Medicine, University of Washington, Seattle, WA, USA

^7^Framingham Heart Study, Framingham, MA, USA

^8^Department of Neurology, Boston University School of Medicine, Boston, Massachusetts, USA ^9^Department of Biostatistics, Boston University School of Public Health, Boston, MA, USA ^10^Faculty of Medicine, University of Iceland, Reykjavik, Iceland

^11^Icelandic Heart Association, Kopavogur, Iceland

^12^Laboratory of Epidemiology and Population Sciences, Intramural Research Program, National Institute of Aging, National Institutes of Health, Bethesda, MD, USA

^13^Human Genetics Center, School of Public Health, The University of Texas Health Science Center at Houston, Houston TX, USA

^14^Department of Psychiatry and Psychotherapy, University Medicine Greifswald, Germany

^15^German Center for Neurodegenerative Diseases (DZNE), Site Rostock/ Greifswald, Rostock, Germany

^16^Department of Epidemiology, University of Washington, Seattle, WA, USA

^17^Department of Health Systems and Population Health, University of Washington, Seattle, WA, USA

^18^Department of Neurology, University of Washington, Seattle, WA, USA

^19^Human Genome Sequencing Center, Baylor College of Medicine, Houston, TX, USA

^20^Department of Epidemiology, Human Genetics and Environmental Sciences, University of Texas Health Science Center at Houston, School of Public Health, Houston, TX

^21^Memory Impairment and Neurodegenerative Dementia (MIND) Center and Department of Medicine, University of Mississippi Medical Center, Jackson, MS, USA

^22^Department of Neurology, School of Medicine, University of Pittsburgh, PA, USA

^23^Nuffield Department of Population Health, University of Oxford, Old Road Campus, Headington, Oxford, OX3 7LF, UK

^24^Big Data Institute, Li Ka Shing Centre for Health Information and Discovery, Old Road Campus, Headington, Oxford, OX3 7LF, UK

^25^Department of Neurology, Erasmus University Medical Centre, Rotterdam, Netherlands

^26^Pôle de Santé Publique Centre Hospitalier Universitaire (CHU) de Bordeaux, Bordeaux, France

^27^CHU de Bordeaux, Department of Neurology, Institute for Neurodegenerative Diseasese, F-33000 Bordeaux, France

^28^Institute of Molecular Medicine, McGovern Medical School, The University of Texas Health Science Center at Houston, Houston TX, USA

^29^Department of Neurology, UT Health San Antonio, 7703 Floyd Curl Drive, San Antonio, TX, USA

## FinnGen

Sami Heikkinen^1^, Hilkka Soininen^2^, Mikko Hiltunen^1^

^1^Institute of Biomedicine, Faculty of Health Sciences, University of Eastern Finland, Kuopio, Finland

^2^Department of Neurology, Institute of Clinical Medicine, University of Eastern Finland, Finland

## GERAD

Atahualpa Castillo Morales^1^, Rebecca Mahoney^1^, Rebecca Sims^2^, Nicola Denning^1^, Alun Meggy^1^, Rachel Marshall^2^, Danielle LeRoux^2^, Catherine Bresner^2^, Julie Williams^1,2^, Peter A Holmans^2^, Valentina Escott-Price^1,2^, Kevin Morgan^3^, Keeley Brookes^4^, Tamar Guetta-Baranes^5^, Clive Holmes^6^, Gill Windle^7,8^, Vanessa Burholt^9,10^, Emma Green^11^, Catherine Macleod^7^, Bob Woods^7^, Simon Mead^12^, Jonathan M Schott^13^, Nick Fox^13^, Patrick G Kehoe^14^, Seth Love^14^

^1^UKDRI@ Cardiff, School of Medicine, Cardiff University, Cardiff, UK

^2^Division of Psychological Medicine and Clinical Neuroscience, School of Medicine, Cardiff University, Cardiff, UK

^3^Human Genetics, School of Life Sciences, University of Nottingham, UK NG7 2UH

^4^Nottingham Trent

^5^Human Genetics. UoN

^6^Clinical and Experimental Science, Faculty of Medicine, University of Southampton, Southampton, UK.

^7^School of Health Sciences, Bangor University, UK

^8^Wales Centre for Ageing & Dementia Research

^9^Faculty of Medical & Health Sciences, University of Auckland, New Zealand ^10^Faculty of Medicine, Health and Life Science, Swansea University ^11^Institute of Public Health, University of Cambridge, UK

^12^MRC Prion Unit at UCL, UCL Institute of Prion Diseases, London, W1W 7FF

^13^Dementia Research Centre, UCL, London, WC1N 3BG

^14^Translational Health Sciences, Bristol Medical School, University of Bristol, Bristol, BS16 1LE, UK

## GR@ACE/DEGESCO

Itziar de Rojas^1,2^, Pablo García-González^1^, Clàudia Olivé^1^, Raquel Puerta^1^, Laura Montrreal^1^, M.Victoria Fernández^1^, Marta Marquié^1,2^, Amanda Cano^1^, Sergi Valero^1,2^, Oscar Sotolongo-Grau^1^, Alba Pérez-Cordón^1^, Ana Espinosa^1,2^, Ángela Sanabria^1,2^, Gemma Ortega^1,2^, Maitée Rosende-Roca^1,2^, Montserrat Alegret^1,2^, Lluís Tárraga^1,2^, Mercè Boada^1,2^, María Eugenia Sáez^3^, Inés Quintela^4^, Ángel Carracedo^5,6^, Luis M Real^7,8,9^, Juan Macías^10^, Anaïs Corma-Gómez^10^, Juan A Pineda^10^, Jose María García-Alberca^11,2^, Silvia Mendoza^12^, Jose Luis Royo^13^, Guillermo Garcia-Ribas^14^, Sebastián García-Madrona^14^, Pablo Mir^2,15^, Emilio Franco-Macías^16,2^, Dolores Buiza-Rueda^15,2^, María Bernal Sánchez-Arjona^16^, Gerard Piñol-Ripoll^17,18^, Raquel Huerto Vilas^17,18^, Alfonso Arias Pastor^17,18^, Pau Pastor^19,20^, Mónica Diez-Fairen^21,22^, Ignacio Alvarez^21,22^, Eloy Rodriguez-Rodriguez^23,2^, Carmen Lage^23,2^, Oriol Dols-Icardo^24,2^, Daniel Alcolea^25,2^, Juan Fortea^25,2^, Alberto Lleó^25,2^, Jordi Pérez-Tur^26,2^, María J. Bullido^27,2,28,29^, Ana Frank-García^30,2,31,29^, Angel Martín Montes^32,2,31^, Raquel Sánchez-Valle^33^, Anna Antonell^33^, Laura Molina-Porcel^34,33^, Victoria Álvarez^35,36^, Manuel Menéndez-González^37,36,38^, Adolfo Lopez de Munain^39,40,2,41^, Fermin Moreno^39,2,41^, Miguel Medina^2^, Pascual Sánchez-Juan^42,2^, Miguel Calero^43,2,44^, Alberto Rábano^42,2^, Ana Belén Pastor^42^, Teodoro del Ser^42^, Florentino Sanchez-Garcia^45^, Carmen Muñoz-Fernandez^45^, M. Candida Deniz-Naranjo^45^, Agustín Ruiz^1,2,46^

^1^Ace Alzheimer Center Barcelona, Universitat Internacional de Catalunya (UIC), Barcelona, Spain.

^2^Networking Research Center on Neurodegenerative Diseases (CIBERNED), Instituto de Salud Carlos III, Madrid, Spain.

^3^CAEBI, Centro Andaluz de Estudios Bioinformáticos, Sevilla, Spain.

^4^Grupo de Medicina Xenómica, Fundación Pública Galega de Medicina Xenómica, Santiago de Compostela, Spain.

^5^Grupo de Medicina Xenómica, Centro de Investigación Biomédica en Red de Enfermedades Raras (CIBERER), Universidade de Santiago de Compostela (CIMUS), Santiago de Compostela, Spain.

^6^Fundación Pública Galega de Medicina Xenómica-Instituto de Investigación Sanitaria de Santiago (IDIS), Santiago de Compostela, Spain.

^7^Instituto de Biomedicina de Sevilla (IBIS), Universidad de Sevilla. Hospital Universitario Virgen de Valme. CIBERINFEC

^8^Depatamento de Bioquímica Médica, Biología Molecular e Inmunología. Facultad de Medicina. Universidad de Sevilla. Sevilla, Spain

^9^Centro de Investigación Biomédica en Red de Enfermedades Infecciosas (CIBERINFEC) Instituto de Salud Carlos III, Madrid. Spain

^10^Unidad Clínica de Enfermedades Infecciosas y Microbiología. Hospital Universitario de Valme, Sevilla, Spain

^11^Alzheimer Research Center & Memory Clinic, Andalusian Institute for Neuroscience, Málaga, Spain ^12^Alzheimer Research Center & Memory Clinic, Instituto Andaluz de Neurociencia, Málaga, Spain ^13^Depatamento de Especialidades Quirúrgicas, Bioquímica e Inmunología. Facultad de Medicina. Universidad de Málaga. Málaga, Spain

^14^Hospital Universitario Ramon y Cajal, IRYCIS, Madrid

^15^Unidad de Trastornos del Movimiento, Servicio de Neurología y Neurofisiología. Instituto de Biomedicina de Sevilla (IBiS), Hospital Universitario Virgen del Rocío/CSIC/Universidad de Sevilla, Seville, Spain

^16^Unidad de Demencias, Servicio de Neurología y Neurofisiología. Instituto de Biomedicina de Sevilla (IBiS), Hospital Universitario Virgen del Rocío/CSIC/Universidad de Sevilla, Seville, Spain

^17^Unitat Trastorns Cognitius, Hospital Universitari Santa Maria de Lleida, Lleida, Spain

^18^Institut de Recerca Biomedica de Lleida (IRBLLeida), Lleida, Spain

^19^Unit of Neurodegenerative diseases, Department of Neurology, Hospital Germans Trias i Pujol, Badalona, Barcelona

^20^Neurodegenerative Diseases Research Laboratory, Germans Trias i Pujol Research Laboratory, Badalona, Barcelona

^21^Fundació Docència i Recerca MútuaTerrassa, Terrassa, Barcelona, Spain

^22^Memory Disorders Unit, Department of Neurology, Hospital Universitari Mutua de Terrassa, Terrassa, Barcelona, Spain

^23^Neurology Service, Marqués de Valdecilla University Hospital (University of Cantabria and IDIVAL), Santander, Spain.

^24^Genetics of Neurodegenerative Diseases Unit IIB Sant Pau, Barcelona, Spain

^25^Department of Neurology, II B Sant Pau, Hospital de la Santa Creu i Sant Pau, Universitat Autònoma de Barcelona, Barcelona, Spain.

^26^Unitat de Genètica Molecular, Institut de Biomedicina de València-CSIC, Valencia, Spain

^27^Centro de Biología Molecular Severo Ochoa (UAM-CSIC)

^28^Instituto de Investigacion Sanitaria ‘Hospital la Paz’ (IdIPaz), Madrid, Spain

^29^Universidad Autónoma de Madrid

^30^Department of Neurology, La Paz University Hospital. Instituto de Investigación Sanitaria del Hospital Universitario La Paz. IdiPAZ.

^31^Hospital La Paz Institute for Health Research, IdiPAZ, Madrid, Spain

^32^Department of Neurology, La Paz University Hospital

^33^Alzheimer’s disease and other cognitive disorders unit. Service of Neurology, Hospital Clínic of Barcelona. Fundació de Recerca Clínic Barcelona-Institut d’Investigacions Biomèdiques August Pi i Sunyer. University of Barcelona, Barcelona, Spain

^34^Neurological Tissue Bank of the Biobanc-Hospital Clinic-IDIBAPS, Institut d’Investigacions Biomèdiques August Pi i Sunyer, Barcelona, Spain

^35^Laboratorio de Genética. Hospital Universitario Central de Asturias, Oviedo, Spain

^36^Instituto de Investigación Sanitaria del Principado de Asturias (ISPA)

^37^Servicio de Neurología. Hospital Universitario Central de Asturias, Oviedo, Spain ^38^Departamento de Medicina, Universidad de Oviedo, Oviedo, Spain ^39^Department of Neurology. Hospital Universitario Donostia. San Sebastian, Spain

^40^Department of Neurosciences. Faculty of Medicine and Nursery. University of the Basque Country, San Sebastián, Spain

^41^Neurosciences Area. Instituto Biodonostia. San Sebastian, Spain

^42^Reina Sofia Alzheimer Center, CIEN Foundation, ISCIII, Madrid, Spain

^43^CIEN Foundation/Queen Sofia Foundation Alzheimer Center/Instituto de Salud Carlos III

^44^UFIEC, Instituto de Salud Carlos III

^45^Department of Immunology, Hospital Universitario Doctor Negrín, Las Palmas de Gran Canaria, Las Palmas, Spain.

^46^Biggs Institute for Alzheimer’s and Neurodegenerative Diseases, University of Texas Health Science Center, San Antonio, Texas, USA.

## PGC-ALZ

Danielle Posthuma^1,2^, Ole A. Andreassen^3^, Douglas P. Wightman^1^, Emil Uffelmann^1^, Hreinn Stefansson^4^, G. Bragi Walters^4^, Kari Stefansson^4^, Jon Snaedal^5^, Helga Eyjólfsdóttir^5^, Nancy L. Pedersen^6^, Chandra A. Reynolds^7^, Ida K. Karlsson^6^, Sara Hägg^6^, Anna Zettergren^8^, Ingmar Skoog^9,10^, Silke Kern^9,10^, Margda Waern^9,11^, Kaj Blennow^12,13,14,15^, Henrik Zetterberg^12,13,16,17,18,19^, Elisa Moreno^20^, Marta Riise Moksnes^20^, Kristian Hveem^20,21,22^, Bendik S Winsvold^20,23,24^, Ben Brumpton^20,21,25^, Geir Selbæk^26,27,28^, Tormod Fladby^29,30^, Dag Aarsland^31,32^, Srdjan Djurovic^33,3^, Arvid Rongve^34,35^, Shahram Bahrami^3^, Alexey A Shadrin^3^, Ingvild Saltvedt^36,37^, Geir Bråthen^38,39^

^1^Department of Complex Trait Genetics, Center for Neurogenomics and Cognitive Research, Amsterdam Neuroscience, Vrije Universiteit Amsterdam

^2^Department of Child and Adolescent Psychiatry and Pediatric Psychology, Section Complex Trait Genetics, Amsterdam Neuroscience, Vrije Universiteit Medical Center, Amsterdam University Medical Center, Amsterdam, The Netherlands

^3^Centre for Precision Psychiatry, Division of Mental Health and Addiction, University of Oslo, and Oslo University Hospital, Oslo, Norway

^4^Amgen deCODE genetics, Sturlugata 8, 102, Reykjavík, Iceland

^5^Department of Geriatric Medicine, Landspitali University Hospital, Reykjavik, Iceland

^6^Department of Medical Epidemiology and Biostatistics, Karolinska Institutet, Stockholm, Sweden ^7^University of Colorado Boulder, Institute for Behavior Genetics and Department of Psychology and Neuroscience, Boulder, Colorado, USA

^8^Neuropsychiatric Epidemiology Unit, Department of Psychiatry and Neurochemistry, Institute of Neuroscience and Physiology, Sahlgrenska Academy, Centre for Ageing and Health (AgeCap) at the University of Gothenburg, Sweden.

^9^Neuropsychiatric Epidemiology Unit, Department of Psychiatry and Neurochemistry, Institute of Neuroscience and Physiology, Sahlgrenska Academy, Centre for Ageing and Health (AgeCap) at the University of Gothenburg, Sweden

^10^Region Västra Götaland, Sahlgrenska University Hospital, Neuropsychiatry Clinic, Gothenburg, Sweden ^11^Region Västra Götaland, Sahlgrenska University Hospital, Psychiatry, Psychosis Clinic, Gothenburg, Sweden

^12^Clinical Neurochemistry Laboratory, Sahlgrenska University Hospital, Mölndal, Sweden

^13^Department of Psychiatry and Neurochemistry, Institute of Neuroscience and Physiology, Sahlgrenska Academy at the University of Gothenburg, Sweden

^14^Paris Brain Institute, ICM, Pitié-Salpêtrière Hospital, Sorbonne University, Paris, France ^15^Neurodegenerative Disorder Research Center, Division of Life Sciences and Medicine, and Department of Neurology, Institute on Aging and Brain Disorders, University of Science and Technology of China and First Affiliated Hospital of USTC, Hefei, P.R. China

^16^Department of Neurodegenerative Disease, UCL Institute of Neurology, Queen Square, London, UK

^17^UK Dementia Research Institute at UCL, London, UK

^18^Hong Kong Center for Neurodegenerative Diseases, Clear Water Bay, Hong Kong, China

^19^Wisconsin Alzheimer’s Disease Research Center, University of Wisconsin School of Medicine and Public Health, University of Wisconsin-Madison, Madison, WI, USA

^20^HUNT Center for Molecular and Clinical Epidemiology, Department of Public Health and Nursing, NTNU, Norwegian University of Science and Technology, Trondheim 7030, Norway

^21^HUNT Research Centre, Department of Public Health and Nursing, NTNU, Norwegian University of Science and Technology, Levanger 7600, Norway

^22^Department of Research, St. Olavs Hospital, Trondheim University Hospital, Trondheim, Norway ^23^Department of Research and Innovation, Division of Clinical Neuroscience, Oslo University Hospital, Oslo, Norway

^24^Department of Neurology, Oslo University Hospital, Oslo, Norway

^25^Clinic of Medicine, St. Olavs Hospital, Trondheim University Hospital, Trondheim 7030, Norway

^26^Norwegian National Centre for Ageing and Health, Vestfold Hospital Trust, Tønsberg, Norway

^27^Institute for Clinical Medicine, University of Oslo, Oslo, Norway

^28^Department of Geriatric Medicine, Oslo University Hospital, Oslo, Norway

^29^Department of Neurology, Akershus University Hospital, Lørenskog, Norway

^30^Institute of Clinical Medicine, University of Oslo, Oslo, Norway

^31^Centre of Age-Related Medicine, Stavanger University Hospital, Norway

^32^Institute of Psychiatry, Psychology and Neurosciences, King’s College London

^33^Department of Medical Genetics, Oslo University Hospital, Oslo, Norway

^34^Department of Research and Innovation, Helse Fonna, Haugesund, Norway

^35^Department of Clinical Medicine 1 (K1), University of Bergen, Bergen, Norway

^36^Department of Neuromedicine and Movement Science (INB), NTNU, Faculty of Medicine and Health Sciences, Trondheim, Norway

^37^Department of Geriatric Medicine, Clinic of Medicine, St. Olavs Hospital, Trondheim University Hospital, Trondheim, Norway

^38^Department of Neurology, St Olav’s Hospital, Trondheim University Hospital, Trondheim, Norway ^39^Department of Neuromedicine and Movement Science, Faculty of Medicine and Health Sciences, Norwegian University of Science and Technology (NTNU), Trondheim, Norway

## Author contributions

**EADB, EADI and Bonn. Project coordination:** C.B. **Consortium coordination:** V.G., K.M., M.V.F., P.G.K., M.T., C.v.D., R.F-S., R.G., P.S-J., K.S., M.I., M.Hil., R.S., W.v.d.F., O.A., A.Rui., A.Ram., J-C.L. **Analysis team:** C.B., A.C., N.A., S.J.v.d.L., M.M., K.P., F.K., B.G-B., S.H., I.d.R., A.N., M.C.D., L.Kle., J.L.B., S.P. **Sample contribution:** S.J.v.d.L., O.P., A.Sch., M.D., D.R., N.Sch., J.D., S.R-H., L.H., L.M.P., E.D., T.G., J.Wilt., S.H-H., S.Moe., M.Sch., T.T., N.Sca., O.D-I., F.M., J.P-T., M.J.B., P.P., R.S-V., V.Á., M.B., P.G-G., R.P., P.Mir., L.M.R., G.P-R., J.M.G-A., E.R-R., H.Soi., A.d.M., S.Meh., J.H., M.V., N.San., J.L., J.Q.T., Y.A.P., H.H., H.See., I.Ram., J.Pap., M.Hul., G-J.B., C.G., H.T., A.U., G.P., V.G., M.L., L.Kil., J.Will., P.H., P.A., A.B., J-F.D., G.N., C.D., F.P., O.H., S.D., E.G., J.Pop., D.G., B.A., P.Mec., V.S., L.P., A.Squ., L.T., B.B., M.W., B.N., M.Spa., D.S., I.Rai., A.D., P.B., C.M., G.R., F.J., K.M., M.V.F., P.G.K., M.T., C.v.D., R.F-S., R.G., P.S-J., K.S., M.I., M.Hil., R.S., W.v.d.F., O.A., A.Rui., A.Ram., J-C.L. **Writing group**: J-C.L, C.B.

**CHARGE. Coordination:** B.F., J.C.B, M.G-G., V.G., L.L., E.B., T.H.M., C.D., S.D., M.F., S.Seshadri. **Data analyses:** B.F., A.Y., M.S., X.J., A.M., J.C.B, J.J.H., H.Z., J.B., S.Sigurdsson, M.G., A.S.B., C.Sarnowski, C.v.D., C.Satizabal, Y.Q. **Sample contribution:** V.G., L.L., H.J.G., M.A.I., B.M.P., W.T.L., F.J.W., E.B., T.H.M., O.L.L., M.K.I., C.D., S.D., M.F., S.Seshadri. **Writing group:** B.F., M.S., X.J., J.C.B, C.D., S.D., M.F., S.Seshadri.

**ADGC. Study Design and Conception:** A.C.N., F.R., N.K., C.Z., W.-P.L., N.R.W., Y.Z., J.J.F., Y.Y.L., R.M.S., T.I., O.V., K.L.L., B.W.K., L.-S.W., L.A.F., J.L.H., R.M., M.A.P.-V., G.D.S. **Sample Contribution:** G.T., J.B.M., Y.E.S., X.Z., E.A., A.A., M.S.A., R.L.A., M.A., L.G.A., S.E.A., S.Asthana, C.S.A., C.T.B., R.C.B., L.L.B., T.G.B., J.T.B., G.W.B., D.Beekly, D.A.Bennett, J.B., T.D.B., D.Blacker, B.F.B., J.D.B., A.Boxer, J.B.B., J.R.B., J.M.B., J.D.Buxbaum, N.J.C., L.B.C., C.Cao, C.S.C., C.M.C., R.M.C., M.M.C., M.-F.C., N.A.C., H.C.C., J.C., S.Craft, P.K.C., D.H.C., E.A.C., C.Cruchaga, M.L.C., M.C., E.D., B.D., P.L.D., C.D., J.C.D., M.Dick, D.W.D., B.A.D., R.S.D., R.D., N.E.-T., D.A.E., K.M.F., T.J.F., K.B.F., D.W.F., M.R.F., T.M.F., M.P.F., D.R.G., M.G., D.H.G., B.G., J.R.G., A.M.G., T.G., N.R.G.-R., N.E.G., J.H.G., H.H., J.H., R.L.H., O.Harari, J.Hardy, E.H., V.H., M.H., L.S.H., R.M.H., M.J.H., B.T.H., L.S.Hynan, L.I., G.P.J., S.J., L.W.J., K.J., L.J., M.I.K., M.J.K., J.S.K., J.A.K., C.D.K., Khaleeq, J.K., N.W.K., J.H.K., W.K., F.M.L., J.J.L., E.B.L., A.L., J.B.L., A.I.L., A.P.L., R.B.L., M.L., O.L.L., C.G.L., D.M., D.C.M., E.R.M., F.M., D.C.Mash, E.M., P.M., A.M., W.C.M., S.M.M., A.C.M., M.Mesulam, B.L.M., C.A.M., J.W.M., T.J.M., E.S.M., J.C.M., S.Mukherjee, A.J.M., T.N., S.O., J.M.O., R.P., J.E.P., H.L.P., V.P., D.P., V.Perez, E.P., R.C.P., M.P., W.W.P., H.P., L.Q., M.Q., J.F.Q., A.R., M.R., E.M.R., B.R., J.S.R., J.M.R., E.D.R., M.Rodriguear, E.R., H.J.R., R.N.R., D.R.R., M.A.S., M.S., A.J.S., J.A.S., L.S.S., W.W.S., S.H.S., S.Small, A.G.S., J.A.Sonnen, P.S.G.-H., R.A.S., S.M.S., D.S., R.H.S., R.E.T., J.T., J.C.T., D.W.T., V.M.VD., L.J.VE., J.M.V., R.V., H.V.V., J.-P.V., S.W., K.A.W.-B., P.L.W., E.M.W., K.C.W., B.W., J.W., H.W., T.S.W., T.W., R.L.W., C.B.W., C.-K.W. S.G.Y., C.-E.Y., L.Y., X.Zhou. **Data Generation:** A.C.N., F.R., J.S., C.Z., W.-P.L., J.Haut, K.H.-N., N.R.W., Y.Z., J.J.F., D.L., R.M.S., O.V., A.B.K., L.S.Hynan, C.D.K. **Analysis.** A.C.N., P.B., L.M.P.S., Q.Q., N.K., S.A.C., Y.K., F.R., C.R., G.R.J., J.S., K.B., C.Z., W.-P.L., J.Haut, K.H.-N., N.R.W., Y.Z., J.J.F., M.A.G., Y.Y.L., P.P.K., D.L., E.L.daF., E.L.P., J.P., R.M.S., T.I., O.V., A.B.K., S.B., B.F.H., T.J.H., K.L.L., B.N.V., B.W.K., W.B., L.-S.W., L.A.F., J.L.H., G.D.S. **Manuscript Preparation:** A.C.N., F.R., N.K., J.S., K.B., C.Z., N.R.W., B.W.K., L.-S.W., L.A.F., J.L.H., M.A.P.-V., G.D.S. **Study Supervision/Management:** A.C.N., F.R., L.A.F., J.L.H., M.A.P.-V., G.D.S.

**FinnGen. Coordination:** M.H. **Data analyses:** S.H. **Sample contribution:** M.H., H.S.

**GERAD. Sample collection:** R.S., N.D., A.M., R.Marshall, D.L., C.B., J.W., K.M., K.B., T.G-B., C.H., G.W., V.B., E.G., C.M., B.W., S.M., J.M.S., N.F., P.G.K., S.L., O.S. **Data analysis.** A.C.M., R.Mahoney, R.S., J.W. **Data production:** A.C.M., R.S., C.B., J.W., P.H., V.E-P. **Supervision:** R.S., S.M.

**GR@ACE/DEGESCO. Data analyses:** I.d.R., P.G-G. **Coordination:** A.R., P.S-J. **Sample contribution:** all GR@ACE/DEGESCO authors.

**PGC-ALZ. Coordination:** D.P., O.A.A., D.W., E.U., B.B., S.B., A.S. **Data analyses:** D.W., H.S., G.B.W., K.S., J.S., H.E., E.M., M.R.M., S.B., A.S. **Sample contribution:** D.P., O.A.A., H.S., B.W., K.S., J.S., H.E., N.P., C.R., I.K., S.H., A.Z., I.Skoog, S.K., M.W., K.B., H.Z., K.H., B.S.W., B.B., G.S., T.F., D.A., S.D., A.R., I.Saltvedt, G.B.

## Disclaimer

The views expressed in this manuscript are those of the authors, and do not represent those of the US Government.

