## Supplementary Text for "Consensus meta-analysis of genome-wide association studies for Alzheimer’s disease and related dementia"

### 1. Sample description and data processing

#### 1.1. European Alzheimer & Dementia Biobank consortium

##### *1.1.1. Study descriptions*

###### *a) European Alzheimer & Dementia Biobank core dataset (EADB-core)*

This consortium groups together 20,464 Alzheimer's disease (AD) cases and 22,244 controls after quality controls from 16 European countries (Austria, Belgium, Bulgaria, Czech Republic, Denmark, Finland, France, Germany, Greece, Italy, Portugal, Spain, Sweden, Switzerland, The Netherlands and the UK). These samples were genotyped in three independent centers (France, Germany and the Netherlands) leading to define three nodes: EADB-France, EADB-Germany and EADB-Netherlands. In addition, EADB also included Australian partners.

**EADB-France.** In the France node, samples were collected from nine countries (39 centers/studies), and after quality controls (QCs), we obtained 13,867 AD cases and 15,310 controls. All these samples were genotyped at the Centre National de Recherche en Génomique Humaine (CNRGH, Evry, France).

<u>Belgium:</u> The participants were part of a large prospective cohort[1] of Belgian AD patients and healthy elderly control individuals. The patients were ascertained at the memory clinic of Middelheim and Hoge Beuken (Hospital Network Antwerp, Belgium) and at the memory clinic of the University Hospitals of Leuven, Belgium. The control individuals were the partners of the patients or volunteers from the Belgian community. The study protocols were approved by the ethics committees of the Antwerp University Hospital and the participating neurological centers at the different hospitals of the BELNEU consortium and by the University of Antwerp.

<u>Czech Republic:</u> The Czech Brain Aging Study (CBAS)[2] is a longitudinal memory-clinic–based study recruiting subjects at risk of dementia (subjects referred for cognitive complaints-SCD, MCI). The CBAS+ study is a cross-sectional study of patients in the early stages of dementia. All subjects signed informed consent and both studies were approved by the local ethics committee.

<u>Denmark:</u> The Copenhagen General Population Study (CGPS) is a prospective study of the Danish general population initiated in 2003 and still recruiting. Individuals were selected randomly based on the national Danish Civil Registration System to reflect the adult Danish population aged 20-100. Data were obtained from a self-administered questionnaire reviewed together with an investigator at the day of attendance, a physical examination, and from blood samples including DNA extraction.

<u>Finland:</u> *The ADGEN cohort*[3]: clinic-based collection of AD patients from Eastern and Northern Finland examined in the Department of Neurology in Kuopio University Hospital and the Department of Neurology in Oulu University Hospital. All the patients were diagnosed with probable AD according to the criteria of the National Institute of Neurological and Communicative Disorders and Stroke and the Alzheimer's disease and Related Disorders Association (NINCDS–ADRDA). The study was approved by the ethics committee of Kuopio University Hospital, Finland (420/2016). *The FINGER study*[4]*:* a Finnish multi-domain lifestyle RCT enrolling 1,260 older adults with an increased risk of dementia from the general population. The intensive

lifestyle intervention lasted for two years, and follow-up extends currently up to seven years. The FINGER study was approved by the coordinating ethics committee of the Hospital District of Helsinki and Uusimaa (94/13/03/00/2009 and HUS/1204/2017), and all the participants gave written informed consent.

France: *The BALTAZAR multicenter (23 memory centers) prospective study*[5]: 1,040 participants from September 2010 to April 2015. They were classified as AD cases (n = 501) according to DSM IV-TR and NINCDS–ADRDA criteria as well as amnestic mild cognitive impairment (MCI) cases (a MCI, n = 417) and non-amnestic MCI cases (na MCI, n = 122) according to Petersen's criteria. A comprehensive battery of cognitive tests was performed, including MMSE, verbal fluency, and FCSRT. All the participants or their legal guardians gave written informed consent. The study was approved by the Paris ethics committee (CPP Ile de France IV Saint Louis Hospital). *MEMENTO*: a clinic-based study[6] aimed at better understanding the natural history of AD, dementia, and related diseases. Between 2011 and 2014, 2,323 individuals presenting either recently diagnosed MCI or isolated cognitive complaints were enrolled in 26 memory centers in France. This study was performed in accordance with the guidelines of the Declaration of Helsinki. The MEMENTO study protocol has been approved by the local ethics committee (Comité de Protection des Personnes Sud-Ouest et Outre Mer III; approval number 2010-A01394-35). All the participants provided written informed consent. *The CNRMAJ-Rouen study*[7]*:* early onset AD patients (n = 870). The patients or their legal guardians provided written informed consent. This study was approved by the ethics committee of CPP Ile de France II.

Italy: The AD cases and controls were collated through Italy in different centers: Brescia, Cagliari, Florence, Milan, Rome, Pertugia, San Giovani Rotondo and Torino. AD cases were diagnosed according to DSM III-R,IV and NINCDS–ADRDA criteria. Controls were defined a minima as subjects without DMS-III-R dementia criteria and with integrity of their cognitive functions (MMS>25).

Spain: The Dementia Genetic Spanish Consortium (DEGESCO) is a national consortium comprising 23 research centers and hospitals across the country, that holds the institutional coverage of The Network Center for Biomedical Research in Neurodegenerative Diseases (CIBERNED). Created in 2013, DEGESCO's objective is the promotion and conduction of genetic studies aimed at understanding the genetic architecture of neurodegenerative dementias in the Spanish population and participates in coordinated actions in national and international frameworks. All DNA samples are in compliance with the Law of Biomedical Research (Law 14/2007) and the Royal Decree on Biobanks (RD 1716/2011). Patients included in the present study met clinical criteria for probable or possible disease established by the National Institute of Neurological and Communication Disorders and Stroke and the Alzheimer Disease and Related Disorders Association (NINCDS-ADRDA). Cognitively healthy controls were unrelated individuals who had a documented MMSE in the normal range. Contributing centers in the France node genotyping were Centro de Biología Molecular Severo Ochoa (CSIC-UAM (Madrid), the Institute Biodonostia, University of Basque Contry (EHU-UPV, San Sebastián), Institut de Biomedicina de Valencia CSIC (València), and Sant Pau Biomedical Research Institute (Barcelona).

Sweden: *Uppsala*. The Swedish AD patients were ascertained at the Memory Disorder Unit at Uppsala University Hospital. For all patients, the diagnosis was established according to the National Institute on Neurological Disorders and Stroke, and the Alzheimer's Disease and Related Disorders Association (NINDS-ADRDA) guidelines[8]. Healthy control subjects were recruited from the same geographic region following advertisements in local newspapers and displayed no signs of dementia upon Mini Mental State

Examination (MMSE). *Karolinska Institutet*, *Stockholm*. AD patients were ascertained at the Memory Clinic at Karolinska University Hospital - Huddinge, Stockholm Sweden. For all patients, the diagnosis was established according to the National Institute on Neurological Disorders and Stroke, and the Alzheimer's Disease and Related Disorders Association (NINDS-ADRDA) guidelines[8] and if available supported by CSF beta-amyloid and phospho-tau biomarker analyses. As controls, DNA from participants in the *Swedish National Study on Aging and Care in Kungsholmen (SNAC-K)* data was collected. The original SNAC-K population consisted of 4590 living and eligible persons who lived on the island of Kungsholmen in Central Stockholm, belonged to pre-specified age strata, and were randomly selected to take part in the study. Between 2001 and 2004, 3363 persons participated in the baseline assessment. They belonged to the age cohorts 60, 66, 72, 78, 81, 84, 87, 90, 93, and 96 years and 99 years and older. The examination consists of three parts: a nurse interview, a medical examination, and a neuropsychological testing session. Altogether, the examination takes about six hours. The participants are reexamined each time they reach the next age cohort. All parts of the SNAC-K project have been approved by the ethical committee at Karolinska Institutet or the regional ethical review board. Informed consent was collected from all the participants or, if the person was severely cognitively impaired, from their next of kin.

<u>The UK</u>: *MRC*. The sample set comprises individuals with AD and healthy controls recruited across the MRC Centre for Neuropsychiatric Genetics and Genomics, Cardiff University, Cardiff, UK; Institute of Psychiatry, London, UK; University of Cambridge, Cambridge, UK. The collection of the samples was through multiple channels, including specialist NHS services and clinics, research registers and Join Dementia Research (JDR) platform. The participants were assessed at home or in research clinics along with an informant, usually a spouse, family member or close friend, who provided information about and on behalf of the individual with dementia. Established measures were used to ascertain the disease severity: Bristol activities of daily living (BADL), Clinical Dementia Rating scale (CDR), Neuropsychiatric Inventory (NPI) and Global Deterioration Scale (GlDS). Individuals with dementia completed the Addenbrooke's Cognitive Examination (ACE-r), Geriatric Depression Scale (GeDS) and National Adult Reading Test (NART) too. Control participants were recruited from GP surgeries and by means of self-referral (including existing studies and Joint Dementia Research platform). For all other recruitment, all AD cases met criteria for either probable (NINCDS-ADRDA, DSM-IV) or definite (CERAD) AD. All elderly controls were screened for dementia using the Mini Mental State Examination (MMSE) or ADAS-cog, were determined to be free from dementia at neuropathological examination or had a Braak score of 2.5 or lower. Control samples were chosen to match case samples for age, gender, ethnicity and country of origin. Informed consent was obtained for all study participants, and the relevant independent ethical committees approved study protocols. *SOTON*, *University of Southampton*, *Southampton*, *UK*. All AD cases met criteria for either probable (NINCDS-ADRDA, DSM-IV) or definite (CERAD) AD. All elderly controls were screened for dementia using the MMSE or ADAS-cog, were determined to be free from dementia at neuropathological examination or had a Braak score of 2.5 or lower. *Nottingham and Manchester*, *University of Nottingham*, *Nottingham*, *UK and Manchester Brain Bank*. All AD cases met criteria for either probable (NINCDS-ADRDA, DSM-IV) or definite (CERAD) AD. All elderly controls were screened for dementia using the MMSE or ADAS-cog, were determined to be free from dementia at neuropathological examination or had a Braak score of 2.5 or lower. *KCL*, *London Neurodegenerative Diseases Brain Bank*. All AD cases met criteria for either probable (NINCDS-ADRDA, DSM-IV) or definite (CERAD) AD. All elderly controls were screened for dementia using the MMSE or ADAS-cog, were determined to be free from dementia at neuropathological examination or had a Braak score of 2.5 or lower. *PRION*, All AD cases met criteria for either probable (NINCDS-ADRDA, DSM-IV) or definite (CERAD) AD. All elderly controls were screened for dementia using the MMSE or ADAS-cog, were

determined to be free from dementia at neuropathological examination or had a Braak score of 2.5 or lower. *CFAS Wales*, The Cognitive Function and Ageing Study Wales (CFAS-Wales) is a longitudinal population-based study of people aged 65 years and over in rural and urban areas of Wales that aims to investigate physical and cognitive health in older age and examine the interactions between health, social networks, activity, and participation. Individuals aged 65 years and over were randomly sampled from general medical practice lists between 2011 and 2013, stratified by age to ensure equal numbers in two age groups, 65-74 years and 75 and over. The baseline sample included 3593 older people and included those living in care homes as well as those living at home. Those who provided written consent to join the study were interviewed in their own homes by trained interviewers and could choose to have the interview conducted through the medium of either English or Welsh. Participants were followed up 2 years later. All AD cases met criteria for either probable (NINCDS-ADRDA, DSM-IV) or definite (CERAD) AD. All elderly controls were screened for dementia using the MMSE or CAMCOG, and were determined to be free from dementia. *UCL-DRC*. the UCL Alzheimer's disease cohort of the Dementia Research Centre (UCL - EOAD DRC) included patients seen at the Cognitive Disorders Clinics at The National Hospital for Neurology and Neurosurgery (Queen Square), or affiliated hospitals. Individuals were assessed clinically and diagnosed as having probable Alzheimer's disease based on contemporary clinical criteria in use at the time, including imaging and neuropsychological testing where appropriate.

**EADB-Germany.** In the German node, samples were collected from seven countries (11 centers/studies) and after QCs, we obtained 4,159 AD cases and 4,545 controls. All these samples were genotyped at Life&brain (Bonn, Germany).

Germany: DELCODE (the multicenter DZNE-Longitudinal Cognitive Impairment and Dementia Study). This is an observational longitudinal memory clinic-based multicenter study in Germany comprising 400 subjects with Subjective cognitive decline (SCD), 200 mild cognitive impairment (MCI) patients, 100 AD dementia patients, 200 control subjects without subjective or objective cognitive decline, and 100 first-degree relatives of patients with a documented diagnosis of AD dementia. All patient groups (SCD, MCI, AD) are referrals, including self-referrals, to the participating memory centers. The control group and the relatives of AD dementia patients are recruited by standardized public advertisement. Ten university-based memory centers are participating, all being collaborators of local DZNE sites. All patient groups (SCD, MCI, AD) were assessed clinically at the respective memory centers before entering DELCODE. The assessments include medical history, psychiatric and neurological examination, neuropsychological testing, blood laboratory work-up, cerebrospinal fluid (CSF) biomarkers, and routine MRI, all according to the local standards. The Consortium to Establish a Registry for Alzheimer's Disease (CERAD) neuropsychological test battery was applied at all memory centers to measure cognitive function. German age, sex, and education-adjusted norms of the CERAD neuropsychological battery are available online (www.memoryclinic.ch). Detail description is of recruitment protocol is reported elsewhere. *The VOGEL study*: The VOGEL study is a prospective, observational, long-term follow-up study with three time points of investigation within 6–8 years. This cohort includes dementia and healthy subjects. Residents of the city of Würzburg born between 1936 and 1941 were recruited. Every participant underwent physical, psychiatric, and laboratory examinations and performed intense neuropsychological testing as well as VSEP and NIRS according to the published procedures. A total of 604 subjects were included. *The Heidelberg/Mannheim memory clinic sample*: This cohort includes 61 subjects from whom 40 MCI patients were recruited and assessed between 2012 and 2016. Some of those patients converted to dementia by AD or other dementias. *The PAGES study:*

This study includes 301 subjects. AD patients were recruited at the memory clinic of the Department of Psychiatry, University of Munich, Germany. Participants in whom dementia associated with AD was diagnosed fulfilled the criteria for probable AD according to the NINCDS–ADRDA. The control group included participants who were randomly selected from the general population of Munich. Controls who had central nervous system diseases or psychotic disorders or who had first-degree relatives with psychotic disorders were excluded. *The Technische Universität München study:* This cohort includes 359 healthy, AD, and other dementias patients recruited from the Centre for Cognitive Disorders. All the participants provided written informed consent. A biobank was submitted to the ethics committee of the Technical University of Munich, School of Medicine (Munich, Germany), which raised no objections and approved the biobank (reference number 347-14). *The Göttingen Universität study:* This study includes 111 in- and outpatients with a healthy or AD dementia status from the Department of Psychiatry of the University of Göttingen. The study's ethical statement was provided locally at the Göttingen University Medical Centre. *The German Dementia Competence Network (DCN) cohort:* Individuals from the DCN cohort were recruited from 14 university hospital memory clinics across Germany between 2003 and 2005[9]. The study was approved by the respective ethics committees, and written informed consent was obtained from all the participants prior to inclusion. *The German Study on Aging, Cognition, and Dementia (AgeCoDe):* The AgeCoDe study is a general practice (GP) registry-based longitudinal study in elderly individuals that recruited patients aged 75 years and above in six German cities from 2003 to 2004[10]. The study was approved by the respective ethics committees, and written informed consent was obtained from all the participants prior to inclusion.

<u>Greece</u>: *the HELIAD study*, comprising 49 AD cases and 1,150 controls. HELIAD is a population-based, multidisciplinary, collaborative study designed to estimate, in the Greek population over the age of 64 years, the prevalence and incidence of MCI, AD, other forms of dementia, and other neuropsychiatric conditions of aging and to investigate associations between nutrition and cognitive dysfunction or age-related neuropsychiatric diseases. The participants were selected through random sampling from the records of two Greek municipalities, Larissa and Marousi. All the participants signed informed consent in Greek.

<u>Portugal:</u> *the Lisbon study* from Portugal, totaling 78 AD cases and 74 controls. This cohort was recruited in 2008–2009 to investigate the connections between oxidative stress and lipid dyshomeostasis in AD. The project includes 190 subjects and was approved by the local ethics committee, and all the participants provided written informed consent. This study includes healthy and dementia-by-AD subjects.

<u>Spain:</u> Those samples are part of DEGESCO. DEGESCO Centers from whom DNA samples were genotyped in the German node (1,778 cases and 470 controls) were the Alzheimer Research Center and Memory Clinic, Fundació ACE, Institut Català de Neurociències Aplicades (Barcelona), the Neurology Service at University Hospital Marqués de Valdecilla (Santander), the Alzheimer's disease and other cognitive disorders, Neurology Department, at Hospital Clínic, IDIBAPS (Barcelona), the Molecular Genetics Laboratory, at the Hospital Universitario Central de Asturias (Oviedo), and Fundació Docència i Recerca Mútua de Terrassa and Movement Disorders Unit, Department of Neurology, University Hospital Mútua de Terrassa (Barcelona).

<u>Switzerland/Austria:</u> Two datasets from Switzerland and Austria were combined, totaling 182 AD cases and 388 controls. *The Lausanne study*: This study includes 137 community-dwelling participants aged 55+ years

with cognitive impairment (memory clinic patients with MCI, dementia) or normal cognition (recruited by advertisement, word of mouth). The study's ethical statement was provided locally at the Department of Psychiatry, Geneva University Centre, Switzerland. *The VITA study:* This is a longitudinal study of 606 individuals (Vienna, Austria) who were 75 years old in 2000, followed up every 30–90 months. This cohort includes dementia and healthy subjects. All the participants gave written informed consent. The study conformed to the latest version of the Declaration of Helsinki and was approved by the ethics committee of the City of Vienna, Austria

**EADB-Netherlands.** In the Dutch node, samples were collected from six organizations in the Netherlands and after QCs, we obtained 2,438 AD cases and 2,389 controls. All these samples were genotyped at the Erasmus Medical University (Rotterdam, The Netherlands). The Medical Ethics Committee (METC) of the local institutes approved the studies. All the participants and/or their legal guardians gave written informed consent for participation in the clinical and genetic studies. Samples from the following institutes were included. 1) *Erasmus Medical Center:* most individuals were selected from population studies from the epidemiology department and accounted for most of the controls, while a smaller subset of samples originated from the neurology department, where AD was diagnosed according to the National Institute of Neurological and Communicative Disorders and Stroke-Alzheimer's Disease and Related Disorders Association (NINCDS-ADRDA) criteria for AD[11]. 2) *The Amsterdam Dementia Cohort (ADC)*[12]*:* This cohort comprises patients who visit the memory clinic of the VU University Medical Centre, the Netherlands. The diagnosis of probable AD is based on the clinical criteria formulated by the NINCDS–ADRDA and based on the NIA–AA. Diagnosis of MCI was made according to Petersen and NIA-AA. Controls presented with subjective cognitive decline at the memory clinic, but performed within normal limits on all clinical investigations. 3) *The 100-Plus study:* This study includes Dutch-speaking individuals who (i) can provide official evidence for being aged 100 years or older, (ii) self-report to be cognitively healthy, which is confirmed by a proxy, (iii) consent to the donation of a blood sample, (iv) consent to (at least) two home visits from a researcher, and (v) consent to undergo an interview and neuropsychological test battery[13]. 4) *Parelsnoer Institute:* a collaboration between 8 Dutch University Medical Centers in which clinical data and biomaterials from patients suffering from chronic diseases (so called "Pearls") are collected according to harmonized protocols. The Pearl Neurodegenerative Diseases[14] includes individuals diagnosed with dementia, mild cognitive impairment, and controls with subjective memory complaints. 5) *The Netherlands Brain Bank:* a non-profit organization that collects human brain tissue of donors with a variety of neurological and psychiatric disorders, but also of non-diseased donors. A clinical diagnosis of AD is based on the clinical criteria of probable AD[8,15]. The selected AD patients for this study all received a definitive diagnosis which was based on autopsy. 6) *Maastricht University Medical Center*: a subset of individuals that were referred to the memory clinic for cognitive complaints were included if they participated in the BioBank-Alzheimer Centrum Limburg (BB-ACL)[16]. Diagnosis of MCI was made according to the criteria of Petersen, and diagnosis of AD-type dementia was made according to the criteria of the DSM-4[17], and the NINCDS-ADRDA[8].

**EADB-Australia.** The Sydney MAS study: a longitudinal study investigating MCI, related syndromes, and age-related cognitive change. Older adults (70–90 years old) were randomly recruited from the community in Sydney, Australia (n = 1,037). An extensive interview was undertaken and questionnaire data collected, including demographics, cognitive performance, and medical history. The majority of participants provided blood samples for genetic analysis. Neuroimaging was performed on a subset of participants. Ethics approval for the study was provided by the ethics committee of the University of New South Wales and the

Illawarra Area Health Service Human Research Ethics Committee. All the participants provided written informed consent to join the study. More information is provided in Sachdev et al[18]. In our study, there were 43 AD cases and 215 controls. Due to the low sample size, the study was not considered in the meta-analysis. However, samples from Sydney MAS study were included in the evaluation of the association of the polygenic risk score with conversion to all-dementia and AD dementia.

**Genotyping.** EADB genomic DNA samples were transferred to 3 genotyping centers and samples that passed the DNA QC were genotyped with the Illumina Infinium Global Screening Array. Raw probe intensities were shared with the CNRGH, which performed the genotype calling on all samples using the same custom cluster file. During the genotyping QC process, three genotyping batches were considered: (1) 49 genotyping chips were identified as possibly problematic and thus were considered as a separate batch, (2) a batch of samples was genotyped and processed after all other samples and (3) the main batch including all other samples. Further details are available in Bellenguez et al.[19].

##### *b) GR@ACE/DEGESCO*

The GR@ACE study[20] recruited Alzheimer's disease (AD) patients from Fundació ACE, Institut Català de Neurociències Aplicades (Catalonia, Spain), and control individuals from three centers: Fundació ACE (Barcelona, Spain), Valme University Hospital (Seville, Spain), and the Spanish National DNA Bank–Carlos III (University of Salamanca, Spain) (http://www.bancoadn.org). Additional cases and controls were obtained from dementia cohorts included in the Dementia Genetics Spanish Consortium (DEGESCO)[21]. At all sites, AD diagnosis was established by a multidisciplinary working group—including neurologists, neuropsychologists, and social workers—according to the DSM-IV criteria for dementia and the National Institute on Aging and Alzheimer's Association's (NIA–AA) 2011 guidelines for diagnosing AD. In our study, we considered as AD cases any individuals with dementia diagnosed with probable or possible AD at any point in their clinical course.

Genotyping was conducted using the Axiom 815K Spanish biobank array (Thermo Fisher) at the Spanish National Centre for Genotyping (CeGEN, Santiago de Compostela, Spain). The genotyping array not only is an adaptation of the Axiom biobank genotyping array but also contains rare population-specific variations observed in the Spanish population.

##### *c) European Alzheimer's Disease Initiative (EADI) Consortium*

EADI is composed of several case-control studies and one population-based cohort, the 3C study[22,23]. Case-control studies are comprised of AD cases and cognitively normal controls across France. The population-based cohort, the 3C study, is a prospective study of the relationship between vascular factors and dementia carried out in the three French cities Bordeaux, Montpellier and Dijon. The AD status was defined based on 12 years follow-up for Dijon participants, 14-15 years follow-up for Montpellier participants and 17-18 years follow-up for Bordeaux participants. All other non demented subjects of 3C were included as controls. All AD cases, both in the case-control studies and the 3C study, were ascertained by neurologists and the clinical diagnosis of probable AD was established according to the DSM-III-R and NINCDS-ADRDA criteria. Samples that passed DNA quality control were genotyped with Illumina Human 610-Quad BeadChips.

***d) Genetic and Environmental Risk in AD (GERAD) Consortium/Defining Genetic, Polygenic, and Environmental Risk for Alzheimer's Disease (PERADES) Consortium***

The GERAD/PERADES sample comprises 3,177 Alzheimer's disease cases and 7,277 controls with available age and gender data[24]. Cases and elderly screened controls were recruited by the Medical Research Council (MRC) Genetic Resource for Alzheimer's disease (Cardiff University; Institute of Psychiatry, London; Cambridge University; Trinity College Dublin), the Alzheimer's Research Trust (ART) Collaboration (University of Nottingham; University of Manchester; University of Southampton; University of Bristol; Queen's University Belfast; the Oxford Project to Investigate Memory and Ageing (OPTIMA), Oxford University); Washington University, St Louis, United States; MRC PRION Unit, University College London; London and the South East Region Alzheimer's disease project (LASER-AD), University College London; Competence Network of Dementia (CND) and Department of Psychiatry, University of Bonn, Germany; the National Institute of Mental Health (NIMH) Alzheimer's disease Genetics Initiative. 6129 population controls were drawn from large existing cohorts with available GWAS data, including the 1958 British Birth Cohort (1958BC) (http://www.b58cgene.sgul.ac.uk), the KORA F4 Study and the Heinz Nixdorf Recall Study. All Alzheimer's disease cases met criteria for either probable (NINCDS-ADRDA, DSM-IV) or definite (CERAD) Alzheimer's disease. All elderly controls were screened for dementia using the MMSE or ADAS-cog, were determined to be free from dementia at neuropathological examination or had a Braak score of 2.5 or lower. Genotypes from all cases and 4617 controls were previously included in the AD GWAS by Harold and colleagues[24]. Genotypes for the remaining 2660 population controls were obtained from WTCCC2.

***e) The Norwegian DemGene Network***

This is a Norwegian network of clinical sites collecting cases from memory clinics based on a standardized examination of cognitive, functional, and behavioral measures and data on the progression of most patients. The Norwegian DemGene Network includes 2,224 cases and 3,089 healthy controls from different studies described elsewhere[25]. The cases were diagnosed according to recommendations from the NIA–AA, the NINCDS–ADRDA criteria, or the ICD-10 research criteria. The controls were screened with a standardized interview and cognitive tests. Additional controls from blood donors of the Oslo University Hospital, Ulleval Hospital, were included ($n$=4992, age between 18-65 years, 48% female). They were thoroughly screened for diseases and medication, and provided blood for DNA analysis, in line with approval from the Regional Committee for Medical and Health Research Ethics. Individuals from the DemGene study and blood donors were genotyped using either the Human Omni Express-24 v1.1 chip (Illumina Inc., San Diego, CA) or the DeCodeGenetics_V1_20012591_A1 chip at deCODE Genetics (Reykjavik, Iceland).

***f) Bonn studies***

<u>DietBB</u>: The DietBB sample included in this GWAS is a subsample extracted from the AgeCoDe cohort in the context of an ongoing genome-wide methylation analysis for dementia. In addition to methylation, the DietBB samples has genome-wide genotype data which was included in this study. The German study on aging, cognition and dementia (AgeCoDe)[10,26] study is a general practice (GP) registry-based longitudinal study in elderly individuals on the identification of predictors of dementia. Participants were recruited in six German cities (Bonn, Dusseldorf, Hamburg, Leipzig, Mannheim, and Munich) with a total of 138 GPs connected to the study sites. The inclusion criteria for this study were an age of 75 years and older, absence of dementia according to GP judgment, and at least one contact with the GP within the past 12 months. Exclusion criteria were GP consultations by home visits only, living in a nursing home, severe illness with an

anticipated fatal outcome within 3 months, language barrier, deafness or blindness, and lack of ability to provide informed consent. Baseline recruitment was performed in 2002 and 2003. The study was approved by the local ethical committees of the Universities of Bonn, Hamburg, Dusseldorf, Heidelberg/Mannheim, and Leipzig, and the Technical University of Munich. A total of 3327 subjects provided informed consent for participation after being provided with a complete description of the study protocol. The study assessments were performed by trained interviewers at the subjects' home. Seventy individuals were excluded after baseline interview because of the presence of dementia according to standard assessment, and 40 subjects were excluded for age less than 75 years. In AgeCoDe, dementia was diagnosed according to the criteria set of DSM-IV in a consensus conference with the interviewer and an experienced geriatrician or geriatric psychiatrist. The etiological diagnosis of dementia in AD was established according to the National Institute of Neurological and Communicative Diseases and Stroke/Alzheimer's Disease and Related Disorders Association (NINCDS-ADRDA) criteria for probable AD[8]. Mixed dementia was diagnosed in cases of cerebrovascular events without temporal relationship to cognitive decline. Mixed dementia and dementia in AD were combined. Dementia diagnosis in subjects who were not interviewed personally was based on the Global Deterioration Scale[27] (score ≥4 points). In these cases, an etiological diagnosis was established only if the information provided was sufficient to judge etiology according to the criteria just described. For DietBB, cohort participants where included if they were dementia-free at baseline and available biomaterial for DNA analysis is available. This criterion led to the selection of 320 participants. In 120 of these participants, dementia of the AD-type occurred at any follow up. The additional 200 remain free of dementia until last follow up of AgeCoDe.

Bonn OMNI cohort: the Bonn OMNI cohort consists of AD patients and controls derived from a larger German GWAS cohort which was recruited from the following three sources: (i) the German Dementia Competence Network; (ii) the German study on Aging, Cognition, and Dementia in primary care patients (AgeCoDe); and (iii) the interdisciplinary Memory Clinic at the University Hospital of Bonn. The control sample comprised of individuals from the population-based study Heinz Nixdorf Recall (HNR) study cohort. This sample was previously used for replication in Lambert et al.[28]. *The German study on aging, cognition and dementia (AgeCoDe)*: see description above. *The German competence network cohort (DCN)*: The DCN cohort includes 1,095 patients with mild cognitive impairment (MCI) and 648 cases with mild Alzheimer's disease (AD) clinical dementia syndrome that were recruited from 14 university hospital memory clinics across Germany between 2003 and 2005[9]. Exclusion criteria were substance abuse or dependence, insufficient German language skills, multi-morbidity, comorbid condition with excess mortality, circumstances that would have made regular attendance at follow-up visits questionable and lack of an informant. The diagnosis of mild dementia according to ICD-10 criteria required a decline of cognitive ability (at least 1 SD) from a previous level in at least 2 domains as evidenced by age-corrected standardized tests, impairment in activities of daily living (i.e. B-ADL > 6), changes in personality, drive, social behavior or control of emotion but no clouding of consciousness. These changes must have persisted for at least 3 months. The etiological diagnosis of AD was assigned according to NINCDS-ADRDA criteria[8]. *Memory clinic Bonn*: The interdisciplinary Memory Clinic of the Department of Psychiatry and Department of Neurology at the University Hospital in Bonn provided further patients. Diagnoses were assigned according the NINCDS/ADRDA criteria[8] and on the basis of clinical history, physical examination, neuropsychological testing (using the CERAD neuropsychological battery, including the MMSE), laboratory assessments, and brain imaging. *Control sample:* In the Heinz Nixdorf Recall (Risk Factors, Evaluation of Coronary Calcification, and Lifestyle) study, participants were randomly sampled in three cities in Germany. The study design has previously been described[29,30]. Briefly, 4814 participants aged 45 to 75 years were enrolled between 2000 and 2003 (t0, baseline). Cognitive performance of participants was evaluated at follow up scheduled 5 years after baseline (t1, n = 4157, 2005–2008) and then again at follow up 5 years after t1 (t2, n = 3087, 2010–2015). Controls sample was selected if participant did not present cognitive impairment as reported at the last available evaluation. Cognitive evaluation has been described extensively previously[31,32]. Herein, cognitive impairment at t1 was defined as a performance of one standard deviation (SD) below the age- and education-adjusted mean except for the clock-drawing test, where a performance ≥3 was rated as impaired (for a detailed description, see the study by Winkler et al.[31]. The study was approved by the University of Duisburg-Essen Institutional Review Board and followed established guidelines of good epidemiological practice.

###### *1.1.2. Data processing*

Data processing of the EADB studies is described in detail in Bellenguez et al.[19]. A brief description is provided below.

*Quality control.* We applied the same QC protocol to all the EADB studies: EADB-core, EADI, GR@ACE/DEGESCO, GERAD, Bonn and DemGene. This QC consisted of assessment of chip's variants, variant intensity QC and autosomal sample QC (exclusion of individuals with high heterozygosity or missingness on the autosomes, of individuals with discordant genetic and clinical sex, of population outliers and of related individuals). Relatedness was inferred within studies, and across each pair of the following studies: Bonn, EADB-core, EADI, DemGene and GERAD.

*TOPMed imputation.* All samples and variants passing the QC were used as the input of the imputation process. The imputation was performed by the Michigan Imputation Server[33] where the TOPMed Freeze5 reference panel was granted to the EADB consortium. The server version used was the 1.2.4 with Eagle v2.4[34] as the phasing software and Minimac4 v4-1.0.2 as the imputation software. Due to the limitation in terms of maximum number of samples per job, the EADB-core samples were split into 5 batches. After the imputation process of all batches, in order to have a global imputation quality, a merged imputation quality was recomputed including all samples using the bcftools impute_info plugin.

*HRC imputation.* A subset of the EADB core samples were imputed with the HRC reference panel and sent to the Sanger Imputation server (https://imputation.sanger.ac.uk/) where the HRC reference panel r1.1 was used. The data were phased using the Eagle v2.0.5 software and the imputation process was performed by PBWT v3.1.

*Association tests.* After QC of the genotyping data, we additionally excluded controls with age below 30 and individuals with known pathogenic mutations. Tests of the association between AD status and autosomal genetic variants were conducted in each dataset by using logistic regression and an additive genetic model, as implemented in SNPTEST 2.5.4-beta3[35]. We analyzed the genotype probabilities using the newml method. Analyses were adjusted for principal components and genotyping centers, when necessary (Supplementary Table 22). The principal components used as adjustment in the analysis were computed using the FlashPCA v2.0 software.

##### 1.2. Alzheimer's Disease Genetics Consortium (ADGC)

Across all datasets used, AD cases and cognitively-normal elders (CNEs) were defined by either (a) pathological confirmation of the presence (cases) or absence (CNEs) of AD pathology or (b) clinical diagnosis using NIA-AA Working Group criteria (cases) or confirmation of normal cognitive status (CNEs) via cognitive evaluation (e.g., mini-mental status exam (MMSE) or modified MMSE (3MS) score ≥ 26). No samples classified as having mild cognitive impairment (MCI) were used in analyses. Details of the ascertainment of individual datasets are described below. Consistency of diagnostic standards across studies was evaluated and confirmed by the ADGC's Clinical Adjudication Board. All subjects were recruited under protocols approved by the appropriate Institutional Review Boards (IRBs).

###### *1.2.1. Study descriptions*

The ADGC dataset comprises subjects from 35 datasets including the Adult Changes in Thought (ACT) cohort study (ACT1); ten waves of cases and cognitively normal controls from the National Institute on Aging (NIA) Alzheimer Disease Centers (ADCs) and coordinated by the National Alzheimer's Coordinating Center (NACC); the Alzheimer Disease Neuroimaging Initiative (ADNI); the Biomarkers of Cognitive Decline Among Normal Individuals (BIOCARD) Cohort; two waves of the Religious Orders Study/Memory and Aging Project (ROSMAP1-2) and the Chicago Health and Aging Project (CHAP) cohort studies at Rush University; the Einstein Aging Study (EAS); the Multi-Site Collaborative Study for Genotype-Phenotype Associations in Alzheimer's Disease (GenADA) Study by GlaxoSmithKline; Mayo Clinic Jacksonville (MAYO) and Rochester (RMAYO) case-control datasets; the Multi-Institutional Research in Alzheimer's Genetic Epidemiology (MIRAGE) study; the NIA Late-Onset Alzheimer's Disease (LOAD) Family Study (NIA-LOAD); the Oregon Health and Science University (OHSU) case-control dataset; the Pfizer case-control dataset; the Texas Alzheimer's Research and Care Consortium (TARCC) dataset; the Translational Genomics Research Institute series 2 (TGEN2) dataset; the University of Miami (UM)/Case Western Reserve University (CWRU)/Mt. Sinai School of Medicine (MSSM) and UM/CWRU/TARCC wave 2 datasets [UM/CWRU/MSSM and UM/CWRU/TARCC2]; the Universitatsklinikum Saarlandes (UKS) case-control dataset; the University of Pittsburgh (UPITT) case-control dataset; Washington University (WASHU) wave 1 and 2 case-control datasets [WASHU1/WASHU2]; and the Washington Heights-Inwood Community Aging Project (WHICAP) study datasets.

Descriptions of the ACT1, ADC waves 1-7, ADNI, BIOCARD, CHAP, EAS, GenADA, MAYO, MIRAGE, NIA-LOAD, OHSU, PFIZER, RMAYO, ROSMAP1, ROSMAP2, TARCC, TGEN2, UKS, UM/CWRU/MSSM, UM/CWRU/TARCC2, UPITT, WASHU1, WASHU2, and WHICAP cohorts were provided in previous ADGC and IGAP studies[28,36–40]. Here we update descriptions of these studies, where applicable, and provide descriptions for ADC waves 8-12, as well as the Combined Small Datasets Collection (CSDC). The CSDC is a harmonized collection of merged small datasets (comprising the existing datasets of BIOCARD, CHAP2, EAS, RMAYO, ROSMAP2, and WASHU2) used in common variant analyses to deal with small case and control counts within the individual studies. All analyses were restricted to individuals of European ancestry.

<u>ACT1/ACT2:</u> The ACT cohort is an urban and suburban elderly population from a stable HMO that includes 2,581 cognitively intact subjects age ≥ 65 years who were enrolled between 1994 and 1998[41,42]. An additional 811 subjects were enrolled in 2000-2002 using the same methods except oversampling clinics with more minorities. All clinical data are reviewed at a consensus conference. Dementia onset is assigned half-way between the prior biennial and the exam that diagnosed dementia. A waiver of consent was obtained from the IRB to enroll deceased ACT participants. In total, ACT contributed data on 532 individuals with probable or possible Alzheimer's disease (70 with autopsy-confirmation) and on 1,571 cognitively normal elders (CNEs; 155 with autopsy-confirmation) who were included in the analyses.

<u>NIA ADC Samples (ADC1-12/NACC)</u>: The NIA ADC cohort included subjects ascertained and evaluated by the clinical and neuropathology cores of the 32 NIA-funded ADCs. Data collection is coordinated by the

National Alzheimer's Coordinating Center (NACC). NACC coordinates collection of phenotype data from the 32 ADCs, cleans all data, coordinates implementation of definitions of Alzheimer's disease cases and controls, and coordinates collection of samples. The ADC cohort consists of 3,311 autopsy-confirmed and 5,656 clinically-confirmed Alzheimer's disease cases, and 247 cognitively normal elders (CNEs) with complete neuropathology data who were older than 60 years at age of death, and 3,687 living CNEs evaluated using the Uniform dataset (UDS) protocol[43,44] who were documented to not have mild cognitive impairment (MCI) and were between 60 and 100 years of age at assessment. Based on the data collected by NACC, the ADGC Neuropathology Core Leaders Subcommittee derived inclusion and exclusion criteria for Alzheimer's disease and control samples. All autopsied subjects were age ≥ 60 years at death. Based on the data collected by NACC, the ADGC Neuropathology Core Leaders Subcommittee derived inclusion and exclusion criteria for Alzheimer's disease and control samples. Alzheimer's disease cases were classified as demented according to NINCDS-ADRDA/DSMIV-V[8] or more recent NIA-AA criteria[11] or Clinical Dementia Rating (CDR) ≥ 1. Neuropathologic stratification of cases followed NIA/Reagan criteria explicitly or used a similar approach when NIA/Reagan criteria were coded as not done, missing, or unknown[45,46]. Cases were intermediate or high likelihood by NIA/Reagan criteria with moderate to frequent amyloid plaques[47] and neurofibrillary tangle (NFT) Braak stage of III-VI[48,49]. Persons with Down's syndrome, non-Alzheimer's disease tauopathies and synucleinopathies were excluded. All autopsied controls had a clinical evaluation within two years of death. Controls did not meet NINCDS-ADRDA/DSMIV-V criteria for dementia, did not have a diagnosis of mild cognitive impairment (MCI), and had a CDR of 0, if performed. Controls did not meet or were low-likelihood Alzheimer's disease by NIA/Reagan criteria, had sparse or no amyloid plaques, and a Braak NFT stage of 0 – II. ADCs sent frozen tissue from autopsied subjects and DNA samples from some autopsied subjects and from living subjects to the ADCs to the National Cell Repository for Alzheimer's Disease (NCRAD). DNA was prepared by NCRAD for genotyping and sent to the genotyping site at Children's Hospital of Philadelphia. ADC samples were genotyped and analyzed in separate batches (waves 1-10). The ADC data used in IGAP discovery analyses (ADC1-12) consist of 6,959 cases and 5,903 CNEs in total.

ADNI: ADNI is a longitudinal, multi-site observational study including Alzheimer's disease, mild cognitive impairment (MCI), and elderly individuals with normal cognition assessing clinical and cognitive measures, MRI and PET scans (FDG and 11C PIB) and blood and CNS biomarkers. For this study, ADNI contributed data on 268 Alzheimer's disease cases with MRI confirmation of Alzheimer's disease diagnosis and 173 healthy controls with Alzheimer's disease-free status confirmed as of most recent follow-up. Alzheimer's disease subjects were between the ages of 55–90, had an MMSE score of 20–26 inclusive, met NINCDS-ADRDA/ NIA-AA criteria for probable Alzheimer's disease[8,11], and had an MRI consistent with the diagnosis of Alzheimer's disease. Control subjects had MMSE scores between 28 and 30 and a Clinical Dementia Rating of 0 without symptoms of depression, MCI or other dementia and no current use of psychoactive medications. According to the ADNI protocol, subjects were ascertained at regular intervals over 3 years, but for the purpose of our analysis we only used the final ascertainment status to classify case-control status. Additional details of the study design are available elsewhere[39,50,51].

BIOCARD (part of CSDC): The BIOCARD study is supported by a grant jointly funded by the National Institute on Aging (NIA) and the National Institute of Mental Health (NIMH). The overarching goal of the BIOCARD Study is to identify biomarkers associated with progression from normal cognitive status to cognitive impairment or dementia, with a particular focus on Alzheimer's Disease. Please see Albert et al. 2014[52] for a detailed description of the study. A total of 354 individuals were initially enrolled in the study. Recruitment was conducted by the staff of the Geriatric Psychiatry Branch (GPB) of the intramural program of the NIMH, beginning in 1995 and ending in 2005. The domains of information collected as part of the study include: cognitive testing, magnetic resonance imaging (MRI), cerebrospinal fluid (CSF), amyloid imaging (using PET-PiB), and blood specimens. Investigators at the Johns Hopkins University School of Medicine began evaluating participants in 2009, and subjects are seen annually. At each visit there are assessments of medical and cognitive status, as well as acquisition of MRI, CSF, PET-PiB, and blood. Each subject in the analyses received a consensus diagnosis by a team of neurologists, neuropsychologists, research nurses and research assistants of the BIOCARD Clinical Core at Johns Hopkins with diagnoses based on evidence of clinical or cognitive dysfunction (i.e., individuals with a CDR score > 0 and/or evidence of decline on cognitive testing). To the extent possible, this diagnosis did not use the cognitive test scores. In brief, (1) clinical data relating to the medical, neurologic and psychiatric status of the subject were examined, (2) reports of changes in cognition by the subject and other sources were examined, and (3) decline in cognitive performance was established. Cognitive test scores were used to: (1) determine whether the subject had become cognitively impaired, and (2) determine the likely etiology of such impairment. These diagnostic procedures are comparable to those implemented in the Alzheimer's Disease Centers (ADC) program, supported by the NIA. For this study, BIOCARD contributed data on 6 Alzheimer's disease cases and 112 healthy controls with Alzheimer's disease-free status confirmed from the most recent follow-up.

CHAP (part of CSDC): CHAP is an on-going, community-based study of individuals from a geographically defined community of 3 neighborhoods in Chicago, Illinois (Morgan Park, Washington Heights, and Beverly), with 6,158 participants in the first phase of the study (78.7% overall; 80.5% of the blacks, 74.6% of the whites)[53]. Data were collected in cycles of approximately 3 years; each consisting of an in-home interview of all participants and clinical evaluation of a random, stratified sample. The baseline cycle measured disease prevalence and provided risk factor data prior to incident disease onset. A cohort of 3,838 persons free of Alzheimer's disease was identified; 729 persons were sampled for baseline clinical evaluation. Persons in the disease-free cohort had either good cognitive function at baseline, or if cognitive function was intermediate or poor, were free from Alzheimer's disease at the baseline clinical evaluation. This disease-free cohort was evaluated for incident disease after an average of 4.1 years. Sampling for incident clinical evaluation was based on age, sex, race, and change in cognitive function (i.e., stable or improved, small decline, or large decline). The sample set available in the ADGC for genetic analyses included 27 Alzheimer's disease cases and 144 persons free of Alzheimer's disease at time of last assessment. All subjects were age 65 years or older at last evaluation.

EAS: (part of CSDC): Based at the Albert Einstein College of Medicine, the EAS is an ongoing community-based cohort study of cognitive aging and Alzheimer's disease in the elderly which began over four decades ago. Please see Barzilai et al. 2004[54] and Katz et al. 2012[55] for details. The EAS cohort has employed systematic recruiting methods to reduce the selection biases that arise from clinic-based samples and to capture the racial diversity within the Bronx community. Since 1993, a total of 1,944 participants have been enrolled. Between 1993 and 2004, Health Care Financing Administration/Centers for Medicaid and Medicare Services (HCFA/CMS) rosters of Medicare eligible persons aged 70 and above were used to develop sampling frames of community residing participants in Bronx County. Since 2004, New York City Board of Elections registered voter lists for the Bronx have been used due to changes in policies for release of HCFA/CMS rosters. Individuals were mailed introductory letters regarding the study and were then telephoned to complete a brief screening interview. Eligible participants were at least 70 years of age, Bronx residents, non-institutionalized, and English speaking. Exclusion criteria included visual or auditory impairments that preclude neuropsychological testing, active psychiatric symptomatology that interfered with the ability to complete assessments, and non-ambulatory status. Written informed consent was obtained at the initial clinic visit. In-person evaluations were completed at baseline and at subsequent 12-month intervals. Functional status was assessed by the self-administered CERAD C1-ALT, a cognitive/functional impairment instrument, and the Instrumental Activities of Daily Living scale (IADL), a subscale on the Lawton Brody Activities of Daily Living Scale. The score on the IADL was based on 5 domains of function that were common to both elderly men and women. Scores for each domain were dichotomized as impaired vs. not impaired and then the domain scores were summed. If the participant agreed, an informant completed the CERAD C2-ALT, a cognitive/functional impairment instrument, and the Informant Questionnaire on Cognitive Decline in the Elderly (IQ-CODE) 14 forms. The standard neurological physical examination was adapted from the Unified Parkinson's Disease Rating Scale. The evaluation assessed the participant's memory for significant recent events in the news and personal events. The coherence and focus of responses, repetitiveness, and language were determined. When possible, informants were interviewed to ascertain whether they noted any cognitive changes in the participant, and to assess accuracy of the participant's responses. The neurologist also assessed each participant for abnormal behaviors, fluctuation in cognition, and history of sleep disturbance and visual/auditory hallucinations. The neurologist assigned an Hachinski Ischemic Score (HIS), the Clinical Dementia Rating (CDR), and provided a clinical impression of presence or absence of dementia. A diagnosis of dementia was based on standardized clinical criteria from the Diagnostic and Statistical Manual, Fourth Edition (DSM-IV) and required impairment in memory plus at least one additional cognitive domain, accompanied by evidence of functional decline. Diagnoses were assigned at consensus case conferences, which included comprehensive review of cognitive test results, relevant neurological signs and symptoms, and functional status. Memory impairment was defined as scores in the impaired range on any of the memory tests in the neuropsychological battery. (FCSRT ≤ 2430 or 1.5 standard deviations (SD) below the age-adjusted mean on Logical Memory) Functional decline was determined at case conference based on information from self or informant report, impairment score on the IADL Lawton Brody Scale, clinical evaluation, and informant questionnaires. Alzheimer's disease was diagnosed in participants with dementia meeting clinical criteria for probable or possible disease established by the NINCDS-ADRDA/ NIA-AA. Incident dementia and Alzheimer's disease were diagnosed in persons free of dementia at baseline who met criteria at follow-up. A subset of individuals who participated in the clinical studies of the EAS came to autopsy, providing an important quality control for diagnostic accuracy. A clinical diagnosis of dementia had a positive predictive value (PPV) of 96% for significant pathology upon autopsy. A clinical diagnosis of possible or probable Alzheimer's disease had a PPV of 79% for the presence of NIA-Reagan intermediate or high likelihood Alzheimer type pathology based on an autopsy sample of 175. For this study, EAS contributed data on 9 Alzheimer's disease cases and 141 healthy controls with Alzheimer's disease-free status confirmed as of most recent follow-up.

GenADA: GenADA study data analyzed included 666 Alzheimer's disease cases and 712 CNEs ascertained from nine memory referral clinics in Canada between 2002 and 2005. Patients and CNEs were of predominantly Europe ancestral genetic background. All patients with Alzheimer's disease satisfied NINCDS-ADRDA/NIA-AA[8,11] and DSM-IV criteria for probable Alzheimer's disease with Global Deterioration Scale scores of 3-7. CNEs had MMSE test scores higher than 25 (mean 29.2 ± 1.1), a Mattis Dementia Rating Scale score of ≥ 136, a Clock Test without error, and no impairments on seven instrumental activities of daily living questions from the Duke Older American Resources and Services Procedures test. Data were collected under an academic-industrial grant from Glaxo-Smith-Kline, Canada by Principal Investigator P. St George-Hyslop. Detailed characteristics of this cohort have been described previously[56].

MAYO/RMAYO (RMAYO part of CSDC): All 671 cases and 1,279 controls consisted of NHW subjects from the United States ascertained at the Mayo Clinic[57]. All subjects were diagnosed by a neurologist at the Mayo Clinic in Jacksonville, Florida (MAYO) or Rochester, Minnesota (RMAYO). The neurologist confirmed a Clinical Dementia Rating score of 0 for all controls; cases had diagnoses of possible or probable Alzheimer's disease made according to NINCDS-ADRDA/NIA-AA criteria[11,57]. Autopsy-confirmed samples (221 cases, 216 CNEs) came from the brain bank at the Mayo Clinic in Jacksonville, FL and were evaluated by a single neuropathologist. The Jacksonville Mayo Clinic contributed 658 cases and 1,046 CNEs, while the Rochester Mayo Clinic contributed 13 cases and 233 CNEs. In clinically-identified cases, the diagnosis of definite Alzheimer's disease was made according to NINCDS-ADRDA criteria. All Alzheimer's disease brains analyzed in the study had a Braak score of 4.0 or greater. Brains employed as controls had a Braak score of 2.5 or lower but often had brain pathology unrelated to Alzheimer's disease and pathological diagnoses that included vascular dementia, frontotemporal dementia, dementia with Lewy bodies, multi-system atrophy, amyotrophic lateral sclerosis, and progressive supranuclear palsy.

MIRAGE: MIRAGE: The MIRAGE study is a family-based genetic epidemiology study of Alzheimer's disease that enrolled Alzheimer's disease cases and unaffected sibling controls at 17 clinical centers in the United States, Canada, Germany, and Greece (details elsewhere[58]), and contributed 1,229 subjects (491 Alzheimer's disease cases and 738 CNEs). For this analysis, a subset of 421 cases and 309 controls that were not related were incorporated into this study[36,39]. Briefly, families were ascertained through a proband meeting the NINCDS-ADRDA/ NIA-AA[8,11] criteria for definite or probable Alzheimer's disease. Unaffected sibling controls were verified as cognitively healthy based on a Modified Telephone Interview of Cognitive Status score ≥ 86[59].

NIA-LOAD: The NIA LOAD Family Study[60] recruited families with two or more affected siblings with LOAD and unrelated, CNEs similar in age and ethnic background. A total of 1,819 cases and 1,969 CNEs from 1,802 families were recruited through the NIA LOAD study, NCRAD, and the University of Kentucky, with 1,798 cases and 1,568 CNEs included for analysis. One case per family was selected after determining the individual with the strictest diagnosis (definite > probable > possible LOAD). If there were multiple individuals with the strictest diagnosis, then the individual with the earliest age of onset was selected. The controls included only those samples that were neurologically evaluated to be normal and were not related to a study participant.

OHSU (part of CSDC): The OHSU dataset includes 132 autopsy-confirmed Alzheimer's disease cases and 153 deceased controls that were evaluated for dementia within 12 months prior to death (age at death > 65 years), which are a subset of the 193 cases and 451 controls examined in our previous study[39] meeting more stringent QC criteria in this study. Subjects were recruited from aging research cohorts at 10 NIA-funded ADC, and did not overlap other samples assembled by the ADGC. A more extensive description of control samples can be found elsewhere[61].

Pfizer: The Pfizer sample collection comprises Alzheimer's disease cases taken from the Lipitor's Effect in Alzheimer's Disease (LEADe) trial, including subjects who converted to Alzheimer's disease after ascertainment as MCI, as well as 216 probable Alzheimer's disease subjects enrolled by PrecisionMed for a case-control study and 149 subjects from a Phase II trial (#A3041005) of CP-457920 (a selective α5 GABAA receptor inverse agonist) in Alzheimer's disease. Samples were collected from multiple clinical sites, and with appropriate IRB/ethics committee approvals at each individual site, with written and informed consent given by subjects for use in follow-up studies. All subjects were diagnosed with probable or possible Alzheimer's disease if they met NINCDS-ADRDA/NIA-AA[8,11] and/or DSM-IV criteria, and had Mini-Mental Status Exam (MMSE) scores < 25 at baseline. The control group included subjects from two studies: 1) the PrecisionMed case-control study (#A9010012), which recruited elderly subjects free of neurological or psychiatric conditions, and 2) 999-GEN-0583-001, which obtained a reference population of cognitively, neurologically, and psychiatrically normal subjects. Controls have no neuropsychiatric conditions or diseases and had MMSE>27 at the time of enrollment. For Alzheimer's disease analysis, all cases with age-at-onset (AAO) less than 65 years were removed to exclude early-onset Alzheimer's disease subjects. All controls were re-matched with remaining cases according to gender, age (all controls are older than cases), and ethnicity (only individuals with NHW background were analyzed). The final Pfizer Alzheimer's disease case-control GWAS dataset included 696 cases and 762 controls. Cases from the PrecisionMed/ A3041005 and LEADe studies and age-matched controls were genotyped using the Illumina HumanHap550 array. *APOE* genotypes were determined from genotypes for rs429358 and rs7412 obtained using Taqman assays.

<u>ROSMAP1/ROSMAP2</u> (ROSMAP2 part of CSDC): ROSMAP are two community-based cohort studies. The ROS has been ongoing since 1993, with a rolling admission. Through July of 2010, 1,139 older nuns, priests, and brothers from across the United States initially free of dementia who agreed to annual clinical evaluation and brain donation at the time of death completed their baseline evaluation. The MAP has been on-going since 1997, also with a rolling admission. Through July of 2010, 1,356 older persons from across northeastern Illinois initially free of dementia who agreed to annual clinical evaluation and organ donation at the time of death completed their baseline evaluation. Details of the clinical and neuropathologic evaluations have been previously reported[62–64]. A total of 1,064 persons passed genotyping QC. Of these, 2885 met clinical criteria for Alzheimer's disease at the time of their last clinical evaluation or time of death and met neuropathologic criteria for Alzheimer's disease for those on whom neuropathologic data were available, and 747 were without dementia or MCI at the time of their last clinical evaluation or time of death and did not meet neuropathologic criteria for Alzheimer's disease for those on whom neuropathologic data were available. A second wave of ROSMAP (referred to as ROSMAP2 in this study) included 59 persons who met clinical criteria for Alzheimer's disease at the time of their last clinical evaluation or time of death and met neuropathologic criteria for Alzheimer's disease for those on whom neuropathologic data were available, and 217 persons who were without dementia or MCI at the time of their last clinical evaluation or time of death and did not meet neuropathologic criteria for Alzheimer's disease for those on whom neuropathologic data were available.

<u>TARCC1/3:</u> The TARCC is a collaborative Alzheimer's research effort directed and funded by the Texas Council on Alzheimer's Disease and Related Disorders (the Council), as part of the Darrell K Royal Texas Alzheimer's Initiative. Composed of Baylor College of Medicine (BCM), Texas Tech University Health Sciences Center (TTUHSC), University of North Texas Health Science Center (UNTHSC), the UT Southwestern Medical Center at Dallas (UTSW), University of Texas Health Science Center at San Antonio (UTHSCSA), Texas A&M Health Science Center (TAMHSC), and the University of Texas at Austin (UTA), this consortium was created to establish a comprehensive research cohort of well characterized subjects to address better diagnosis, treatment, and ultimately prevention of Alzheimer's disease[65]. The resulting prospective cohort, the Texas Harris Alzheimer's Research Study, contains clinical, neuropsychiatric, genetic, and blood biomarker data on more than 3,000 participants diagnosed with Alzheimer's disease, mild cognitive

impairment (MCI), and cognitively normal individuals. Longitudinal data/sample collection and follow-up on participants occurs on an annual basis. Three waves of sample data from TARCC were examined as part of genetic analyses in the ADGC. Data from the TARCC included 323 cases and 181 CNEs in the first wave (included in the TARCC1 cohort); 84 cases and 115 CNEs in the second wave (included in the UM/CWRU/TARCC2 cohort); and an additional 268 cases and 211 CNEs in TARCC3. All TARCC subjects were greater than 65 years of age at disease onset (cases) or at last disease-free exam (non-cases).

TGEN2: Among the TGEN2 data analyzed were 668 clinically- and neuropathologically-characterized brain donors, and 365 CNEs without dementia or significant Alzheimer's disease pathology. Of these cases and CNEs, 667 were genotyped as a part of the TGEN1 series[66]. Samples were obtained from twenty-one different National Institute on Aging-supported Alzheimer's disease Center brain banks and from the Miami Brain Bank as previously described[66–70]. Additional individual samples from other brain banks in the United States, United Kingdom, and the Netherlands were also obtained in the same manner. The criteria for inclusion were as follows: self-defined ethnicity of European descent, neuropathologically confirmed Alzheimer's disease or neuropathology present at levels consistent with status as a control, and age of death greater than 65. Autopsy diagnosis was performed by board-certified neuropathologists and was based on the presence or absence of the characterization of probable or possible Alzheimer's disease. Where possible, Braak staging and/or CERAD classification were employed. Samples derived from subjects with a clinical history of stroke, cerebrovascular disease, comorbidity with any other known neurological disease, or with the neuropathological finding of Lewy bodies were excluded.

UKS: The UKS cohort is a thoroughly diagnosed case-control cohort from Universitatsklinikum Saarlandes, consisting of individuals clinically diagnosed with sporadic Alzheimer's disease (N = 596; age-at-onset [mean ± SD]: 72.2 ± 6.6 years) and cognitively healthy, age-, gender-, and ethnicity-matched population-based controls (N = 170; age-at-exam: 64.1 ± 3.0 years).

UM/CWRU/MSSM: The UM/CWRU/MSSM dataset (formerly UM/VU/MSSM)[71–74] contains 1,177 cases and 1,126 CNEs ascertained at the University of Miami, Case Western Reserve University and Mt. Sinai School of Medicine, including 409 autopsy-confirmed cases and 136 controls, primarily from the Mt. Sinai School of Medicine[75]. An additional 16 cases were included and 34 controls excluded from the data analyzed in the Jun et al. 2010 study[39]. Each affected individual met NINCDS-ADRDA/NIA-AA[8,11] criteria for probably or definite Alzheimer's disease with age at onset greater than 60 years as determined from specific probe questions within the clinical history provided by a reliable family informant or from documentation of significant cognitive impairment in the medical record. Cognitively healthy controls were unrelated individuals from the same catchment areas and frequency matched by age and gender, and had a documented MMSE or 3MS score in the normal range. Cases and controls had similar demographics: both had similar ages-at-onset/ages-at-exam of 71.1 (±17.4 SD) for cases and 73.5 (±10.6 SD) for controls, and cases and controls were 64.5% and 61.3% female, respectively.

UM/CWRU/TARCC2: The UMCWRUTARCC2 sample included 256 cases and 189 controls from the University of Miami, Case Western Reserve University, and the Texas Alzheimer's Research Care Consortium (wave 2). All Alzheimer's disease cases had onset of disease symptoms after age 65 years and met NINCDS-ADRDA/NIA-AA[8,11] criteria for probable or possible Alzheimer's disease. Controls were adjudicated to have MMSE scores greater than 28 and no clinically identified signs of cognitive impairment. Additional details of subject recruitment at these sites are described in the UM/CWRU/MSSM (formerly UM/VU/MSSM) and TARCC cohort descriptions in this supplement and elsewhere[36–38].

UPITT: The University of Pittsburgh dataset contains 1,254 NHW Alzheimer's disease cases (of which 277 were autopsy-confirmed) recruited by the University of Pittsburgh Alzheimer's Disease Research Center, and 828 NHW, CNEs ages 60 and older (2 were autopsy-confirmed). All Alzheimer's disease cases met NINCDS-ADRDA/NIA-AA[8,11] criteria for probable or definite Alzheimer's disease. Additional details of the cohort used for GWAS have been previously published[76].

WASHU/WASHU2 (WASHU2 part of CSDC): A LOAD case-control dataset consisting of two waves (WASHU1 with 339 cases and 281 healthy elderly controls and WASHU2 with 38 cases and 94 healthy elderly controls) was used in analyses for this study. Participants were recruited as part of a longitudinal study of healthy aging and dementia. Diagnosis of dementia etiology was made in accordance with standard criteria and methods[44]. Severity of dementia was assessed using the Clinical Dementia Rating scale[77].

WHICAP: WHICAP[78,79] is a community-based longitudinal study of aging and dementia among elderly, urban-dwelling residents. Beginning enrolment in 1989, WHICAP has followed more than 5,900 residents over 65 years of age, including white, African American, and Hispanic participants. Detailed clinical assessments were performed at approximately 24-month intervals over the seven years of the initial study. All interviews were conducted in either English or Spanish. The choice of language was decided by the subject to ensure the best performance, and the majority of assessments were performed in the subject's home, which included medical, neurological, and neuropsychological evaluations. Results of the neurological, psychiatric, and neuropsychological assessments were reviewed in a consensus conference comprised of neurologists, psychiatrists, and neuropsychologists. Based on this review all participants were assigned to one of three categories: dementia, cognitive impairment, or normal cognitive function. The sample set available in the ADGC for genetic analyses included 73 Alzheimer's disease cases and 560 subjects with normal cognitive function.

CSDC: The Combined Small Datasets Collection is a harmonized dataset including all data from six separately ascertained datasets or waves of datasets already described above (BIOCARD, CHAP2, EAS, RMAYO, ROSMAP2, and WASHU2). None of these datasets were separately incorporated into analyses and were only analyzed in the CSDC. Datasets or waves were incorporated into the CSDC if they included fewer than 100 cases and/or 100 CNEs. As all datasets were genotyped separately but imputed to the same dataset, genotyped SNVs overlapping all of the high-density genotyping platforms used were extracted and a set of ~20,000 variants were used to estimate both population substructure and explore potential heterogeneity between datasets using principal components analysis (PCA) in the 'smartpca.pl' program in EIGENSTRAT/EIGENSOFT[80,81]. Data-set level association analyses similar to those described for all other cohorts and datasets were performed, though covariate adjustment additionally included indicator variables for study to adjust for residual batch effects not captured in PCs.

###### *1.2.2. Data processing*

*Genotyping*. Genotyping chips are provided in Supplementary Table 22.

*Quality control*. Standard QC was performed on individual datasets using PLINK v1.9[82–84] and including filtering and re-estimating all quality metrics after excluding variants with a missingness rate of >10% of genotype calls. QC filters included exclusions on SNPs with call rates below 98% for Illumina and 95% for Affymetrix panels; SNPs with departure from Hardy-Weinberg Equilibrium (HWE) of $P<10^{-6}$ among cognitively-normal elders (CNEs, either non-cases or controls) for variants of MAF>0.01; and SNPs with informative missingness by case-CNE status of $P<10^{-6}$. Samples were dropped if the individual call rate was <95%; if X chromosome heterozygosity indicated inconsistency between predicted and reported sex; or if population substructure analyses (described below) indicated the sample did not cluster with 1000 Genomes Phase 3 populations of European ancestry.

*Relatedness check*. Relatedness was assessed using the "--genome" function of PLINK v1.9. Using ~20,000 LD-pruned SNPs sampled from among genotyped variants, pi-hat (the proportion of alleles shared IBD) was estimated across all pairs of subjects across all ADGC datasets. Among pairs of subjects with no known familial relationships, one sample was excluded among pairs with pi-hat>0.95 if phenotype and covariate data matched, otherwise both samples were excluded; among all pairs with pi-hat>0.4 but less than 0.95, one sample was kept giving preference to cases over CNEs, age (earlier age-at-onset among case pairs, later age-at-exam among CNE pairs). Pairs of relatives were dropped from family datasets if pi-hat differed substantially from expectation based on their reported relationships.

*Populations substructure*. To identify samples of non-European ancestry, we performed a principal components (PCs) analysis using 'smartpca' in EIGENSOFT v7.2.1[80,81] on the subset of ~20,000 LD-pruned SNPs used for relatedness checks on genotypes from all samples within each individual dataset and from the 1000 Genomes Phase III reference panels. Subjects not clustering with European ancestry groups were excluded from analysis. To account for the effects of population substructure in our analysis, a second PC analysis was performed using only the remaining subjects in each dataset. PCs 1-10 were examined for association with AD case-control status and eigenvector loading, and only PCs showing nominal association with AD (P<0.05) and eigenvector loadings >3 were used in covariate adjustment for populations substructure (average number of PCs used is 3; range: 2-4).

*Imputation*. For each dataset, SNPs not directly genotyped were imputed using the TOPMed (Version 2 on GRC38) reference panel.

*Association tests*. Single variant-based association analysis on datasets of unrelated cases and CNEs were performed in SNPTEST 2.5.4-beta3 using logistic regression under an additive model, with adjustment for PCs only. We analyzed the genotype probabilities using the newml method. Family-based datasets were analyzed using GMMAT 1.4.2.

#### 1.3. Cohorts for Heart and Aging Research in Genomic Epidemiology (CHARGE)

##### *1.3.1. Cardiovascular Health Study (CHS)*

*Setting*. The Cardiovascular Health Study (CHS) is a population-based cohort study of risk factors for coronary heart disease and stroke in adults ≥65 years conducted across four field centers[85]. The original predominantly European ancestry cohort of 5,201 persons was recruited in 1989-1990 from random samples of the Medicare eligibility lists; subsequently, an additional predominantly African-American cohort of 687 persons was enrolled for a total sample of 5,888. Genotyping was performed using the Illumina 370CNV BeadChip system (for European ancestry participants, in 2007) or the Illumina HumanOmni1-Quad_v1 BeadChip system (for African-American participants, in 2010). CHS was approved by institutional review committees at each field center and individuals in the present analysis had available DNA and gave informed consent including consent to use of genetic information for the study of cardiovascular disease.

*Quality control.* The following exclusions were applied to identify a final set of 306,655 autosomal SNPs: call rate < 97%, HWE P < $10^{-5}$, > 2 duplicate errors or Mendelian inconsistencies (for reference CEPH trios), heterozygote frequency = 0, SNP not found in imputation reference panel.

*Imputation.* Imputation to the TOPMed Freeze 5 panel was performed on the Michigan imputation server. SNPs were excluded for variance on the allele dosage ≤0.01. These analyses were limited to the 2152 European ancestry participants from the CHS Memory Study[86] with successful genotyping.

##### *1.3.2. Framingham Heart Study (FHS)*

*Setting*. Framingham Heart Study (FHS) samples consist of 4350 well genotyped individuals from Original and Offspring cohorts[87,88]. The details of recruitment and surveillance of AD, and genotyping in FHS have been detailed previously[40]. Briefly, the Original cohort of the FHS has been evaluated biennially since 1948, was screened for prevalent dementia and AD since 1974-76. The Offspring cohort (offspring of original cohorts and spouse of offspring), recruited in 1971 and examined once every 4 years, have been screened for prevalent dementia with a neuropsychological battery and brain MRI. The AD status used in this study was taken from surveillance up to 2017. FHS participants had DNA extracted and provided consent for genotyping in the 1990s. Genotyping using the Affymetrix GeneChip® Human Mapping 500K Array Set and 50K Human Gene Focused Panel.® was attempted in 5293 Original and Offspring cohort participants.

*Imputation.* Imputation to the TOPMed Freeze 5 panel was performed on the Michigan imputation server.

*Association tests.* GWAS was carried out using a logistic regression model fitted via generalized estimating equations, with each family as a cluster, minimally adjusting for cohort status and the first and ninth PCs that were associated with the outcome.

##### *1.3.3. The Rotterdam Study (RS)*

*Setting*. The Rotterdam Study (RS) is a prospective population-based cohort study among middle-aged and older adults that started in 1990 in the district of Ommoord, in Rotterdam, The Netherlands. The study includes 14,926 participants and has three subcohorts[89]. At start of the study, all inhabitants of the district of Ommoord who were aged 55 years and older were invited to participate. At baseline, in 1990-1993, of the 10,215 invited inhabitants, 7,983 agreed to participate in the baseline examination (RS1, response rate 78%). In 2000, the cohort was extended with 3,011 participants (RS2, 67% of invitees). This extension consisted of all persons living in the study district who had become 55 years and older or had moved into the study district. As of 2008, 14,926 subjects aged 45 years or over comprise the Rotterdam Study cohort. As of 2016, the cohort has been expanded by persons aged 40 years and over. A second extension was initiated in 2006, in which 3,932 participants (RS3, 65% of invitees) who were 45 years and older were included. Study rounds consist of a home interview and visits with extensive investigations at the dedicated research centre. Rounds are repeated every 4-6 years. Participants are continuously monitored for diseases and mortality through linkage of the medical records from the general practitioners and municipality records. The Rotterdam Study has been approved by the Medical Ethics Committee of the Erasmus MC and by the Ministry of Health, Welfare and Sport of The Netherlands. All participants provided written informed consent to participate in the study and to obtain information from their treating physicians.

*Quality control.* For this study, we included a total of 7,421 participants from the first two subcohorts of the RS (RS1 and RS2), that were genotyped and passed genotyping quality control (92% of all subjects with genotyping)[90]. Exclusion criteria were a call rate <98%, Hardy–Weinberg p-value $<10^{-6}$, minor allele frequency <0.01%, excess autosomal heterozygosity >0.336, sex mismatch, and outlying identity-by-state clustering estimates.

*Phenotype definition:* Methods of surveillance for dementia have been published earlier[91]. Participants were cognitively screened during their visits to the study research center, both at baseline and subsequent center visits. Individuals who scored <26 on the Mini-mental state examination (MMSE) test or >0 on the Geriatric Mental State Schedule organic assessment were administered the Cambridge Mental Disorders of the Elderly Examination by a research physician. In addition, linkage of the study database with general practitioner files and the regional institute for outpatient mental health care allowed for continuous monitoring of incident dementia. A consensus panel headed by a consultant neurologist established the final diagnosis according to standard criteria. We studied the outcomes of all-cause dementia (DSM-III-R), and Alzheimer's disease (NINCDS–ADRDA). For the assessment of dementia, and type of dementia, the latest follow-up information with available data was used to determine the disease state. Follow-up for dementia was near complete until January 1st, 2018. Within this period, participants were censored at date of dementia diagnosis, death, or loss to follow-up. For this study, we only included participants from the first and second subcohort (N=7,421).

*Imputation.* Imputation to the TOPMed Freeze 5 panel were performed using the Michigan Imputation Server[33] following a two steps procedure using Eagle v2.4[34] and Minimac4 for the phasing and imputation respectively. No imputation quality filter was applied and the appropriate reference population was selected as per-imputation quality control procedures. For this study the European reference panel was chosen.

*Association tests.* Tests of the association between AD status and autosomal genetic variants were conducted in each dataset by using logistic regression and an additive genetic model, as implemented in SNPTEST 2.5.6[35]. We analyzed the genotype probabilities using the newml method. Analyses were adjusted for the first five principal components (Supplementary Table 22).

##### ***1.3.4. Atherosclerosis Risk in Communities (ARIC) Study***

*Setting.* The Atherosclerosis Risk in Communities (ARIC) study is a prospective cohort study initiated in 1987 and conducted in four U.S. communities (Forsyth County, North Carolina; Jackson, Mississippi; Minneapolis, Minnesota; and Washington County, Maryland)[92]. Since the baseline exam, eleven subsequent follow-up visits have been carried out. At the fifth visit (2011–2013), a subset of surviving participants completed an extensive neuropsychological battery[93], and expert classification of cognitive status (normal/MCI/dementia) was obtained[94]. Diagnoses of dementia for those who were not evaluated in person or by telephone were established following an informant interview. Data used in this manuscript are from visit 5 and include White participants with genotype and dementia assessment.

*Quality control.* Genomic DNA from whole blood was genotyped using the Affymetrix Genome-Wide Human SNP array 6.0 (Affymetrix, Santa Clara, CA, USA). Quality control was carried out before imputation:

single nucleotide polymorphisms (SNPs) were excluded if they had call rate <95%, Hardy–Weinberg equilibrium P values <0.0001, or minor allele frequencies <1%. Individuals with cryptic relatedness, defined as an identity-by-state distance >0.8, generated using PLINK, were also excluded.

*Imputation.* Race-specific imputation of variant dosages were carried out using the TopMed reference panel (freeze 5b) on the Michigan server.

*Association tests.* Tests of the association between AD status and autosomal genetic variants were conducted in each dataset by using logistic regression and an additive genetic model, as implemented in SNPTEST 2.5.6[35]. We analyzed the genotype probabilities using the newml method. Analyses were adjusted for principal components and center (Supplementary Table 22).

###### *1.3.5. Amsterdam UMC*

The Amsterdam UMC provided additional cases and controls from the Amsterdam Dementia Cohort (ADC)[12], not previously genotyped as part of EADB. The Quality control and imputation is described elsewhere[95]. The Sample was imputed to TOPMED (v5) and PLINK –firth-fallback was used for association testing. Variants with the PLINK error code "unfinished" were excluded.

##### 1.4. Psychiatric Genomics Consortium – Alzheimer's Disease Working Group (PGC-ALZ)

###### *1.4.1. Study descriptions*

###### *a) Swedish Twin Studies of Aging (STSA)*

The Swedish Twin Studies of Aging (STSA) (n cases = 398, n controls = 1,079) includes three sub-studies of aging within the Swedish Twin Registry[96]: The Swedish Adoption/Twin Study of Aging (SATSA)[97], Aging in Women and MEN (GENDER)[98], and The Study of Dementia in Swedish Twins (HARMONY)[99]. Informed consent was obtained from all participants and the studies were approved by the Regional Ethics Board in Stockholm and the Institutional Review Board at the University of Southern California. DNA was extracted from blood samples and genotyped using Illumina Infinium PsychArray. Alzheimer's disease patients were diagnosed as part of the studies according to the NINCDS/ADRDA criteria[8]. In addition, information on disease after last study participation was retrieved from three population-based health care registers: The National Patient Register, the Causes of Death Register, and the Prescribed Drug Register.

###### *b) TwinGene*

TwinGene[96] is a population-based study of older twins drawn from the Swedish Twin Registry. Written informed consent was obtained from all participants and the study was approved by the Regional Ethics Board in Stockholm. DNA was extracted from blood samples and genotyped using Illumina Human OmniExpress for 1,791 individuals. Information about Alzheimer's disease (n cases = 343, n controls = 9070) was extracted from the National Patient Register, the Causes of Death Register, and the Prescribed Drug Register, all of which are population-based health care registers with nationwide coverage.

###### *c) Gothenburg*

The Gothenburg H70 Birth Cohort Studies and Clinical AD from Sweden (Gothenburg) AD cases originate from Sweden and were either collected in memory clinics (in different parts of Sweden) or as a part of two population-based epidemiological studies in Gothenburg; the Prospective Population Study of Women (PPSW) and the Gothenburg Birth Cohort Studies (H70, H85 and 95+), described in detail previously[100–103]. Controls originate from the Gothenburg Birth Cohort Studies and PPSW. Individuals of non-European descent were excluded as part of the QC of the GWAS-data. AD diagnosis was based on National Institute of Neurological and Communicative Disorders and Stroke-Alzheimer's Disease and Related Disorders (NINCDS-ADRA) criteria. All control samples were clinically investigated and free from dementia. The individuals were genotyped using the Illumina Neurochip array.

##### *1.4.2. Data processing*

*Quality control.* Genotype data were obtained in Plink v1.90 binary format and, if necessary, were converted to build GRCh37 using the UCSC LiftOver tool. The raw genotypes were processed using the Psychiatric Genomics Consortium (PGC) Ricopili pipeline version 2019_Aug_16.001. The quality control (QC) procedure initially removed SNPs with a missingness > 0.95, then kept individuals with a SNP missingness < 0.05 and an autosomal heterozygosity deviation (Fhet) < 0.2. Finally, SNPs with a missingness > 0.02, a difference in SNP missingness between cases and controls > 0.02; and deviation from Hardy-Weinberg equilibrium ($P < 10^{-6}$ in controls or $P < 10^{-10}$ in cases) were removed. Next, non-European individuals within the datasets were removed based on principle component analysis (PCA), using the 1KG Phase 3 dataset as a reference. The PCA pipeline was repeated including all European individuals in all genotype level datasets to identify individuals across the datasets with a pihat > 0.2 for exclusion from the analysis. PCA was additionally performed within each European dataset to create principal component covariates for logistic regression. These genotype data were lifted over to build GRCh38 using the UCSC LiftOver tool.

*Imputation.* Prior to imputation, genotype data were quality controlled using McCarthy group tools (https://www.well.ox.ac.uk/~wrayner/tools/) using the TOPMed freeze 5 hg38 reference panel variant list (ALL.TOPMed_freeze5_hg38_dbSNP.vcf.gz). Using this tool, variants were coded to fit the TOPMed format (including removing ambiguous variants, flipping strands, and recoding SNP names) then exported as VCF format for upload to the Michigan Imputation Server. The data was then imputed to topmed-r2 v1.0.0 using Minimac4 v1.5.7. Only variants with an INFO score >0.3 were kept.

*Association tests.* The imputed data were analysed in VCF format using SNPTEST v2.5.6, under an additive genetic model. We analyzed the genotype probabilities using the newml method. PCs 1-10 were included as covariates (PCs 11, 13, 15, 16, and 17 were included in the Gothenburg analysis).

#### 1.5. The Trøndelag Health Study (HUNT) – Alzheimer's disease

*Setting*. The Trøndelag Health Study (HUNT) is an ongoing population based cohort study from the county of Trøndelag in Norway[104,105]. All inhabitants aged 20 years or older were invited to participate in the HUNT1 survey (1984-1986), the HUNT2 survey (1995-1997), the HUNT3 survey (2006-2008), and the HUNT4 survey (2017-2019). In addition, all inhabitants aged 13 to 19 years were invited to participate in the Young-HUNT1 survey (1995-1997), the Young-HUNT2 survey (2000-2001), the Young-HUNT3 survey (2006-2008) and the Young-HUNT4 survey (2017-2019). Approximately 150,000 inhabitants have participated in at least one survey. All participants have provided questionnaire, interview, and measurement data, which can be found at the HUNT databank [https://hunt-db.medisin.ntnu.no/hunt-db/]. In addition, participants in HUNT

2-4 have provided biological samples for storage at the HUNT biobank [https://www.ntnu.edu/hunt/hunt-biobank]. The Norwegian Identification Number can be used to link data from local and national registries to data from the HUNT database and the HUNT biobank.

*Genotyping and quality control.* A total of 71,860 participants in the HUNT 2 and 3 studies have been genotyped with Illumina HumanCoreExome arrays (HumanCoreExome12 v1.0, HumanCoreExome12 v1.1, or UM HUNT Biobank v1.0)[106]. We excluded participants whose genotypes had 1) call rates <99%, 2) contamination >2.5% as estimated with BAF Regress[107], 3) large chromosomal copy number variants, 4) lower call rate of technical duplicate pair or twins, 5) uncommon sex chromosomal constellations (i.e. others than XX or XY), or 6) discrepancies with reported gender. The remaining genotypes were analyzed in a second round of genotype calling following the Genome Studio quality control protocol[108] [https://www.illumina.com/techniques/microarrays/array-data-analysis-experimental-design/genomestudio.html]. We used BLAT[109] to determine genomic position, strand orientation and reference allele of all genotyped variants, using the Genome Reference Consortium Human genome build 37 [http://genome.ucsc.edu] and the revised Cambridge Reference Sequence of the Human Mitochondrial DNA [http://mitomap.org] as reference. Variants were excluded if they had 1) call rates <99%, 2) been genotyped in another assay with higher call rates, 3) probe sequences not mapping to the reference genome, 4) cluster separation <0.3, 5) Gentrain score < 0.15, or 6) Hardy Weinberg equilibrium deviation from unrelated samples of European ancestry with p-value <0.0001. To harmonize the three arrays (HumanCoreExome12 v1.0, HumanCoreExome12 v1.1, or UM HUNT Biobank v1.0), we removed variants with frequency differences >15% between the datasets or variants monomorphic in one dataset and had MAF > 1% in one of the other data sets. We inferred ancestry using PLINK v1.90[83], projecting the genotype samples into the space of the principal components of the Human Genome Diversity Project (HGDP) reference panel (938 unrelated individuals; downloaded from http://csg.sph.umich.edu/chaolong/LASER/)[110,111]. Recent European ancestry was defined as samples that fell into an ellipsoid spanning exclusively European populations of the HGDP panel.

*Imputation.* Eagle2 v2.3[34] were then used to phase the data. A total of 69,716 samples passed the quality control. Imputation of genetic variants from the TOPMed reference panel (freeze 5)[112] was performed using the Minimac4 v1.0 software [https://genome.sph.umich.edu/wiki/Minimac4]. Variants with estimated squared correlations between imputed and true genotypes ($R^2$) <0.3 or with MAF < 0.00084 (corresponding to estimated MAC <3 in cases) were excluded, resulting in a total of ~14.7 million well-imputed variants.

*Association testing.* We used the Scalable and Accurate Implementation of GEneralized mixed model (SAIGE) v.0.35.8.3[113] for testing of associations between binary traits and common genetic variants. The SAIGE method is tailored for GWAS analyses in population-based data from a restricted geographic area as it controls for case-control imbalance and relatedness. We removed samples overlapping with the DemGene study. Sex, batch, age, and the first 10 principal components were used as covariates. The principal components were calculated by projecting all samples into the space of the principal components of unrelated HUNT samples, using directly genotyped variants in PLINK v1.90[83].

*Phenotype definition.* The health care system in Norway is publicly funded. Levanger hospital and Namsos hospital, which are the two only hospitals in Nord-Trøndelag, have catchment area responsibilities for the whole county. Diagnoses of Alzheimer's disease are mainly made at geriatric, neurological, and old age psychiatric wards and outpatient clinics. We obtained data from hospital registries on ICD-9 and ICD-10 codes from all inpatient and outpatient contacts from 1987 through 2018 for all genotyped participants in the HUNT study. A subset of these diagnoses have been validated against the ICD-10 criteria for Alzheimer's disease by four specialists in geriatrics and old age psychiatry as part of the Health and Memory Study[114]. A proportion of the genotyped individuals also participated in HUNT4. As part of this study all participants ≥70 years of age (n=19,403) were invited to a substudy (HUNT4 70+), which included cognitive tests by trained health personnel at a field station, at homes, or at their nursing home and a structured carer interview in regard to participants suspected of having dementia. Clinical experts made diagnoses of dementia and dementia subtypes according to DSM-5 criteria[115]. In total 9,930 participants were included in this substudy, and 840 were diagnosed with Alzheimer's disease, of which 718 had valid genotype data. We defined participants as cases if they 1) had one or more hospital contact(s) due to a diagnosis of ICD-9 331.0 Alzheimer's disease, ICD-10 G30 Alzheimer's disease, or ICD-10 F00 Dementia in Alzheimer's disease or 2) had a validated diagnosis of Alzheimer's disease in the HUNT4 70+ study. This resulted in a total of 1776 cases, of which 718 (40,4%) were diagnosed in the HUNT4 70+ study. We defined participants as controls if they were not defined as a case, did not have a validated diagnosis of dementia in HUNT4 70+, and had age >80 years when last seen as participants in a HUNT survey or diagnosed in a hospital.

*Ethics.* The current study is approved by the Regional Committee for Medical and Health Research Ethics (ref. 2017/1031).

##### 1.6. deCODE

*Setting*. Data from the deCODE study included 7,002 Alzheimer's patients (5,098 of whom were chip-typed) and 181,573 controls (88,739 of whom were chip-typed). In 15% of patients, the diagnosis of Alzheimer's disease was established at the Memory Clinic of the University Hospital according to the criteria for definite, probable, or possible Alzheimer's disease of the National Institute of Neurological and Communicative Disorders and Stroke and the Alzheimer's Disease and Related Disorders Association (NINCDS-ADRDA). In 80% of patients, the diagnosis has been registered according to the criteria for code 331.0 in ICD-9, or for F00 and G30 in ICD-10 in health records. Five percent of the patients were identified in the Directorate of Health medication database as having been prescribed Donepezil (Aricept). The controls were drawn from various research projects at deCODE Genetics. The study was approved by the National Bioethics Committee and the Icelandic Data Protection Authority. Written informed consent was obtained from all participants or their guardians before blood samples were drawn. All sample identifiers were encrypted in accordance with the regulations of the Icelandic Data Protection Authority.

*Genotyping, quality control and imputation.* Chip-typing and long-range phasing of 155,250 individuals was carried out as described previously[116]. Imputation of the variants found in 28,075 whole-genome sequenced individuals into the chip-typed individuals and 285,664 close relatives was performed as detailed earlier[116].

*Association tests.* Association analysis in the deCODE sample was carried out using logistic regression with AD status as the response and genotype counts and a set of nuisance variables, including sex, county of birth, and current age, as predictors[117]. Correction for inflation of test statistics due to relatedness and population stratification in this Icelandic cohort was performed using the intercept estimate (1.30) from LD score regression[118].

##### 1.7. UK Biobank

*Phenotype definitions.* We used the February 2022 data release of the UK Biobank (UKBB, application number 61054). For the UKBB-diagnosed phenotype, AD cases were extracted from UK Biobank self-report,

ICD10 code G30 for diagnoses, primary care and cause of death. Our analysis included 2,917 diagnosed cases and 429,538 controls. For the UKBB-proxy analysis, proxy AD/dementia cases included participants who reported at least one biological relative (parents and siblings) affected with dementia, either at baseline or follow up. Participants who answered “Do not know” or “Prefer not to answer” were excluded from analyses. Individuals who did not report AD or any family history of dementia were used as proxy controls. All proxy controls whose parents age/age at death was missing or < 60 were removed. We also excluded individuals from siblings that reported discordant AD status of parents. Our analysis included 55,960 proxy cases of dementia and 291,526 proxy controls.

*Quality control.* The quality control (QC) of the UKBB is described in Bycroft et al.[119]. The QC includes first a marker-based QC testing for batch, plates, and sex effect (genotype frequency differences), departure from HWE within each batch and discordance across control replicates. The genotype calls of the variants failing at least one test were set to missing. The p-value threshold used for the marker-based QC was set to $10^{-12}$. Then, a sample QC was performed, removing samples with poor quality genotype calls, related individuals, population outliers, PC-adjusted heterozygosity above the mean (0.1903) and high missingness in the autosomes (0.05). For the analyses, all individuals of non-European ancestry were excluded. For the UKBB-proxy analysis, all related individuals were excluded using a kinship threshold of 0.25 (controls were excluded over proxy AD cases).

*Imputation.* UKBB dataset was phased with SHAPEIT3 and imputed with a new version of the IMPUTE2 program referred to as IMPUTE4[119]. The imputation panel used is a combination of HRC, UK10K and 1000 Genomes. All samples and variants passing the UKBB QC were used as the input of the imputation process: related individuals and population outliers were not excluded.

*Association test in UKBB with diagnosed cases.* We performed a regression of the AD status on the genetic variants with an additive genetic model using a logistic mixed model as implemented in SAIGE (v1.0.9). Dosages were used. Analyses were adjusted on principal components, genotyping center and sex. The genetic relatedness, used in the first step of the SAIGE analysis, was constructed from autosomal variants:

- that were genotyped
- with MAF ≥1%
- with HWE P >= $1 \times 10^{-15}$
- with missingness < 0.01
- not involved in inter-chromosomal LD; the list of those variants is available in the Supplementary Table 19 of the REGENIE paper[120]
- not in the *APOE* region (40 to 50 Mb on chr 19 in GrCh37 and GrCh38)
- not in regions of high LD;
- remaining after LD pruning using a $r^2$ threshold of 0.9 with a window size of 1,000 markers and a step size of 100 markers.

In step 2, we set the option « impute_method » to « best guessed ». A leave-one-chromosome-out (LOCO) scheme was considered, and effect sizes of variants with p-value <= 0.1 were estimated through the Firth’s Bias-Reduced Logistic Regression.

*Association test in UKBB with proxy cases.* The association test on proxy status in UKBB was performed using the SAIGE protocol described above, with an adjustment on principal components and genotyping center. A correction factor of 2 was applied to the association statistics (effect sizes and standard errors)[121,122].

*Variant filtering*. The chr8:125446584:C:T *TRIB1* variant was excluded from the UKBB summary statistics, because its frequency was very different in the UKBB ($3x10^{-4}$) compared to the other datasets (20%).

##### 1.8. FinnGen

*Setting*. Patients and control subjects in FinnGen provided informed consent for biobank research, based on the Finnish Biobank Act. Alternatively, separate research cohorts, collected prior the Finnish Biobank Act came into effect (in September 2013) and start of FinnGen (August 2017), were collected based on study-specific consents and later transferred to the Finnish biobanks after approval by Fimea (Finnish Medicines Agency), the National Supervisory Authority for Welfare and Health. Recruitment protocols followed the biobank protocols approved by Fimea. The Coordinating Ethics Committee of the Hospital District of Helsinki and Uusimaa (HUS) statement number for the FinnGen study is Nr HUS/990/2017.

The FinnGen study is approved by Finnish Institute for Health and Welfare (permit numbers: THL/2031/6.02.00/2017, THL/1101/5.05.00/2017, THL/341/6.02.00/2018, THL/2222/6.02.00/2018, THL/283/6.02.00/2019, THL/1721/5.05.00/2019 and THL/1524/5.05.00/2020), Digital and population data service agency (permit numbers: VRK43431/2017-3, VRK/6909/2018-3, VRK/4415/2019-3), the Social Insurance Institution (permit numbers: KELA 58/522/2017, KELA 131/522/2018, KELA 70/522/2019, KELA 98/522/2019, KELA 134/522/2019, KELA 138/522/2019, KELA 2/522/2020, KELA 16/522/2020), Findata permit numbers THL/2364/14.02/2020, THL/4055/14.06.00/2020,,THL/3433/14.06.00/2020, THL/4432/14.06/2020, THL/5189/14.06/2020, THL/5894/14.06.00/2020, THL/6619/14.06.00/2020, THL/209/14.06.00/2021, THL/688/14.06.00/2021, THL/1284/14.06.00/2021, THL/1965/14.06.00/2021, THL/5546/14.02.00/2020, THL/2658/14.06.00/2021, THL/4235/14.06.00/2021 and Statistics Finland (permit numbers: TK-53-1041-17 and TK/143/07.03.00/2020 (earlier TK-53-90-20) TK/1735/07.03.00/2021).

The Biobank Access Decisions for FinnGen samples and data utilized in FinnGen Data Freeze 8 include: THL Biobank BB2017_55, BB2017_111, BB2018_19, BB_2018_34, BB_2018_67, BB2018_71, BB2019_7, BB2019_8, BB2019_26, BB2020_1, Finnish Red Cross Blood Service Biobank 7.12.2017, Helsinki Biobank HUS/359/2017, Auria Biobank AB17-5154 and amendment #1 (August 17 2020), AB20-5926 and amendment #1 (April 23 2020), Biobank Borealis of Northern Finland_2017_1013, Biobank of Eastern Finland 1186/2018 and amendment 22 § /2020, Finnish Clinical Biobank Tampere MH0004 and amendments (21.02.2020 & 06.10.2020), Central Finland Biobank 1-2017, and Terveystalo Biobank STB 2018001.

FinnGen research project is a public-private partnership combining genotype data from Finnish biobanks and digital health record data from Finnish health registries (Kurki et al., 2022). FinnGen utilizes biobank samples that consist of 1) prospective samples (‘new samples’) and 2) legacy samples. 'New samples' can be collected from voluntary individuals thought Hospital biobank, Terveystalo Biobank or Blood Service Biobank. Legacy samples are older sample cohorts that have been collected for a specific research project before the Finnish Biobank Act came into effect (September 2013) and have then been transferred to a biobank according to the Finnish Biobank Act 13 §. The ‘new samples’ were genotyped with FinnGen ThermoFisher Axiom custom array at the ThermoFisher genotyping service in San Diego, CA, US. The ‘legacy samples’ were genotyped over the years using various generations of Illumina and Affymetrix GWAS arrays.

*Phenotype definition:* We used the AD cases from the FinnGen Data Freeze 8 using G6_ALZHEIMER where cases are defined by having ICD-10 code G10 or ICD-9 code 3310 in either hospital discharge records or as the cause of death. In total, the G6_ALZHEIMER has 7,759 cases and 334,740 controls with high-quality genotypes and genotype-verified sex.

*Quality control.* The genotype data processing is described in detail in: https://finngen.gitbook.io/finngen-handbook/finngen-data-specifics/red-library-data-individual-level-data/genotype-data/description-of-how-the-data-is-processed-in-refinery). Individuals with ambiguous sex, high genotype missingness (>5%), excess heterozygosity (+-3SD) and non-Finnish ancestry were excluded, and variants with high missingness (>2%), low HWE P-value (<1e-6) and low minor allele count (MAC<3) were excluded.

*Imputation.* The genotype data were imputed with a Finnish population specific reference panel, Sisu (V4), described in https://finngen.gitbook.io/finngen-handbook/finngen-data-specifics/red-library-data-individual-level-data/genotype-data/imputation-panel. Genotype imputation process is described in https://dx.doi.org/10.17504/protocols.io.xbgfijw.

*Association tests.* We used the FinnGen R8 summary statistics, obtained by performing a logistic mixed model as implemented in regenie 2.2.4[120]. For regenie step 1 LOCO prediction computation, age, sex, 10 PCs and genotyping batch were used as covariates. For calculating genetic relatedness in regenie step 1, the following variants were included: 1) variants imputed with an INFO score > 0.95 in all batches and 2) variants with > 97 % non-missing genotypes and 3) variants with MAF > 1 %. The remaining variants were LD pruned with a 1Mb window and r2 threshold of 0.1. This resulted in a set of 61,289 well-imputed not rare variants for relatedness calculation. Association tests were then run with regenie for each variant with a minimum allele count of 5 among cases and controls. The approximate Firth test was used for variants with an initial p-value of less than 0.01 and the standard error was computed based on effect size and likelihood ratio test p-value (regenie options –firth--approx --pThresh 0.01 –firth-se).

#### 2. Effective sample size

The effective sample size represents the sample size for a hypothetical GWAS with equivalent power as our meta-analysis, but with a balanced sample (50% cases, 50% controls) and with diagnosed cases only, so without any proxy cases. The effective sample size Neff was computed for each study included in the meta-analysis using this formula: Neff = 4 / ( 1 / N_cases + 1 / N_controls ) with N_cases and N_controls the number of cases and controls respectively. Neff for the proxy UK Biobank study was computed by dividing the raw number of proxy cases and proxy controls by four[121,123]. Neff of the meta-analysis was then computed as the sum of Neff across studies[124]. This estimation does not consider relatedness in biobanks.

#### 3. Detailed meta-analyses results

##### 3.1. Main meta-analysis

With the clumping strategy, we detected 96 loci at the genome-wide significance level of $5 \times 10^{-8}$ (Figure 1, Supplementary Tables 2 and 3, Supplementary Figures 3-55). Two independent loci, each with a lead variant, were annotated as the *IGH* gene cluster and merged in the reporting. The two *IGH* loci were considered separately though in the conditional analyses (see below). In a sensitivity meta-analysis, we did not filter the per-study GWAS on the effective allele count, except in FHS. The *TREM2* locus was represented by a different lead variant in the two meta-analyses: chr6:40974457:G:A in the main one and chr6:41161514:C:T (also named R47H) in the sensitivity analysis; both variants were in high LD ($r^2$=0.887 in the EADB-core samples). Additionally, the *SORT1* chr1:109345810:T:C lead variant was only identified in the sensitivity analysis. The effect sizes of the three variants being similar in both meta-analyses (Supplementary Table 23), we considered the sensitivity meta-analysis results for those two loci. Finally, 97 lead variants were identified in 96 loci. Eighteen of the loci were novel in European samples although two of them, *ADGRL3* and *TRIB1*, were recently identified in a trans-ethnic GWAS[125].

We performed a stepwise conditional analysis within each locus (excluding *APOE* and *ADGRL3*), considering a suggestive threshold of $1x10^{-5}$. We identified 157 variants, including the 95 lead variants of the loci (with two lead variants in the *IGH* locus). Overall, we thus identified 159 independent signals in 96 loci at the suggestive significance level. Among them, 24 secondary signals in 15 previously known loci were genome-wide significant (GWS): *NCK2*, *BIN1*, *RHOH*, *HLA*, *TREM2*, *CLU/PTK2B*, *SHARPIN*, *PICALM*, *SORL1*, *SLC24A4/RIN3*, *ADAM10*, *PLCG2*, *ACE*, *ABCA7* and *CD33* (Figure 1, Supplementary Table 3). Considering the two signals in the *IGH* locus, we thus have 25 GWS secondary signals in 16 loci. This is the first time a secondary signal is detected in *CD33*, *HLA*, *PICALM* and *RHOH*, while we detect more secondary signals than previously in *ABCA7* (4), *BIN1* (3), *PTK2B/CLU* (3), *NCK2* (3) and *PLCG2* (5)[19,21,126,127]. All those secondary signals were validated in a stepwise conditional analysis performed on variants meeting more stringent criteria on imputation quality and effective sample size, except two rare variants in *ABCA7*; for such rare variants, it is however more difficult to have a tag variant meeting the stringent criteria (Supplementary Table 3). Some secondary signals in *BIN1*, *CLU/PTK2B*, and *PLCG2* were GWS after conditional analysis only, and there was a difference of more than 1 log between the results of the single variant analysis and the stepwise conditional analysis. The exact conditional analysis performed in a subset of the samples confirmed that the single variant analysis results were deflated at those index variants (Supplementary Table 3). On the contrary, four lead variants were not GWS anymore after conditional analysis - although they still had $P < 1x10^{-5}$ - in the two known loci *SEC61G/EGFR* and *SPPL2A/USP8/USP50*, and in the two novel loci *TRIB1* and *AXIN1*. For *SEC61G/EGFR*, *TRIB1* and *AXIN1*, the results of the stepwise conditional analyses might have been impacted by imperfect LD measurement due to imputation, or different effective sample sizes between variants. Indeed, the strict stepwise conditional analyses did not validate a secondary signal at those loci. However, for all those loci, the single variant results might also have been slightly inflated[128]. Indeed, the decrease in the significance of the index variants between the single variant analysis and the stepwise conditional analysis was also noticed in the exact conditional analyses performed in a subset of the samples (Supplementary Table 3).

We then performed a joint analysis of all the 158 index variants of the non-*APOE* loci to validate the independence of the different loci. For all loci except 5 (*SORT1*, *TREM2*, *KAT8/BCKDK*, *KLF16/REXO1* and *VAV1*), the index variants were GWS in both the within-loci conditional analysis and the joint analysis performed across all loci. For *SORT1* and *TREM2*, the conditional analysis was performed on the meta-analysis not filtered on effective allele count, while the joint analysis was performed on the one filtered on an effective allele count of 5; this explains the loss of GWS in the joint analysis. For *KLF16/REXO1* and *VAV1*, the results are borderline GWS after the joint analysis; the loss of significance might just be due to the approximation of the GCTA joint analysis (for example due to imperfect LD measurement in an imputed reference panel). For *KAT8/BCKDK*, we performed an approximate conditional analysis of the lead variant on the one of the neighboring locus *DOC2A*, which confirmed a decrease in the effect size and in the significance of the signals at both loci (Supplementary Table 4). This was also confirmed by an exact conditional analysis of both index variants in a subset of the studies (Supplementary Table 4). Overall, the signal at *KAT8/BCKDK* and *DOC2A* seems to be slightly inflated in the single variant analysis, and the

*KAT8/BCKDK* signal requires further validation. We observed the same behavior for the *APH1B* and *SNX1/CIAO2A* signals, though both signals remain GWS after conditional analysis (Supplementary Table 4).

Overall, we considered that 91 loci were genuine Tier 1 GWS signals, and we classified as Tier 2 the GWS signals at *SEC61G/EGFR*, *SPPL2A/USP8/USP50*, *TRIB1*, *AXIN1* and *KAT8/BCKDK*, considering that they required further external validation (Figure 1, Supplementary Tables 2 and 3). In the Tier 1 loci, 117 index variants were lead variants or showed a GWS signal in the joint analysis. Among those 117 index variants, 74 variants were located in a protein-coding gene: 18 missense variants (in genes from 15 loci: *MME*, *TREM2*, *DOCK4*, *SHARPIN*, *SPI1* in the *CELF1/SPI1* locus, *MS4A6A* in the *MS4A* locus, *APH1B*, *CTSH*, *DOC2A*, *PLCG2*, *ABI3*, *ABCA7*, *APOE*, *SORT1* and *VAV1*), 1 splice donor variant (in *ABCA7*), eight 3' or 5' UTR variants, and 47 intronic variants (Supplementary Table 3). Three of the 18 missense variants had a REVEL score greater than 0.25 (in the *MME*, *TREM2* and *ABCA7* loci), and only seven of the 117 Tier 1 GWS index variants were rare (in 6 loci: *SORT1*, *NCK2*, *ADGRL3*, *TREM2*, *PLCG2* and 2 in *ABCA7*). Among the remaining 42 index variants, 27 were located in a gene, including 2 missense variants, two 3' or 5' UTR variants, 1 synonymous variant and 22 intronic variants. The two missense variants were located in *RIN3* in the *SLC24A4/RIN3* locus (with a REVEL score of 0.576) and in *LILRB4* in the *LILRB1* locus. Two of the 42 suggestive variants were rare (in the *TREM2* locus and the missense variant in *RIN3*).

Four loci identified in the two largest AD GWAS meta-analyses on European ancestry samples[19,127] were not GWS in the EADB-IGAP-PGC meta-analysis: *HAVCR2*, *SLC2A4RG/LIME1*, *FOXF1* and *NTN5* (Supplementary Figure 56).

##### 3.2. Sensitivity meta-analyses

Among the 91 Tier 1 loci, 75 (82.4%) and 56 (61.5%) were GWS in the no-proxy and no-biobank meta-analyses respectively (Figure 1, Supplementary Table 7). The *LILRB2* and *LILRB1/LILRB4* loci were identified as a single locus in both sensitivity meta-analyses (according to the start and end position of the loci), but the *LILRB1/LILRB4* lead variant was GWS only in the no-proxy meta-analysis. Among the 91 Tier 1 loci, only *ABCA1* was GWS in the no-biobank meta-analysis, but not in the no-proxy meta-analysis, although borderline ($P=5.49 \times 10^{-8}$, Supplementary Table 7). Although some lead variants were different across the main, no-proxy and no-biobank analyses, they were tagging the same signal, except at the *NCK2*, *PLCG2* and *LILRB2* loci (Supplementary Table 7). In the *NCK2* locus, the lead variant of the main and no-biobank meta-analyses (chr2:105749599:T:C) was identified as a secondary GWS signal in the no-proxy meta-analysis, while the lead variant of the no-proxy meta-analysis (chr2:105732208:T:C) was identified as a GWS signal in the main and no-biobank meta-analyses (Supplementary Table 7). In the *PLCG2* locus, the lead variants of the main (chr16:81908423:C:G), no-proxy (chr16:81942226:T:G) and no-biobank (chr16:81739398:G:A) meta-analyses were tagging three different signals; however, each of those lead signals was identified as a GWS secondary signal in the other meta-analyses (Supplementary Table 7). In the *LILRB2* locus, the lead variants of the main (chr19:54305226:T:C) and the no-proxy meta-analyses (chr19:54302959:T:C) were tagging the same signal. However this signal seems independent from the lead (chr19:54267597:C:T) and secondary (chr19:54565959:G:A) signals detected in the no-biobank meta-analysis, as a signal remains at those two variants (p-value in the order of $10^{-7}$) despite the adjustment on the lead variant of the main meta-analysis (Supplementary Table 7). Finally, we note that, in the *ATP8B4* gene, the lead variant detected in the no-proxy and no-biobank was a rare missense variant predicted pathogenic (chr15:49972642:C:T, REVEL score of 0.859, MAF=0.0087). This variant was already detected in the EADB meta-analysis[19], but was not considered as a novel locus because it was not nominally significant in Stage 2. The evidence for the signal increased in this new analysis ($P=4.3 \times 10^{-11}$ vs $P=1.26 \times 10^{-9}$ in EADB).

Ten of the 75 GWS loci in both the main and no-proxy meta-analyses were novel: *PTPRC*, *MGAT5*, *FAM193B*, *TMEM184A*, *DOCK4*, *IPMK*, *UBFD1*, *LRRC25*, and *CEP89* and *LILRB1/LILRB4*. The *UBFD1* and *LRRC25* novel loci were also GWS in the no-biobank meta-analysis. This provides further support for those novel loci. We note that the Tier 2 locus *SPPL2A/USP8/USP50* was GWS in the no-proxy meta-analysis, and the Tier 2 locus *TRIB1* was GWS in both the no-proxy and the no-biobank meta-analyses, supporting that some Tier 2 loci are genuine.

Of note, nine loci were identified in the no-proxy meta-analysis only - *MFSD10*, *ILRUN*, *AKR1D1*, *ANO5*, *RIC8B*, *GTF2H3*, *C16orf95*, *SNAI1* and *PSMG1* -, while four loci were identified in the no-biobank meta-analysis only - *C1S*, *MEFV*, *ZCCHC2* and *RIPOR3* (Figure 1, Supplementary Table 7, Supplementary Figures 57 - 61). All were novel loci, and were classified as Tier 2 loci. The lead variant of the *MEFV* locus was rare (MAF=0.0023). *C16orf95* is associated with phosphorylated tau (pTau) levels in cerebrospinal fluid (CSF); the AD and pTau index variants tag the same signal ($r^2$=0.779 and D'=0.997 in TOPMed[129])[130]. However, the directions of effect are counterintuitive: the minor allele of the AD index variant decreases AD risk, while the minor allele of the pTau index variant increases CSF pTau levels. This discrepancy was already observed in Jansen et al, as the minor allele of the pTau index variant is also associated with a smaller ventricular volume, implying less neurodegeneration (in agreement with the effect direction for AD risk)[130,131]. This latter observation supports the validity of some of those Tier 2 loci, despite the absence of signal in the UKBB-proxy.

Among the 25 GWS secondary signals, 18 and 12 were identified or tagged in the no-proxy and no-biobank meta-analyses respectively. Among them, 11 signals were detected in all 3 analyses (in *NCK2*, *TREM2*, *CLU/PTK2B*, *SHARPIN*, *SORL1*, *IGH* gene cluster, *ADAM10/ALDH1A2*, *PLCG2*, *ACE* and *ABCA7)* , while one of the secondary signal in the *TREM2* locus was detected only in the no-biobank meta-analysis. Six secondary GWS signals - in *BIN1*, *ATP8B4*, *LILRB2*, *HLA*, *ABI3* and *ADAM10/ALDH1A2* - were detected in the no-proxy and the no-biobank meta-analyses only.

#### 4. Single-cell enrichment analysis

##### 4.1. Methods

We performed a three-step process to assess the association of gene overexpression and gene association with AD for each version of the 6 models (primary, common-only, no-apoe, larger window, no-proxy and no-biobank, see Online Methods). First, FUMA cell type analysis performed an association analysis between the MAGMA gene Z-scores and the degree of overexpression of a gene in a cell type compared to the average expression of that gene across all cell types in a dataset. The model was defined using the following command:

```
magmav1.08 --gene-results [input file name].genes.raw \
--gene-covar [file name of selected scRNA-seq data set] \
--model condition-hide=Average direction=greater \
--out [output file name]
```

Significant cell types were selected after Bonferroni correction for the total number of cell types tested across all datasets within each model ($0.05/136=3.68x10^{-4}$). This is a stringent threshold for multiple testing correction within each analysis given the high correlation between gene expression in cell types across datasets, some of which included the same individuals.

Secondly, within dataset conditional analyses were performed where the association analysis above was repeated except each significant cell type association was conditioned on one of the other significantly associated cell types from the same dataset. The proportional significance was calculated as the ratio of conditional significance to marginal significance (-log10(P_conditioned)/-log10(P_marginal)) and was used to forward select independent cell types from each dataset. The criteria for selecting cell types is described here https://fuma.ctglab.nl/tutorial#workflow. In summary, cell types which were the less significant cell type of a pair were not selected unless the proportional significance of both cell types in the pair were above 0.8 or 0.5 after conditioning on each other.

Thirdly, the cell types selected from step 2 were used to perform across dataset conditional analysis. This step performed the same pairwise conditional analysis as above except the association between a cell type gene expression and MAGMA gene association was also conditioned on the two average cell type gene-expression values within each dataset. The model for an example analysis for conditioning cell type 1 in dataset 1 on cell type 2 in dataset 2 is below:

Celltype1_Dataset1_expression ~ AD_Z_score + Celltype2_Dataset2_expression + Dataset1_Average_expression + Dataset2_Average_expression

Proportional significance was calculated by comparing the conditional significance from the model described above to the significance when only conditioning on the average gene-expression of all cell types in each of the datasets (model below):

Celltype1_Dataset1_expression ~ AD_Z_score + Dataset1_Average_expression + Dataset2_Average_expression

The formulas for these calculations are available at https://fuma.ctglab.nl/tutorial#celltype. The three-step process was repeated using the 6 MAGMA gene level results described above as input. The cell labels were assigned in the original publications and in our analyses we interpreted the following labels as microglia: MG, Mgl, Micro, Micro_C1QB, Micro_L1.3_TYROBP, Micro_out, microglia. The figures were generated using ggplot2 in R v4.1.3.

#### 4.2. Detailed results

##### *4.2.1. Primary analysis*

In step 1 of the primary analysis, 4 cell type dataset pairs were identified as significant after Bonferroni correction for 136 cell type dataset pairs ($0.05/136=3.68x10^{-4}$) (Supplementary Figure 62; Supplementary Table 9). All four of these cell types were microglia (Micro_L1.3_TYROBP in Allen_Human_MTG_level2, MG in DroNc_Human_Hippocampus, Microglia in PsychENCODE_Adult, Micro in GSE168408_Human_Prefrontal_Cortex_level2_Adult) and had P-values ranging from $1.13x10^{-13}$ to $6.06x10^{-8}$. The only 2 datasets which did not show a significant association were Allen_Human_MTG_level1 and GSE168408_Human_Prefrontal_Cortex_level1_Adult and this was likely due to the cell types in these datasets being defined too broadly (all non-neuronal cells being aggregated together). The non-microglia cell type with the lowest P-value was an endothelial cell type (Endo_L2.6_NOSTRIN in

Allen_Human_MTG_level2) which had a P-value of 0.0022. The strength of the association with microglia and the lack of association signal in other cell types suggests that only microglia (in these datasets) overexpress ADD relevant genes. Since only one significant cell type was identified in each dataset, all 4 cell type associations were taken forward for analysis to step 3, and step 2 was not performed.

In step 3, the 4 significantly associated cell types were conditioned on each other to determine whether these associations were independent from each other. Proportional significance was calculated (-log10(P_conditioned)/-log10(P_marginal)) to determine how independent the datasets were from each other, with larger values (0-1) indicating higher independence. All 4 of the cell types had proportional significance <0.2 after conditioning on the most significant cell type dataset pair (Micro in GSE168408_Human_Prefrontal_Cortex_level2_Adult) (Supplementary Figure 62; Supplementary Table 10). This suggests that more than 80% of the original association of each of these cell types across the datasets was captured by the association of Micro in GSE168408_Human_Prefrontal_Cortex_level2_Adult. In combination, the results from step 1 and step 3 show that genes associated with ADD are overexpressed in microglia in 4 different datasets spanning different brain regions and that the association of microglia across the datasets is caused by the same set of genes.

##### *4.2.2. Sensitivity analyses*

All of the four significant cell type dataset pairs from the primary results were also significant (P<0.05/136) in the five sensitivity analyses (Supplementary Figure 63, Supplementary Table 9) and no other cell types were significant in any of the five sensitivity analyses. Generally, the larger variant to gene mapping windows yielded more significant results compared to the primary analysis. This was also true for the exclusion of the *APOE* region. Conversely, only including common variants or reducing the sample size of the meta-analysis (exclusion of biobank or proxy datasets) caused a decrease in significance of the cell type associations compared to the primary analysis. The step 3 results from the sensitivity analyses only differed slightly compared to the primary results (Supplementary Figure 64, Supplementary Table 10), with more than 75% of the cell type associations being explained by the association of Micro in GSE168408_Human_Prefrontal_Cortex_level2_Adult.

#### 5. Polygenic scores results

The signals for the three AD-related NPEs and LATE-NC were in the the same direction in ADC/NACC and the smaller ACT study (Figure 2a, Supplementary Figure 65, Supplementary Table 17). There was some evidence of heterogeneity between ACT and ADC/NACC, however this phenomena is difficult to assess for only two studies. Heterogeneity, if present, appeared a bit reduced after adjusting on AD diagnosis (Supplementary Figure 65b).

In ADC/NACC, only the associations with Braak stage and CERAD score remained significant after adjustment on the (other) AD NPEs (Figure 2b, Supplementary Table 17). The association with amyloid-β plaques was not significant anymore, while the association with LATE-NC remained significant at the nominal level only ($P=3.78 \times 10^{-2}$, Supplementary Table 17), with an OR decreasing from 1.056 [1.025-1.088] to 1.035 [1.00-1.068]. When adjusting the analysis of Braak stage and CERAD score on LATE-NC in addition to the two other AD NPEs, CERAD score remained significant. Braak stage was only nominally significant ($P=5.86 \times 10^{-3}$), with a very similar OR with and without adjustment on LATE-NC (OR=1.064 [1.018-1.112] vs

OR=1.067 [1.044-1.091]), indicating that the loss of significance is due to a loss of power, as the sample size for LATE-NC is much lower than for the other NPEs (Supplementary Table 16). Overall, we conclude that the PGS is associated with Braak stage and CERAD score independently of the other AD NPEs or LATE-NC in ADC/NACC, but a larger sample size is required to assess if the association with LATE-NC is due or not to the frequent co-occurrence of LATE-NC and AD NPEs. There was no significant interaction ($P \leq 2.27 \times 10^{-3}$) between the PGS and the number of APOE ε4 and ε2 alleles for Braak stage and CERAD score (Supplementary Table 17). In the ADC/NACC dataset, compared to the individuals with a main PGS in the median quintile (40 to 60%), individuals in the 10$^{th}$ decile had a risk increased by 2.05-fold (95% CI=[1.47-2.85]) and 1.96-fold (95% CI=[1.39-2.78]) for Braak stage 5 to 6 and moderate/severe neuritic amyloid plaque pathology at death respectively, while individuals in the 1$^{st}$ decile had a risk decreased by 0.47-fold (95% CI=[0.37-0.61]) and 0.43-fold (95% CI=[0.33-0.56]) respectively (Figure 3 and Supplementary Table 18). To allow comparison with the smaller ACT study, association results are also provided per quintiles (Supplementary Table 19, Supplementary Figure 67).

#### 6. List of software and URLs

Bedtools: https://bedtools.readthedocs.io
BCFtools: http://samtools.github.io/bcftools/bcftools.html
Gene Ontology (downloaded from the NCBI on June 19th, 2023): http://geneontology.org/docs/download-ontology/
Reactome (July 17th, 2023): https://reactome.org/download-data
KEGG and Pathway Interaction Database (PID) pathways (MSigDB v2023.1.Hs updated March 2023): https://www.gsea-msigdb.org/gsea/msigdb/index.jsp
LocusZoom: https://github.com/statgen/locuszoom-standalone
Sanger imputation server: https://imputation.sanger.ac.uk/
Michigan imputation server: https://imputationserver.sph.umich.edu/
SNPTEST 2.5.4-beta1, 2.5.4-beta3, and 2.5.6: https://www.chg.ox.ac.uk/~gav/snptest/
FlashPCA2: https://github.com/gabraham/flashpca
EIGENSOFT: https://www.hsph.harvard.edu/alkes-price/software/
PLINK 1.9: https://www.cog-genomics.org/plink/
PLINK 2.0: https://www.cog-genomics.org/plink/2.0/
GMMAT 1.4.2: https://cran.r-project.org/web/packages/GMMAT/index.html
SAIGE v.0.35.8.3 and 1.0.9: https://saigegit.github.io/SAIGE-doc/
regenie v2.2.4: https://rgcgithub.github.io/regenie/
GCTA-COJO: https://yanglab.westlake.edu.cn/software/gcta/#COJO
FUMA v1.5.2 and v1.6.1: https://fuma.ctglab.nl/
UCSC LiftOver: https://genome.ucsc.edu/cgi-bin/hgLiftOver
McCarthy group tools: https://www.well.ox.ac.uk/~wrayner/tools/
Human Genome Diversity Project (HGDP) reference panel: http://csg.sph.umich.edu/chaolong/LASER/
Minimac4: https://genome.sph.umich.edu/wiki/Minimac4
LD Score Regression: https://github.com/bulik/ldsc
METAL v2020-05-05: https://github.com/statgen/METAL/tree/master
LDstore 2: http://www.christianbenner.com/
emeraLD: http://github.com/statgen/emeraLD
GWAS catalog version e112_r2024-07-08: https://www.ebi.ac.uk/gwas/

VEP: https://www.ensembl.org/info/docs/tools/vep/index.html
PhenoGram: https://ritchielab.org/software/phenogram-downloads
ADES-ADSP summary statistics: https://doi.org/10.5281/zenodo.6818051
MAGMA v1.08: https://cncr.nl/research/magma/
ontologyIndex R package: https://cran.r-project.org/web/packages/ontologyIndex/index.html
qqman R package: https://cran.r-project.org/web/packages/qqman/index.html
GenABEL R package (estlambda function): https://github.com/GenABEL-Project/GenABEL
rmeta R package (forestplot function): https://cran.r-project.org/web/packages/rmeta/index.html
ggplot2 R package: https://cran.r-project.org/web/packages/ggplot2/index.html
1000 Genomes data, Phase 3: https://www.internationalgenome.org/data
pROC R package: https://cran.r-project.org/web/packages/pROC/index.html
rcompanion R package: https://cran.r-project.org/web/packages/rcompanion/index.html

#### 7. Supplementary List of Authors

##### EADB

Sonia Moreno-Grau[1,2], Rafael Campos-Martin[3,4], Dag Aarsland[5,6], Carla Abdelnour[1,2], Emilio Alarcón-Martín[1,7], Daniel Alcolea[8,2], Montserrat Alegret[1,2], Ignacio Alvarez[9,10], Nicola J. Armstrong[11], Tsolaki Anthoula[12,13], Ildebrando Appollonio[14,15], Marina Arcaro[16], Silvana Archetti[17], Alfonso Arias Pastor[18,19], Lavinia Athanasiu[20], Henri Bailly[21], Nerisa Banaj[22], Miquel Baquero[23], Ana Belén Pastor[24], Sonia Bellini[25], Claudine Berr[26], Céline Besse[27], Valentina Bessi[28,29], Giuliano Binetti[25,30], Alessandra Bizarro[31], Rafael Blesa[8,2], Silvia Boschi[32], Paola Bossù[33], Geir Bråthen[34,35], Catherine Bresner[36], Henry Brodaty[11,37], Keeley J. Brookes[38], Dolores Buiza-Rueda[2,39], Katharina Bûrger[40,41], Vanessa Burholt[42,43], Miguel Calero[2,44,45], Geneviève Chene[46,47], Ángel Carracedo[48,49], Roberta Cecchetti[50], Laura Cervera-Carles[8,2], Camille Charbonnier[51], Caterina Chillotti[52], Simona Ciccone[53], Jurgen A.H.R. Claassen[54], Jordi Clarimon[8,2], Christopher Clark[55], Elisa Conti[14], Anaïs Corma-Gómez[56], Guido Maria Giuffre[57,58], Carlo Custodero[59], Delphine Daian[27], Efthimios Dardiotis[60], Jean-François Dartigues[61], Peter Paul de Deyn[62], Teodoro del Ser[24], Nicola Denning[63], Janine Diehl-Schmid[64], Mónica Diez-Fairen[9,10], Paolo Dionigi Rossi[53], Srdjan Djurovic[20], Emmanuelle Duron[21], Sebastiaan Engelborghs[65,66,67,68], Valentina Escott-Price[63,36], Ana Espinosa[1,2], Michael Ewers[40,41], Tagliavini Fabrizio[69], Lucia Farotti[70], Chiara Fenoglio[71], Marta Fernández-Fuertes[56], Catarina B Ferreira[72], Evelyn Ferri[53], Bertrand Fin[27], Peter Fischer[73], Tormod Fladby[74], Klaus Fließbach[75,76], Juan Fortea[8,2], Tatiana M. Foroud[77], Silvia Fostinelli[25], Nick C. Fox[78], Emlio Franco-Macías[79], Ana Frank-García[2,80,81], Lutz Froelich[82], Jose Maria García-Alberca[2,83], Sebastian Garcia-Madrona[84], Guillermo Garcia-Ribas[84], Ina Giegling[85], Giaccone Giorgio[69], Oliver Goldhardt[64], Antonio González-Pérez[86], Giulia Grande[87], Emma Green[88], Tamar Guetta-Baranes[89], Annakaisa Haapasalo[90], Georgios Hadjigeorgiou[91], Harald Hampel[92,93], John Hardy[94], Annette M. Hartmann[85], Janet Harwood[36], Seppo Helisalmi[95,96], Michael T. Heneka[97,98], Isabel Hernández[1,2], Martin J. Herrmann[99], Per Hoffmann[100], Clive Holmes[101], Raquel Huerto Vilas[18,19], Marc Hulsman[102,103], Geert Jan Biessels[104], Charlotte Johansson[105,106], Lena Kilander[107], Anne Kinhult Ståhlbom[105,106], Miia KIVIPELTO[108,109,110,111], Anne Koivisto[95,112,113], Johannes Kornhuber[114], Mary H. Kosmidis[115], Carmen Lage[2,116], Erika J Laukka[87,117], Alessandra Lauria[31], Jenni LEHTISALO[95,118], Ondrej LERCH[119,120], Alberto Lleó[8,2], Adolfo Lopez de Munain[2,121], Seth Love[122], Malin Löwemark[107], Lauren Luckcuck[36], Juan Macías[56], Catherine A. MacLeod[123], Wolfgang Maier[124,76], Francesca MANGIALASCHE[108], Marco Spallazzi[125], Marta Marquié[1,2], Rachel Marshall[36], Angel Martín Montes[2,80,81], Carmen Martínez Rodríguez[126], Simon Mead[127], Miguel Medina[2,44], Alun Meggy[63], Silvia Mendoza[83], Manuel Menéndez-González[126], Merel Mol[128], Laura Montrreal[1], Kevin Morgan[129], Markus M Nöthen[100], Tiia NGANDU[118], Robert Olaso[27], Adelina Orellana[1,2], Michela Orsini[130], Gemma Ortega[1,2], Alessandro Padovani[131], Paolo Caffarra[132], Alba Pérez-Cordón[1], Juan A Pineda[56], Claudia Pisanu[133], Thomas Polak[99], Danielle Posthuma[134], Josef Priller[135,136], Olivier Quenez[51], Inés Quintela[48], Alberto Rábano[24,2], Marcel J.T. Reinders[137],

Peter Riederer[138], Natalia Roberto[1], Arvid Rongve[139,140], Irene Rosas Allende[141,142], Maitée Rosende-Roca[1,2], Jose Luis Royo[143], Elisa Rubino[144], María Eugenia Sáez[86], Paraskevi Sakka[145], Ingvild Saltvedt[35,146], Ángela Sanabria[1,2], María Bernal Sánchez-Arjona[79], Florentino Sanchez-Garcia[147], Sigrid B Sando[34,35], Michela Scamosci[50], Elio Scarpini[16,71], Martin Scherer[148], Jonathan M. Schott[78], Alexey A Shadrin[20], Olivia Skrobot[122], Alina Solomon[95,108], Sandro Sorbi[28,149], Oscar Sotolongo-Grau[1], Annika Spottke[98,150], Eystein Stordal[151], Juan Pablo Tartan[1], Lluís Tárraga[1,2], Niccolo Tesí[102,103], Anbupalam Thalamuthu[11], Tegos Thomas[12], Latchezar Traykov[152], Andre Uitterlinden[153], Abbe Ullgren[105], Ingun Ulstein[154], Sergi Valero[1,2], Christine Van Broeckhoven[155,156,157], Jasper Van Dongen[158,155,156], Rik Vandenberghe[159,160], Jean-Sébastien Vidal[21], Jonathan Vogelgsang[161,162], Michael Wagner[75,98], Leonie Weinhold[163], Gill Windle[123], Bob Woods[123], Mary Yannakoulia[164], Miren Zulaica[2,165]

[1]Research Center and Memory clinic Fundació ACE, Institut Català de Neurociències Aplicades, Universitat Internacional de Catalunya, Barcelona, Spain
[2]CIBERNED, Network Center for Biomedical Research in Neurodegenerative Diseases, National Institute of Health Carlos III, Madrid, Spain
[3]Division of Neurogenetics and Molecular Psychiatry, Department of Psychiatry and Psychotherapy, Faculty of Medicine and University Hospital Cologne, University of Cologne, Cologne, Germany
[4]Estudios en Neurociencias y Sistemas Complejos (ENyS) CONICET-HEC-UNAJ
[5]Centre of Age-Related Medicine, Stavanger University Hospital, Norway
[6]Institute of Psychiatry, Psychology & Neuroscience, PO 70, 16 De Crespigny Park, London, SE58AF
[7]Department of Surgery, Biochemistry and Molecular Biology, School of Medicine, University of Málaga, Málaga, Spain.
[8]Department of Neurology, II B Sant Pau, Hospital de la Santa Creu i Sant Pau, Universitat Autònoma de Barcelona, Barcelona, Spain.
[9]Fundació Docència i Recerca MútuaTerrassa and Movement Disorders Unit, Department of Neurology, University Hospital MútuaTerrassa, Terrassa 08221, Barcelona, Spain
[10]Memory Disorders Unit, Department of Neurology, Hospital Universitari Mutua de Terrassa, Terrassa, Barcelona, Spain.
[11]Centre for Healthy Brain Ageing, School of Psychiatry, Faculty of Medicine, University of New South Wales, Sydney, Australia
[12]1st Department of Neurology, Medical school, Aristotle University of Thessaloniki, Thessaloniki, Makedonia, Greece
[13]Alzheimer Hellas, Thessaloniki, Makedonia, Greece
[14]School of Medicine and Surgery, University of Milano-Bicocca, Italy
[15]Neurology Unit, "San Gerardo" hospital, Monza, Italy
[16]Fondazione IRCCS Ca' Granda, Ospedale Policlinico, Milan, Italy
[17]Department of Laboratory Diagnostics, III Laboratory of Analysis, Brescia Hospital, Brescia, Italy
[18]Unitat Trastorns Cognitius, Hospital Universitari Santa Maria de Lleida, Lleida, Spain
[19]Institut de Recerca Biomedica de Lleida (IRBLLeida), Lleida, Spain
[20]NORMENT Centre, University of Oslo, Oslo, Norway
[21]Université de Paris, EA 4468, APHP, Hôpital Broca, Paris, France
[22]Laboratory of Neuropsychiatry, Department of Clinical and Behavioral Neurology, IRCCS Santa Lucia Foundation, Rome, Italy
[23]Servei de Neurologia, Hospital Universitari i Politècnic La Fe, Valencia, Spain.
[24]Reina Sofia Alzheimer Center, CIEN Foundation, ISCIII, Madrid, Spain
[25]Molecular Markers Laboratory, IRCCS Istituto Centro San Giovanni di Dio Fatebenefratelli, Brescia
[26]Univ. Montpellier, Inserm U1061, Neuropsychiatry: epidemiological and clinical research, PSNREC, Montpellier, France
[27]Université Paris-Saclay, CEA, Centre National de Recherche en Génomique Humaine, 91057, Evry, France
[28]Department of Neuroscience, Psychology, Drug Research and Child Health University of Florence, Florence Italy
[29]Azienda Ospedaliero-Universitaria Careggi, , Florence, Italy

[30]MAC - Memory Clinic, IRCCS Istituto Centro San Giovanni di Dio Fatebenefratelli, Brescia
[31]Geriatrics Unit Fondazione Policlinico A. Gemelli IRCCS, Rome , Italy
[32]Department of Neuroscience “Rita Levi Montalcini”, University of Torino, Torino, Italy
[33]Experimental Neuro-psychobiology Laboratory,Department of Clinical and Behavioral Neurology, IRCCS Santa Lucia Foundation, Rome, Italy
[34]Department of Neurology and Clinical Neurophysiology, University Hospital of Trondheim, Trondheim, Norway
[35]Department of Neuromedicine and Movement Science, Norwegian University of Science and Technology, Trondheim, Norway
[36]MRC Centre for Neuropsychiatric Genetics and Genomics, , Division of Psychological Medicine and Clinical Neuroscience, School of Medicine, Cardiff University, Cardiff, UK
[37]Dementia Centre for Research Collaboration, School of Psychiatry, University of New South Wales, Sydney, Australia
[38]Biosciences, School of Science and Technology, Nottingham Trent University, Nottingham UK
[39]Unidad de Trastornos del Movimiento, Servicio de Neurología y Neurofisiología. Instituto de Biomedicina de Sevilla (IBiS), Hospital Universitario Virgen del Rocío/CSIC/Universidad de Sevilla, Seville, Spain
[40]Institute for Stroke and Dementia Research, Klinikum der Universität München, Ludwig-Maximilians-Universität LMU, Munich, Germany.
[41]German Center for Neurodegenerative Diseases (DZNE, Munich), Munich, Germany.
[42]Faculty of Medical & Health Sciences, University of Auckland, New Zealand
[43]Wales Centre for Ageing & Dementia Research, Swansea University, Wales, New Zealand
[44]CIEN Foundation/Queen Sofia Foundation Alzheimer Center
[45]UFIEC, Instituto de Salud Carlos III, , Madrid, Spain
[46]Inserm, Bordeaux Population Health Research Center, UMR 1219, Univ. Bordeaux, ISPED, CIC 1401-EC, Univ Bordeaux,Bordeaux, France
[47]CHU de Bordeaux, Pole santé publique, Bordeaux, France
[48]Grupo de Medicina Xenómica, Centro Nacional de Genotipado (CEGEN-PRB3-ISCIII). Universidade de Santiago de Compostela, Santiago de Compostela, Spain.
[49]Fundación Pública Galega de Medicina Xenómica- CIBERER-IDIS, University of Santiago de Compostela, Santiago de Compostela, Spain.
[50]Institute of Gerontology and Geriatrics, Department of Medicine and Surgery, University of Perugia Perugia (Italy)
[51]Normandie Univ, UNIROUEN, Inserm U1245 and CHU Rouen, Department of Genetics and CNR-MAJ, Rouen, France
[52]Unit of Clinical Pharmacology, University Hospital of Cagliari, Cagliari, Italy
[53]Geriatic Unit, Fondazione Cà Granda, IRCCS Ospedale Maggiore Policlinico, Milan, Italy
[54]Radboudumc Alzheimer Center, Department of Geriatrics, Radboud University Medical Center, Nijmegen, the Netherlands
[55]Institute for Regenerative Medicine, University of Zürich, Schlieren, Switzerland
[56]Unidad Clínica de Enfermedades Infecciosas y Microbiología. Hospital Universitario de Valme, Sevilla, Spain
[57]Department of Neuroscience, Università Cattolica del Sacro Cuore, Rome, Italy
[58]Neurology Unit, IRCCS Fondazione Policlinico Universitario A. Gemelli, Rome, Italy
[59]University of Bari, “A. Moro”
[60]School of Medicine, University of Thessaly, Larissa, Greece
[61]University Bordeaux, Inserm, Bordeaux Population Health Research Center, France
[62]Department of Neurology, University Medical Center Groningen, the Netherlands
[63]UKDRI@ Cardiff, School of Medicine, Cardiff University, Cardiff, UK
[64]Technical University of Munich, School of Medicine, Klinikum rechts der Isar, Department of Psychiatry and Psychotherapy
[65]Center for Neurosciences, Vrije Universiteit Brussel (VUB), Brussels, Belgium
[66]Reference Center for Biological Markers of Dementia (BIODEM), Institute Born-Bunge, University of Antwerp, Antwerp, Belgium

[67]Institute Born-Bunge, University of Antwerp, Antwerp, Belgium
[68]Department of Neurology, UZ Brussel, Brussels, Belgium
[69]Fondazione IRCCS, Istituto Neurologico Carlo Besta, Milan Italy
[70]Centre for Memory Disturbances, Lab of Clinical Neurochemistry, Section of Neurology, University of Perugia, Italy
[71]University of Milan, Milan, Italy
[72]Faculty of Medicine, University of Lisbon, Portugal
[73]Department of Psychiatry, Social Medicine Center East- Donauspital, Vienna, Austria
[74]Institute of Clinical Medicine, University of Oslo, Oslo, Norway.
[75]Department of Neurodegeneration and Geriatric Psychiatry, University of Bonn, 53127 Bonn, Germany.
[76]German Center for Neurodegenerative Diseases (DZNE Bonn), Bonn, Germany
[77]Department of Medical and Molecular Genetics, Indiana University, Indianapolis, Indiana, USA
[78]Dementia Research Centre, UCL Queen Square Institute of Neurology, London, United Kingdom
[79]Unidad de Demencias, Servicio de Neurología y Neurofisiología. Instituto de Biomedicina de Sevilla (IBiS), Hospital Universitario Virgen del Rocío/CSIC/Universidad de Sevilla, Seville, Spain
[80]Instituto de Investigacion Sanitaria 'Hospital la Paz' (IdIPaz), Madrid, Spain
[81]Hospital Universitario la Paz, Madrid, Spain
[82]Department of geriatric Psychiatry, Central Institute for Mental Health, Mannheim, University of Heidelberg, Germany
[83]Alzheimer Research Center & Memory Clinic, Andalusian Institute for Neuroscience, Málaga, Spain.
[84]Hospital Universitario Ramon y Cajal, IRYCIS, Madrid
[85]Department of Psychiatry and Psychotherapy, Medical University of Vienna, Vienna, Austria
[86]CAEBI, Centro Andaluz de Estudios Bioinformáticos, Sevilla, Spain.
[87]Aging Research Center, Department of Neurobiology, Care Sciences and Society, Karolinska Institutet and Stockholm University, Stockholm, Sweden
[88]Institute of Public Health, University of Cambridge, UK
[89]Human Genetics, School of Life Sciences, Life Sciences Building, University Park, University of Nottingham, Nottingham, UK
[90]A.I Virtanen Institute for Molecular Sciences, University of Eastern Finland, Kuopio, Finland
[91]Department of Neurology, Medical School, University of Cyprus, Cyprus
[92]Sorbonne University, GRC n° 21, Alzheimer Precision Medicine Initiative (APMI), AP-HP, Pitié-Salpêtrière Hospital, Boulevard de l'hôpital, F-75013, Paris, France
[93]Eisai Inc., Neurology Business Group, 100 Tice Blvd, Woodcliff Lake, NJ 07677, USA
[94]Reta Lila Weston Research Laboratories, Department of Molecular Neuroscience, UCL Institute of Neurology, London, UK.
[95]Insitute of Clinical Medicine - Neurology, University of Eastern, Kuopio, Finland
[96]Institute of Clinical Medicine – Internal Medicine, University of Eastern Finland, Kuopio, Finland
[97]Department of Neurodegeneration and Geriatric Psychiatry, University of Bonn, Bonn, Germany.
[98]German Center for Neurodegenerative Diseases (DZNE, Bonn), Bonn, Germany.
[99]Department of Psychiatry, Psychosomatics and Psychotherapy, Center of Mental Health, University Hospital, Wuerzburg
[100]Institute of Human Genetics, University of Bonn, School of Medicine & University Hospital Bonn, Bonn, Germany
[101]Clinical and Experimental Science, Faculty of Medicine, University of Southampton, Southampton, UK.
[102]Alzheimer Center Amsterdam, Department of Neurology, Amsterdam Neuroscience, Vrije Universiteit Amsterdam, Amsterdam UMC, Amsterdam, The Netherlands
[103]Section Genomics of Neurdegenerative Diseases and Aging, Department of Human Genetics Amsterdam UMC, Vrije Universiteit Amsterdam, Amsterdam UMC, Amsterdam, The Netherlands
[104]Department of Neurology, UMC Utrecht Brain Center, Utrecht, the Netherlands
[105]Karolinska Institutet, Center for Alzheimer Research, Department NVS, Division of Neurogeriatrics, Stockholm, Sweden
[106]Unit for Hereditary dementias, Karolinska University Hospital-Solna, Stockholm, Sweden

[107]Dept.of Public Health and Carins Sciences / Geriatrics, Uppsala University
[108]Division of Clinical Geriatrics, Center for Alzheimer Research, Care Sciences and Society (NVS), Karolinska Institutet, Stockholm, Sweden
[109]Institute of Public Health and Clinical Nutrition, University of Eastern Finland, Kuopio, Finland
[110]Neuroepidemiology and Ageing Research Unit, School of Public Health, Imperial College London, London, United Kingdom
[111]Stockholms Sjukhem, Research & Development Unit, Stockholm, Sweden
[112]Department of Neurology, Kuopio University Hospital, Kuopio, Finland
[113]Department of Neurosciences, University of Helsinki and Department of Geriatrics, Helsinki University Hospital, Helsinki, Finland
[114]Department of Psychiatry and Psychotherapy, Universitätsklinikum Erlangen, and Friedrich-Alexander Universität Erlangen-Nürnberg, Erlangen, Germany.
[115]Laboratory of Cognitive Neuroscience, School of Psychology, Aristotle University of Thessaloniki, Thessaloniki, Greece
[116]Neurology Service, Marqués de Valdecilla University Hospital (University of Cantabria and IDIVAL), Santander, Spain.
[117]Stockholm Gerontology Research Center, Stockholm, Sweden
[118]Public Health Promotion Unit, Finnish Institute for Health and Welfare, Helsinki, Finland
[119]Memory Clinic, Department of Neurology, Charles University, 2nd Faculty of Medicine and Motol University Hospital, Czech Republic
[120]International Clinical Research Center, St. Anne's University Hospital Brno, Brno, Czech Republic
[121]Department of Neurology. Hospital Universitario Donostia. OSAKIDETZA-Servicio Vasco de Salud, San Sebastian, Spain
[122]Translational Health Sciences, Bristol Medical School, University of Bristol, Bristol, BS16 1LE, UK
[123]School of Health Sciences, Bangor University, UK
[124]Department of Neurodegenerative Diseases and Geriatric Psychiatry, University Hospital Bonn, Bonn, Germany
[125]Unit of Neurology, University of Parma and AOU, Parma, Italy
[126]Servicio de Neurología HOspital Universitario Central de Asturias- Oviedo and Instituto de Investigación Biosanitaria del Principado de Asturias, Oviedo, Spain
[127]MRC Prion Unit at UCL, UCL Institute of Prion Diseases, London, W1W 7FF
[128]Department of Neurology, ErasmusMC
[129]Human Genetics, School of Life Sciences, University of Nottingham, UK NG7 2UH
[130]Department of Neuroscience, Catholic University of Sacred Heart, Fondazione Policlinico Universitario A. Gemelli IRCCS, Rome, Italy
[131]Centre for Neurodegenerative Disorders, Department of Clinical and Experimental Sciences, University of Brescia, Brescia, Italy
[132]DIMEC, University of Parma, Parma, Italy
[133]Department of Biomedical Sciences, University of Cagliari, Italy
[134]Department of Complex Trait Genetics, Center for Neurogenomics and Cognitive Research, Amsterdam Neuroscience, Vrije University, Amsterdam, The Netherlands.
[135]German Center for Neurodegenerative Diseases (DZNE), Berlin, Germany.
[136]Department of Neuropsychiatry and Laboratory of Molecular Psychiatry, Charité, Charitéplatz 1, 10117 Berlin, Germany
[137]Delft Bioinformatics Lab, Delft University of Technology, Delft, The Netherlands
[138]Center of Mental Health, Clinic and Policlinic of Psychiatry, Psychosomatics and Psychotherapy, University Hospital of Würzburg, Wuerzburg, Germany
[139]Department of Research and Innovation, Helse Fonna, Haugesund Hospital, Haugesund, Norway.
[140]The University of Bergen, Institute of Clinical Medicine (K1), Bergen Norway
[141]Laboratorio de Genética. Hospital Universitario Central de Asturias, Oviedo, Spain
[142]Instituto de Investigación Sanitaria del Principado de Asturias (ISPA)

[143]Departamento de Especialidades Quirúrgicas, Bioquímicas e Inmunología, School of Medicine, University of Málaga, Málaga, Spain.
[144]Department of Neuroscience and Mental Health, AOU Città della Salute e della Scienza di Torino, Torino, Italy
[145]Athens Association of Alzheimer’s disease and Related Disorders, Athens, Greece
[146]Department of Geriatrics, St. Olav’s Hospital, Trondheim University Hospital, Norway
[147]Department of Immunology, Hospital Universitario Doctor Negrín, Las Palmas de Gran Canaria, Spain.
[148]Department of Primary Medical Care, University Medical Centre Hamburg-Eppendorf, 20246 Hamburg, Germany.
[149]IRCCS Fondazione Don Carlo Gnocchi, Florence, Italy
[150]Department of Neurology, University of Bonn, Bonn, Germany.
[151]Department of Psychiatry, Namsos Hospital, Namsos, Norway
[152]Clinic of Neurology, UH "Alexandrovska", Medical University - Sofia, Sofia, Bulgaria
[153]Department of Internal medicine and Biostatistics, ErasmusMC
[154]Department of Geriatric Medicine, Oslo University Hospital, Oslo, Norway
[155]Laboratory of Neurogenetics, Institute Born - Bunge, Antwerp, Belgium
[156]Department of Biomedical Sciences, University of Antwerp, Neurodegenerative Brain Diseases Group, Center for Molecular Neurology, VIB, Antwerp, Belgium
[157]Neurodegenerative Brain Diseases Group, VIB Center for Molecular Neurology, VIB, Antwerp, Belgium
[158]Complex Genetics of Alzheimer's Disease Group, VIB Center for Molecular Neurology, VIB, Antwerp, Belgium
[159]Laboratory for Cognitive Neurology, Department of Neurosciences, University of Leuven, Belgium
[160]Neurology Department, University Hospitals Leuven, Leuven, Belgium
[161]Department of Psychiatry and Psychotherapy, University Medical Center Goettingen, Goettingen, Germany
[162]Department of Psychiatry, Harvard Medical School, McLean Hospital, Belmont, MA, USA
[163]Institute of Medical Biometry, Informatics and Epidemiology, University Hospital of Bonn, Bonn, Germany.
[164]Department of Nutrition and Diatetics, Harokopio University, Athens, Greece
[165]Neurosciences Area. Instituto Biodonostia. San Sebastian, Spain

**EADI**

Céline Bellenguez[1], Benjamin Grenier-Boley[1], Jacques Epelbaum[2], David Wallon[3], Didier Hannequin[3], Florence Pasquier[4], Claudine Berr[5], Jean-Francois Dartigues[6], Dominique campion[7], Christophe Tzourio[8], Vincent Dermecourt[4], Nathalie Fievet[1], Olivier Hanon[9], Carole Dufouil[8], Alexis Brice[10], Bruno Dubois[11], Karen Ritchie[5], Phillippe Amouyel[1], Jean-Charles Lambert[1]

1. Univ. Lille, Inserm, CHU Lille, Institut Pasteur Lille, U1167-RID-AGE - Facteurs de risque et déterminants moléculaires des maladies liées au vieillissement, F-59000 Lille, France
2. UMR 894, Center for Psychiatry and Neuroscience, INSERM, Université Paris Descartes, F-75000 Paris, France
3. Normandie Univ, UNIROUEN, Inserm U1245, CHU Rouen, Department of Neurology and CNR-MAJ, F 76000, Normandy Center for Genomic and Personalized Medicine, Rouen, France
4. Univ. Lille, Inserm, CHU Lille, UMR1172, Resources and Research Memory Center (MRRC) of Distalz, Licend, Lille France
5. Univ. Montpellier, Inserm U1061, Neuropsychiatry: epidemiological and clinical research, PSNREC, Montpellier, France
6. University Bordeaux, Inserm, Bordeaux Population Health Research Center, France
7. Normandie Univ, UNIROUEN, Inserm U1245 and CHU Rouen, Department of Genetics and CNR-MAJ, Rouen, France
8. University Bordeaux, Inserm, Bordeaux Population Health Research Center, France
9. Université de Paris, EA 4468, APHP, Hôpital Broca, Paris, France

10. Inserm U1127, CNRS UMR7225, Sorbonne Universités, UPMC Univ Paris 06, UMR_S1127, Institut du Cerveau et de la Moelle épinière, F-75013, Paris, France; APHP, Department of genetics, Pitié-Salpêtrière Hospital, 75013, Paris, France
11. Institut de la Mémoire et de la Maladie d'Alzheimer (IM2A), Département de Neurologie, Hôpital de la Pitié-Salpêtrière, AP-HP, Paris, France; Institut des Neurosciences Translationnelles de Paris (IHU-A-ICM), Institut du Cerveau et de la Moelle Epinière (ICM), Paris, France; INSERM, CNRS, UMR-S975, Institut du Cerveau et de la Moelle Epinière (ICM), Paris, France; Sorbonne Universités, Université Pierre et Marie Curie, Hôpital de la Pitié-Salpêtrière, AP-HP, Paris, France

**DEGESCO consortium**

Adarmes-Gómez Astrid Daniela[1,2], Aguilar Miquel[3,4], Aguilera Nuria[5], Alcolea Daniel[6,2], Alegret Montserrat[5,7], Alonso María Dolores[8], Alvarez Ignacio[3,4], Álvarez Victoria[9,10], Amer-Ferrer Guillermo[11], Antequera Martirio[12], Antonell Anna[13], Antúnez Carmen[14], Arias Pastor Alfonso[15,16], Baquero Miquel[17], Belbin Olivia[6,2], Bernal Sánchez-Arjona María[18], Boada Mercè[5,7], Buendia Mar[5], Buiza-Rueda Dolores[1,2], Bullido María Jesús[19,2,20,21], Buongiorno Mariateresa[3,4], Calero Miguel[22,2,23], Camara Ana[24,2], Cano Amanda[5], Cañabate Pilar[5], Cardona Serrate Fernando[25,2,26], Carracedo Ángel[27,28], Carrillo Fátima[1,2], Casajeros María José[29], Chafer Pericás Consuelo[17], Compta Yaroslau[24], Corbatón-Anchuelo Arturo[30,31], Corma-Gómez Anaïs[32], De la Guía Paz[33], de Rojas Itziar[5,7], del Ser Teodoro[34], Díaz Belloso Rafael[1,2], Diego Susana[5], Diez-Fairen Mónica[3,4], Dols-Icardo Oriol[6,2], Erro Aguirre María Elena[35,36,37], Espinosa Ana[5,7], Fernandez Manuel[24,2,38], Fernández Maria Victoria[5], Fernández-Fuertes Marta[32], Fortea Juan[6,2], Franco-Macías Emilio[18,2], Frank-García Ana[39,2,40,21], Gailhajenet Anna[5], García-Alberca Jose María[33], García-González Pablo[5], García-Madrona Sebastián[29,41], Garcia-Ribas Guillermo[29,41], Garrote-Espina Lorena[1,2], Gómez-Garre Pilar[1,2], Guitart Marina[5], Huerto Vilas Raquel[15,16], Ibarria Marta[5], Illán-Gala Ignacio[42,2], Jesús Silvia[1,2], Kulisevsky Jaime[43,2], Labrador Espinosa Miguel Angel[1,2], Lafuente Asunción[5], Lage Carmen[44,2], Legaz Agustina[12], Lleó Alberto[6,2], López-García Sara[45,2], Lopez de Munain Adolfo[46,47,2,48], Macías Juan[32], Macias-García Daniel[1,2], Manzanares Salvadora[12], Marín Marta[18], Marín-Muñoz Juan[12], Maroñas Olalla[49], Marquié Marta[5,7], Marti Maria[24], Martín Elvira[5], Martín Montés Angel[50,2,40], Martínez Begoña[12], Martínez Victoriana[12], Martínez-Lage Álvarez Pablo[51], Martínez-Larrad María Teresa[30,31], Martinez de Pancorbo Marian[52], Martínez Rodríguez Carmen[53,10], Medina Miguel[2], Mendioroz Iriarte Maite[35,36,37], Mendoza Silvia[33], Menéndez-González Manuel[54,10,55], Mir Pablo[1,2,56], Molina-Porcel Laura[57,13], Montrreal Laura[5], Moreno Mariola[5], Moreno Fermin[46,2,48], Muñoz Esteban[24], Muñoz-Delgado Laura[1,2], Noguera Perea Fuensanta[12], Olivé Clàudia[5], Ortega Gemma[5,7], Pagonabarraga Javier[43,2], Painous Celia[24], Pancho Ana[5], Pastor Ana Belén[58,59], Pastor Pau[60,61], Pelejá Ester[5], Pérez-Cordón Alba[5], Pérez-Tur Jordi[25,2,26], Periñán María Teresa[1,2], Pineda Juan Antonio[32], Pineda-Sánchez Rocío[1,2], Piñol-Ripoll Gerard[15,16], Preckler Silvia[5], Puerta Raquel[5], Quintela Inés[62], Rábano Alberto[58,59,2], Real Luis Miguel[32], Real de Asúa Diego[63], Rodriguez-Rodriguez Eloy[44,2], Rosas Allende Irene[9,10], Rosende-Roca Maitée[5,7], Royo Jose Luís[64], Ruiz Agustín[5,7,65], Sáez María Eugenia[66], Sanabria Ángela[5,7], Sánchez-Juan Pascual[67,2], Sánchez-Valle Raquel[13], Sánchez Ruiz de Gordoa Javier[35], Sastre Isabel[19,2], Sotolongo-Grau Oscar[5], Tárraga Lluís[5,7], Valero Sergi[5,7], Valldeoriola Francesc[24], Vargas Liliana[5], Vicente María Pilar[12], Vivancos-Moreau Laura[12]

[1]Unidad de Trastornos del Movimiento, Servicio de Neurología y Neurofisiología. Instituto de Biomedicina de Sevilla (IBiS), Hospital Universitario Virgen del Rocío/CSIC/Universidad de Sevilla, Seville, Spain, [2]CIBERNED, Network Center for Biomedical Research in Neurodegenerative Diseases, National Institute of Health Carlos III, Madrid, Spain, [3]Fundació Docència i Recerca MútuaTerrassa, Terrassa, Barcelona, Spain, [4]Memory Disorders Unit, Department of Neurology, Hospital Universitari Mutua de Terrassa, Terrassa, Barcelona, Spain, [5]Ace Alzheimer Center Barcelona, Universitat Internacional de Catalunya (UIC), Barcelona, Spain., [6]Department of Neurology, II B Sant Pau, Hospital de la Santa Creu i Sant Pau, Universitat Autònoma de Barcelona, Barcelona, Spain., [7]Networking Research Center on Neurodegenerative Diseases (CIBERNED), Instituto de Salud Carlos III, Madrid, Spain., [8]Servei de Neurologia. Hospital Clínic Universitari de València, [9]Laboratorio de Genética. Hospital Universitario Central de Asturias, Oviedo, Spain, [10]Instituto de Investigación Sanitaria del Principado de Asturias (ISPA), [11]Department of Neurology, Hospital Universitario

Son Espases, Palma, Spain, [12]Unidad de Demencias. Hospital Clínico Universitario Virgen de la Arrixaca, Palma, Spain, [13]Alzheimer's disease and other cognitive disorders unit. Service of Neurology. Hospital Clínic of Barcelona. Institut d'Investigacions Biomèdiques August Pi i Sunyer, University of Barcelona, Barcelona, Spain, [14]Unidad de Demencias, Hospital Clínico Universitario Virgen de la Arrixaca, Murcia, Spain., [15]Unitat Trastorns Cognitius, Hospital Universitari Santa Maria de Lleida, Lleida, Spain, [16]Institut de Recerca Biomedica de Lleida (IRBLLeida), Lleida, Spain, [17]Servei de Neurologia, Hospital Universitari i Politècnic La Fe, Valencia, Spain, [18]Unidad de Demencias, Servicio de Neurología y Neurofisiología. Instituto de Biomedicina de Sevilla (IBiS), Hospital Universitario Virgen del Rocío/CSIC/Universidad de Sevilla, Seville, Spain, [19]Centro de Biología Molecular Severo Ochoa (UAM-CSIC), [20]Instituto de Investigacion Sanitaria 'Hospital la Paz' (IdIPaz), Madrid, Spain, [21]Universidad Autónoma de Madrid, [22]CIEN Foundation/Queen Sofia Foundation Alzheimer Center/Instituto de Salud Carlos III, [23]UFIEC, Instituto de Salud Carlos III, [24]Parkinson's Disease & Movement Disorders Unit, Neurology Service, Hospital Clínic I Universitari de Barcelona; IDIBAPS, CIBERNED (CB06/05/0018-ISCIII), ERN- RND, Institut Clínic de Neurociències (Maria de Maeztu Excellence Centre), Universitat de Barcelona. Barcelona, Catalonia, Spain, [25]Unitat de Genètica Molecular, Institut de Biomedicina de València-CSIC, Valencia, Spain, [26]Unidad Mixta de Neurologia Genètica, Instituto de Investigación Sanitaria La Fe, Valencia, Spain., [27]Grupo de Medicina Xenómica, CIBERER, CIMUS. Universidade de Santiago de Compostela, Santiago de Compostela, Spain., [28]Fundación Pública Galega de Medicina Xenómica- IDIS, Santiago de Compostela, Spain., [29]Hospital Universitario Ramon y Cajal, IRYCIS, Madrid, [30]Instituto de Investigación Sanitaria, Hospital Clínico San Carlos (IdISSC), Madrid, Spain, [31]Spanish Biomedical Research Centre in Diabetes and Associated Metabolic Disorders(CIBERDEM), Madrid, Spain, [32]Unidad Clínica de Enfermedades Infecciosas y Microbiología. Hospital Universitario de Valme, Sevilla, Spain, [33]Alzheimer Research Center & Memory Clinic, Instituto Andaluz de Neurociencia, Málaga, Spain., [34]Department of Neurology/CIEN Foundation/Queen Sofia Foundation Alzheimer Center, [35]Navarrabiomed, Pamplona, Spain, [36]Department of Neurology, Hospital Universitario de Navarra, Pamplona, Spain, [37]Instituto de Investigación Sanitaria de Navarra (IDISNA), Pamplona, Spain, [38]Universitat de Barcelona (UB), [39]Department of Neurology, La Paz University Hospital. Instituto de Investigación Sanitaria del Hospital Universitario La Paz. IdiPAZ., [40]Hospital La Paz Institute for Health Research, IdiPAZ, Madrid, Spain, [41]None, [42]Sant Pau Memory Unit, Department of Neurology, Institut de Recerca de Sant Pau; Hospital de Sant Pau, Barcelona, Spain, [43]Movement Disorders Unit, Department of Neurology, Institut de Recerca de Sant Pau; Hospital de Sant Pau, Barcelona, Spain, [44]Neurology Service, Marqués de Valdecilla University Hospital (University of Cantabria and IDIVAL), Santander, Spain., [45]Service of Neurology, University Hospital Marqués de Valdecilla, IDIVAL, University of Cantabria, Santander, Spain, [46]Department of Neurology. Hospital Universitario Donostia. San Sebastian, Spain, [47]Department of Neurosciences. Faculty of Medicine and Nursery. University of the Basque Country, San Sebastián, Spain, [48]Neurosciences Area. Instituto Biodonostia. San Sebastian, Spain, [49]Grupo de Medicina Xenómica, Centro Nacional de Genotipado (CEGEN-PRB3-ISCIII). Universidade de Santiago de Compostela, Santiago de Compostela, Spain., [50]Department of Neurology, La Paz University Hospital, [51]Centro de Investigación y Terapias Avanzadas. Fundación CITA-alzheimer, San Sebastian, Spain, [52]BIOMICS País Vasco; Centro de investigación Lascaray, Universidad del País Vasco UPV/EHU, Vitoria-Gasteiz, Spain, [53]Hospital de Cabueñes, Gijón, Spain, [54]Servicio de Neurología. Hospital Universitario Central de Asturias, Oviedo, Spain, [55]Departamento de Medicina, Universidad de Oviedo, Oviedo, Spain, [56]Departamento de Medicina, Facultad de Medicina, Universidad de Sevilla, Seville, Spain., [57]Neurological Tissue Bank of the Biobanc-Hospital Clinic-IDIBAPS, Institut d'Investigacions Biomèdiques August Pi i Sunyer, Barcelona, Spain, [58]CIEN Foundation/Queen Sofia Foundation Alzheimer Center, [59]BT-CIEN, [60]Unit of Neurodegenerative diseases, Department of Neurology, Hospital Germans Trias i Pujol , Badalona, Barcelona, [61]Neurodegenerative Diseases Research Laboratory, Germans Trias i Pujol Research Laboratory, Badalona, Barcelona, [62]Grupo de Medicina Xenómica, Fundación Pública Galega de Medicina Xenómica, Santiago de Compostela, Spain., [63]Hospital Universitario La Princesa, Madrid, Spain, [64]Departamento de Especialidades Quirúrgicas, Bioquímica e Inmunología. School of Medicine. University of Malaga. Málaga, Spain, [65]Biggs Institute for Alzheimer's and Neurodegenerative Diseases, University of Texas Health Science Center, San Antonio, Texas, USA., [66]CAEBI, Centro Andaluz de Estudios Bioinformáticos, Sevilla, Spain., [67]Alzheimer's Centre Reina Sofia-CIEN Foundation-ISCIII, Madrid, Spain

**FinnGen**

Aarno Palotie[1], Mark Daly[1], Bridget Riley-Gills[2], Howard Jacob[2], Coralie Viollet[3], Slavé Petrovski[3], Chia-Yen Chen[4], Sally John[4], George Okafo[5], Robert Plenge[6], Joseph Maranville[6], Mark McCarthy[7], Rion Pendergrass[7], Jonathan Davitte[8], Kirsi Auro[9], Simonne Longerich[10], Anders Mälarstig[11], Anna Vlahiotis[11], Katherine Klinger[12], Clement Chatelain[12], Jorg Blankenstein[12], Karol Estrada[13], Robert Graham[13], Dawn Waterworth[14], Chris O ´Donnell[15], Nicole Renaud[15], Tomi P. Mäkelä[16], Jaakko Kaprio[17], Minna Ruddock[18], Petri Virolainen[19], Antti Hakanen[19], Terhi Kilpi[20], Markus Perola[20], Jukka Partanen[21], Taneli Raivio[22], Jani Tikkanen[23], Raisa Serpi[23], Kati Kristiansson[24], Veli-Matti Kosma[25], Jari Laukkanen[26], Marco Hautalahti[27], Outi Tuovila[28], Jeffrey Waring[2], Bridget Riley-Gillis[2], Fedik Rahimov[2], Ioanna Tachmazidou[3], Zhihao Ding[5], Marc Jung[5], Hanati Tuoken[5], Shameek Biswas[6], Neha Raghavan[10], Adriana Huertas-Vazquez[10], Jae-Hoon Sul[10], Xinli Hu[11], Åsa Hedman[11], Ma´en Obeidat[15], Jonathan Chung[15], Jonas Zierer[15], Mari Niemi[15], Samuli Ripatti[17], Johanna Schleutker[29], Mikko Arvas[21], Olli Carpén[22], Reetta Hinttala[23], Johannes Kettunen[23], Arto Mannermaa[25], Katriina Aalto-Setälä[30], Mika Kähönen[24], Johanna Mäkelä[27], Reetta Kälviäinen[31], Valtteri Julkunen[31], Hilkka Soininen[31], Anne Remes[32], Mikko Hiltunen[33], Jukka Peltola[34], Minna Raivio[35], Pentti Tienari[35], Juha Rinne[36], Roosa Kallionpää[36], Juulia Partanen[37], Adam Ziemann[2], Nizar Smaoui[2], Anne Lehtonen[2], Susan Eaton[4], Heiko Runz[4], Sanni Lahdenperä[4], Natalie Bowers[7], Edmond Teng[7], Fanli Xu[38], David Pulford[39], Laura Addis[38], John Eicher[38], Qingqin S Li[40], Karen He[14], Ekaterina Khramtsova[14], Martti Färkkilä[35], Jukka Koskela[35], Sampsa Pikkarainen[35], Airi Jussila[34], Katri Kaukinen[34], Timo Blomster[32], Mikko Kiviniemi[31], Markku Voutilainen[36], Tim Lu[7], Linda McCarthy[38], Amy Hart[14], Meijian Guan[14], Jason Miller[10], Kirsi Kalpala[11], Melissa Miller[11], Kari Eklund[35], Antti Palomäki[36], Pia Isomäki[34], Laura Pirilä[36], Oili Kaipiainen-Seppänen[31], Johanna Huhtakangas[32], Nina Mars[17], Apinya Lertratanakul[2], Marla Hochfeld[6], Jorge Esparza Gordillo[38], Fabiana Farias[10], Nan Bing[11], Tarja Laitinen[34], Margit Pelkonen[31], Paula Kauppi[35], Hannu Kankaanranta[41], Terttu Harju[32], Riitta Lahesmaa[36], Hubert Chen[7], Joanna Betts[38], Rajashree Mishra[38], Majd Mouded[42], Debby Ngo[42], Teemu Niiranen[43], Felix Vaura[43], Veikko Salomaa[43], Kaj Metsärinne[36], Jenni Aittokallio[36], Jussi Hernesniemi[34], Daniel Gordin[35], Juha Sinisalo[35], Marja-Riitta Taskinen[35], Tiinamaija Tuomi[35], Timo Hiltunen[35], Amanda Elliott[44], Mary Pat Reeve[17], Sanni Ruotsalainen[17], Dirk Paul[3], Audrey Chu[38], Dermot Reilly[45], Mike Mendelson[46], Jaakko Parkkinen[11], Tuomo Meretoja[47], Heikki Joensuu[48], Johanna Mattson[35], Eveliina Salminen[35], Annika Auranen[49], Peeter Karihtala[48], Päivi Auvinen[31], Klaus Elenius[36], Esa Pitkänen[17], Relja Popovic[2], Margarete Fabre[50], Jennifer Schutzman[7], Diptee Kulkarni[38], Alessandro Porello[14], Andrey Loboda[10], Heli Lehtonen[11], Stefan McDonough[11], Sauli Vuoti[51], Kai Kaarniranta[52], Joni A Turunen[53], Terhi Ollila[35], Hannu Uusitalo[34], Juha Karjalainen[17], Mengzhen Liu[2], Stephanie Loomis[4], Erich Strauss[7], Hao Chen[7], Kaisa Tasanen[32], Laura Huilaja[32], Katariina Hannula-Jouppi[35], Teea Salmi[34], Sirkku Peltonen[36], Leena Koulu[36], David Choy[7], Ying Wu[11], Pirkko Pussinen[35], Aino Salminen[35], Tuula Salo[35], David Rice[35], Pekka Nieminen[35], Ulla Palotie[35], Maria Siponen[31], Liisa Suominen[31], Päivi Mäntylä[31], Ulvi Gursoy[36], Vuokko Anttonen[32], Kirsi Sipilä[54], Hannele Laivuori[17], Venla Kurra[34], Laura Kotaniemi-Talonen[34], Oskari Heikinheimo[35], Ilkka Kalliala[35], Lauri Aaltonen[35], Varpu Jokimaa[36], Marja Vääräsmäki[32], Outi Uimari[32], Laure Morin-Papunen[32], Maarit Niinimäki[32], Terhi Piltonen[32], Katja Kivinen[17], Elisabeth Widen[17], Taru Tukiainen[17], Niko Välimäki[55], Eija Laakkonen[56], Jaakko Tyrmi[57], Heidi Silven[58], Eeva Sliz[58], Riikka Arffman[58], Susanna Savukoski[58], Triin Laisk[59], Natalia Pujol[59], Janet Kumar[8], Iiris Hovatta[60], Erkki Isometsä[35], Hanna Ollila[17], Jaana Suvisaari[43], Antti Mäkitie[61], Argyro Bizaki-Vallaskangas[34], Sanna Toppila-Salmi[62], Tytti Willberg[36], Elmo Saarentaus[17], Antti Aarnisalo[35], Elisa Rahikkala[32], Kristiina Aittomäki[63], Fredrik Åberg[64], Mitja Kurki[65], Aki Havulinna[66], Juha Mehtonen[17], Priit Palta[17], Shabbeer Hassan[17], Pietro Della Briotta Parolo[17], Wei Zhou[67], Mutaamba Maasha[67], Susanna Lemmelä[17], Manuel Rivas[68], Aoxing Liu[17], Arto Lehisto[17], Andrea Ganna[17], Vincent Llorens[17], Henrike Heyne[17], Joel Rämö[17], Satu Strausz[17], Tuula Palotie[69], Kimmo Palin[55], Javier Garcia-Tabuenca[70], Harri Siirtola[70], Tuomo Kiiskinen[17], Jiwoo Lee[65], Kristin Tsuo[65], Kati Hyvärinen[71], Jarmo Ritari[71], Katri Pylkäs[58], Minna Karjalainen[58], Tuomo Mantere[23], Eeva Kangasniemi[24], Sami Heikkinen[33], Nina Pitkänen[19], Samuel Lessard[12], Clément Chatelain[12], Lila Kallio[19], Tiina Wahlfors[20], Eero Punkka[22], Sanna Siltanen[24], Tiina Jokela[26], Anu Jalanko[17], Auli Toivola[17], Huei-Yi Shen[17], Risto Kajanne[17], Rodos Rodosthenous[17], Mervi Aavikko[17], Helen Cooper[17], Denise Öller[17], Rasko Leinonen[72], Henna Palin[24], Malla-Maria Linna[22], Masahiro Kanai[67], Zhili Zheng[67], L. Elisa Lahtela[17], Mari Kaunisto[17], Elina

Kilpeläinen[17], Tianduanyi Wang[17], Timo P. Sipilä[17], Oluwaseun Alexander Dada[17], Awaisa Ghazal[17], Anastasia Kytölä[17], Rigbe Weldatsadik[17], Jaska Uimonen[17], Kati Donner[17], Anu Loukola[22], Päivi Laiho[20], Tuuli Sistonen[20], Essi Kaiharju[20], Markku Laukkanen[20], Elina Järvensivu[20], Sini Lähteenmäki[20], Lotta Männikkö[20], Regis Wong[20], Minna Brunfeldt[20], Sami Koskelainen[20], Tero Hiekkalinna[20], Teemu Paajanen[20], Shuang Luo[17], Shanmukha Sampath Padmanabhuni[17], Marianna Niemi[70], Javier Gracia-Tabuenca[70], Mika Helminen[70], Tiina Luukkaala[70], Iida Vähätalo[70], Iina Laak[17], Saija Haapa-Paananen[73], Sarah Smith[73], Tom Southerington[73], Meri Lähteenmäki[73]

[1]Institute for Molecular Medicine Finland (FIMM), HiLIFE, University of Helsinki, Helsinki, Finland; Broad Institute of MIT and Harvard; Massachusetts General Hospital
[2]Abbvie, Chicago, IL, United States
[3]Astra Zeneca, Cambridge, United Kingdom
[4]Biogen, Cambridge, MA, United States
[5]Boehringer Ingelheim, Ingelheim am Rhein, Germany
[6]Bristol Myers Squibb, New York, NY, United States
[7]Genentech, San Francisco, CA, United States
[8]GlaxoSmithKline, Collegeville, PA, United States
[9]GlaxoSmithKline, Espoo, Finland
[10]Merck, Kenilworth, NJ, United States
[11]Pfizer, New York, NY, United States
[12]Translational Sciences, Sanofi R&D, Framingham, MA, USA
[13]Maze Therapeutics, San Francisco, CA, United States
[14]Janssen Research & Development, LLC, Spring House, PA, United States
[15]Novartis Institutes for BioMedical Research, Cambridge, MA, United States
[16]HiLIFE, University of Helsinki, Finland, Finland
[17]Institute for Molecular Medicine Finland (FIMM), HiLIFE, University of Helsinki, Helsinki, Finland
[18]Arctic biobank / University of Oulu
[19]Auria Biobank / University of Turku / Hospital District of Southwest Finland, Turku, Finland
[20]THL Biobank / Finnish Institute for Health and Welfare (THL), Helsinki, Finland
[21]Finnish Red Cross Blood Service / Finnish Hematology Registry and Clinical Biobank, Helsinki, Finland
[22]Helsinki Biobank / Helsinki University and Hospital District of Helsinki and Uusimaa, Helsinki
[23]Northern Finland Biobank Borealis / University of Oulu / Northern Ostrobothnia Hospital District, Oulu, Finland
[24]Finnish Clinical Biobank Tampere / University of Tampere / Pirkanmaa Hospital District, Tampere, Finland
[25]Biobank of Eastern Finland / University of Eastern Finland / Northern Savo Hospital District, Kuopio, Finland
[26]Central Finland Biobank / University of Jyväskylä / Central Finland Health Care District, Jyväskylä, Finland
[27]FINBB - Finnish biobank cooperative
[28]Business Finland, Helsinki, Finland
[29]Auria Biobank / Univ. of Turku / Hospital District of Southwest Finland, Turku, Finland
[30]Faculty of Medicine and Health Technology, Tampere University, Tampere, Finland
[31]Northern Savo Hospital District, Kuopio, Finland
[32]Northern Ostrobothnia Hospital District, Oulu, Finland
[33]University of Eastern Finland, Kuopio, Finland
[34]Pirkanmaa Hospital District, Tampere, Finland
[35]Hospital District of Helsinki and Uusimaa, Helsinki, Finland
[36]Hospital District of Southwest Finland, Turku, Finland
[37]Institute for Molecular Medicine Finland, HiLIFE, University of Helsinki, Finland
[38]GlaxoSmithKline, Brentford, United Kingdom
[39]GlaxoSmithKline, Stevenage, United Kingdom
[40]Janssen Research & Development, LLC, Titusville, NJ 08560, United States
[41]University of Gothenburg, Gothenburg, Sweden/ Seinäjoki Central Hospital, Seinäjoki, Finland/ Tampere University, Tampere, Finland

[42]Novartis, Basel, Switzerland
[43]Finnish Institute for Health and Welfare (THL), Helsinki, Finland
[44]Institute for Molecular Medicine Finland (FIMM), HiLIFE, University of Helsinki, Helsinki, Finland; Broad Institute, Cambridge, MA, USA and Massachusetts General Hospital, Boston, MA, USA
[45]Janssen Research & Development, LLC, Boston, MA, United States
[46]Novartis, Boston, MA, United States
[47]Department of Breast Surgery, Helsinki University Hospital Comprehensive Cancer Center and University of Helsinki, Helsinki, Finland
[48]Department of Oncology, Helsinki University Hospital Comprehensive Cancer Center and University of Helsinki, Helsinki, Finland
[49]Pirkanmaa Hospital District , Tampere, Finland
[50]AstraZeneca, Cambridge, United Kingdom
[51]Janssen-Cilag Oy, Espoo, Finland
[52]Northern Savo Hospital District, Kuopio, Finland; Department of Molecular Genetics, University of Lodz, Lodz, Poland
[53]Helsinki University Hospital and University of Helsinki, Helsinki, Finland; Eye Genetics Group, Folkhälsan Research Center, Helsinki, Finland
[54]Research Unit of Oral Health Sciences Faculty of Medicine, University of Oulu, Oulu, Finland; Medical Research Center, Oulu, Oulu University Hospital and University of Oulu, Oulu, Finland
[55]University of Helsinki, Helsinki, Finland
[56]University of Jyväskylä, Jyväskylä, Finland
[57]University of Oulu, Oulu, Finland / University of Tampere, Tampere, Finland
[58]University of Oulu, Oulu, Finland
[59]Estonian biobank, Tartu, Estonia
[60]University of Helsinki, Finland
[61]Department of Otorhinolaryngology - Head and Neck Surgery, University of Helsinki and Helsinki University Hospital, Helsinki, Finland
[62]University of Eastern Finland and Kuopio University Hospital, Department of Otorhinolaryngology, Kuopio, Finland and Department of Allergy, Helsinki University Hospital and University of Helsinki, Finland
[63]Department of Medical Genetics, Helsinki University Central Hospital, Helsinki, Finland
[64]Transplantation and Liver Surgery Clinic, Helsinki University Hospital, Helsinki University, Helsinki, Finland
[65]Institute for Molecular Medicine Finland (FIMM), HiLIFE, University of Helsinki, Helsinki, Finland; Broad Institute, Cambridge, MA, United States
[66]Institute for Molecular Medicine Finland (FIMM), HiLIFE, University of Helsinki, Helsinki, Finland; Finnish Institute for Health and Welfare (THL), Helsinki, Finland
[67]Broad Institute, Cambridge, MA, United States
[68]University of Stanford, Stanford, CA, United States
[69]University of Helsinki and Hospital District of Helsinki and Uusimaa, Helsinki, Finland
[70]University of Tampere, Tampere, Finland
[71]Finnish Red Cross Blood Service, Helsinki, Finland
[72]Institute for Molecular Medicine Finland (FIMM), HiLIFE, University of Helsinki, Helsinki, Finland; European Molecular Biology Laboratory, European Bioinformatics Institute, Cambridge, UK
[73]Finnish Biobank Cooperative – FINBB

**GERAD/MRC/UCL-DRC**

Richard Abraham[1], Ammar Al-Chalabi[2], Nicholas J Bass[3], Carol Brayne[4], Kristelle S Brown[5], John Collinge[6], David Craig[7], Carlos Cruchaga[8,9], Panagiotis Deloukas[10], Martin Dichgans[11,12], Nick C Fox[13], Lutz Frölich[14], Amy Gerrish[13], Michael Gill[15,16], Alison M Goate[17,18], Rita Guerreiro[19,20,21], Rhian Gwilliam[10], Harald Hampel[22,23,24], John Hardy[20], Denise Harold[25], Reinhard Heun[26], Isabella Heuser[27], Paul Hollingworth[1], Michael Hüll[28], Frank Jessen[29,26,30], Karl-Heinz Jöckel[31], Janet A Johnston[32], Lesley Jones[1], John SK Kauwe[33], Norman Klopp[34],

Johannes Kornhuber[35], Brian Lawlor[15,16], Gill Livingston[3], Simon Lovestone[36], Michelle K Lupton[37,38], Aoibhinn Lynch[15,16], Wolfgang Maier[39,26], David Mann[40], Kevin Mayo[41], Bernadette McGuinness[32], Andrew McQuillin[3], Simon Mead[6], Susanne Moebus[42], John C Morris[8,9], Markus M Nöthen[43], Michael C O'Donovan[1], Sara Ortega-Cubero[44,45,46], Michael J Owen[1], Peter Passmore[32], John F Powell[37,38], Petra Proitsi[37,38], Martin Rossor[13], David C Rubinsztein[47], Britta Schürmann[48], Christopher E Shaw[2,49], Andrew B Singleton[50], A David Smith[51], Hendrik van den Bussche[52], Jens Wiltfang[53], Dan Rujescu[54], Petra Nowotny[55], H-Erich Wichmann[56,57,58], Hugh Gurling[59], Thomas W Mühleisen[60], Stephen Todd[61], Catherine Mummery[62], Natalie Ryan[62], Lauren Luckcuck[63]

[1]Division of Psychological Medicine and Clinial Neurosciences, MRC Centre for Neuropsychiatric Genetics and Genomics, Cardiff University, UK
[2]Kings College London, Institute of Psychiatry, Psychology and Neuroscience, UK
[3]Division of Psychiatry, University College London, UK
[4]Institute of Public Health, University of Cambridge, Cambridge, UK
[5]Institute of Genetics, Queens Medical Centre, University of Nottingham, Nottingham, UK
[6]MRC Prion Unit at UCL, Institute of Prion Diseases, London
[7]Ageing Group, Centre for Public Health, School of Medicine, Dentistry and Biomedical Sciences, Queen's University Belfast, UK
[8]Department of Psychiatry, Washington University School of Medicine, St. Louis Missouri, USA
[9]Hope Center Program on Protein Aggregation and Neurodegeneration, Washington University School of Medicine, St. Louis, Missouri, USA
[10]The Wellcome Trust Sanger Institute, Wellcome Trust Genome Campus, Hinxton, Cambridge, UK.
[11]Institute for Stroke and Dementia Research, Klinikum der Universität München, Munich, Germany
[12]German Center for Neurodegenerative Diseases (DZNE, Munich), Munich, 80336, Germany
[13]Dementia Research Centre, Department of Neurodegenerative Disease, UCL Institute of Neurology, London, UK
[14]Central Institute of Mental Health, Medical Faculty Mannheim, University of Heidelberg, Germany.
[15]Mercer's Institute for Research on Ageing, St James' Hospital, Dublin 8
[16]James Hospital and Trinity College, Dublin, Ireland
[17]Icahn School of Medicine at Mount Sinai, New York, NY, USA
[18]Hope Center Program on Protein Aggregation and Neurodegeneration, Washington University School of Medicine, St Louis, Missouri, USA
[19]Dementia Research Institute at UCL
[20]Department of Molecular Neuroscience, UCL, Institute of Neurology, London, UK
[21]Department of Medical Sciences, Institute of Biomedicine iBiMED, University of Aveiro, 3810-193 Aveiro, Portugal
[22]AXA Research Fund & UPMC Chair, Paris, France
[23]Sorbonne Universités, Université Pierre et Marie Curie, Paris
[24]Institut de la Mémoire et de la Maladie d'Alzheimer (IM2A) & Institut du Cerveau et de la Moelle épinière (ICM), Département de Neurologie, Hôpital de la Pitié-Salpêtrière, Paris, France
[25]School of Biotechnology, Dublin City University, Dublin, ROI.
[26]Department of Psychiatry and Psychotherapy, University of Bonn, 53127, Bonn, Germany
[27]Department of Psychiatry, Charité Berlin, Germany
[28]Department of Psychiatry, University of Freiburg, Freiburg, Germany (M.H.)
[29]German Center for Neurodegenerative Diseases (DZNE), 53175, Bonn, Germany
[30]Department of Psychiatry and Psychotherapy, University of Cologne, 50937 Cologne, Germany
[31]Institute for Medical Informatics, Biometry and Epidemiology, University Hospital of Essen, University Duisburg-Essen, Hufelandstr. 55, D-45147 Essen, Germany.
[32]Centre for Public Health, School of Medicine, Dentistry and Biomedical Sciences, Queens University, Belfast, UK
[33]Department of Biology, Brigham Young University, Provo, Utah, USA
[34]Institute of Epidemiology, Helmholtz Zentrum München, German Research Center for Environmental Health, Neuherberg, Germany

[35]Department of Psychiatry and Psychotherapy, University of Erlangen-Nuremberg, Germany
[36]Department of Psychiatry, University of Oxford, Oxford, UK
[37]Department of Basic and Clinical Neuroscience, Institute of Psychiatry, Psychology and Neuroscience, Kings College London, London UK; 44
[38]Genetic Epidemiology, QIMR Berghofer Medical Research Institute, Herston, Queensland, Australia
[39]German Center for Neurodegenerative Diseases (DZNE), 53127 Bonn, Germany
[40]Division of Neuroscience and Experimental Psychology, School of Biological Sciences, Faculty of Biology, Medicine and Health, University of Manchester, Manchester Academic Health Science Centre, Manchester M13 9PT, UK
[41]Departments of Psychiatry, Neurology and Genetics, Washington University School of Medicine, St Louis, MO 63110, US.
[42]Institute for Medical Informatics, Biometry and Epidemiology, University Hospital of Essen, University Duisburg-Essen, Hufelandstr, 55, D-45147 Essen, Germany
[43]Institute of Human Genetics, Department of Genomics, Life and Brain Center, University of Bonn, Bonn, Germany
[44]Neurogenetics Laboratory, Division of Neurosciences, Centre for Applied Medical Research, University of Navarra School of Medicine, Pamplona, Spain
[45]CIBERNED, Centro de Investigación Biomédica en Red de Enfermedades Neurodegenerativas, Instituto de Salud Carlos III, Madrid, Spain
[46]Department of Neurology, Complejo Asistencial Universitario de Palencia, Spain
[47]Cambridge Institute for Medical Research and UK Dementia Research Institute, University of Cambridge, Cambridge, UK
[48]Department of Psychiatry and Psychotherapy, University of Bonn, Germany
[49]UK Dementia Research Institute at King's College London, UK
[50]Molecular Genetics Section, Laboratory of Neurogenetics, National Institute on Aging, National Institutes of Health, Bethesda, MD 20892, USA
[51]Oxford Project to Investigate Memory and Ageing (OPTIMA), University of Oxford, Level 4, John Radcliffe Hospital, Oxford OX3 9DU, UK
[52]Institute of Primary Medical Care, University Medical Center Hamburg-Eppendorf, Germany
[53]LVR-Hospital Essen, Department of Psychiatry and Psychotherapy, University Duisburg-Essen, Germany
[54]Alzheimer Memorial Center and Geriatric Psychiatry Branch, Department of Psychiatry, Ludwig-Maximilian University, Munich, Germany
[55]Departments of Psychiatry, Neurology and Genetics, Washington University School of Medicine, St. Louis, Missouri, USA
[56]Institute of Epidemiology, Helmholtz Zentrum München, German Research Center for Environmental Health, Neuherberg, Germany
[57]Institute of Medical Informatics, Biometry and Epidemiology, Ludwig-Maximilians-Universität, Munich, Germany.
[58]Klinikum Grosshadern, Munich, Germany.
[59]Department of Mental Health Sciences, University College London, London, UK
[60]Institute of Human Genetics, University of Bonn, Bonn, Germany.
[61]Ageing Group, Centre for Public Health, School of Medicine, Dentistry and Biomedical Sciences, Queen's University Belfast, Belfast, UK.
[62]Dementia Research Centre, UCL, London, WC1N 3BG
[63]Division of Psychological Medicine and Clinical Neuroscience, School of Medicine, Cardiff University, Cardiff, UK

**The GR@ACE study group**

Aguilera Nuria[1], Alegret Montserrat[1,2], Bein Natali[1], Blazquez-Folch Josep[1], Boada Mercè[1,2], Buendia Mar[1], Cano Amanda[1], Cañabate Pilar[1,2], Capdevila-Bayo Maria[1], Carracedo Angel[3,4], Corbatón-Anchuelo A[5], Casales Federico[1], de Rojas Itziar[1,2], Diego Susana[1], Espinosa Ana[1,2], Fernandez Maria Victoria[1], Gailhajenet Anna[1],

García-González Pablo[1], García-Gutiérrez Fernando[1], Guitart Marina[1], Ibarria Marta[1], Lafuente Asunción[1], Lleonart Núria[1], Macias Juan[6], Maroñas Olalla[3], Martín Elvira[1], Martínez Maria Teresa[5], Marquié Marta[1,2], Miguel Andrea[1], Montrreal Laura[1], Morató Xavier[1,2], Moreno Mariola[1], Muñoz Nathalia[1], Olivé Clàudia[1], Ortega Gemma[1,2], Pancho Ana[1], Pelejà Ester[1], Pérez-Cordon Alba[1], Pineda Juan A[6], Puerta Raquel[1], Preckler Silvia[1], Quintela Inés[3], Real Luis Miguel[6], Rodríguez José Nelet[1], Rosende-Roca Maitee[1],Ruiz Agustín[1,2,7], Sáez Maria Eugenia[8], Sanabria Angela[1,2], Seguer Susanna[1], Sotolongo-Grau Oscar[1], Tárraga Luís[1,2], Tartari Juan Pablo[1], Valero Sergi[1,2], Vargas Liliana[1].

[1]Ace Alzheimer Center Barcelona, Universitat Internacional de Catalunya (UIC), Barcelona, Spain. [2]Networking Research Center on Neurodegenerative Diseases (CIBERNED), Instituto de Salud Carlos III, Madrid, Spain. [3]Grupo de Medicina Xenómica, Centro Nacional de Genotipado (CEGEN-PRB3-ISCIII). Universidad de Santiago de Compostela, Santiago de Compostela, Spain. [4]Fundación Pública Galega de Medicina Xenómica- CIBERER-IDIS, Santiago de Compostela, Spain. [5]Centro de Investigación Biomédica en Red de Diabetes y Enfermedades Metabólicas Asociadas, CIBERDEM, Spain, Hospital Clínico San Carlos, Madrid, Spain. [6]Unidad Clínica de Enfermedades Infecciosas y Microbiología. Hospital Universitario de Valme, Sevilla, Spain. [7]Biggs Institute for Alzheimer's and Neurodegenerative Diseases, University of Texas Health Science Center, San Antonio, Texas, USA. [8]CAEBI. Centro Andaluz de Estudios Bioinformáticos, Sevilla, Spain.

#### 8. Supplementary Acknowledgments

**EADB.** We thank the many study participants, researchers and staff for collecting and contributing to the data, the high-performance computing service at the University of Lille and the staff at CEA-CNRGH for their help with sample preparation and genotyping and excellent technical assistance. We thank Antonio Pardinas for his help. We thank the Netherlands Brain Bank. This research was conducted using the UKBB resource (application number 61054). This work was funded by a grant (EADB) from the EU Joint Programme – Neurodegenerative Disease Research. Inserm UMR1167 is also funded by the INSERM, Institut Pasteur de Lille, Lille Métropole Communauté Urbaine and French government's LABEX DISTALZ program (development of innovative strategies for a transdisciplinary approach to AD). The work was further supported by the CoSTREAM project (www.costream.eu) and funding from the European Union's Horizon 2020 research and innovation programme under grant agreement No 667375. This work is also funded by la Fondation pour la Recherche Médicale (FRM) (EQU202003010147); this work was also supported by Italian Ministry of Health (Ricerca Corrente) (R.G., IRCCS Istituto Centro San Giovanni di Dio Fatebenefratelli, Brescia); Ministero dell'Istruzione, dell'Università e della Ricerca–MIUR project "Dipartimenti di Eccellenza 2018–2022" to Department of Neuroscience "Rita Levi Montalcini", University of Torino (IR), and AIRAlzh Onlus-ANCC-COOP (SB); Partly supported by "Ministero della Salute", I.R.C.C.S. Research Program, Ricerca Corrente 2018-2020, Linea n. 2 "Meccanismi genetici, predizione e terapie innovative delle malattie complesse" and by the "5 x 1000" voluntary contribution to the Fondazione I.R.C.C.S. Ospedale "Casa Sollievo della Sofferenza"; RF-2018-12366665, Fondi per la ricerca 2019 (Sandro Sorbi), and RF-2021-12374301 (Paola Bossù). Copenhagen General Population Study (CGPS): We thank staff and participants of the CGPS for their important contributions. Karolinska Institutet AD cohort: Dr. C.G. and co-authors of the Karolinska Institutet AD cohort report grants from Swedish Research Council (VR) 2015-02926, 2018-02754, 2015-06799, Swedish Alzheimer Foundation, Region Stockholm ALF-project and research school, Karolinska Institutet StratNeuro, Swedish Demensfonden, and Swedish brain foundation, during the conduct of the study. ADGEN: This work was supported by Academy of Finland (grant numbers 307866); Sigrid Jusélius Foundation; the Strategic Neuroscience Funding of the University of Eastern Finland; EADB project in the

JPNDCO-FUND program (grant number 301220). CBAS: Supported by project nr. LX22NPO5107 (MEYS): financed by European Union – Next Generation EU; supported by the project no. LQ1605 from the National Program of Sustainability II (MEYS CR); supported by Ministry of Health of the Czech Republic, grant nr. NV19-04-00270 (All rights reserved), Grant Agency of Charles University Grants No. 693018 and 654217; the Ministry of Health, Czech Republic—conceptual development of research organization, University Hospital Motol, Prague, Czech Republic Grant No. 00064203; the Czech Ministry of Health Project AZV Grant No. 16—27611A; and Institutional Support of Excellence 2. LF UK Grant No. 699012. CNRMAJ-Rouen: This study received fundings from the Centre National de Référence Malades Alzheimer Jeunes (CNRMAJ). The Finnish Geriatric Intervention Study for the Prevention of Cognitive Impairment and Disability (FINGER) data collection was supported by grants from the Academy of Finland, La Carita Foundation, Juho Vainio Foundation, Novo Nordisk Foundation, Finnish Social Insurance Institution, Ministry of Education and Culture Research Grants, Yrjö Jahnsson Foundation, Finnish Cultural Foundation South Osthrobothnia Regional Fund, and EVO/State Research Funding grants of University Hospitals of Kuopio, Oulu and Turku, Seinäjoki Central Hospital and Oulu City Hospital, Alzheimer's Research & Prevention Foundation USA, AXA Research Fund, Knut and Alice Wallenberg Foundation Sweden, Center for Innovative Medicine (CIMED) at Karolinska Institutet Sweden, and Stiftelsen Stockholms sjukhem Sweden. FINGER cohort genotyping was funded by EADB project in the JPND CO-FUND (grant number 301220). Research at the Belgian EADB site is funded in part by the Alzheimer Research Foundation (SAO-FRA), The Research Foundation Flanders (FWO), and the University of Antwerp Research Fund. FK receives a postdoctoral fellowship (BOF 49758) from the University of Antwerp Research Fund. SNAC-K is financially supported by the Swedish Ministry of Health and Social Affairs, the participating County Councils and Municipalities, and the Swedish Research Council. BDR Bristol: We would like to thank the South West Dementia Brain Bank (SWDBB) for providing brain tissue for this study. The SWDBB is part of the Brains for Dementia Research programme, jointly funded by Alzheimer's Research UK and Alzheimer's Society and is supported by BRACE (Bristol Research into Alzheimer's and Care of the Elderly) and the Medical Research Council. BDR Nottingham and Manchester: We would like to thank the Manchester Brain Bank for providing brain tissue for this study. The Manchester Brain Bank is part of the Brains for Dementia Research programme, jointly funded by Alzheimer's Research UK and Alzheimer's Society. The University of Nottingham acknowledges support from Alzheimer's Research UK. BDR KCL: Tissue samples were supplied by the London Neurodegenerative Diseases Brain Bank at KCL, which receives funding from the Medical Research Council (MRC) and as part of the Brains for Dementia Research program, jointly funded by Alzheimer's Research UK and Alzheimer's Society. The CFAS Wales study was funded by the ESRC (RES-060-25-0060) and HEFCW as 'Maintaining function and well-being in later life: a longitudinal cohort study'. We are grateful to the NISCHR Clinical Research Centre for their assistance in tracing participants and in interviewing and in collecting blood samples, and to general practices in the study areas for their cooperation. MRC: We thank all individuals who participated in this study. Cardiff University was supported by the Alzheimer's Society (AS; grant RF014/164) and the Medical Research Council (MRC; grants G0801418/1, MR/K013041/1, MR/L023784/1) (R.S. is an AS Research Fellow). Cardiff University was also supported by the European Joint Programme for Neurodegenerative Disease (JPND; grant MR/L501517/1), Alzheimer's Research UK (ARUK; grant ARUK-PG2014-1), the Welsh Assembly Government (grant SGR544:CADR), Brain's for dementia Research and a donation from the Moondance Charitable Foundation. Cardiff University acknowledges the support of the UK Dementia Research Institute, which receives its funding from UK DRI Ltd, funded by the UK Medical Research Council, Alzheimer's Society and Alzheimer's Research UK. Cambridge University acknowledges support from the MRC. Join Dementia Research (JDR) is funded by the Department of Health and delivered by the National Institute for Health Research in partnership with Alzheimer Scotland, Alzheimer's Research UK and

Alzheimer's Society. UCL-DRC: Funding for DNA extraction from whole blood, storage and database management was provided by the MRC (UK) and the National Institute of Health Research's Biomedical Research Centre at University College London Hospitals NHS Foundation Trust, through the MRC Prion Unit at UCL/Institute of Prion Diseases. Patient recruitment for the MRC Prion Unit/UCL Department of Neurodegenerative Disease collection was supported by the UCLH/UCL Biomedical Centre and NIHR Queen Square Dementia Biomedical Research Unit. The University of Southampton acknowledges support from the Alzheimer's Society. King's College London was supported by the NIHR Biomedical Research Centre for Mental Health and the Biomedical Research Unit for Dementia at the South London and Maudsley NHS Foundation Trust and by King's College London and the MRC. ARUK and the Big Lottery Fund provided support to Nottingham University. A.Ram. : Part of the work was funded by the JPND EADB grant (German Federal Ministry of Education and Research (BMBF) grant: 01ED1619A). A. Ram. is also supported by the German Research Foundation (DFG) grants Nr: RA 1971/6-1, RA1971/7-1, and RA 1971/8-1. German Study on Ageing, Cognition and Dementia in Primary Care Patients (AgeCoDe): This study/publication is part of the German Research Network on Dementia (KND), the German Research Network on Degenerative Dementia (KNDD; German Study on Ageing, Cognition and Dementia in Primary Care Patients; AgeCoDe), and the Health Service Research Initiative (Study on Needs, health service use, costs and health-related quality of life in a large sample of oldestold primary care patients (85+; AgeQualiDe)) and was funded by the German Federal Ministry of Education and Research (grants KND: 01GI0102, 01GI0420, 01GI0422, 01GI0423, 01GI0429, 01GI0431, 01GI0433, 01GI0434; grants KNDD: 01GI0710, 01GI0711, 01GI0712, 01GI0713, 01GI0714, 01GI0715, 01GI0716; grants Health Service Research Initiative: 01GY1322A, 01GY1322B, 01GY1322C, 01GY1322D, 01GY1322E, 01GY1322F, 01GY1322G). VITA study: The support of the Ludwig Boltzmann Society and the AFI Germany have supported the VITA study. The former VITA study group should be acknowledged: W. Danielczyk, G. Gatterer, K Jellinger, S Jugwirth, KH Tragl, S Zehetmayer. Vogel Study: This work was financed by a research grant of the ''Vogelstiftung Dr. Eckernkamp''. HELIAD study: This study was supported by the grants: IIRG-09-133014 from the Alzheimer's Association, 189 10276/8/9/2011 from the ESPA-EU program Excellence Grant (ARISTEIA) and the ΔΥ2β/οικ.51657/14.4.2009 of the Ministry for Health and Social Solidarity (Greece). Biobank Department of Psychiatry, UMG: Prof. Jens Wiltfang is supported by an Ilídio Pinho professorship and iBiMED (UID/BIM/04501/2013), and FCT project PTDC/DTP_PIC/5587/2014 at the University of Aveiro, Portugal. Lausanne study: This work was supported by grants from the Swiss National Research Foundation (SNF 320030_141179). PAGES study: Harald Hampel is an employee of Eisai Inc. During part of this work he was supported by the AXA Research Fund, the "Fondation partenariale Sorbonne Université" and the "Fondation pour la Recherche sur Alzheimer", Paris, France. Mannheim, Germany Biobank: Department of geriatric Psychiatry, Central Institute for Mental Health, Mannheim, University of Heidelberg, Germany. Genotyping for the Swedish Twin Studies of Aging was supported by NIH/NIA grant R01 AG037985. Genotyping in TwinGene was supported by NIH/NIDDK U01 DK066134. WvdF is recipient of Joint Programming for Neurodegenerative Diseases (JPND) grants PERADES (ANR-13-JPRF-0001) and EADB (733051061). Gothenburg Birth Cohort (GBC) Studies: We would like to thank UCL Genomics for performing the genotyping analyses. The studies were supported by The Stena Foundation, The Swedish Research Council (2015-02830, 2013-8717), The Swedish Research Council for Health, Working Life and Wellfare (2013-1202, 2005-0762, 2008-1210, 2013-2300, 2013- 2496, 2013-0475), The Brain Foundation, Sahlgrenska University Hospital (ALF), The Alzheimer's Association (IIRG-03-6168), The Alzheimer's Association Zenith Award (ZEN-01-3151), Eivind och Elsa K:son Sylvans Stiftelse, The Swedish Alzheimer Foundation. Clinical AD, Sweden: We would like to thank UCL Genomics for performing the genotyping analyses. Barcelona Brain Biobank: Brain Donors of the Neurological Tissue Bank of the Biobanc-Hospital Clinic-IDIBAPS and their families for their generosity. Hospital Clínic de Barcelona Spanish

Ministry of Economy and Competitiveness-Instituto de Salud Carlos III and Fondo Europeo de Desarrollo Regional (FEDER), Unión Europea, “Una manera de hacer Europa” grants (PI16/0235 to Dr. R. Sánchez-Valle and PI17/00670 to Dr. A.Antonelli). AA is funded by Departament de Salut de la Generalitat de Catalunya, PERIS 2016-2020 (SLT002/16/00329). Work at JP-T laboratory was possible thanks to funding from Ciberned and generous gifts from Consuelo Cervera Yuste and Juan Manuel Moreno Cervera. Sydney Memory and Ageing Study (Sydney MAS): We gratefully acknowledge and thank the following for their contributions to Sydney MAS: participants, their supporters and the Sydney MAS Research Team (current and former staff and students). Funding was awarded from the Australian National Health and Medical Research Council (NHMRC) Program Grants (350833, 568969, 109308). This work was supported by InnoMed (Innovative Medicines in Europe), an integrated project funded by the European Union of the Sixth Framework program priority (FP6-2004- LIFESCIHEALTH-5). Oviedo: This work was partly supported by Grant from Fondo de Investigaciones Sanitarias-Fondos FEDER EuropeanUnion to V.A. PI15/00878. Project MinE: The ProjectMinE study was supported by the ALS Foundation Netherlands and the MND association (UK) (Project MinE, www.projectmine.com). The SPIN cohort: We are indebted to patients and their families for their participation in the “Sant Pau Initiative on Neurodegeneration cohort”, at the Sant Pau Hospital (Barcelona). This is a multimodal research cohort for biomarker discovery and validation that is partially funded by Generalitat de Catalunya (2017 SGR 547 to OB), as well as by the Fondo de Investigaciones Sanitario, Carlos III Health Institute (INT21/00073, PI20/01473 and PI23/01786 to J.F., PI18/00435 to D.A., PI20/01330 to A.L. and PI21/01395 to O.D.-I.) and the Centro de Investigación Biomédica en Red sobre Enfermedades Neurodegenerativas Program 1, partly jointly funded by Fondo Europeo de Desarrollo Regional, Unión Europea, Una Manera de Hacer Europa. O.D.-I. receives funding from the Alzheimer’s Association (AARF-22-924456) and the Jerome Lejeune Foundation postdoctoral fellowship. We would also like to thank the Fundació Bancària Obra Social La Caixa (DABNI project) to JF and AL; and Fundación BBVA (to AL), for their support in funding this follow-up study. Adolfo López de Munain is supported by Fundación Salud 2000 (PI2013156), CIBERNED and Diputación Foral de Gipuzkoa (Exp.114/17). P.S.J. is supported by CIBERNED and Carlos III Institute of Health, Spain (PI08/0139, PI12/02288, and PI16/01652, PI20/01011), jointly funded by Fondo Europeo de Desarrollo Regional (FEDER), Unión Europea, “Una manera de hacer Europa”. We thank Biobanco Valdecilla for their support. Amsterdam dementia Cohort (ADC): Research of the Alzheimer center Amsterdam is part of the neurodegeneration research program of Amsterdam Neuroscience. The AlzheimerCenter Amsterdam is supported by Stichting Alzheimer Nederland and Stichting VUmc fonds. The clinical database structure was developed with funding from Stichting Dioraphte. Genotyping of the Dutch case-control samples was performed in the context of EADB (European Alzheimer&Dementia biobank) funded by the JPco-fuND FP-829-029 (ZonMW project number #733051061). This research is performed by using data from the Parelsnoer Institute an initiative of the Dutch Federation of University Medical Centres (www.parelsnoer.org). 100-Plus study: We are grateful for the collaborative efforts of all participating centenarians and their family members and/or relations. We thank the Netherlands Brain Bank for supplying DNA for genotyping. This work was supported by Stichting AlzheimerNederland (WE09.2014-03), Stichting Diorapthe, Horstingstuit foundation, Memorabel (ZonMW project number #733050814, #733050512) and Stichting VUmcFonds. Additional support for EADB cohorts: WF, SL, HH are recipients of ABOARD, a public-private partnership receiving funding from ZonMW (#73305095007) and Health~Holland, Topsector Life Sciences & Health (PPP-allowance; #LSHM20106). The DELCODE study was funded by the German Center for Neurodegenerative Diseases (Deutsches Zentrum für Neurodegenerative Erkrankungen (DZNE)), reference number BN012.

**EADI.** This work has been developed and supported by the LABEX (laboratory of excellence program investment for the future) DISTALZ grant (Development of Innovative Strategies for a Transdisciplinary approach to ALZheimer's disease) including funding from MEL (Metropole européenne de Lille), ERDF (European Regional Development Fund) and Conseil Régional Nord Pas de Calais. This work was supported by INSERM, the National Foundation for Alzheimer's disease and related disorders, the Institut Pasteur de Lille and the Centre National de Recherche en Génomique Humaine, CEA, the JPND PERADES, the Laboratory of Excellence GENMED (Medical Genomics) grant no. ANR-10-LABX-0013 managed by the National Research Agency (ANR) part of the Investment for the Future program, and the FP7 AgedBrainSysBio. The Three-City Study was performed as part of collaboration between the Institut National de la Santé et de la Recherche Médicale (Inserm), the Victor Segalen Bordeaux II University and Sanofi-Synthélabo. The Fondation pour la Recherche Médicale funded the preparation and initiation of the study. The 3C Study was also funded by the Caisse Nationale Maladie des Travailleurs Salariés, Direction Générale de la Santé, MGEN, Institut de la Longévité, Agence Française de Sécurité Sanitaire des Produits de Santé, the Aquitaine and Bourgogne Regional Councils, Agence Nationale de la Recherche, ANR supported the COGINUT and COVADIS projects. Fondation de France and the joint French Ministry of Research/INSERM "Cohortes et collections de données biologiques" programme. Lille Génopôle received an unconditional grant from Eisai. The Three-city biological bank was developed and maintained by the laboratory for genomic analysis LAG-BRC - Institut Pasteur de Lille.

**Bonn.** This group would like to thank Dr. Heike Koelsch for her scientific support. The Bonn group was funded by the German Federal Ministry of Education and Research (BMBF): Competence Network Dementia (CND) grant number 01GI0102, 01GI0711, 01GI042

**FinnGen.** We want to acknowledge the participants and investigators of the FinnGen study. The FinnGen project is funded by two grants from Business Finland (HUS 4685/31/2016 and UH 4386/31/2016) and the following industry partners: AbbVie Inc., AstraZeneca UK Ltd, Biogen MA Inc., Bristol Myers Squibb (and Celgene Corporation & Celgene International II Sàrl), Genentech Inc., Merck Sharp & Dohme LCC, Pfizer Inc., GlaxoSmithKline Intellectual Property Development Ltd., Sanofi US Services Inc., Maze Therapeutics Inc., Janssen Biotech Inc, Novartis AG, and Boehringer Ingelheim International GmbH. Following biobanks are acknowledged for delivering biobank samples to FinnGen: Auria Biobank (www.auria.fi/biopankki), THL Biobank (www.thl.fi/biobank), Helsinki Biobank (www.helsinginbiopankki.fi), Biobank Borealis of Northern Finland (https://www.ppshp.fi/Tutkimus-ja-opetus/Biopankki/Pages/Biobank-Borealis-briefly-in-English.aspx), Finnish Clinical Biobank Tampere (www.tays.fi/en-US/Research_and_development/Finnish_Clinical_Biobank_Tampere), Biobank of Eastern Finland (www.ita-suomenbiopankki.fi/en), Central Finland Biobank (www.ksshp.fi/fi-FI/Potilaalle/Biopankki), Finnish Red Cross Blood Service Biobank (www.veripalvelu.fi/verenluovutus/biopankkitoiminta), Terveystalo Biobank (www.terveystalo.com/fi/Yritystietoa/Terveystalo-Biopankki/Biopankki/) and Arctic Biobank (https://www.oulu.fi/en/university/faculties-and-units/faculty-medicine/northern-finland-birth-cohorts-and-arctic-biobank). All Finnish Biobanks are members of BBMRI.fi infrastructure (https://www.bbmri-eric.eu/national-nodes/finland/). Finnish Biobank Cooperative -FINBB (https://finbb.fi/) is the coordinator of BBMRI-ERIC operations in Finland. The Finnish biobank data can be accessed through the Fingenious® services (https://site.fingenious.fi/en/) managed by FINBB.

**GERAD.** Data used in the preparation of this article were obtained from the Genetic and Environmental Risk for Alzheimer's disease (GERAD1) Consortium. Cardiff University was supported by the Wellcome Trust, Medical Research Council (MRC), Alzheimer's Research UK (ARUK) and the Welsh Assembly Government. ARUK supported sample collections at the Kings College London, the South West Dementia Bank, Universities of Cambridge, Nottingham, Manchester and Belfast. The Belfast group acknowledges support from the Alzheimer's Society, Ulster Garden Villages, N.Ireland R&D Office and the Royal College of Physicians/Dunhill Medical Trust. The MRC and Mercer's Institute for Research on Ageing supported the Trinity College group. The South West Dementia Brain Bank acknowledges support from Bristol Research into Alzheimer's and Care of the Elderly. The Charles Wolfson Charitable Trust supported the OPTIMA group. Washington University was funded by NIH grants, Barnes Jewish Foundation and the Charles and Joanne Knight Alzheimer's Research Initiative. Patient recruitment for the MRC Prion Unit/UCL Department of Neurodegenerative Disease collection was supported by the UCLH/UCL Biomedical Centre. LASER-AD was funded by Lundbeck SA. The Bonn group was supported by the German Federal Ministry of Education and Research (BMBF), Competence Network Dementia and Competence Network Degenerative Dementia, and by the Alfried Krupp von Bohlen und Halbach-Stiftung. The GERAD1 Consortium also used samples ascertained by the NIMH AD Genetics Initiative.

**GR@ACE.** We would like to thank patients and controls who participated in this project. The Genome Research @ Ace Alzheimer Center Barcelona project (GR@ACE) is supported by Grifols SA, Fundación bancaria 'La Caixa', Ace Alzheimer Center Barcelona and CIBERNED. Ace Alzheimer Center Barcelona is one of the participating centers of the Dementia Genetics Spanish Consortium (DEGESCO). AR and MB receive support from the European Union / EFPIA Innovative Medicines Initiative joint undertaking ADAPTED and MOPEAD projects (grant numbers 115975 and 115985, respectively). MB and AR are also supported by national grants PI13/02434, PI16/01861, PI17/01474, PI19/01240, PI19/01301 and PI22/01403. Acción Estratégica en Salud is integrated into the Spanish National R+D+I Plan and funded by ISCIII –Subdirección General de Evaluación and the Fondo Europeo de Desarrollo Regional (FEDER–'Una manera de hacer Europa'). AR is also funded by JPco-fuND-2 "Multinational research projects on Personalized Medicine for Neurodegenerative Diseases" ADPriOMICs project (AC23_2/00038), PREADAPT project (ISCIII grant: AC19/00097), EURONANOMED III Joint Transnational call for proposals (2017) for European Innovative Research & Technological Development Projects in Nanomedicine (ISCIII grant: AC17/00100), the ISCIII national grant PMP22/00022, funded by the European Union (NextGenerationEU), The support of CIBERNED (ISCIII) under the grants CB06/05/2004 and CB18/05/00010. AR also received support from HARPONE project, Agency for Innovation and Entrepreneurship (VLAIO, Belgium) grant N° PR067/21, Janssen and DESCARTES project funded by German Research Foundation (DFG, Germany). I.dR. is supported by a national grant from the Instituto de Salud Carlos III FI20/00215. Ace Alzheimer Center Barcelona research receives support from Roche, Janssen, Life Molecular Imaging, Araclon Biotech, Alkahest, Laboratorio de Análisis Echevarne, and IrsiCaixa. Some control samples and data from patients included in this study were provided in part by the National DNA Bank Carlos III (www.bancoadn.org, University of Salamanca, Spain) and Hospital Universitario Virgen de Valme (Sevilla, Spain); they were processed following standard operating procedures with the appropriate approval of the Ethical and Scientific Committee.

**CHARGE.** This project was conducted within the neurology working group of the Cohorts for Heart and Aging Research in Genomic Epidemiology (*CHARGE*) Consortium. The CHARGE cohorts are supported in part by the National Heart, Lung, and Blood Institute (NHLBI) infrastructure grants R01HL105756 (Psaty) and

RC2HL102419 (Boerwinkle), and the neurology working group is supported by the National Institute on Aging (NIA) R01 grant AG033193 (Seshadri). ARIC: The Atherosclerosis Risk in Communities Study is carried out as a collaborative study supported by National Heart, Lung, and Blood Institute contracts (75N92022D00001, 75N92022D00002, 75N92022D00003. 75N922D00004, 75N92022D00005). The ARIC Neurocognitive Study is supported by U01HL096812, U01HL096814, U01HL096899, U01HL096902, and U01HL096917 from the NIH (NHLBI, NINDS, NIA and NIDCD). Funding was also supported by R01HL087641, and R01HL086694; National Human Genome Research Institute contract U01HG004402; and National Institutes of Health contract HHSN268200625226C. Infrastructure was partly supported by Grant Number UL1RR025005, a component of the National Institutes of Health and NIH Roadmap for Medical Research. Funding support for "Building on GWAS for NHLBI-diseases: the U.S. CHARGE consortium" was provided by the NIH through the American Recovery and Reinvestment Act of 2009 (ARRA) (5RC2HL102419). This project was funded from R01-NS087541 to Myriam Fornage and Eric Boerwinkle. The authors thank the staff and participants of the ARIC study for their important contributions. CHS: This CHS research was supported by NHLBI contracts HHSN268201200036C, HHSN268200800007C, HHSN268201800001C, N01HC55222, N01HC85079, N01HC85080, N01HC85081, N01HC85082, N01HC85083, N01HC85086, 75N92021D00006; and NHLBI grants U01HL080295, R01HL087652, R01HL105756, R01HL103612, R01HL120393, and U01HL130114 with additional contribution from the National Institute of Neurological Disorders and Stroke (NINDS). Additional support was provided through R01AG023629, R01AG20098, R01AG15928, and R01AG033193 from the National Institute on Aging (NIA). A full list of principal CHS investigators and institutions can be found at CHS-NHLBI.org. The provision of genotyping data was supported in part by the National Center for Advancing Translational Sciences, CTSI grant UL1TR001881, and the National Institute of Diabetes and Digestive and Kidney Disease Diabetes Research Center (DRC) grant DK063491 to the Southern California Diabetes Endocrinology Research Center. FHS: FHS is supported by the National Heart, Lung and Blood Institute's Framingham Heart Study (Contracts No. N01-HC-25195, No. HHSN268201500001I and No. 75N92019D00031), and its contract with Affymetrix, Inc. for genotyping services (Contract No. N02-HL-6-4278). A portion of this research utilized the Linux Cluster for Genetic Analysis (LinGA-II) funded by the Robert Dawson Evans Endowment of the Department of Medicine at Boston University School of Medicine and Boston Medical Center. This study was also supported by grants from the National Institute of Aging (R01s AG033040, AG033193, AG054076, AG049607, AG059421, U01 AG058589, AG061872 and U01-AG049505) and the National Institute of Neurological Disorders and Stroke (R01-NS017950, UH2 NS100605). Dr. DeCarli is supported by the Alzheimer's Disease Center (P30 AG 010129). We thank the study participants, as well as the study team (especially the investigators and staff of the neurology team) for their contributions and dedication to the study. The authors are pleased to acknowledge that the computational work reported on in this paper was performed on the Shared Computing Cluster that is administered by Boston University Research Computing Services. URL: www.bu.edu/tech/support/research/.

**Rotterdam study.** This study was funded by the Netherlands Organisation for Health Research and Development (ZonMW) as part of the Joint Programming for Neurological Disease (JPND) as part of the PERADES Program (Defining Genetic Polygenic, and Environmental Risk for Alzheimer's disease using multiple powerful cohorts, focused Epigenetics and Stem cell metabolomics), Project number 733051021. This work was funded also by the European Union Innovative Medicine Initiative (IMI) programme under grant agreement No. 115975 as part of the Alzheimer's Disease Apolipoprotein Pathology for Treatment Elucidation and Development (ADAPTED, https://www.imi-adapted.eu) and the European Union's Horizon 2020 research and innovation programme as part of the Common mechanisms and pathways in Stroke and

Alzheimer's disease CoSTREAM project (www.costream.eu, grant agreement No. 667375). The current study is supported by the Deltaplan Dementie and Memorabel supported by ZonMW (Project number 733050814) and Alzheimer Nederland. The Rotterdam Study is funded by Erasmus Medical Center and Erasmus University, Rotterdam, Netherlands Organization for the Health Research and Development (ZonMw), the Research Institute for Diseases in the Elderly (RIDE), the Ministry of Education, Culture and Science, the Ministry for Health, Welfare and Sports, the European Commission (DG XII), and the Municipality of Rotterdam. The authors are grateful to the study participants, the staff from the Rotterdam Study and the participating general practitioners and pharmacists. The generation and management of GWAS genotype data for the Rotterdam Study (RS-I, RS-II, RS-III) was executed by the Human Genotyping Facility of the Genetic Laboratory of the Department of Internal Medicine, Erasmus MC, Rotterdam, The Netherlands. The GWAS datasets are supported by the Netherlands Organization of Scientific Research NWO Investments (Project number 175.010.2005.011, 911-03-012), the Genetic Laboratory of the Department of Internal Medicine, Erasmus MC, the Research Institute for Diseases in the Elderly (014-93-015; RIDE2), the Netherlands Genomics Initiative (NGI)/Netherlands Organization for Scientific Research (NWO) Netherlands Consortium for Healthy Aging (NCHA), project number 050-060-810. We thank Pascal Arp, Mila Jhamai, Marijn Verkerk, Lizbeth Herrera and Marjolein Peters, MSc, and Carolina Medina-Gomez, MSc, for their help in creating the GWAS database, and Karol Estrada, PhD, Yurii Aulchenko, PhD, and Carolina Medina- Gomez, MSc, for the creation and analysis of imputed data.

**DemGene.** The project has received funding from The Research Council of Norway (RCN) Grant Nos. 213837, 223273, 225989, 248778, and 251134 and EU JPND Program RCN Grant Nos. 237250, 311993, the South-East Norway Health Authority Grant No. 2013-123, the Norwegian Health Association, and KG Jebsen Foundation. The RCN FRIPRO Mobility grant scheme (FRICON) is co-funded by the European Union's Seventh Framework Programme for research, technological development and demonstration under Marie Curie grant agreement No 608695. European Community's grant PIAPP-GA-2011-286213 PsychDPC.

**PGC-ALZ.** This work was supported by BRAINSCAPES: A Roadmap from Neurogenetics to Neurobiology (grant no. 024.004.012) and a European Research Council advanced grant (grant no. ERC-2018-AdG GWAS2FUNC 834057). The Trøndelag Health Study (The HUNT Study) is a collaboration between HUNT Research Centre (Faculty of Medicine and Health Sciences, NTNU, Norwegian University of Science and Technology), Trøndelag County Council, Central Norway Regional Health Authority, and the Norwegian Institute of Public Health. The genotyping in HUNT was financed by the National Institutes of Health; University of Michigan; the Research Council of Norway; the Liaison Committee for Education, Research and Innovation in Central Norway; and the Joint Research Committee between St Olavs hospital and the Faculty of Medicine and Health Sciences, NTNU. The genetic investigations of the HUNT Study are a collaboration between researchers from the HUNT Center for Molecular and Clinical Epidemiology (formerly known as the K.G. Jebsen Center for Genetic Epidemiology as of August 1st 2023), NTNU, and the University of Michigan Medical School and the University of Michigan School of Public Health. We thank HUNT participants for donating their time, samples, and information to help others; clinicians and other employees at Nord-Trøndelag Hospital Trust for their support and for contributing to data collection.

**ADGC.** The National Institutes of Health, National Institute on Aging (NIH-NIA) supported this work through the following grants: ADGC, U01 AG032984, RC2 AG036528; Samples from the National Centralized Repository for Alzheimer's Disease and Related Dementias (NCRAD), which receives government support under a cooperative agreement grant (U24 AG021886) awarded by the National Institute on Aging (NIA),

were used in this study. We thank contributors who collected samples used in this study, as well as patients and their families, whose help and participation made this work possible; Data for this study were prepared, archived, and distributed by the National Institute on Aging Alzheimer’s Disease Data Storage Site (NIAGADS) at the University of Pennsylvania (U24 AG041689); GCAD, U54 AG052427; Additional University of Pennsylvania grants supporting this work included R01 AG054060 and RF1 AG061351 (PI/MPI Adam Naj).
**ACT - Adult Changes in Thought -** Kaiser Permanente Washington, U19AG06656
**ADC - Alzheimer's Disease Centers/NACC National Alzheimer’s Coordinating Center -** The NACC database is funded by NIA/NIH Grant U24 AG072122. NACC data are contributed by the NIA-funded ADRCs: P30 AG062429 (PI James Brewer, MD, PhD), P30 AG066468 (PI Oscar Lopez, MD), P30 AG062421 (PI Bradley Hyman, MD, PhD), P30 AG066509 (PI Thomas Grabowski, MD), P30 AG066514 (PI Mary Sano, PhD), P30 AG066530 (PI Helena Chui, MD), P30 AG066507 (PI Marilyn Albert, PhD), P30 AG066444 (PI David Holtzman, MD), P30 AG066518 (PI Lisa Silbert, MD, MCR), P30 AG066512 (PI Thomas Wisniewski, MD), P30 AG066462 (PI Scott Small, MD), P30 AG072979 (PI David Wolk, MD), P30 AG072972 (PI Charles DeCarli, MD), P30 AG072976 (PI Andrew Saykin, PsyD), P30 AG072975 (PI Julie A. Schneider, MD, MS), P30 AG072978 (PI Ann McKee, MD), P30 AG072977 (PI Robert Vassar, PhD), P30 AG066519 (PI Frank LaFerla, PhD), P30 AG062677 (PI Ronald Petersen, MD, PhD), P30 AG079280 (PI Jessica Langbaum, PhD), P30 AG062422 (PI Gil Rabinovici, MD), P30 AG066511 (PI Allan Levey, MD, PhD), P30 AG072946 (PI Linda Van Eldik, PhD), P30 AG062715 (PI Sanjay Asthana, MD, FRCP), P30 AG072973 (PI Russell Swerdlow, MD), P30 AG066506 (PI Glenn Smith, PhD, ABPP), P30 AG066508 (PI Stephen Strittmatter, MD, PhD), P30 AG066515 (PI Victor Henderson, MD, MS), P30 AG072947 (PI Suzanne Craft, PhD), P30 AG072931 (PI Henry Paulson, MD, PhD), P30 AG066546 (PI Sudha Seshadri, MD), P30 AG086401 (PI Erik Roberson, MD, PhD), P30 AG086404 (PI Gary Rosenberg, MD), P20 AG068082 (PI Angela Jefferson, PhD), P30 AG072958 (PI Heather Whitson, MD), P30 AG072959 (PI James Leverenz, MD).
**ADNI - Alzheimer's Disease Neuroimaging Initiative -** Data collection and sharing for the Alzheimer's Disease Neuroimaging Initiative (ADNI) is funded by the National Institute on Aging (National Institutes of Health Grant U19 AG024904). The grantee organization is the Northern California Institute for Research and Education. In the past, ADNI has also received funding from the National Institute of Biomedical Imaging and Bioengineering, the Canadian Institutes of Health Research, and private sector contributions through the Foundation for the National Institutes of Health (FNIH) including generous contributions from the following: AbbVie, Alzheimer’s Association; Alzheimer’s Drug Discovery Foundation; Araclon Biotech; BioClinica, Inc.; Biogen; Bristol-Myers Squibb Company; CereSpir, Inc.; Cogstate; Eisai Inc.; Elan Pharmaceuticals, Inc.; Eli Lilly and Company; EuroImmun; F. Hoffmann-La Roche Ltd and its affiliated company Genentech, Inc.; Fujirebio; GE Healthcare; IXICO Ltd.; Janssen Alzheimer Immunotherapy Research & Development, LLC.; Johnson & Johnson Pharmaceutical Research &Development LLC.; Lumosity; Lundbeck; Merck & Co., Inc.; Meso Scale Diagnostics, LLC.; NeuroRx Research; Neurotrack Technologies; Novartis Pharmaceuticals Corporation; Pfizer Inc.; Piramal Imaging; Servier; Takeda Pharmaceutical Company; and Transition Therapeutics;
**BIOCARD - Biomarkers of Cognitive Decline Among Normal Individuals -** Johns Hopkins University, U19AG033655;
**CHAP - Chicago Health and Aging Project -** Rush University, R01 AG11101;
**EAS - Einstein Aging Study -** Einstein College of Medicine, P01 AG03949;
**EFIGA - Estudio Familiar de Influencia Genetica en Alzheimer -** Columbia University, RF1AG015473;
**GenerAAtions - Genetic & Environmental Risk Factors for AD Among African Americans -** Johns Hopkins BIOCARD R01AG020688;

The **GSK** dataset is from the Multi-Site Collaborative Study for Genotype-Phenotype Associations in Alzheimer's disease (Genetic Alzheimer's Disease Associations – **GenADA),** funded by GlaxoSmithKline.
**Indianapolis - Indianapolis African Americans/Ibadan Study of Aging -** Indianapolis-Ibadan Dementia Project, R01AG009956;
We are very grateful to the members of **Japanese Genetic Study Consortium of Alzheimer's Disease (JGSCAD)** for the collection of blood samples. The members were listed in previous JGSCAD publications [Kuwano R, 2006; Miyashita A,2007].
**Japanese Alzheimer's Disease Neuroimaging Initiative (J-ADNI)** was supported by the following funding sources: the Translational Research Promotion Project from the New Energy and Industrial Technology Development Organization of Japan; Research on Dementia, Health Labor Sciences Research Grant; the Life Science Database Integration Project of Japan Science and Technology Agency; the Research Association of Biotechnology (Astellas Pharma Inc., Bristol-Myers Squibb, Daiichi-Sankyo, Eisai, Eli Lilly and Company, Merck-Banyu, Mitsubishi Tanabe Pharma, Pfizer Inc., Shionogi & Co., Ltd., Sumitomo Dainippon, and Takeda Pharmaceutical Company), Japan; and a grant from an anonymous foundation. The investigators within J-ADNI contributed to the design and implementation of J-ADNI and/or provided data but did not participate in the analysis or the writing of this report. A complete listing of J-ADNI investigators can be found at https://humandbs.biosciencedbc.jp/en/ hum0043-j-adni-authors
**MAYO - Mayo Clinic –** Mayo Clinic, R01 AG032990, U01 AG046139, R01 NS080820, RF1 AG051504, P50 AG016574;
**MIRAGE - Multi-Institutional Research in Alzheimer's Genetic Epidemiology -** Boston University, R01-AG048927, U01-AG082655;
**NIA-LOAD/NCRAD - NIA-LOAD/National Centralized Repository for Alzheimer's Disease/National Centralized Repository for Alzheimer's Disease (NCRAD) -** Columbia University, U24AG056270, P30AG072976;
**OHSU - Oregon Health and Science University -** Oregon Health and Science University, P30 AG008017, R01 AG026916;
**PRADI - Puerto Rican Alzheimer's Disease Initiative –** University of Miami, RF1AG054074;
**REAAADI - Research in African-American Alzheimer's Disease Initiative -** Wakeforest University & University of Miami, AG052410;
**RMAYO - Rochester Mayo Clinic -** Mayo Clinic, P50 AG016574; P30 AG062677;
**ROS/MAP - Religious Orders Study/Memory and Aging Project –** Rush University, P30AG10161, P30AG72975, R01AG15819, R01AG17917, U01AG46152, and U01AG61356
**TARCC - Texas Alzheimer's Research and Care Consortium -** The Texas Alzheimer's Research and Care Consortium (TARCC) is funded by the state of Texas through the Texas Council on Alzheimer's Disease and Related Disorders;
**TGEN - Translational Genomics Research Institute -** TGEN - AG041232, Genotyping of the TGEN2 cohort was supported by Kronos Science. The TGen series was also funded by NIA grant AG041232 to AJM and MJH, The Banner Alzheimer's Foundation;
**UM/CWRU/MSSM: Subset A-C and MTV/MTC-** University of Miami/Case Western Reserve University/Mount Sinai School of Medicine - University of Miami, Case Western Reserve University, Mount Sinai School of Medicine, Texas Alzheimer's Research and Care Consortium, R01 AG027944, R01 AG028786, R01 AG019085;
**UPITT -** University of Pittsburgh, P30 AG066468, R01 AG030653, AG064877;
**WASHU - Washington University -** Washington University, P50 AG05681, P01 AG03991, P01 AG026276;
**WHICAP - Washington Heights/Inwood Columbia Aging Project –** Columbia University, RF1 AG054023;

Additional grants supporting this work included: National Institute on Aging grants R01-AG048927, U01-AG082665, U01-AG062602, U01-AG081230, P30-AG072978 and U01-AG032984 (Boston University); U01HG008657 (University of Washington); U24 NS072026, P30 AG019610 and P30 AG072980 (Thomas G. Beach); AG069008 (Amanda J. Myers); RF1AG082339, R01AG082730 and F30NS124136 (David W. Fardo). A.M.G acknowledges funding from the Sanford J Grossman Charitable Trust.
