## Supplementary Figures for "Consensus meta-analysis of genome-wide association studies for Alzheimer’s disease and related dementia"

**Supplementary Figure 1:** QQplots of the main meta-analysis results. The *APOE* locus was excluded (43 to 47 Mb on chromosome 19). a) All variants. Genomic inflation factor lambda=1.084. b) Frequent variants (minor allele frequency > 0.01). Genomic inflation factor lambda=1.202, LDSC intercept=1.088, partitioned LDSC intercept=1.055

a)

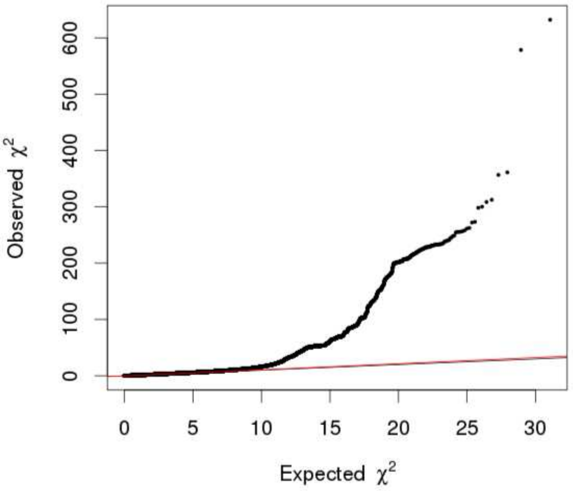


b)

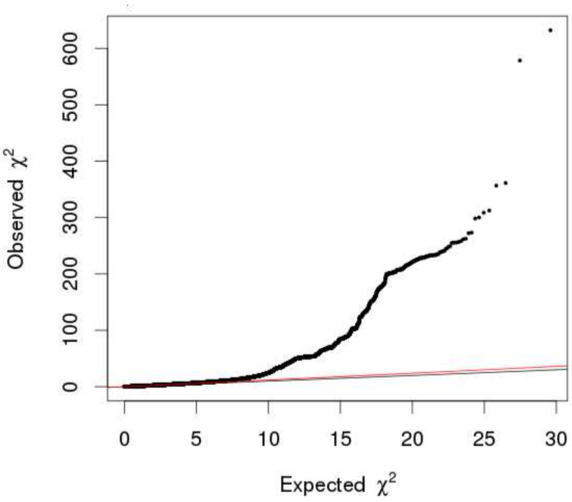

**Supplementary Figure 2:** Manhattan plots of the a) main b) no-proxy and c) no-biobank meta-analyses results. The y-axis was cut at 100. p: two-sided raw P-values derived from a fixed-effect meta-analysis.

a)

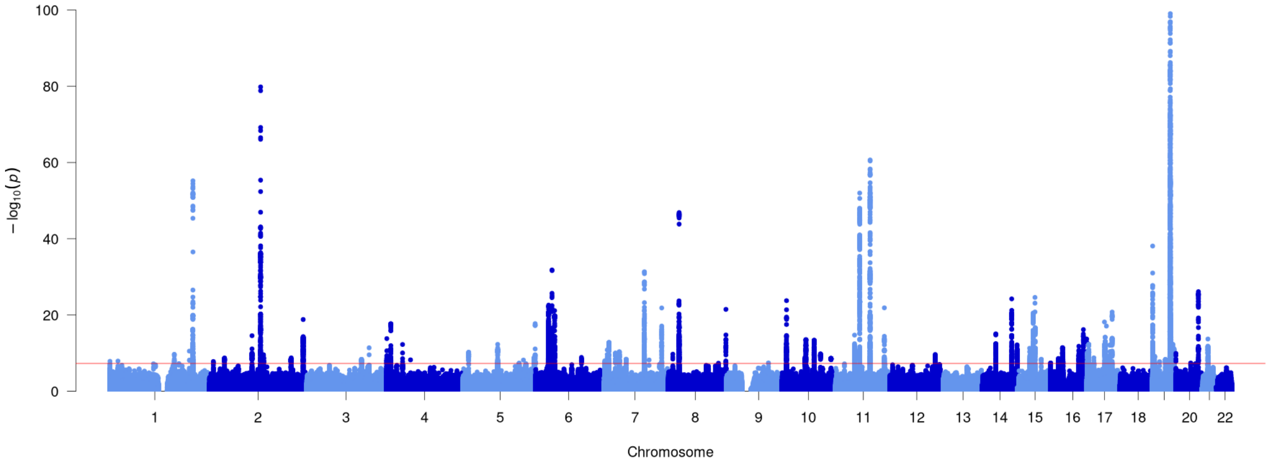


b)

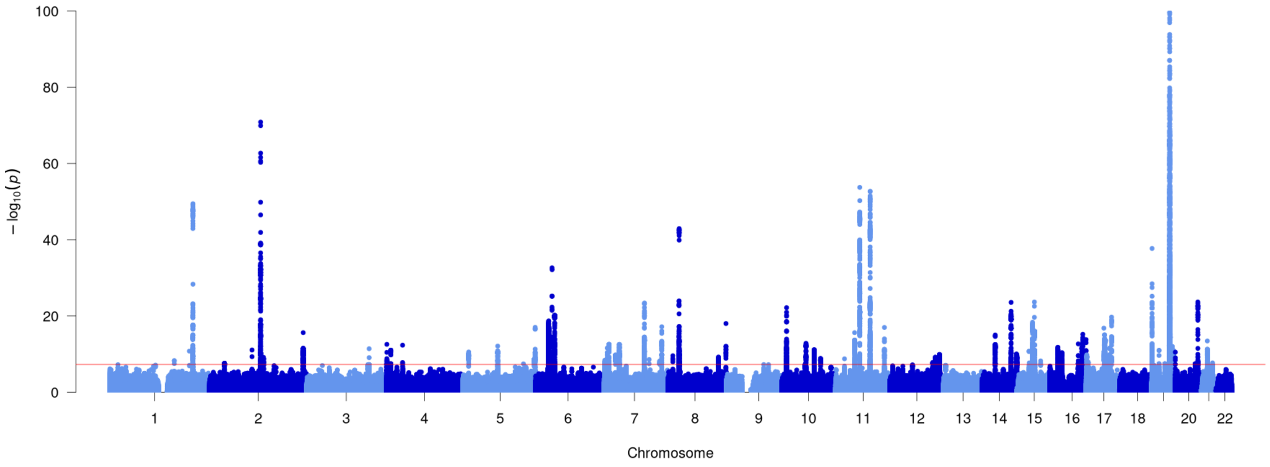


c)

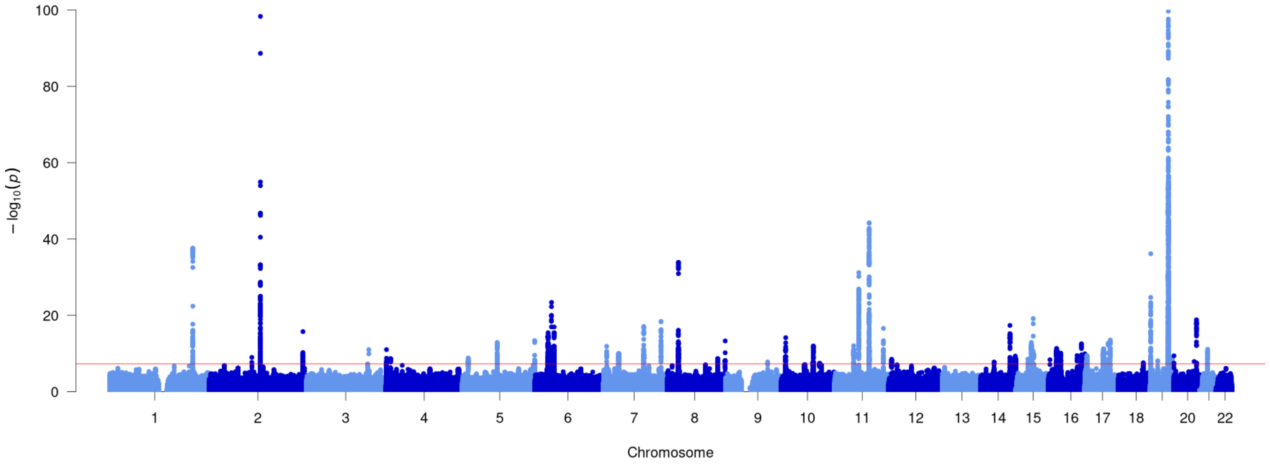

**Supplementary Figure 3:** LocusZoom and forest plots of the main analysis results for (a) *AGRN*, (b) *EIF4G3* (lead signal) and (c) *EIF4G3* (secondary signal) loci. The forest plots also show the results of the no-proxy and no-biobank meta-analyses (where the UKBB diagnosed rather than the UKBB proxy results were used). In the forest plots, data are presented as odds-ratio with 95% confidence interval. P values are two-sided raw P values derived from a fixed-effect meta-analysis. OR: odds ratio, CI: confidence interval, EA: effect allele, EAF: effect allele frequency range across all studies, HetP: heterogeneity P value, HetISq: heterogeneity statistic.

a)

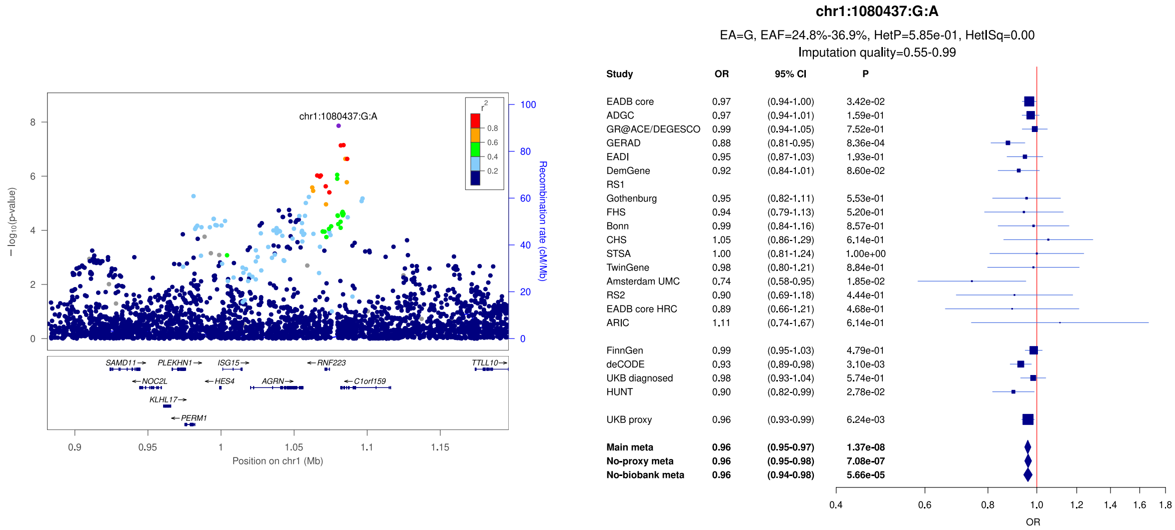


b)

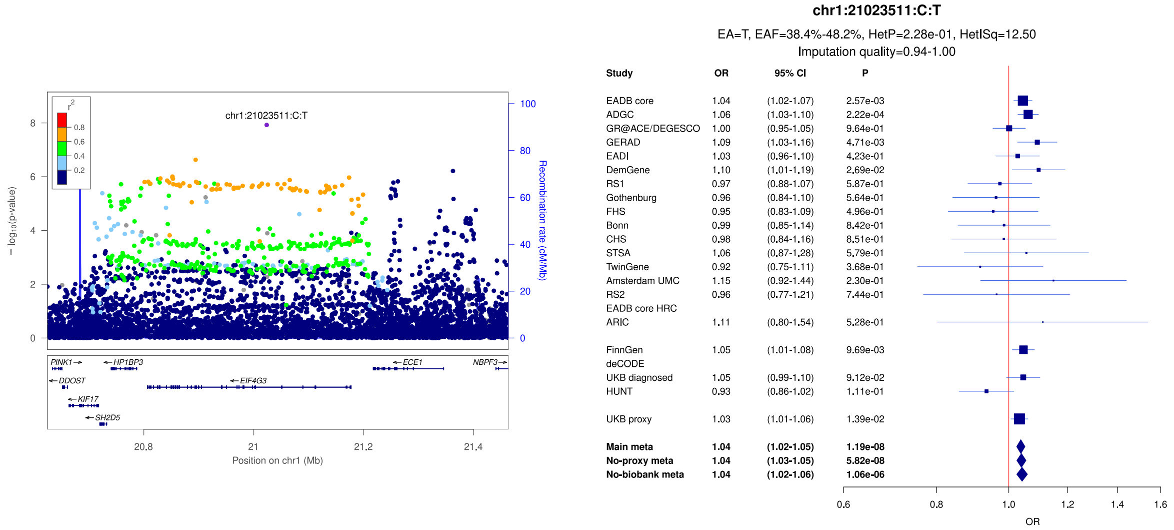


c)

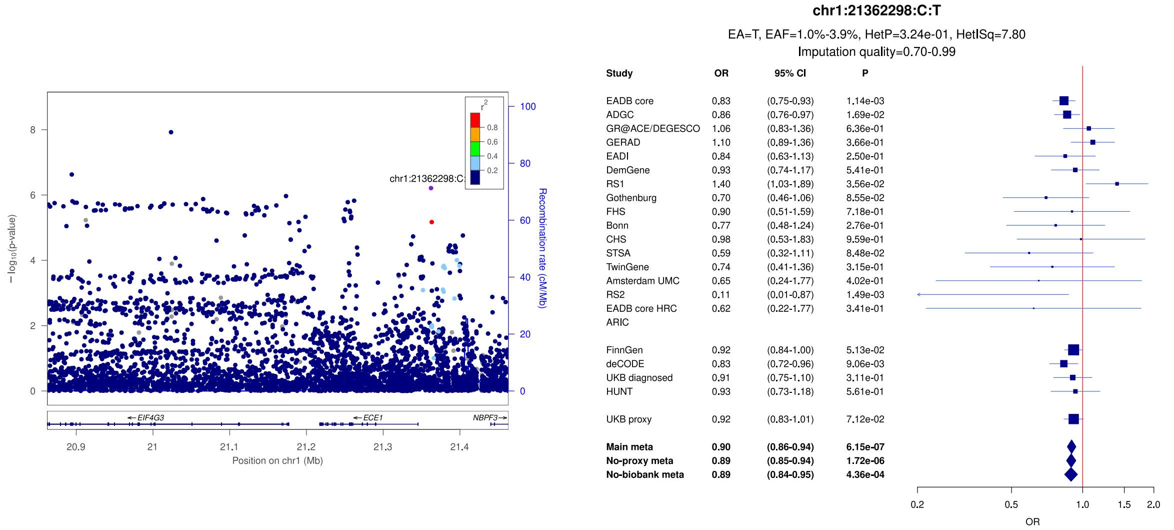

**Supplementary Figure 4:** LocusZoom and forest plots of the main analysis results for (a) *SORT1*, (b) *ADAMTS4* and (c) *PTPRC* loci. The forest plots also show the results of the no-proxy and no-biobank meta-analyses (where the UKBB diagnosed rather than the UKBB proxy results were used). In the forest plots, data are presented as odds-ratio with 95% confidence interval. P values are two-sided raw P values derived from a fixed-effect meta-analysis. OR: odds ratio, CI: confidence interval, EA: effect allele, EAF: effect allele frequency range across all studies, HetP: heterogeneity P value, HetISq: heterogeneity statistic.

a)

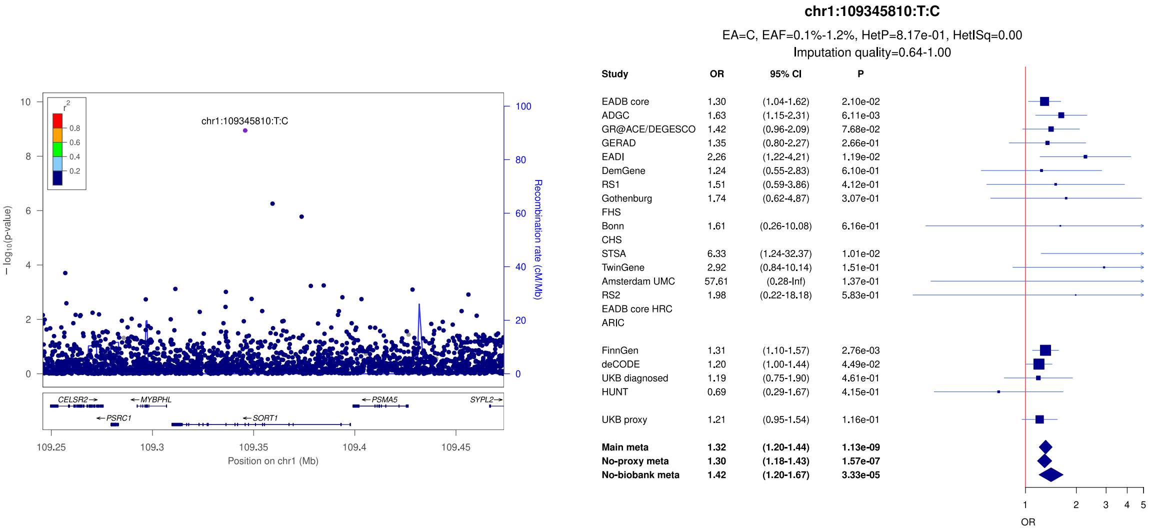


b)

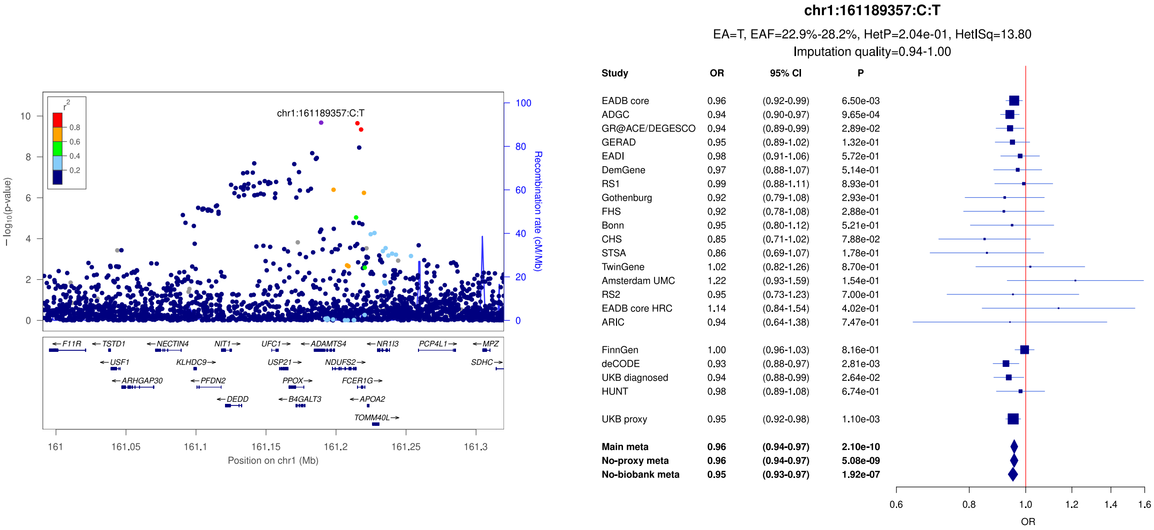


c)

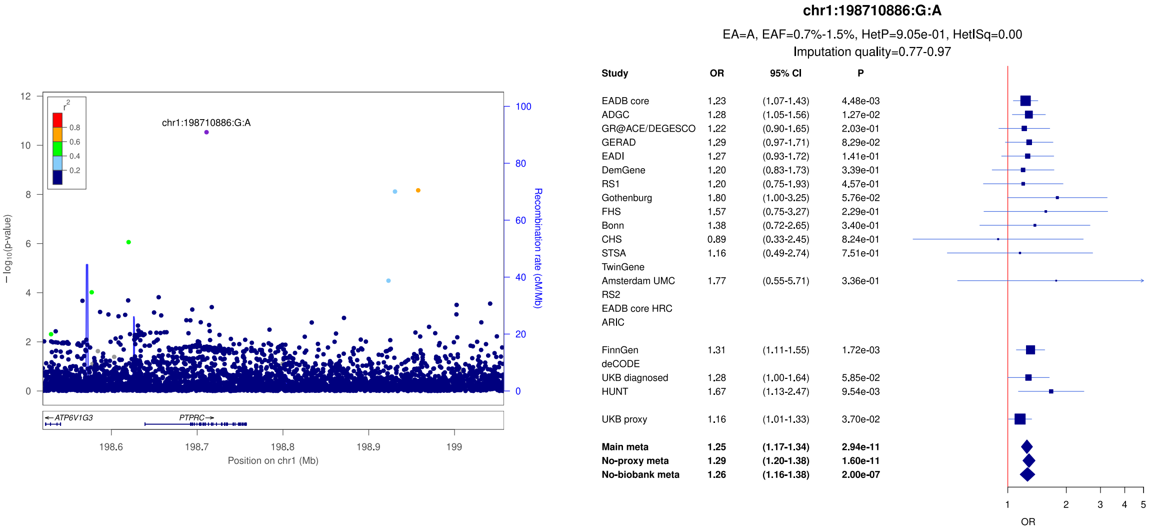

**Supplementary Figure 5:** LocusZoom and forest plots of the main analysis results for (a) *CR1*, (b) *ADAM17* and (c) *PRKD3* loci. The forest plots also show the results of the no-proxy and no-biobank meta-analyses (where the UKBB diagnosed rather than the UKBB proxy results were used). In the forest plots, data are presented as odds-ratio with 95% confidence interval. P values are two-sided raw P values derived from a fixed-effect meta-analysis. OR: odds ratio, CI: confidence interval, EA: effect allele, EAF: effect allele frequency range across all studies, HetP: heterogeneity P value, HetISq: heterogeneity statistic.

a)

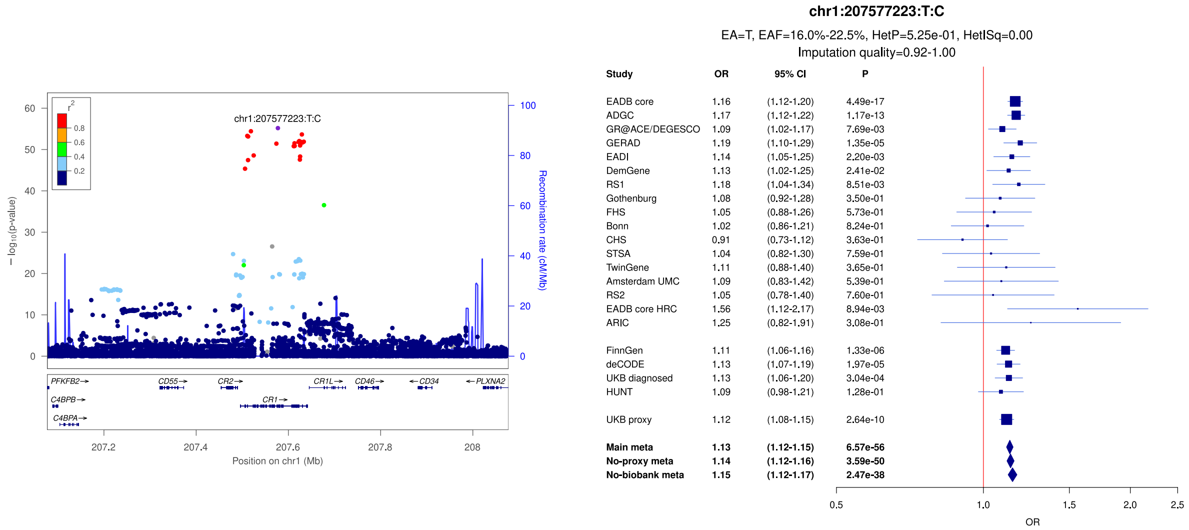


b)

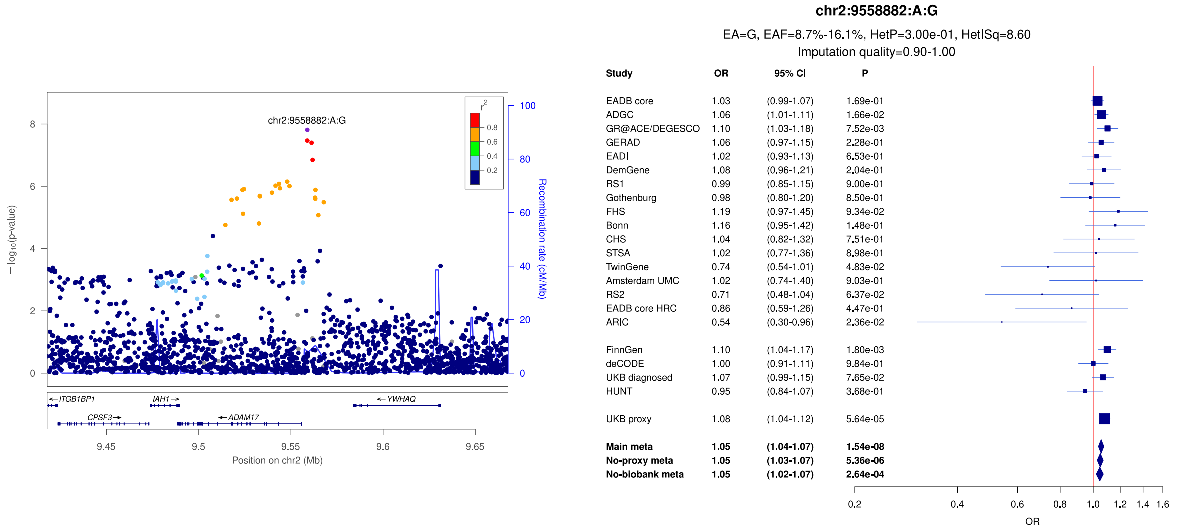


c)

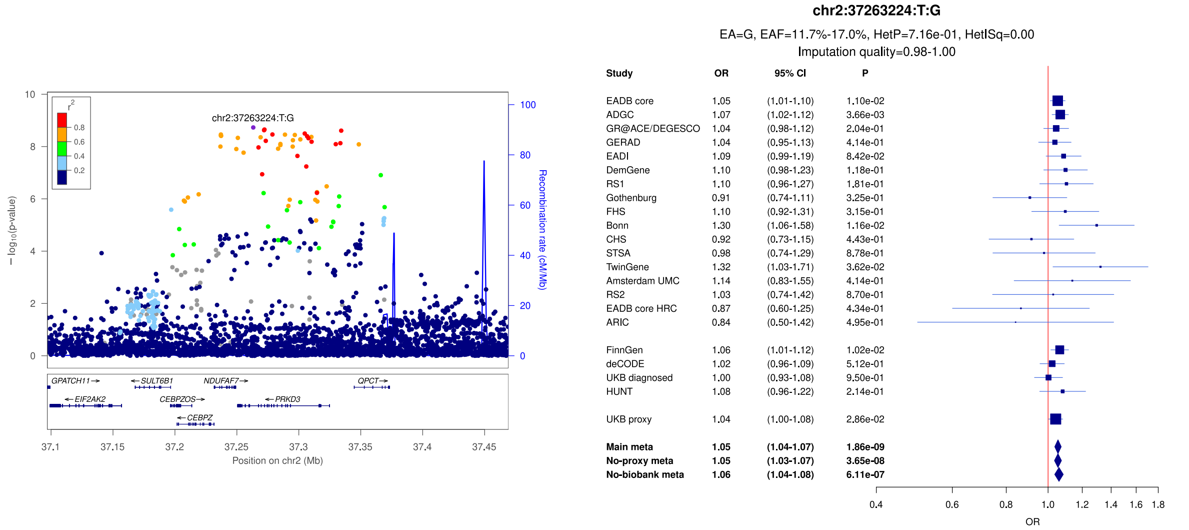

**Supplementary Figure 6:** LocusZoom and forest plots of the main analysis results for *NCK2* lead (b) and secondary (a and c) signals. The forest plots also show the results of the no-proxy and no-biobank meta-analyses (where the UKBB diagnosed rather than the UKBB proxy results were used). In the forest plots, data are presented as odds-ratio with 95% confidence interval. P values are two-sided raw P values derived from a fixed-effect meta-analysis. OR: odds ratio, CI: confidence interval, EA: effect allele, EAF: effect allele frequency range across all studies, HetP: heterogeneity P value, HetISq: heterogeneity statistic.

a)

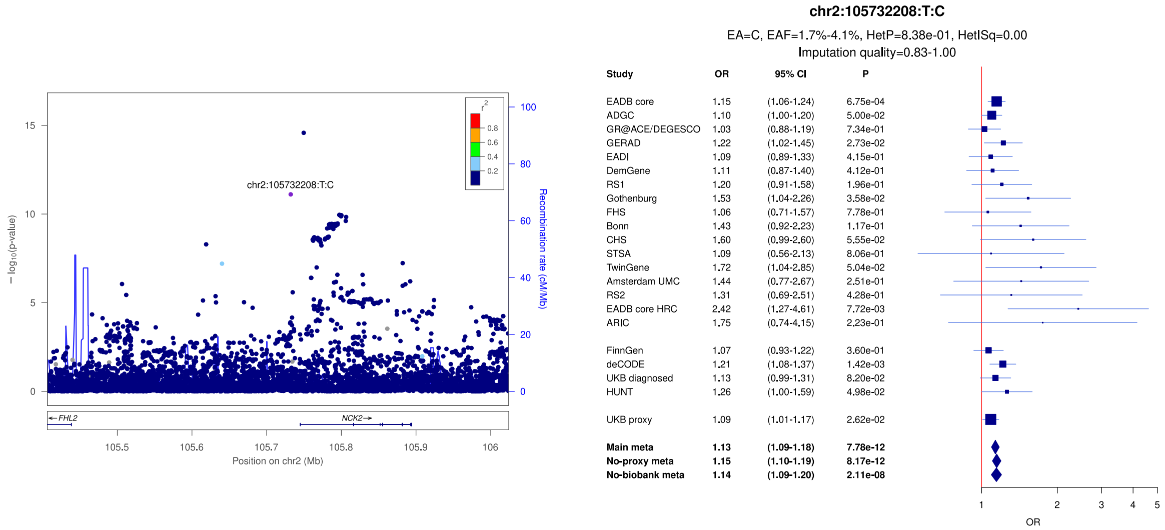


b)

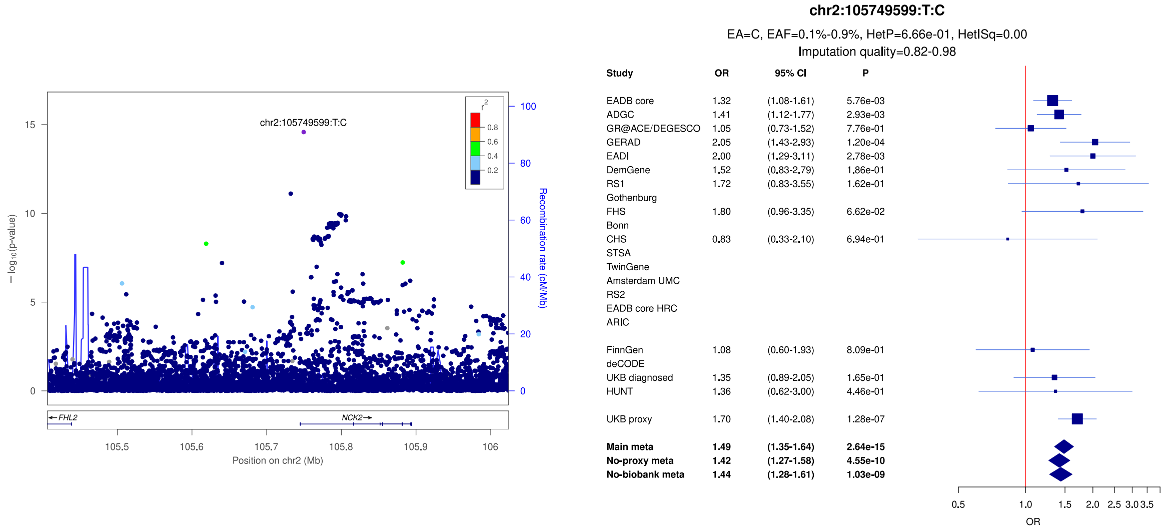


c)

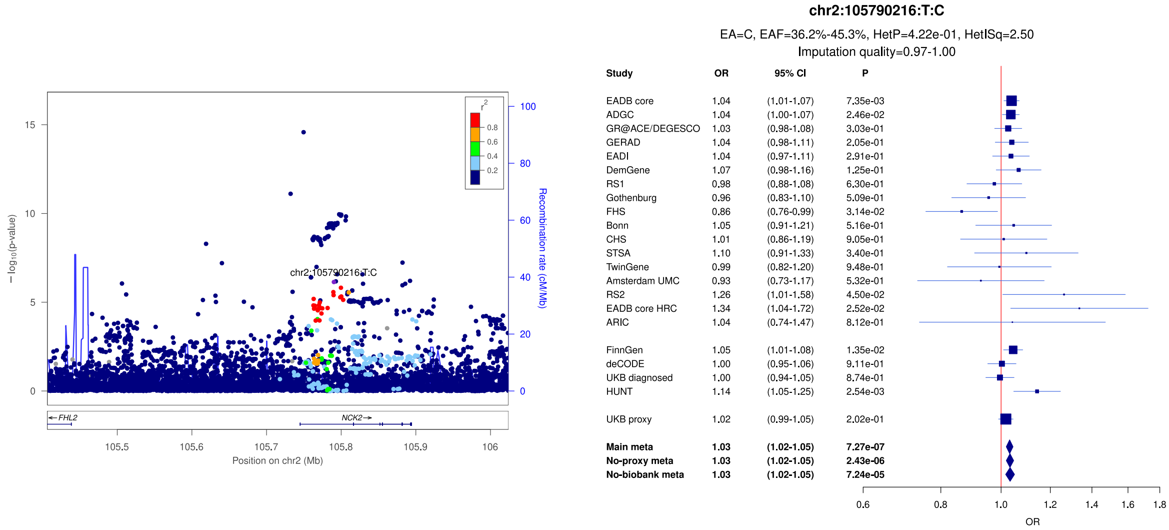

**Supplementary Figure 7:** LocusZoom and forest plots of the main analysis results for (a) *NCK2* (secondary signal) (b) *BIN1* (secondary signal) and (c) *BIN1* (lead signal). The forest plots also show the results of the no-proxy and no-biobank meta-analyses (where the UKBB diagnosed rather than the UKBB proxy results were used). In the forest plots, data are presented as odds-ratio with 95% confidence interval. P values are two-sided raw P values derived from a fixed-effect meta-analysis. OR: odds ratio, CI: confidence interval, EA: effect allele, EAF: effect allele frequency range across all studies, HetP: heterogeneity P value, HetISq: heterogeneity statistic.

a)

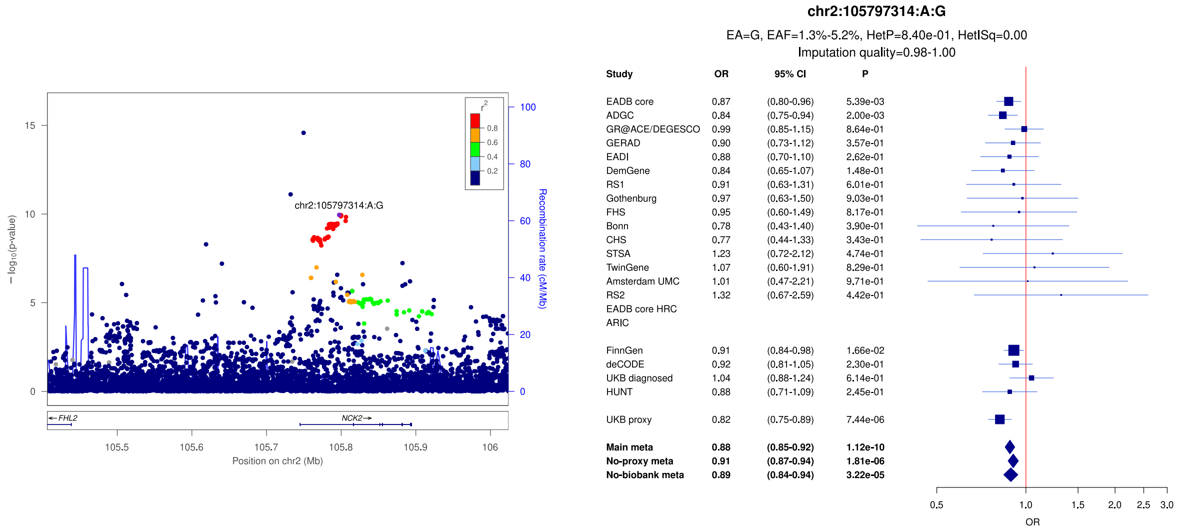


b)

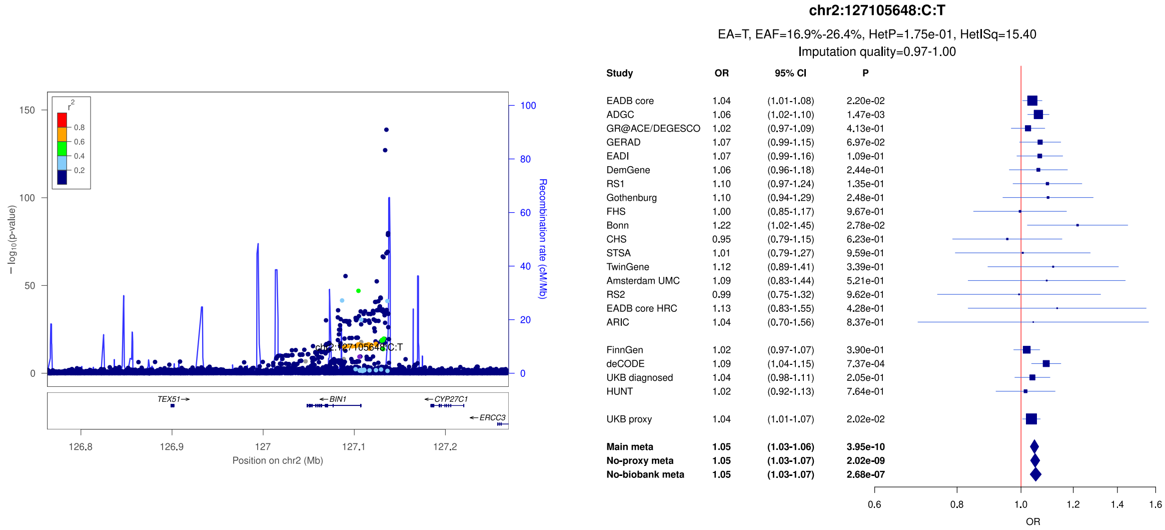


c)

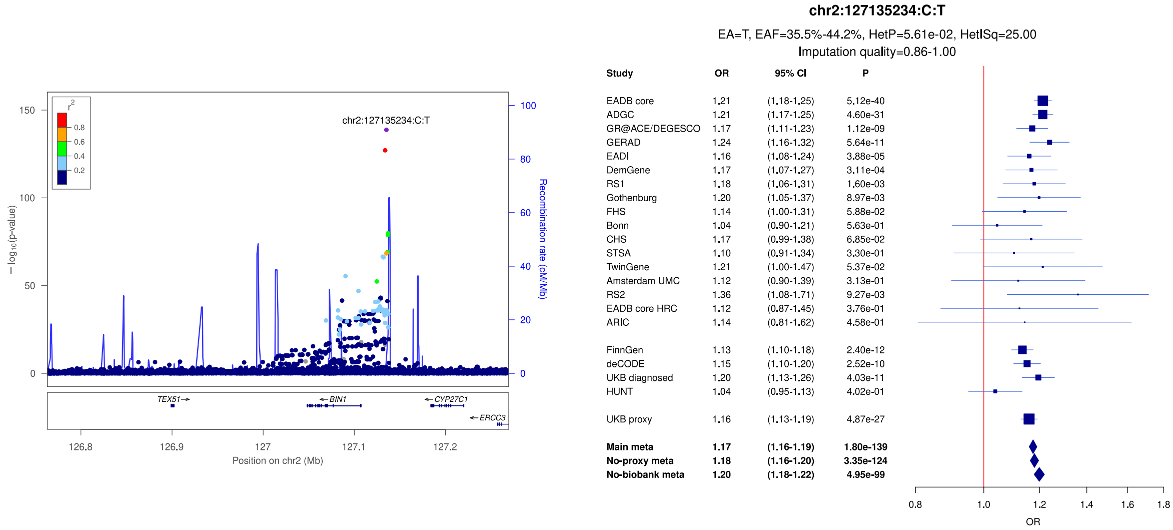

**Supplementary Figure 8:** LocusZoom and forest plots of the main analysis results for (a) *BIN1* (secondary signal) (b) *MGAT5* and (c) *WDR12/ICA1L/CYP20A1*. The forest plots also show the results of the no-proxy and no-biobank meta-analyses (where the UKBB diagnosed rather than the UKBB proxy results were used). In the forest plots, data are presented as odds-ratio with 95% confidence interval. P values are two-sided raw P values derived from a fixed-effect meta-analysis. OR: odds ratio, CI: confidence interval, EA: effect allele, EAF: effect allele frequency range across all studies, HetP: heterogeneity P value, HetISq: heterogeneity statistic.

a)

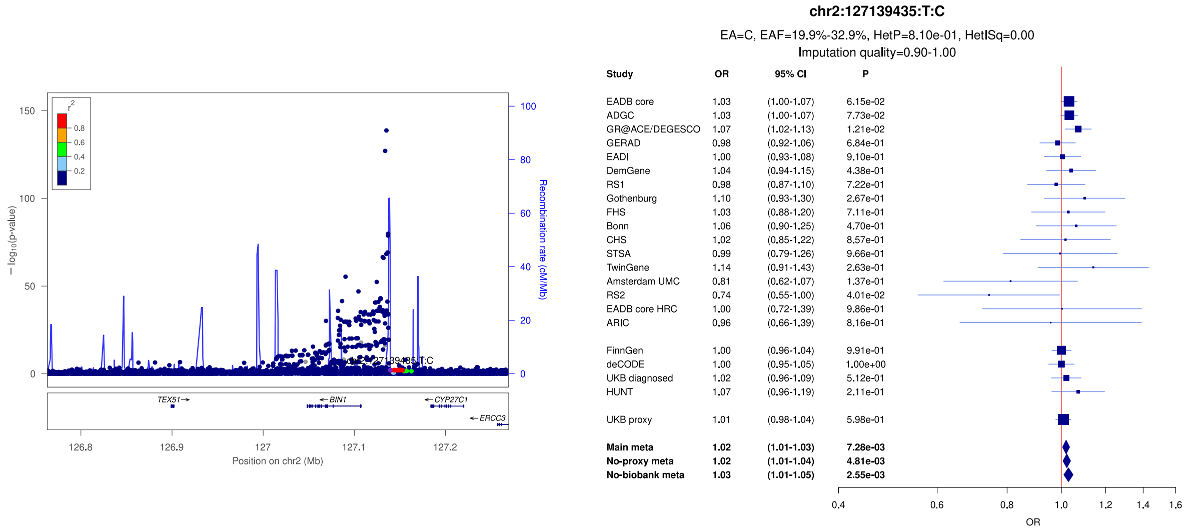


b)

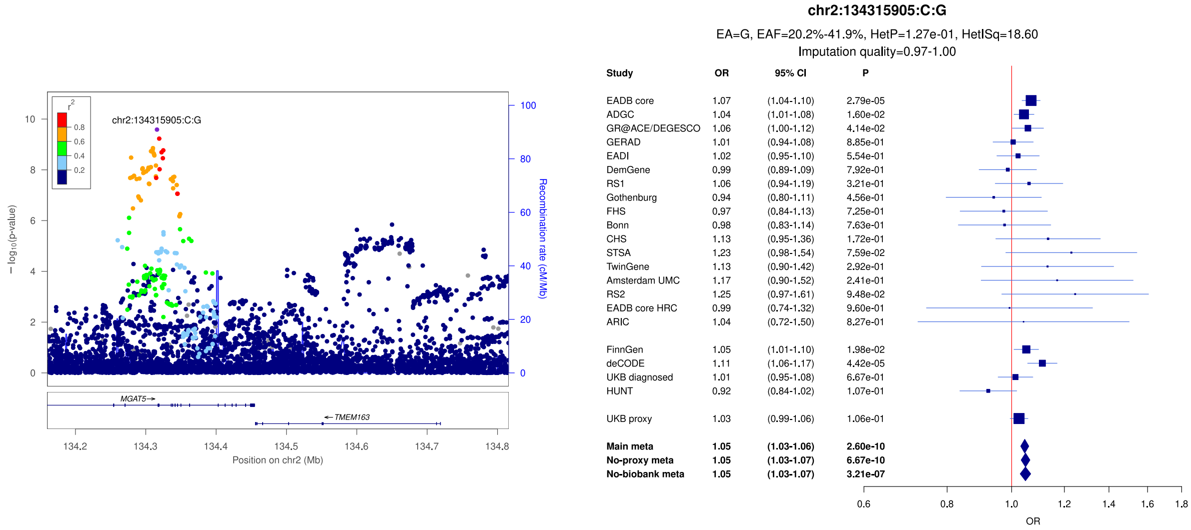


c)

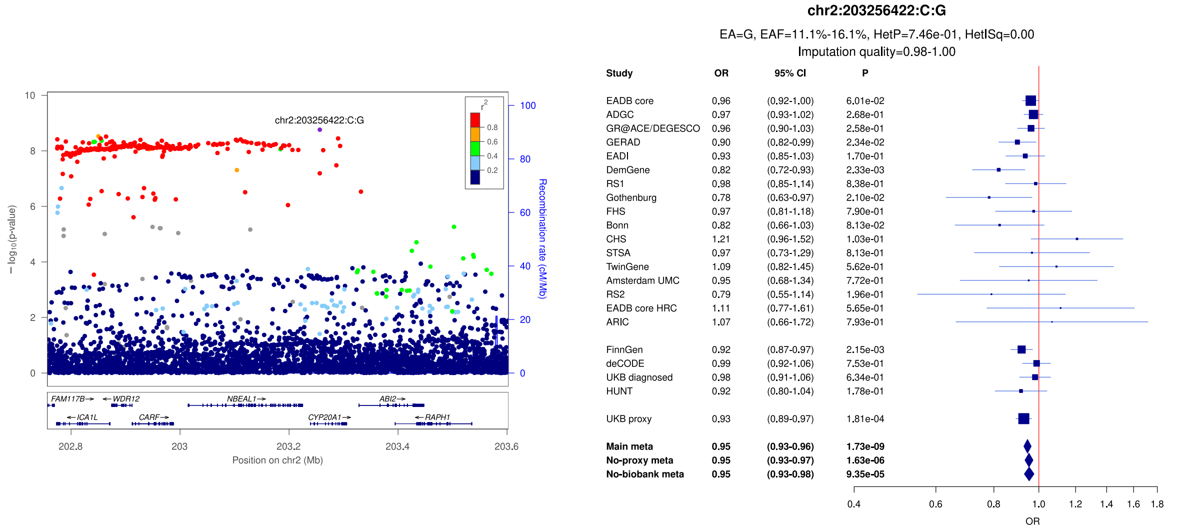

**Supplementary Figure 9:** LocusZoom and forest plots of the main analysis results for (a) *INPP5D* (lead signal) (b) *INPP5D* (secondary signal) and (c) *PPP2R3A*. The forest plots also show the results of the no-proxy and no-biobank meta-analyses (where the UKBB diagnosed rather than the UKBB proxy results were used). In the forest plots, data are presented as odds-ratio with 95% confidence interval. P values are two-sided raw P values derived from a fixed-effect meta-analysis. OR: odds ratio, CI: confidence interval, EA: effect allele, EAF: effect allele frequency range across all studies, HetP: heterogeneity P value, HetISq: heterogeneity statistic.

a)

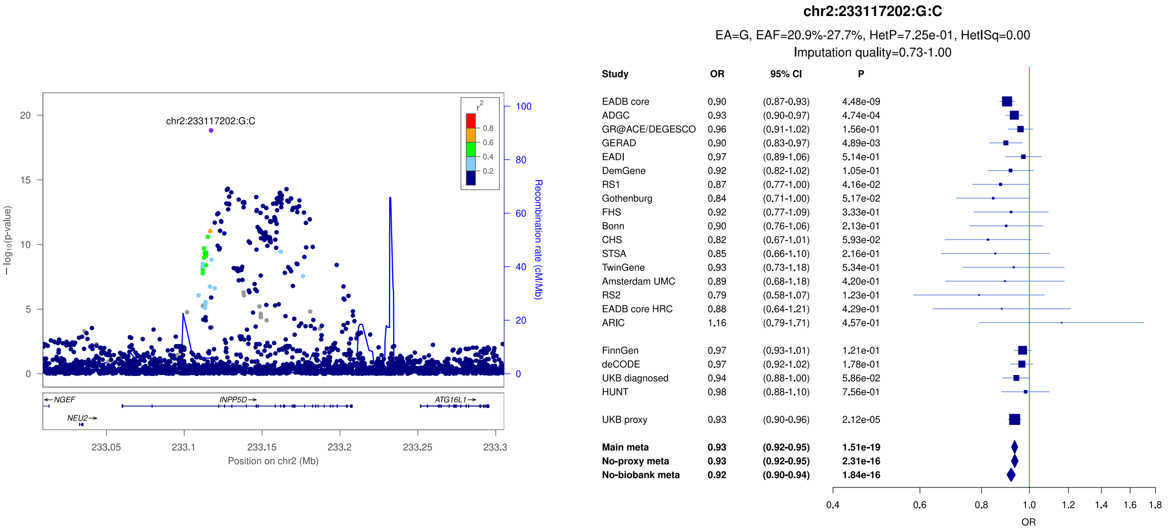


b)

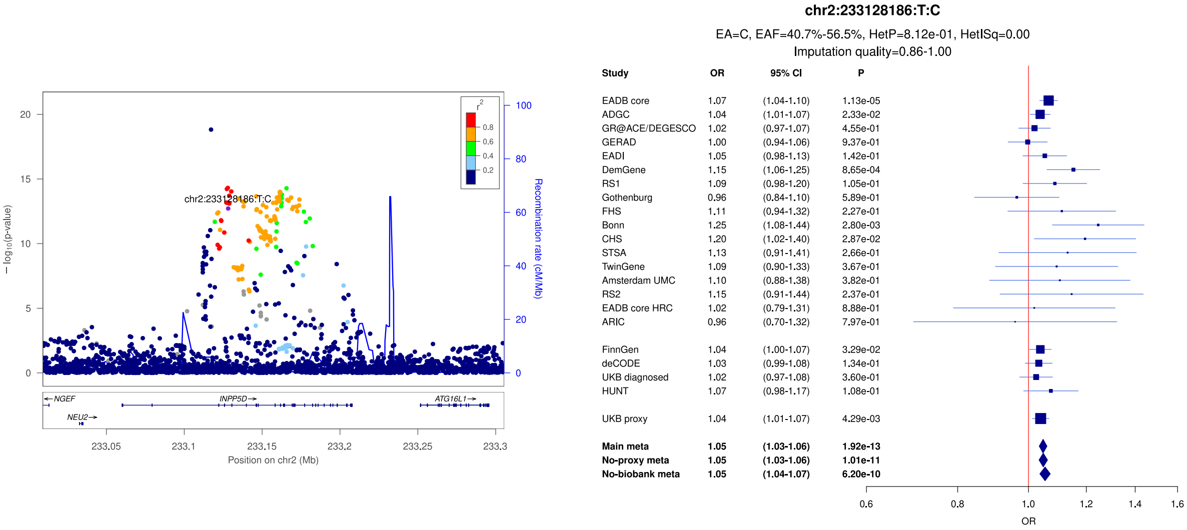


c)

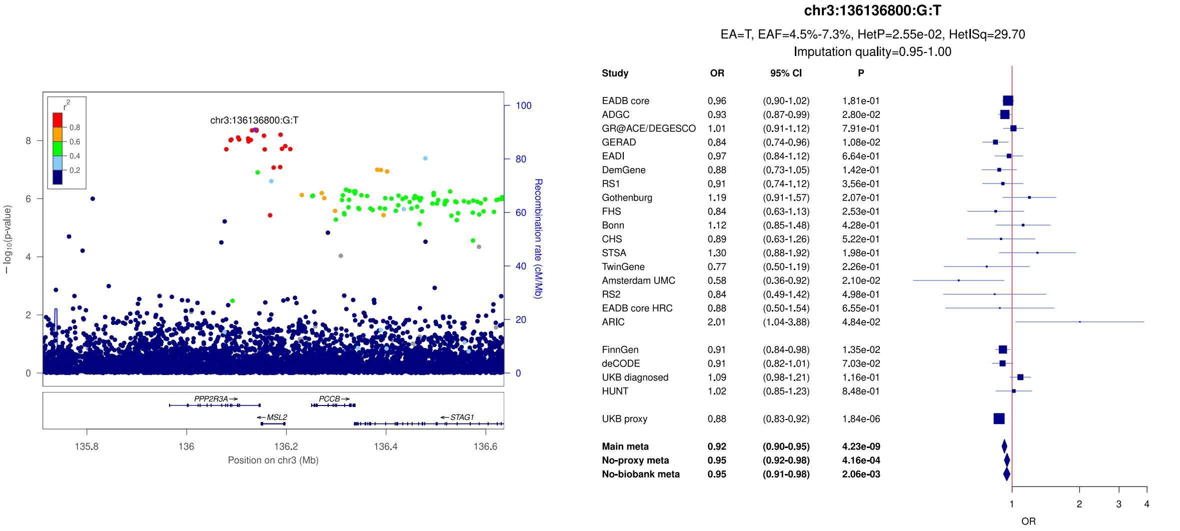

**Supplementary Figure 10:** LocusZoom and forest plots of the main analysis results for (a) *MME* (secondary signal) (b) *MME* (lead signal) and (c) *IDUA/DGKQ* (lead signal). The forest plots also show the results of the no-proxy and no-biobank meta-analyses (where the UKBB diagnosed rather than the UKBB proxy results were used). In the forest plots, data are presented as odds-ratio with 95% confidence interval. P values are two-sided raw P values derived from a fixed-effect meta-analysis. OR: odds ratio, CI: confidence interval, EA: effect allele, EAF: effect allele frequency range across all studies, HetP: heterogeneity P value, HetISq: heterogeneity statistic.

a)

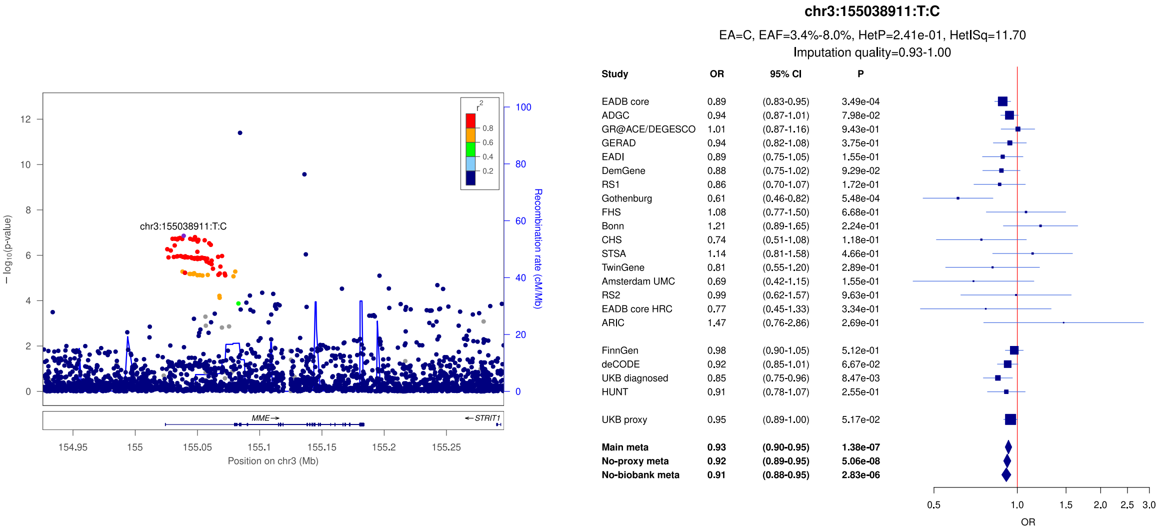


b)

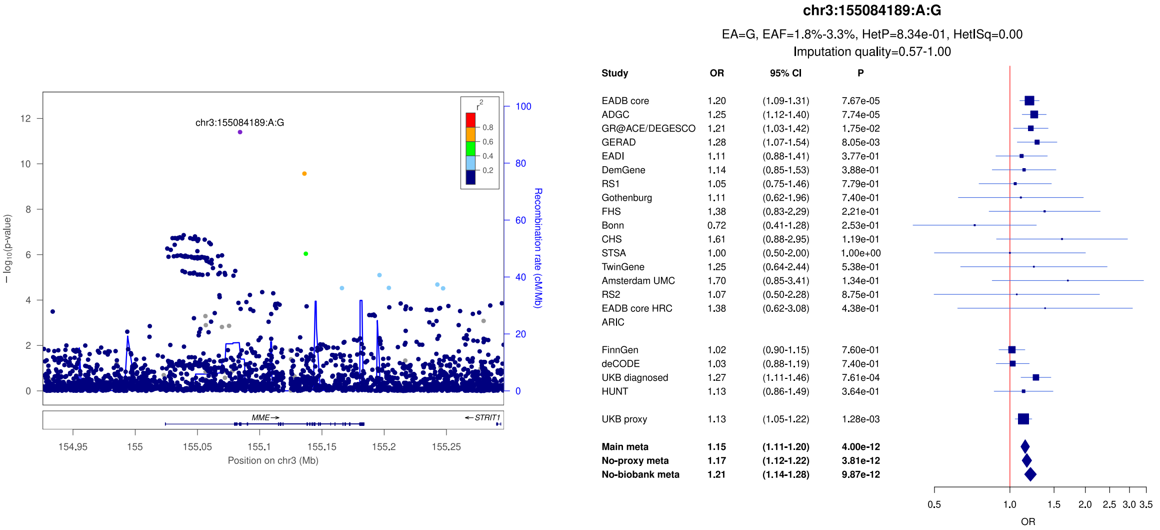


c)

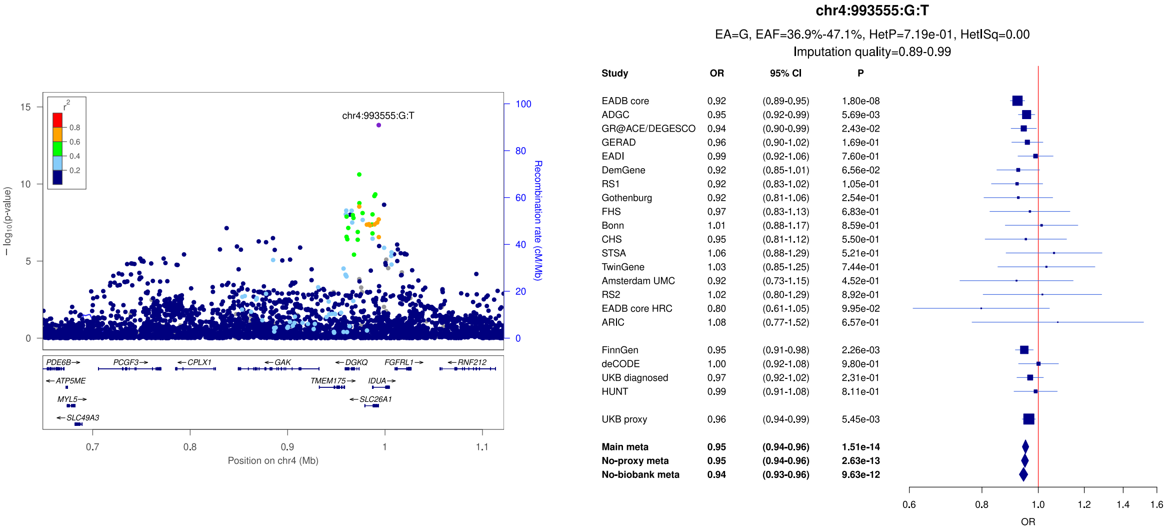

**Supplementary Figure 11:** LocusZoom and forest plots of the main analysis results for (a) *IDUA/DGKQ* (secondary signal) (b) *CLNK* and (c) *RHOH* (lead signal). The forest plots also show the results of the no-proxy and no-biobank meta-analyses (where the UKBB diagnosed rather than the UKBB proxy results were used). In the forest plots, data are presented as odds-ratio with 95% confidence interval. P values are two-sided raw P values derived from a fixed-effect meta-analysis. OR: odds ratio, CI: confidence interval, EA: effect allele, EAF: effect allele frequency range across all studies, HetP: heterogeneity P value, HetISq: heterogeneity statistic.

a)

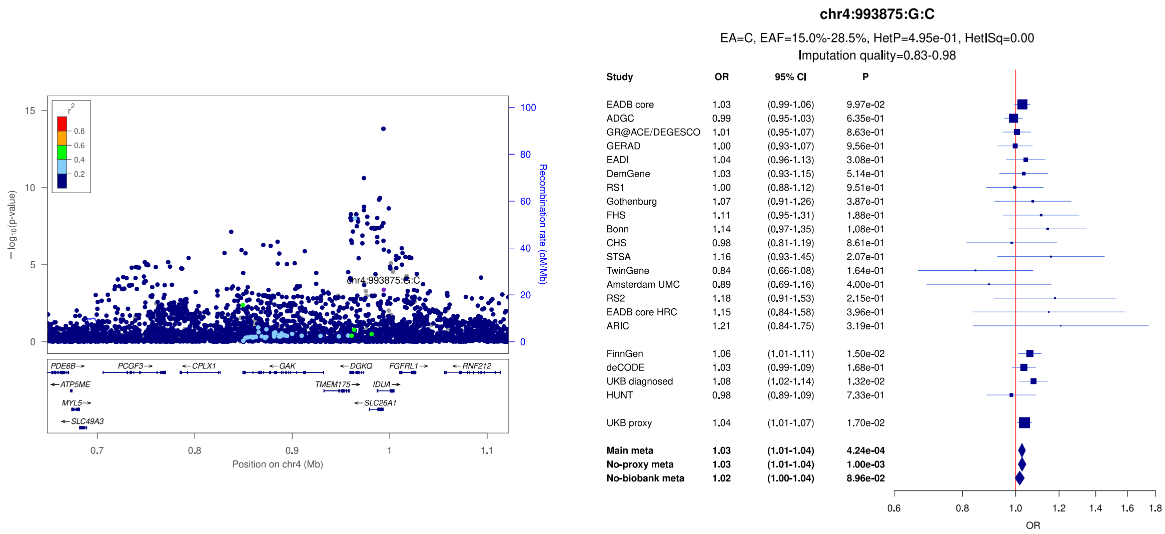


b)

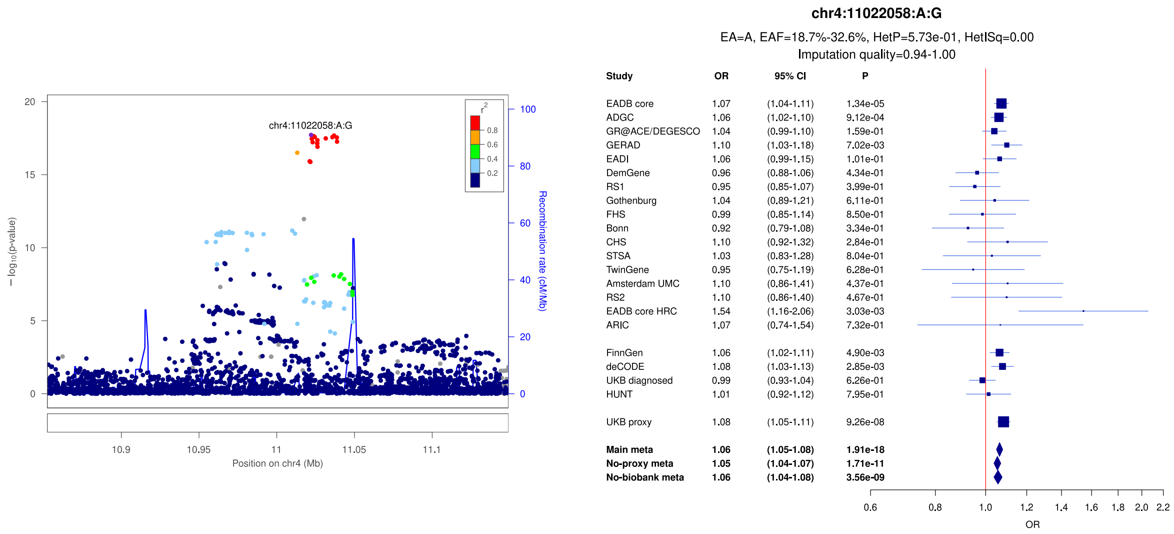


c)

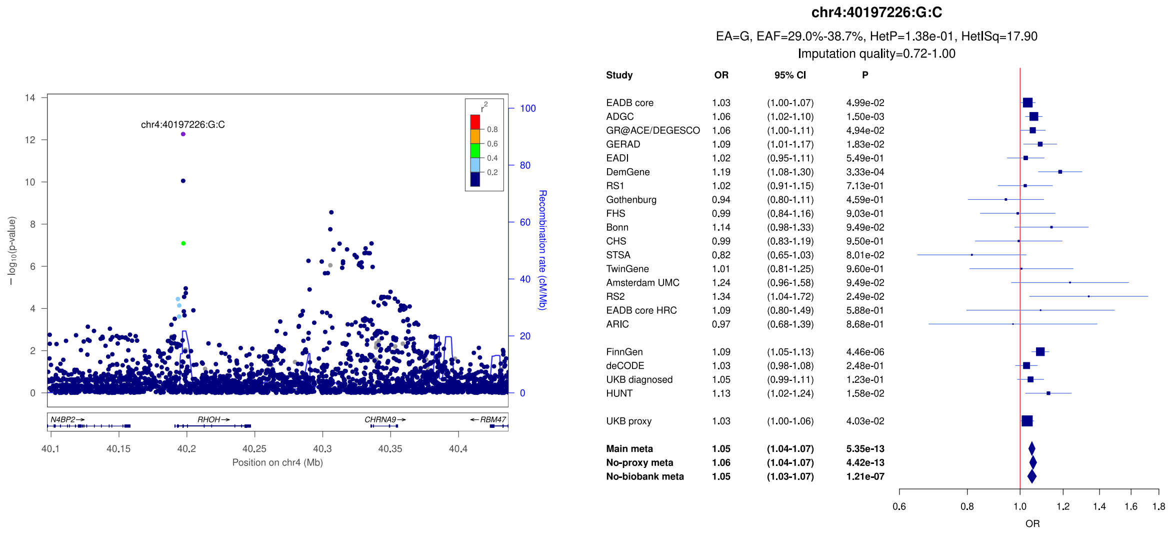

**Supplementary Figure 12:** LocusZoom and forest plots of the main analysis results for (a) *RHOH* (secondary signal) (b) *ADGRL3* and (c) *ANKH/OTULIN*. The forest plots also show the results of the no-proxy and no-biobank meta-analyses (where the UKBB diagnosed rather than the UKBB proxy results were used). In the forest plots, data are presented as odds-ratio with 95% confidence interval. P values are two-sided raw P values derived from a fixed-effect meta-analysis. OR: odds ratio, CI: confidence interval, EA: effect allele, EAF: effect allele frequency range across all studies, HetP: heterogeneity P value, HetISq: heterogeneity statistic.

a)

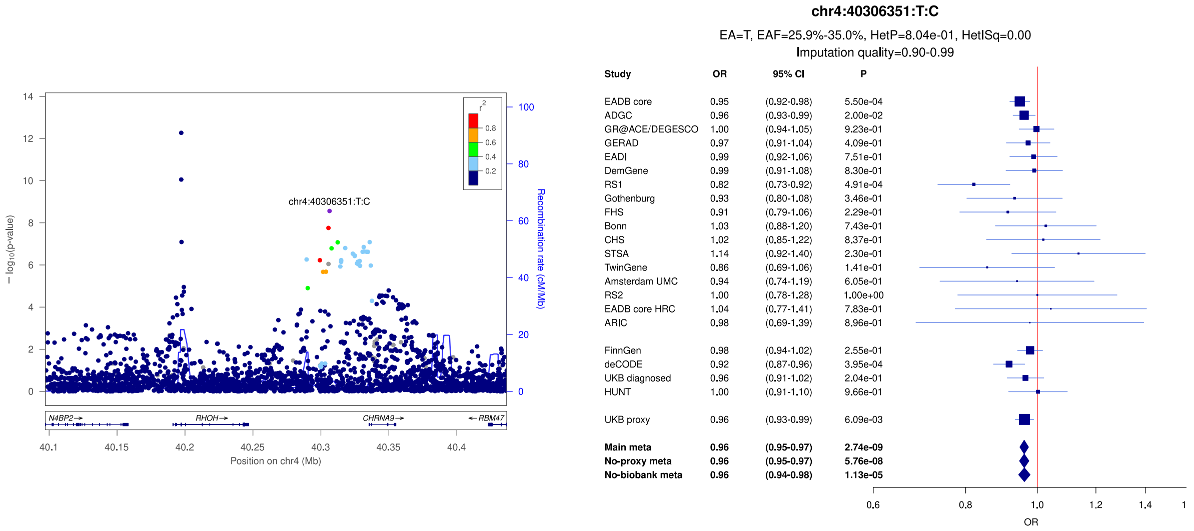


b)

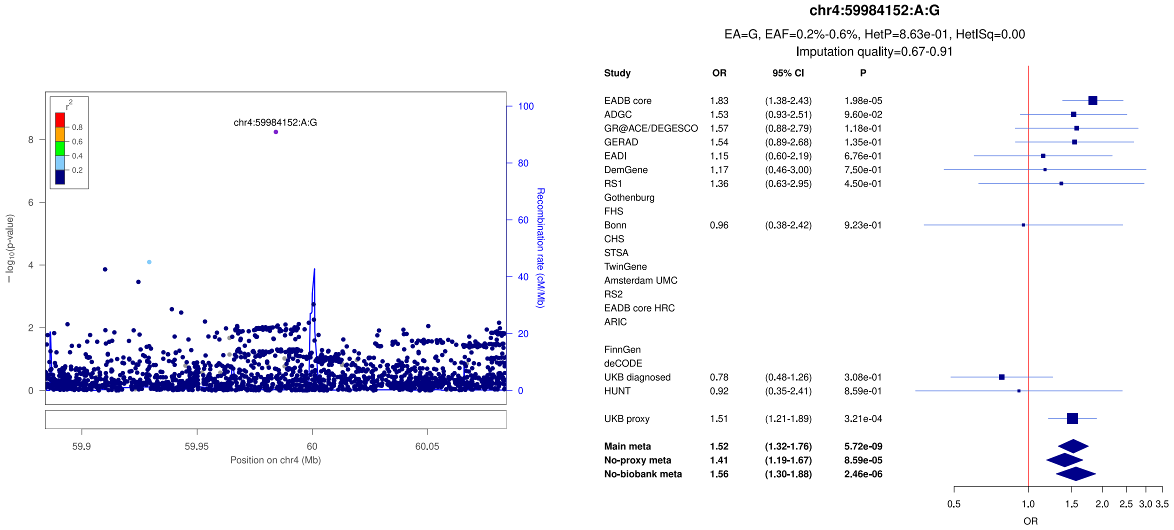


c)

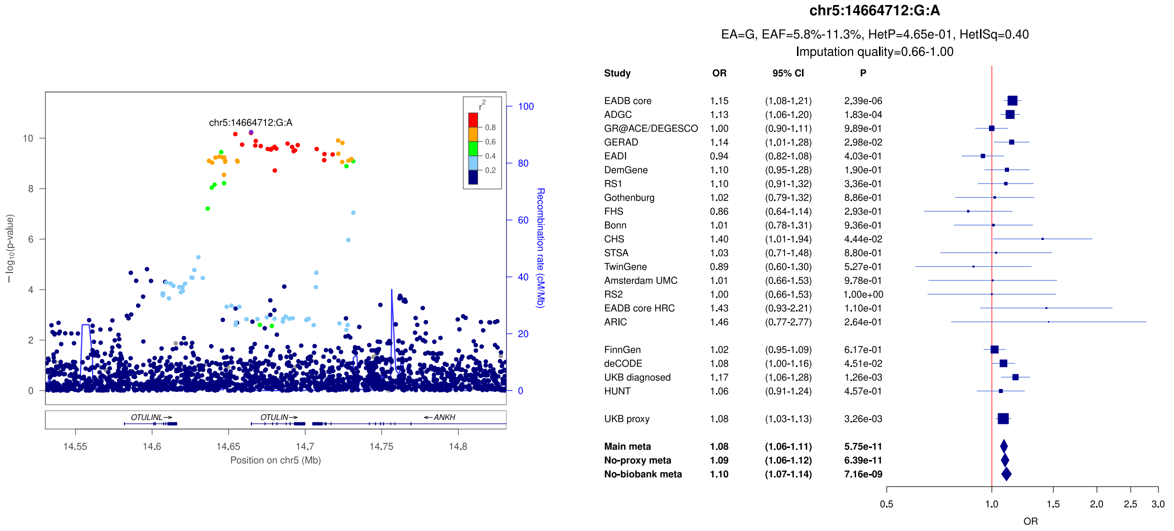

**Supplementary Figure 13:** LocusZoom and forest plots of the main analysis results for (a) *COX7C* (b) *HBEGF* and (c) *TNIP1*. The forest plots also show the results of the no-proxy and no-biobank meta-analyses (where the UKBB diagnosed rather than the UKBB proxy results were used). In the forest plots, data are presented as odds-ratio with 95% confidence interval. P values are two-sided raw P values derived from a fixed-effect meta-analysis. OR: odds ratio, CI: confidence interval, EA: effect allele, EAF: effect allele frequency range across all studies, HetP: heterogeneity P value, HetISq: heterogeneity statistic.

a)

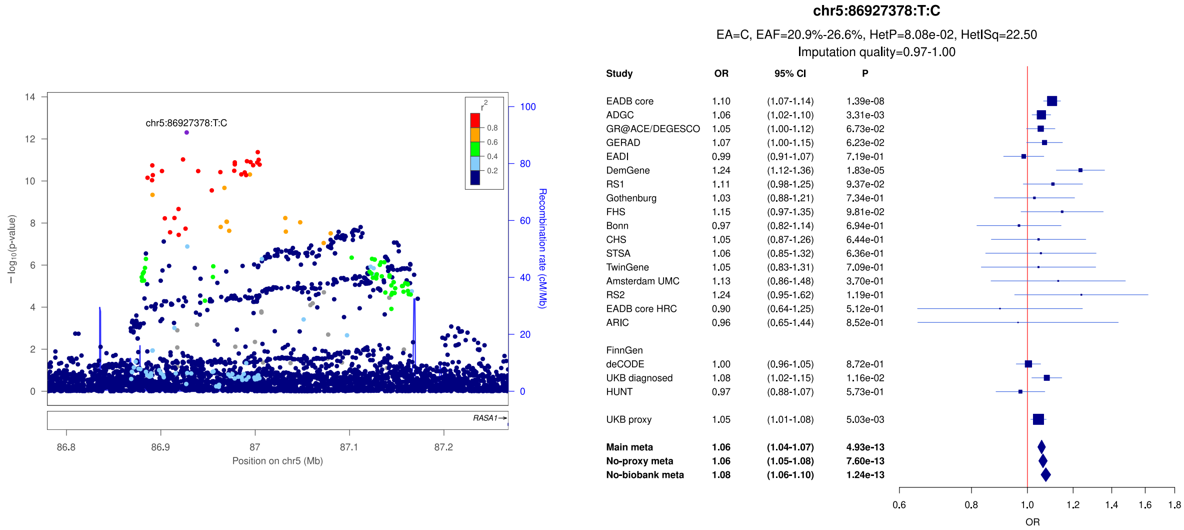


b)

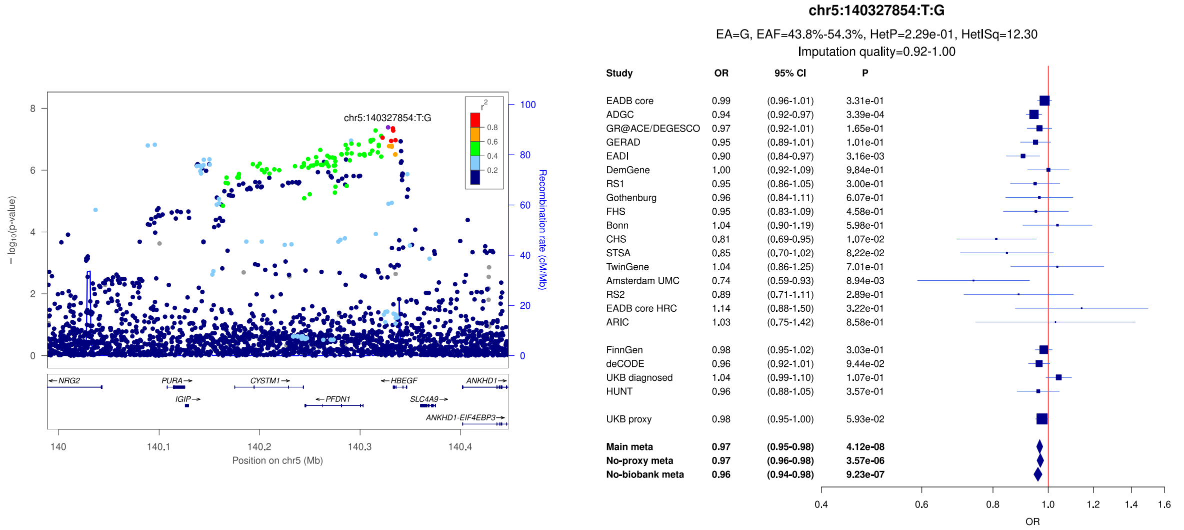


c)

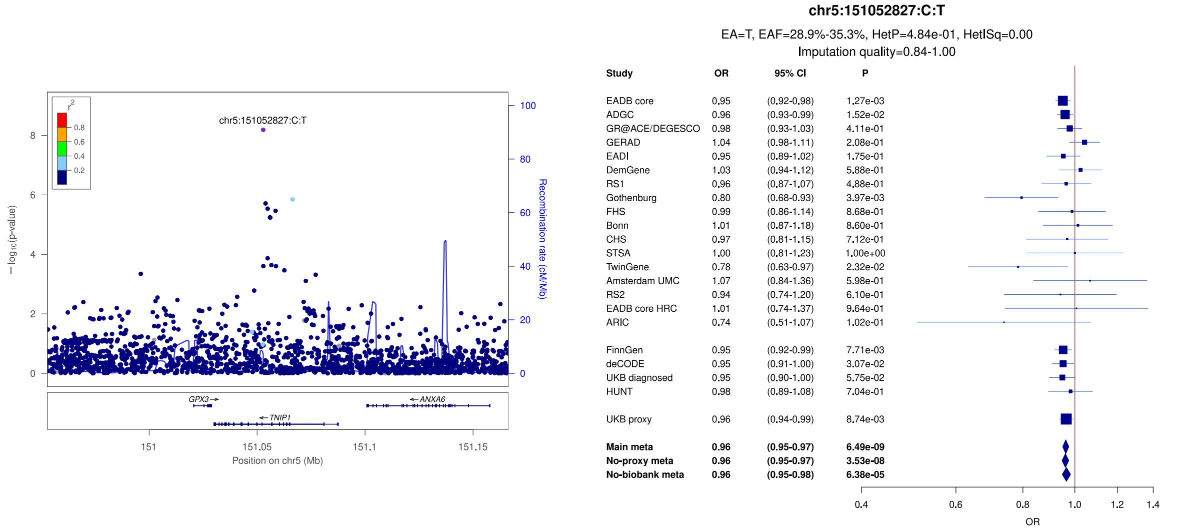

**Supplementary Figure 14:** LocusZoom and forest plots of the main analysis results for (a) *FAM193B* (secondary signal) (b) *FAM193B* (lead signal) and (c) *RASGEF1C* (secondary signal). The forest plots also show the results of the no-proxy and no-biobank meta-analyses (where the UKBB diagnosed rather than the UKBB proxy results were used). In the forest plots, data are presented as odds-ratio with 95% confidence interval. P values are two-sided raw P values derived from a fixed-effect meta-analysis. OR: odds ratio, CI: confidence interval, EA: effect allele, EAF: effect allele frequency range across all studies, HetP: heterogeneity P value, HetISq: heterogeneity statistic.

a)

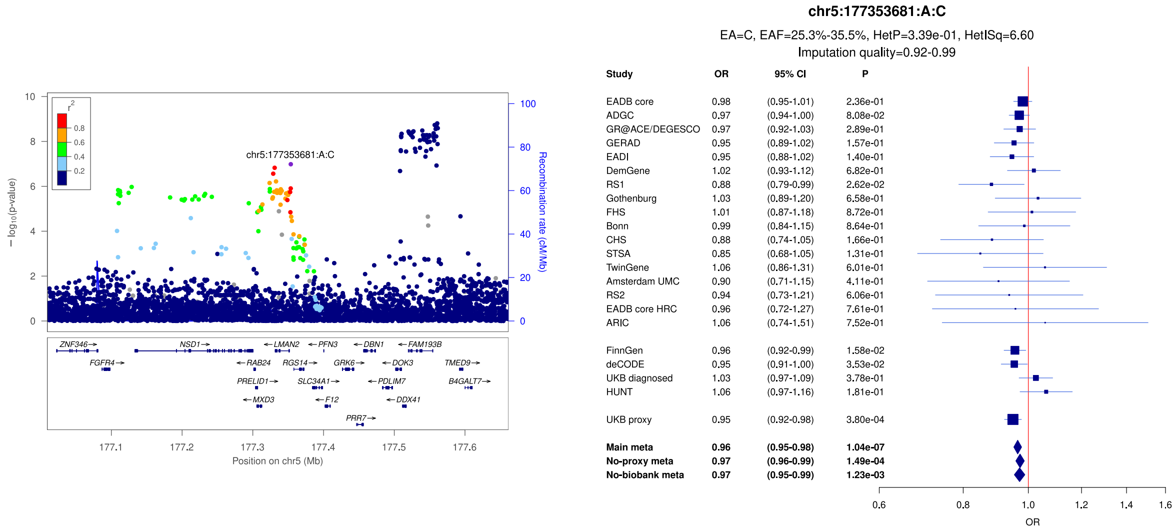


b)

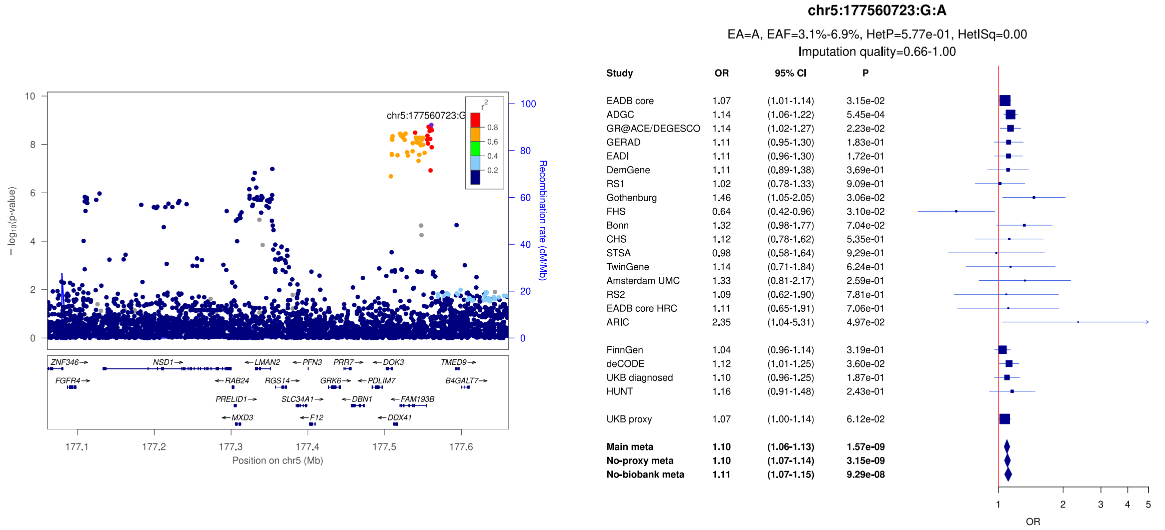


c)

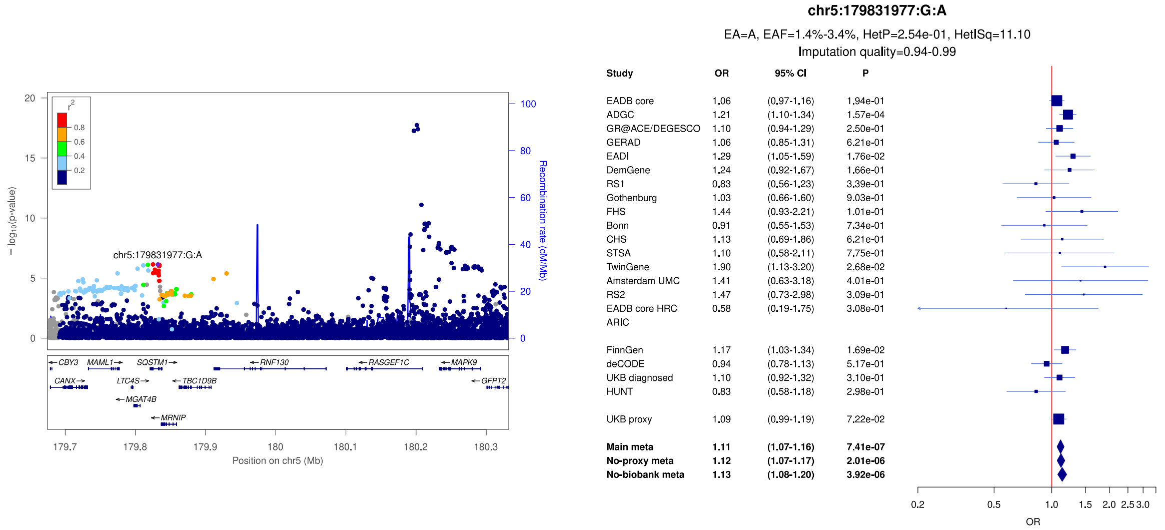

**Supplementary Figure 15:** LocusZoom and forest plots of the main analysis results for (a) *RASGEF1C* (lead signal) (b) *HLA* (secondary signal) and (c) *HLA* (secondary signal). The forest plots also show the results of the no-proxy and no-biobank meta-analyses (where the UKBB diagnosed rather than the UKBB proxy results were used). In the forest plots, data are presented as odds-ratio with 95% confidence interval. P values are two-sided raw P values derived from a fixed-effect meta-analysis. OR: odds ratio, CI: confidence interval, EA: effect allele, EAF: effect allele frequency range across all studies, HetP: heterogeneity P value, HetISq: heterogeneity statistic.

a)

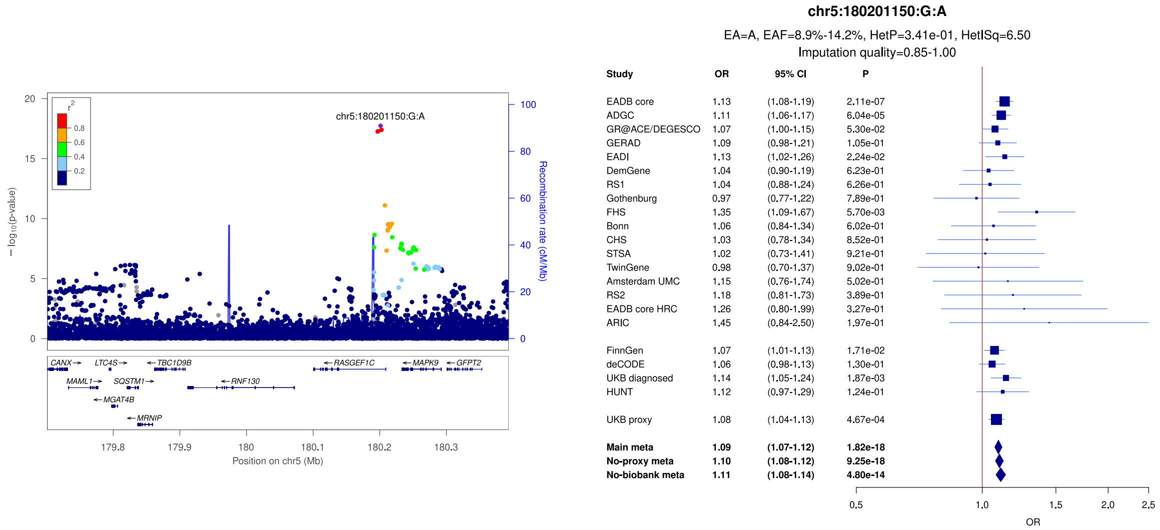


b)

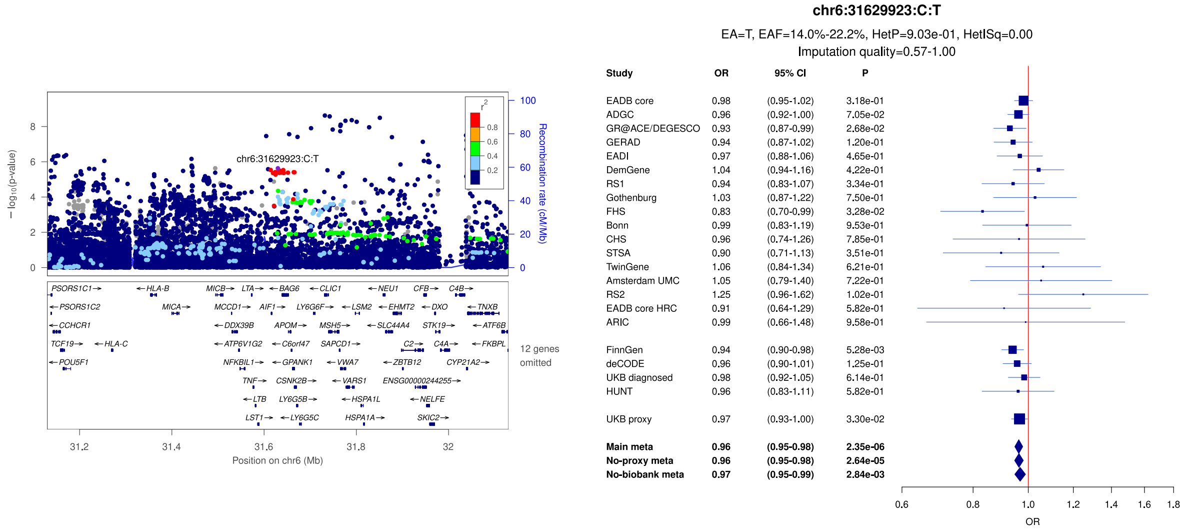


c)

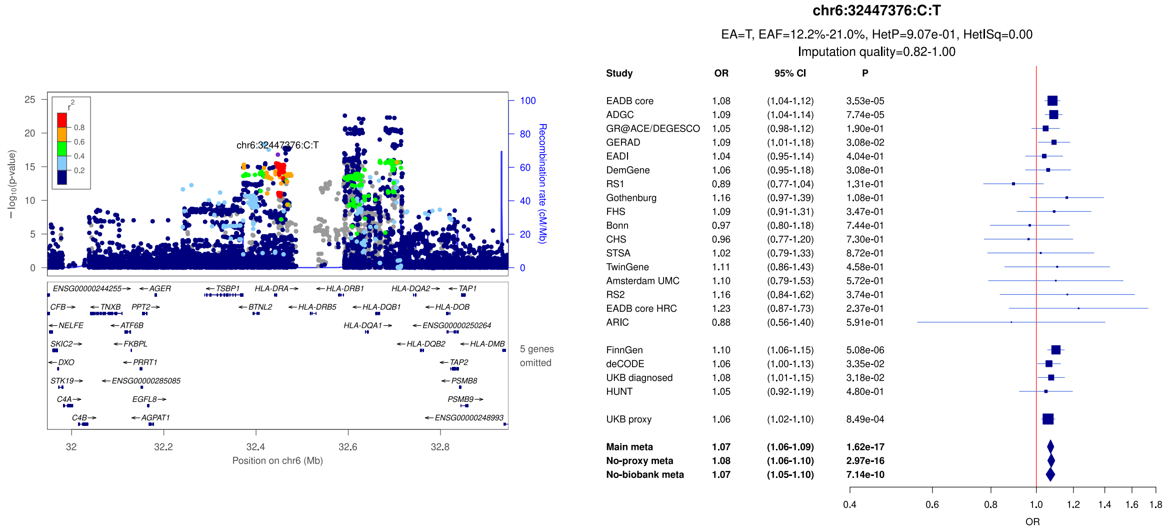

**Supplementary Figure 16:** LocusZoom and forest plots of the main analysis results for (a) *HLA* (lead signal) (b) *TREM2* (secondary signal) and (c) *TREM2* (secondary signal). The forest plots also show the results of the no-proxy and no-biobank meta-analyses (where the UKBB diagnosed rather than the UKBB proxy results were used). In the forest plots, data are presented as odds-ratio with 95% confidence interval. P values are two-sided raw P values derived from a fixed-effect meta-analysis. OR: odds ratio, CI: confidence interval, EA: effect allele, EAF: effect allele frequency range across all studies, HetP: heterogeneity P value, HetISq: heterogeneity statistic.

a)

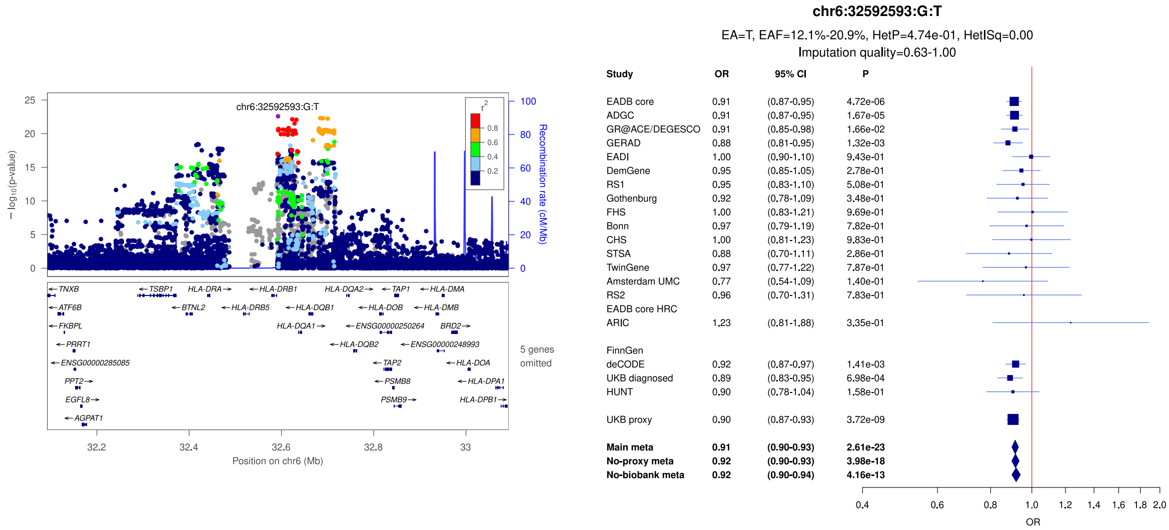


b)

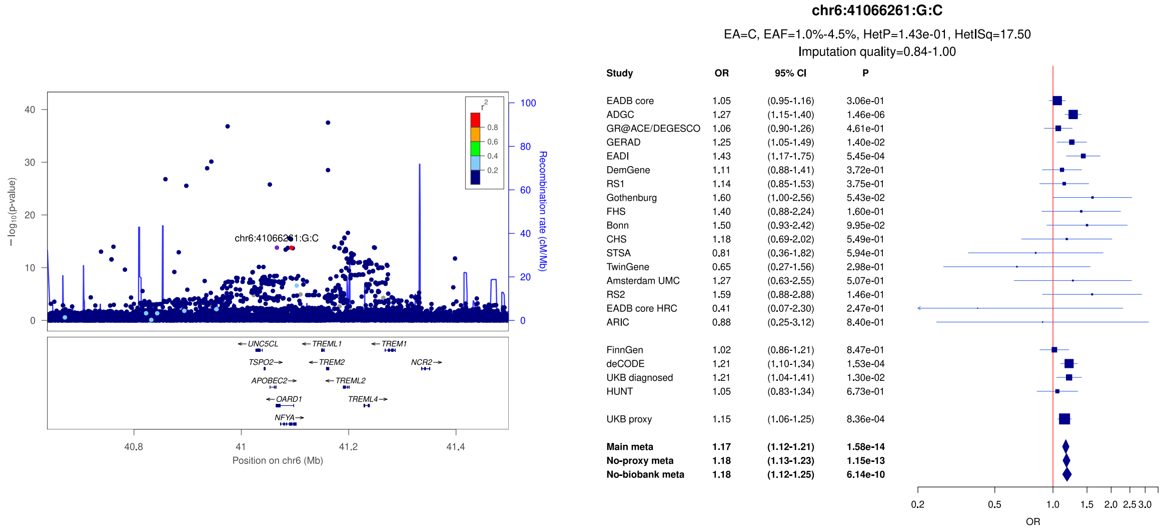


c)

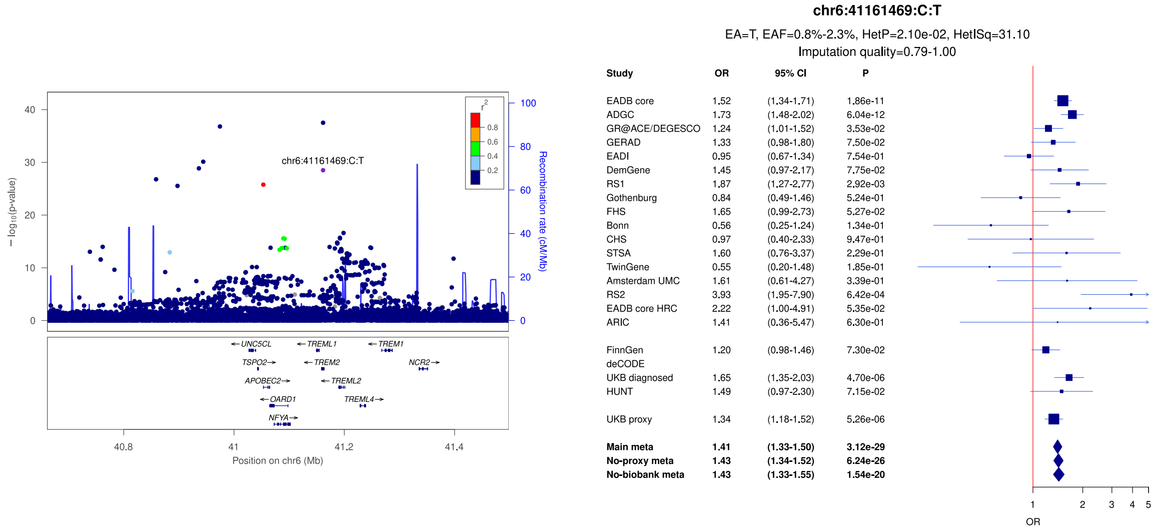

**Supplementary Figure 17:** LocusZoom and forest plots of the main analysis results for (a) *TREM2* (lead signal) (b) *TREM2* (secondary signal) and (c) *TREM2* (secondary signal). The forest plots also show the results of the no-proxy and no-biobank meta-analyses (where the UKBB diagnosed rather than the UKBB proxy results were used). In the forest plots, data are presented as odds-ratio with 95% confidence interval. P values are two-sided raw P values derived from a fixed-effect meta-analysis. OR: odds ratio, CI: confidence interval, EA: effect allele, EAF: effect allele frequency range across all studies, HetP: heterogeneity P value, HetISq: heterogeneity statistic.

a)

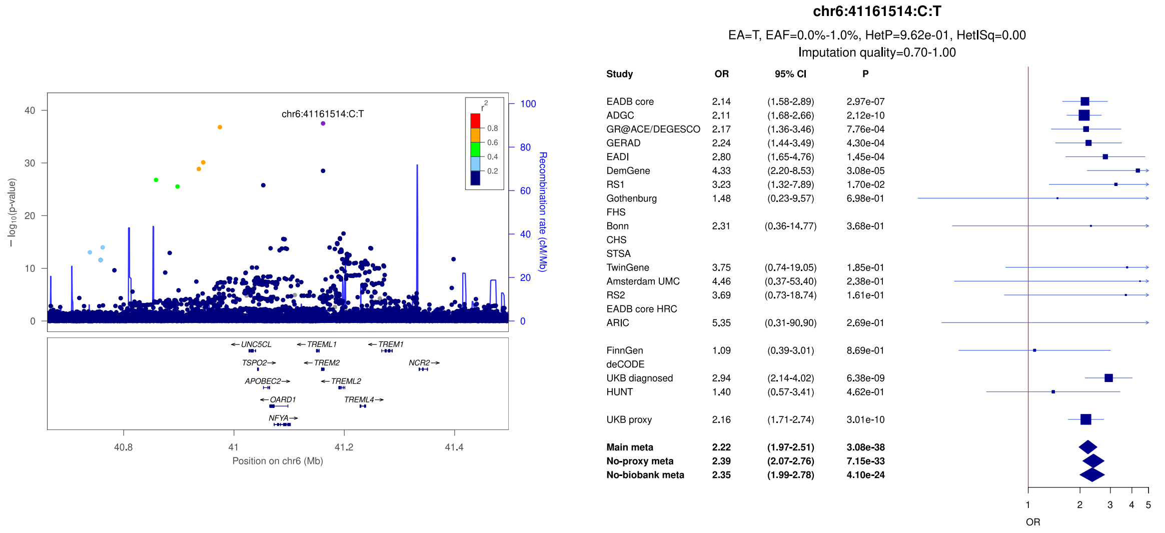


b)

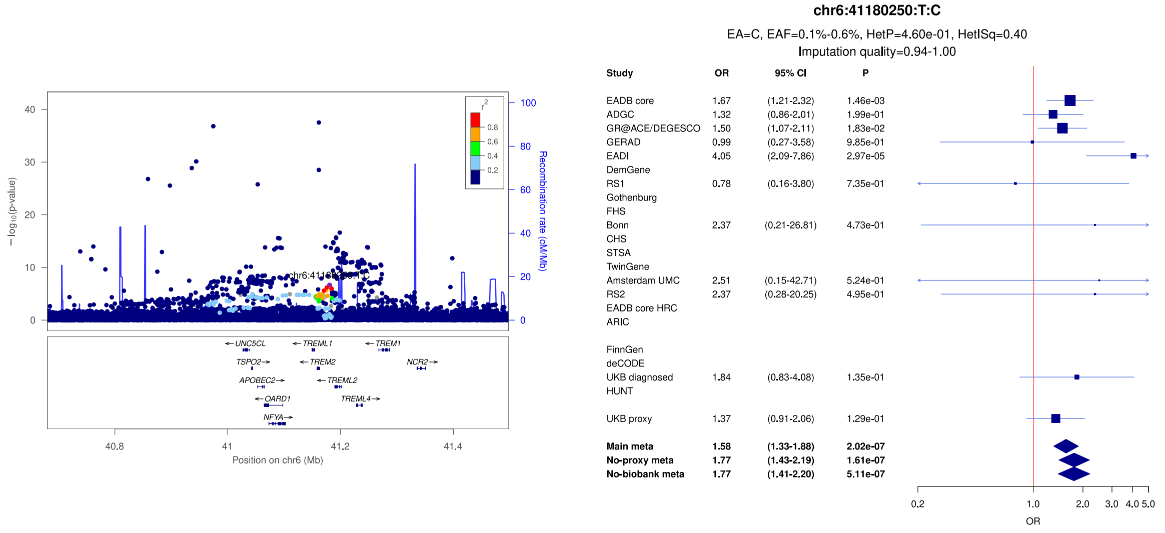


c)

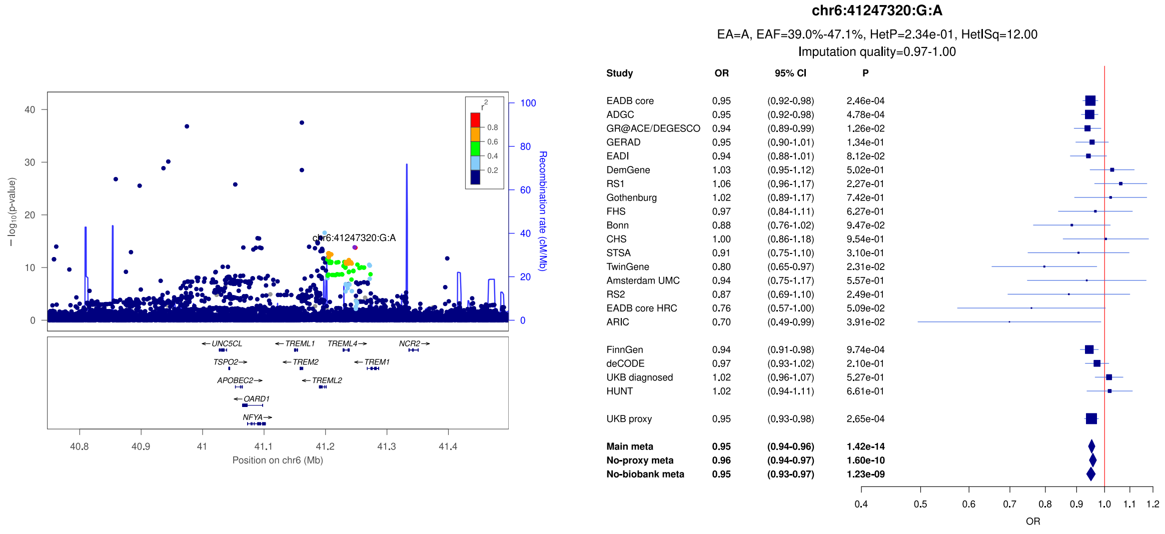

**Supplementary Figure 18:** LocusZoom and forest plots of the main analysis results for (a) *CD2AP* (b) *HS3ST5* and (c) *TMEM184A*. The forest plots also show the results of the no-proxy and no-biobank meta-analyses (where the UKBB diagnosed rather than the UKBB proxy results were used). In the forest plots, data are presented as odds-ratio with 95% confidence interval. P values are two-sided raw P values derived from a fixed-effect meta-analysis. OR: odds ratio, CI: confidence interval, EA: effect allele, EAF: effect allele frequency range across all studies, HetP: heterogeneity P value, HetISq: heterogeneity statistic.

a)

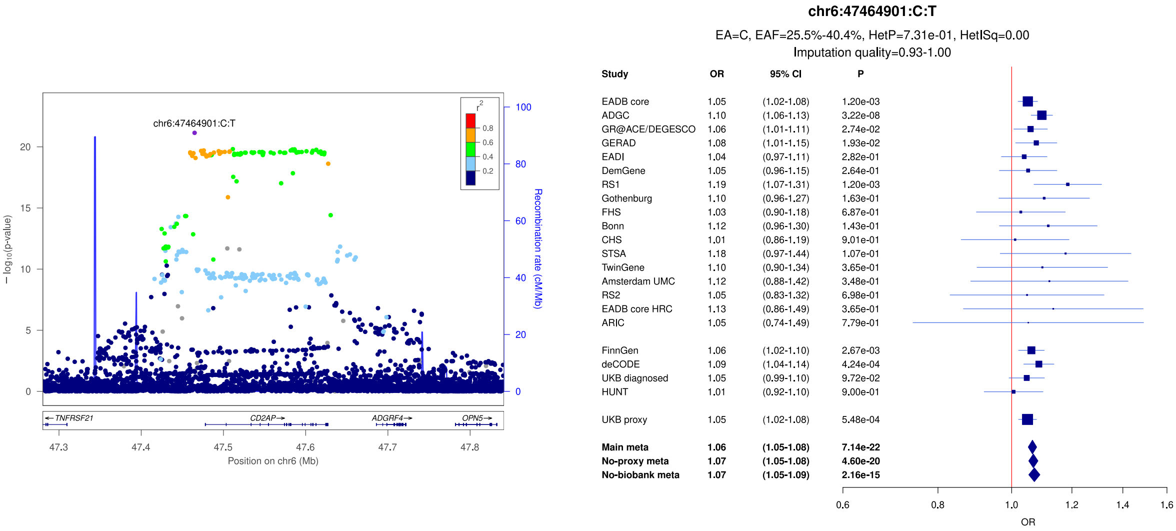


b)

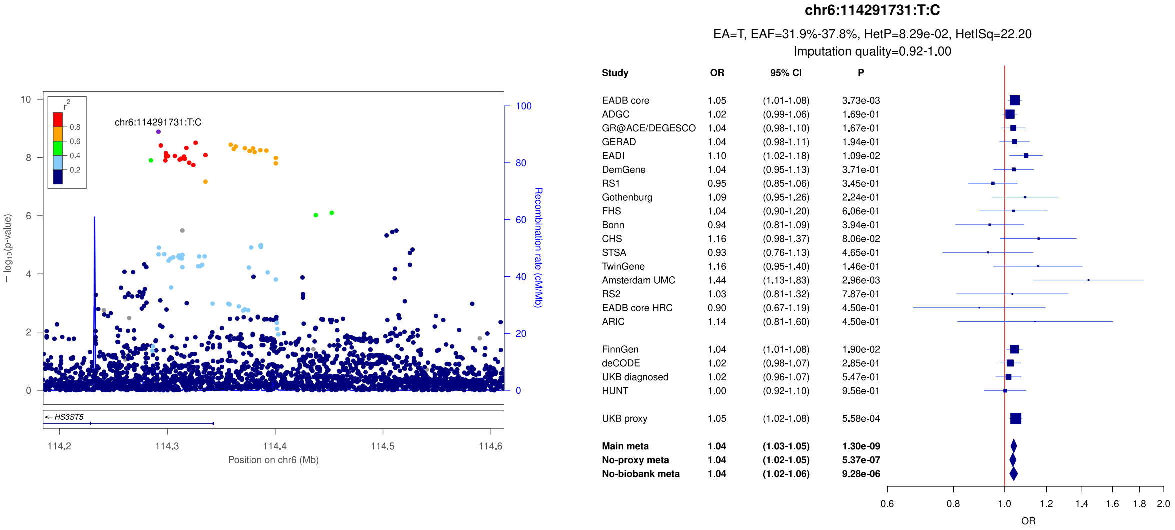


c)

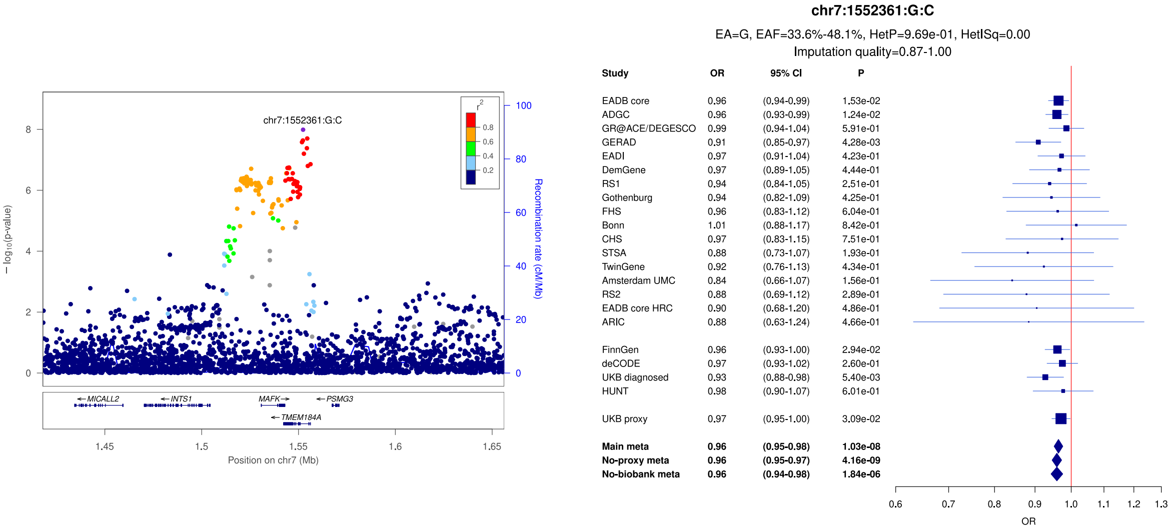

**Supplementary Figure 19:** LocusZoom and forest plots of the main analysis results for (a) *ICA1/UMAD1* (lead signal) (b) *ICA1/UMAD1* (secondary signal) and (c) *TMEM106B*. The forest plots also show the results of the no-proxy and no-biobank meta-analyses (where the UKBB diagnosed rather than the UKBB proxy results were used). In the forest plots, data are presented as odds-ratio with 95% confidence interval. P values are two-sided raw P values derived from a fixed-effect meta-analysis. OR: odds ratio, CI: confidence interval, EA: effect allele, EAF: effect allele frequency range across all studies, HetP: heterogeneity P value, HetISq: heterogeneity statistic.

a)

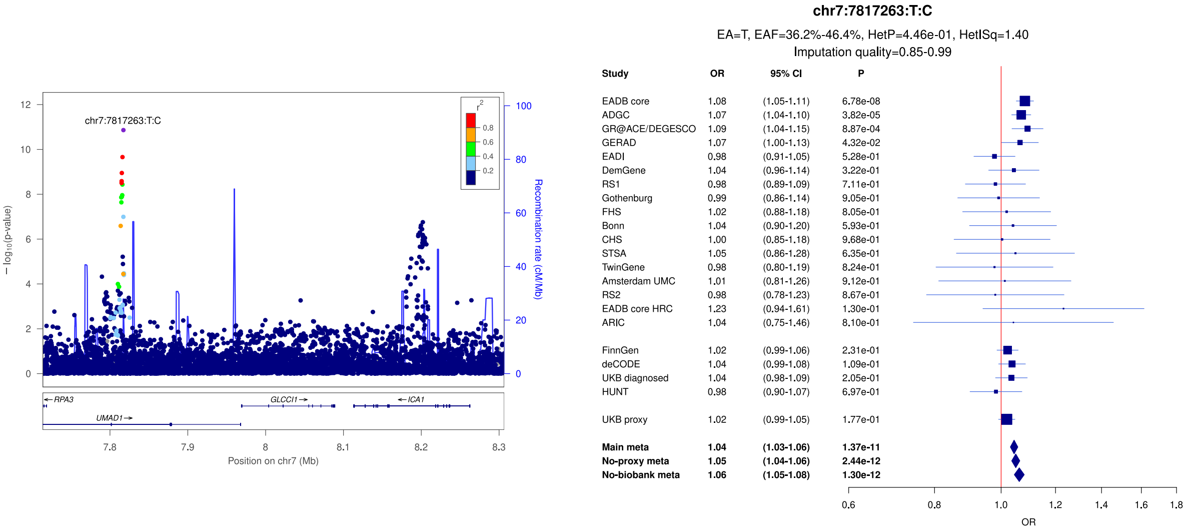


b)

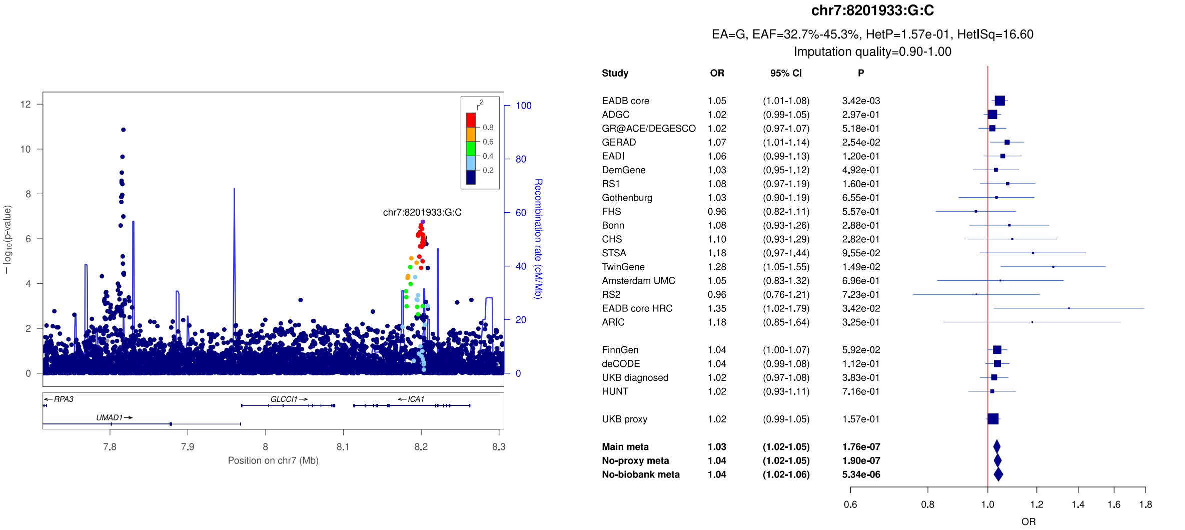


c)

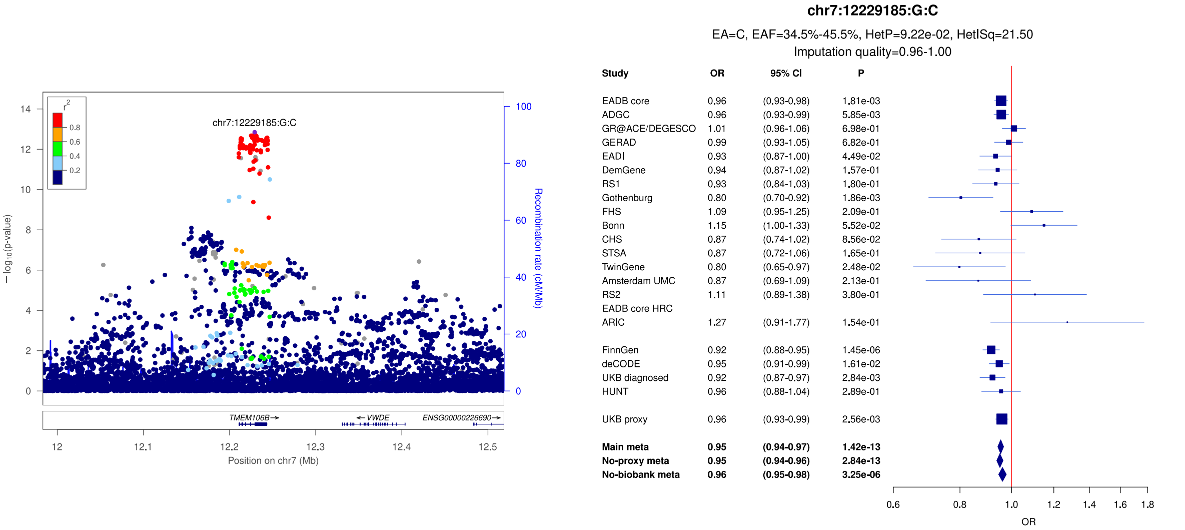

**Supplementary Figure 20:** LocusZoom and forest plots of the main analysis results for (a) *JAZF1* (b) *NME8* (secondary signal) and (c) *NME8* (lead signal). The forest plots also show the results of the no-proxy and no-biobank meta-analyses (where the UKBB diagnosed rather than the UKBB proxy results were used). In the forest plots, data are presented as odds-ratio with 95% confidence interval. P values are two-sided raw P values derived from a fixed-effect meta-analysis. OR: odds ratio, CI: confidence interval, EA: effect allele, EAF: effect allele frequency range across all studies, HetP: heterogeneity P value, HetISq: heterogeneity statistic.

a)

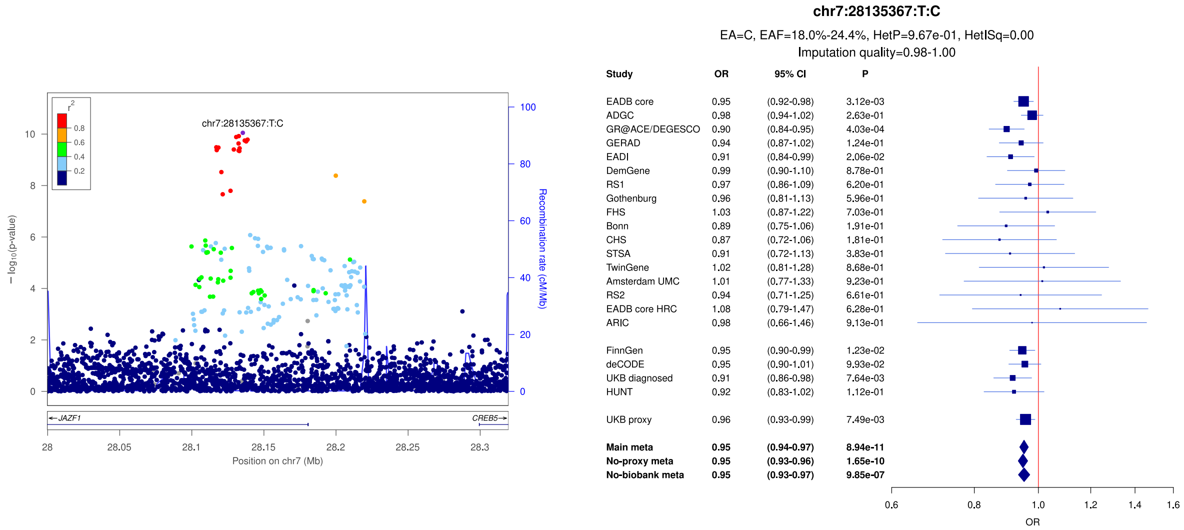


b)

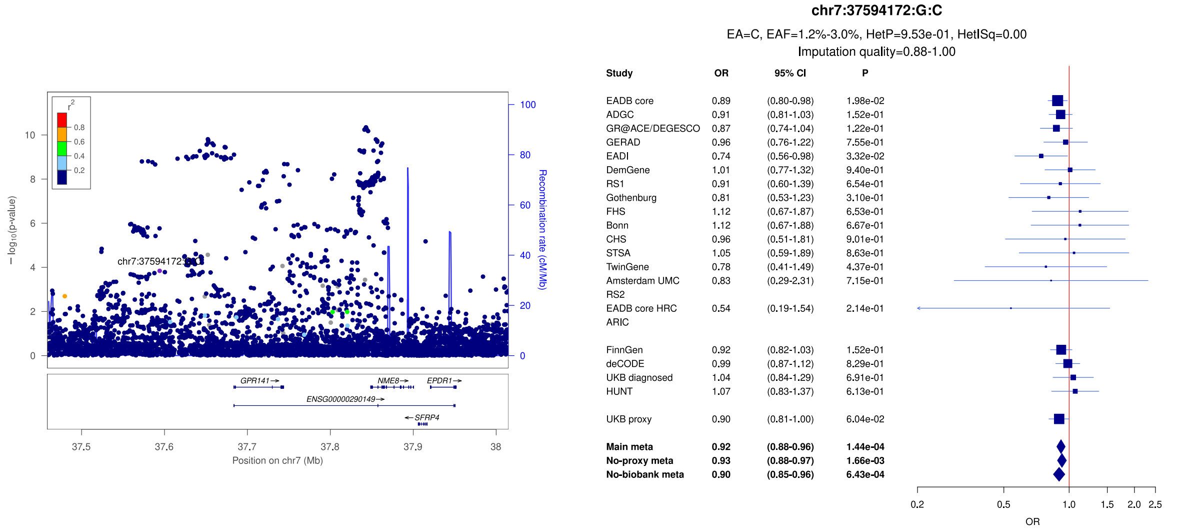


c)

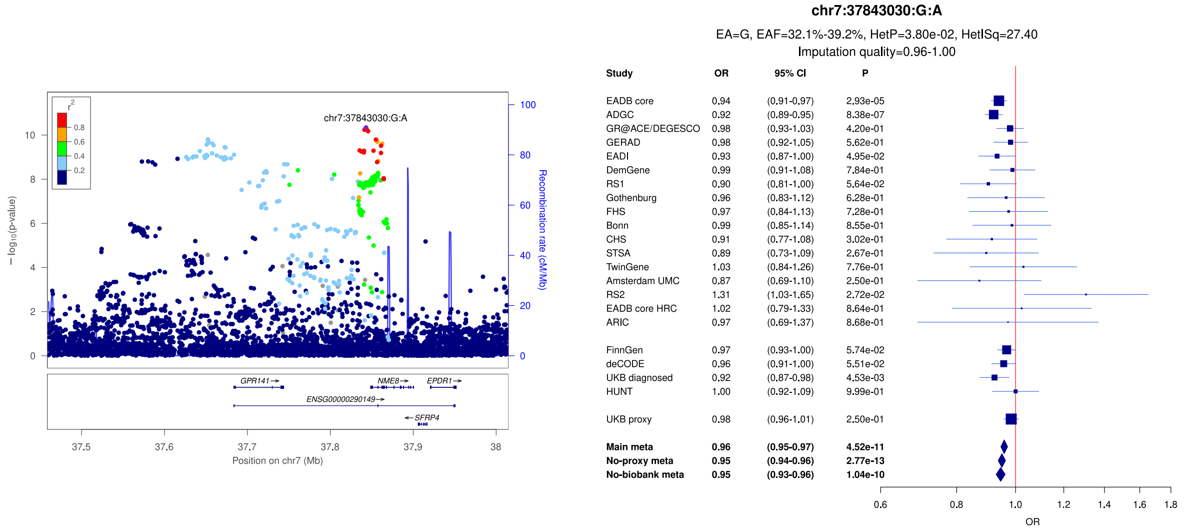

**Supplementary Figure 21:** LocusZoom and forest plots of the main analysis results for (a) *SEC61G /EGFR* (lead signal) (b) *SEC61G/EGFR* (secondary signal) and (c) *ZCWPW1/NYAP1* (secondary signal). The forest plots also show the results of the no-proxy and no-biobank meta-analyses (where the UKBB diagnosed rather than the UKBB proxy results were used). In the forest plots, data are presented as odds-ratio with 95% confidence interval. P values are two-sided raw P values derived from a fixed-effect meta-analysis. OR: odds ratio, CI: confidence interval, EA: effect allele, EAF: effect allele frequency range across all studies, HetP: heterogeneity P value, HetISq: heterogeneity statistic.

a)

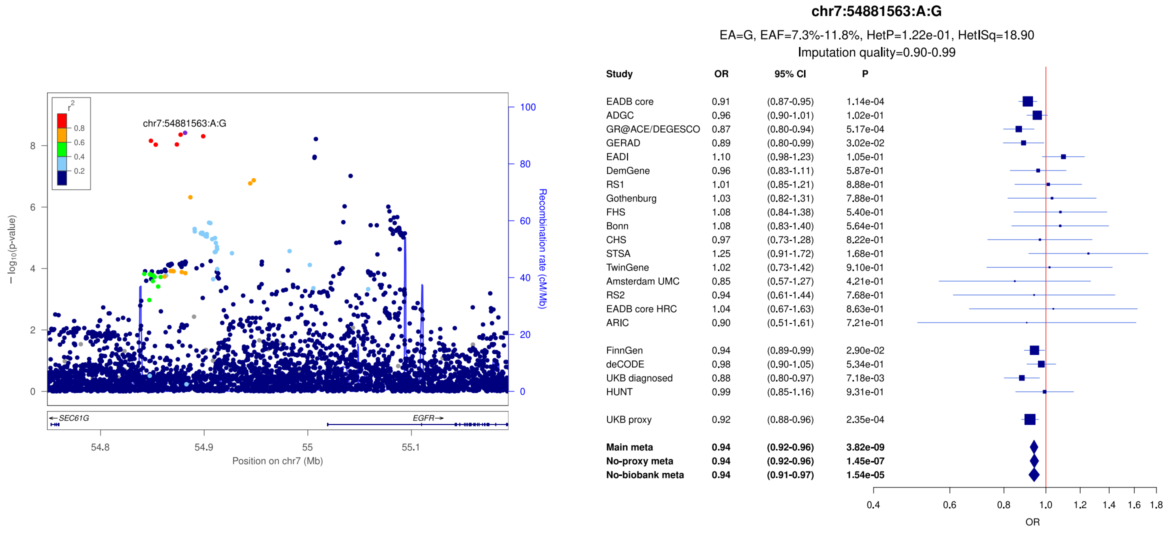


b)

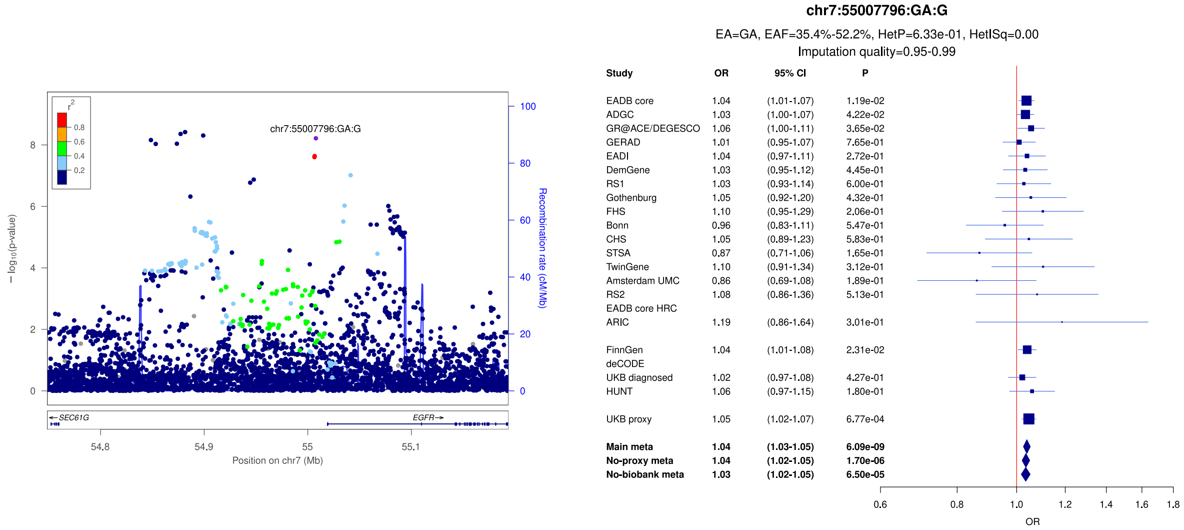


c)

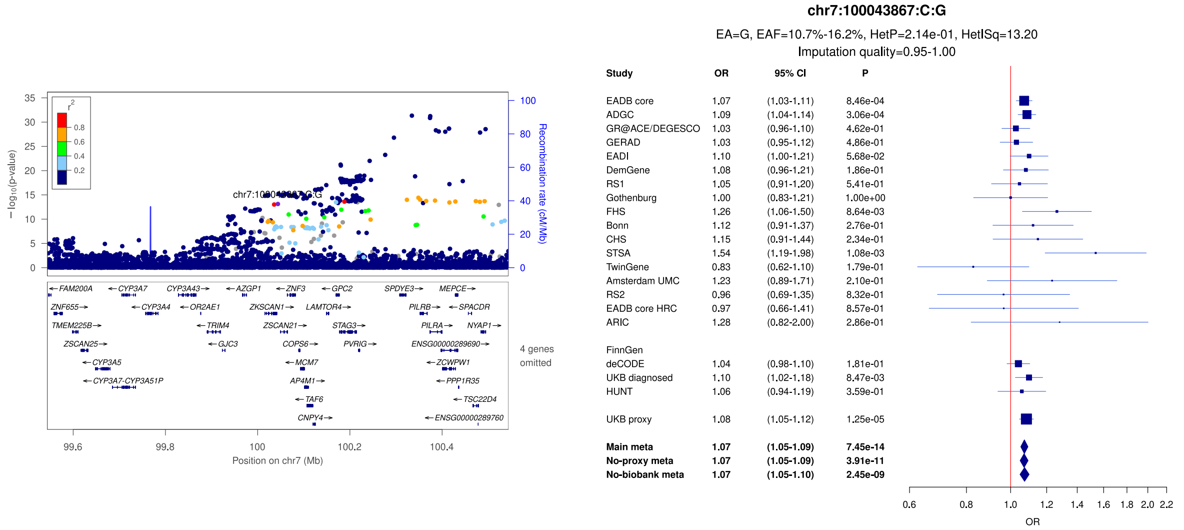

**Supplementary Figure 22:** LocusZoom and forest plots of the main analysis results for (a) *ZCWPW1/NYAP1* (lead signal) (b) *DOCK4* and (c) *EPHA1*. The forest plots also show the results of the no-proxy and no-biobank meta-analyses (where the UKBB diagnosed rather than the UKBB proxy results were used). In the forest plots, data are presented as odds-ratio with 95% confidence interval. P values are two-sided raw P values derived from a fixed-effect meta-analysis. OR: odds ratio, CI: confidence interval, EA: effect allele, EAF: effect allele frequency range across all studies, HetP: heterogeneity P value, HetISq: heterogeneity statistic.

a)

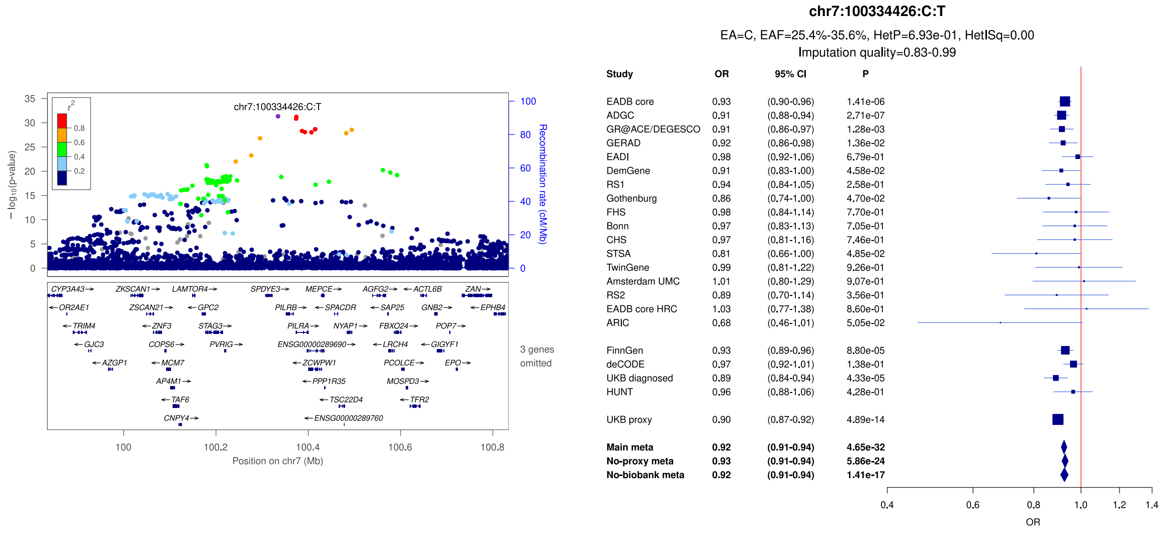


b)

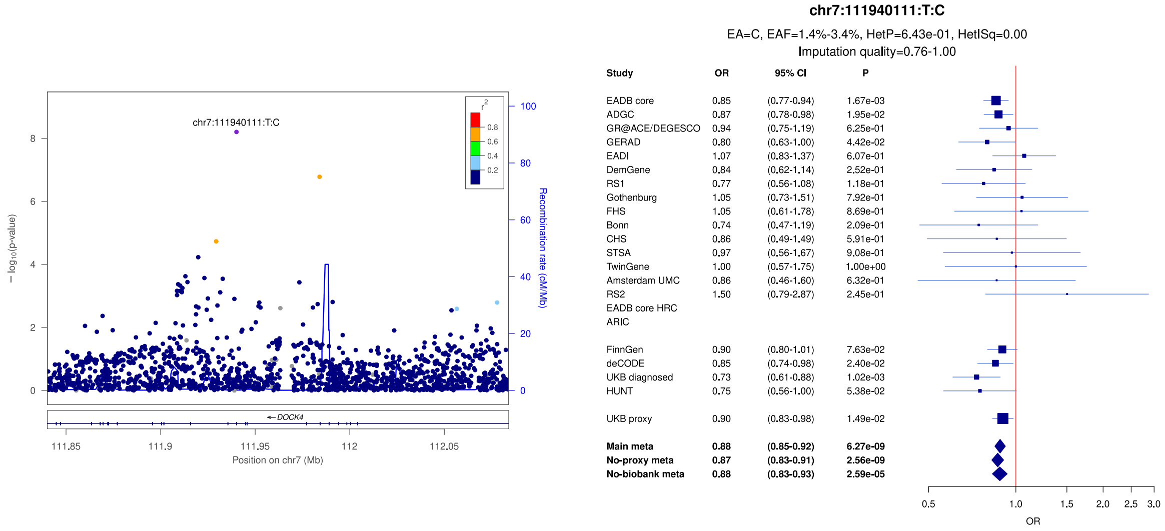


c)

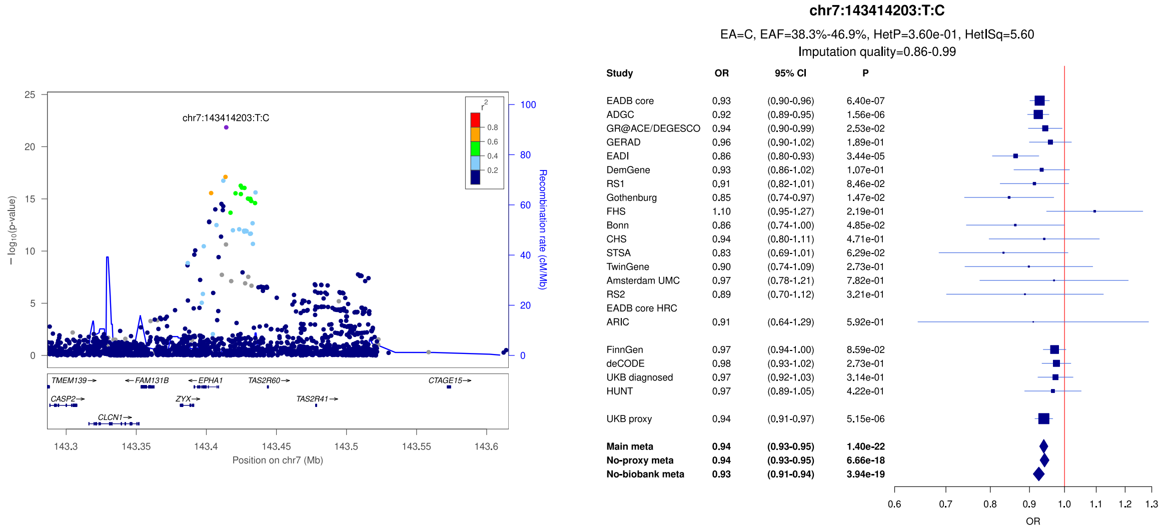

**Supplementary Figure 23:** LocusZoom and forest plots of the main analysis results for (a) *CTSB* (secondary signal) (b) *CTSB* (lead signal) and (c) *CLU/PTK2B* (secondary signal). The forest plots also show the results of the no-proxy and no-biobank meta-analyses (where the UKBB diagnosed rather than the UKBB proxy results were used). In the forest plots, data are presented as odds-ratio with 95% confidence interval. P values are two-sided raw P values derived from a fixed-effect meta-analysis. OR: odds ratio, CI: confidence interval, EA: effect allele, EAF: effect allele frequency range across all studies, HetP: heterogeneity P value, HetISq: heterogeneity statistic.

a)

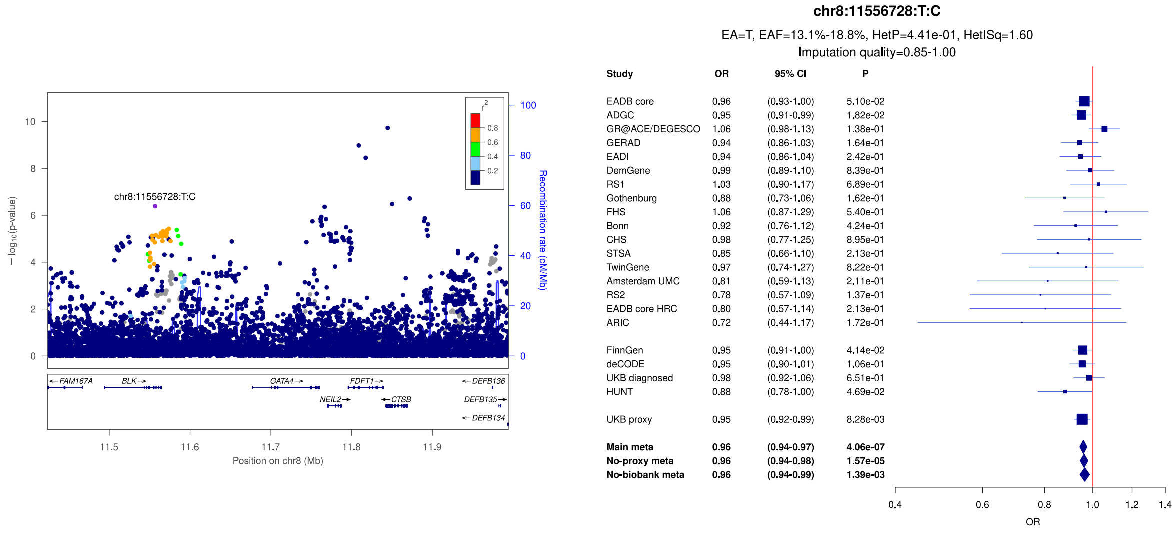


b)

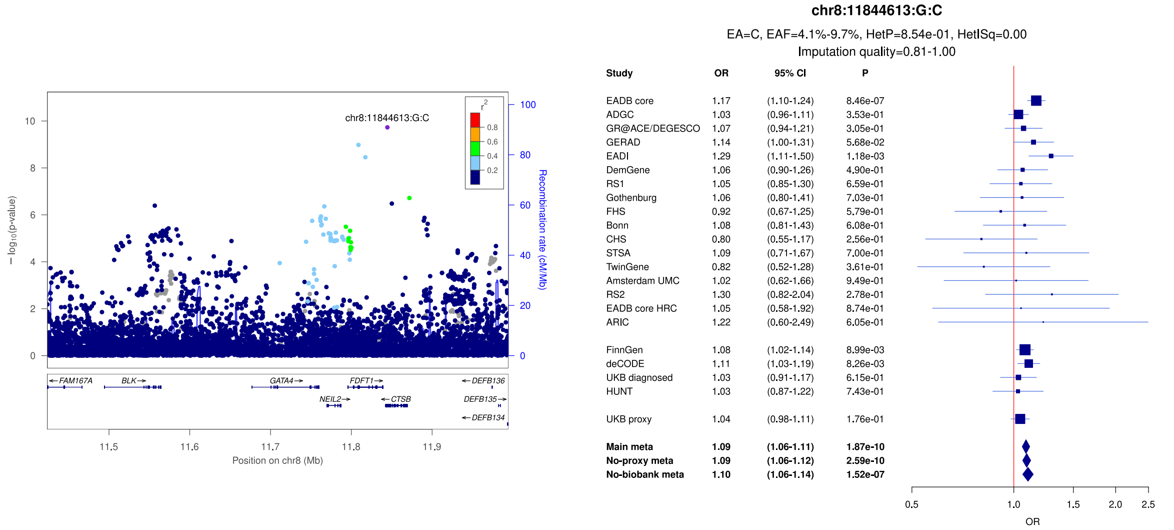


c)

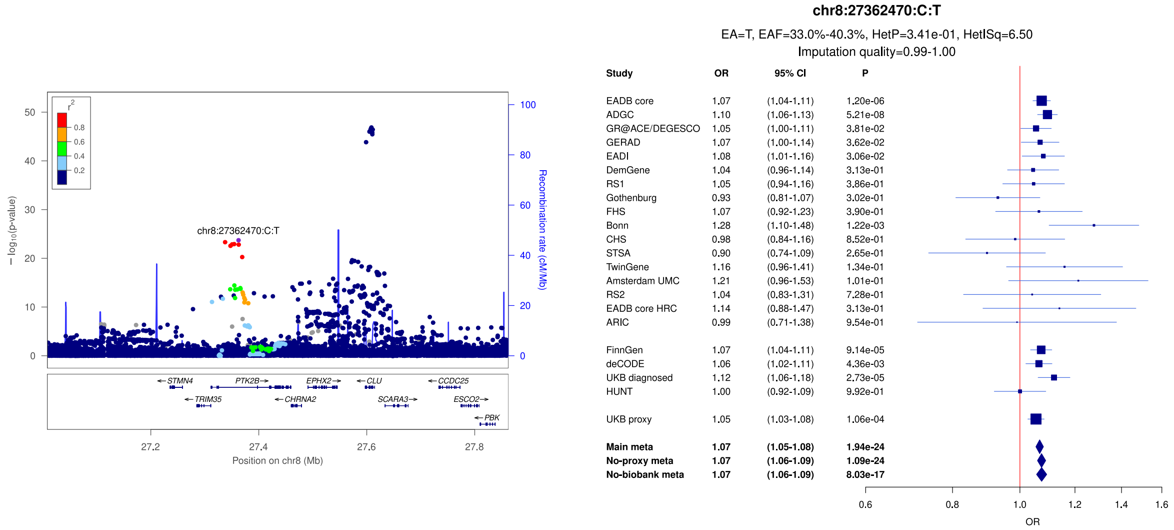

**Supplementary Figure 24:** LocusZoom and forest plots of the main analysis results for (a) *CLU/PTK2B* (lead signal) (b) *CLU/PTK2B* (secondary signal) and (c) *TRIB1* (lead signal). The forest plots also show the results of the no-proxy and no-biobank meta-analyses (where the UKBB diagnosed rather than the UKBB proxy results were used). In the forest plots, data are presented as odds-ratio with 95% confidence interval. P values are two-sided raw P values derived from a fixed-effect meta-analysis. OR: odds ratio, CI: confidence interval, EA: effect allele, EAF: effect allele frequency range across all studies, HetP: heterogeneity P value, HetISq: heterogeneity statistic.

a)

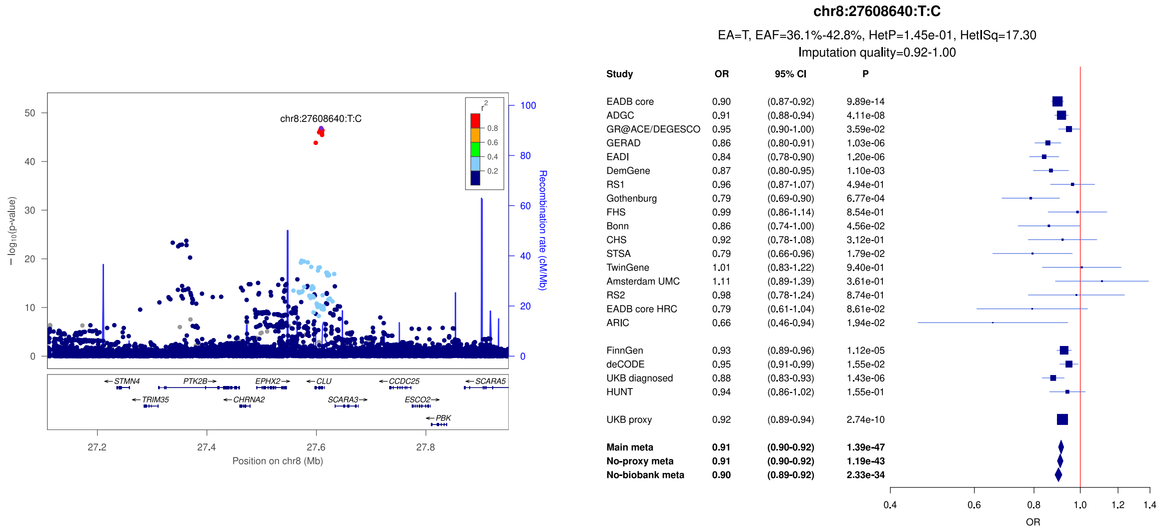


b)

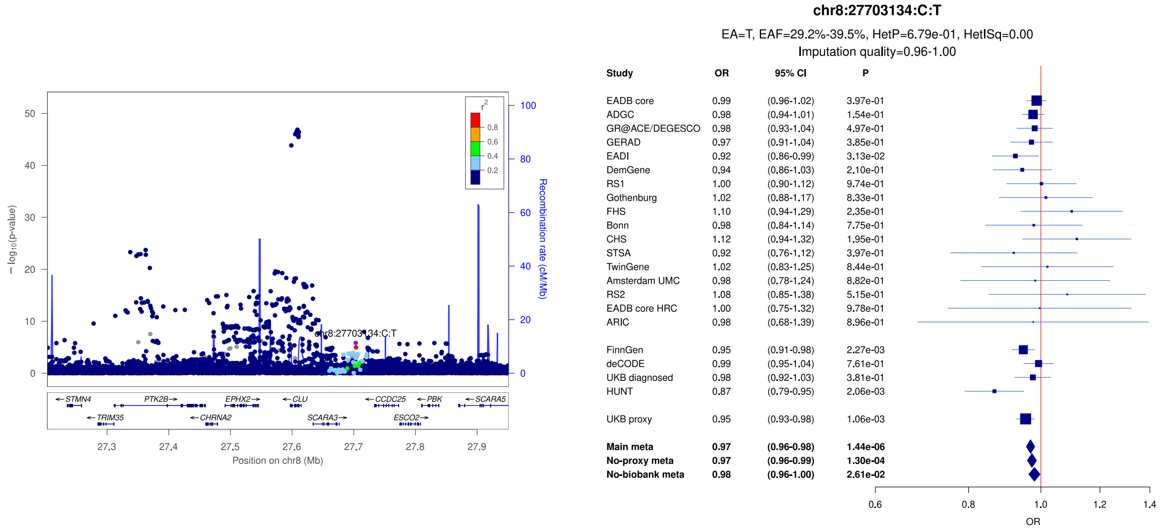


c)

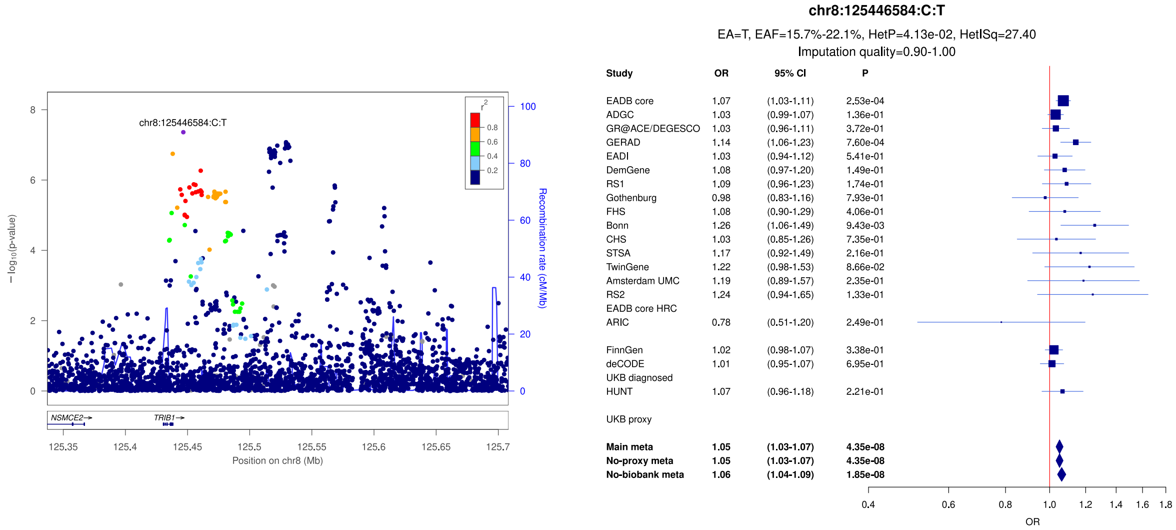

**Supplementary Figure 25:** LocusZoom and forest plots of the main analysis results for (a) *TRIB1* (secondary signal) (b) *SHARPIN* (secondary signal) and (c) *SHARPIN* (secondary signal). The forest plots also show the results of the no-proxy and no-biobank meta-analyses (where the UKBB diagnosed rather than the UKBB proxy results were used). In the forest plots, data are presented as odds-ratio with 95% confidence interval. P values are two-sided raw P values derived from a fixed-effect meta-analysis. OR: odds ratio, CI: confidence interval, EA: effect allele, EAF: effect allele frequency range across all studies, HetP: heterogeneity P value, HetISq: heterogeneity statistic.

a)

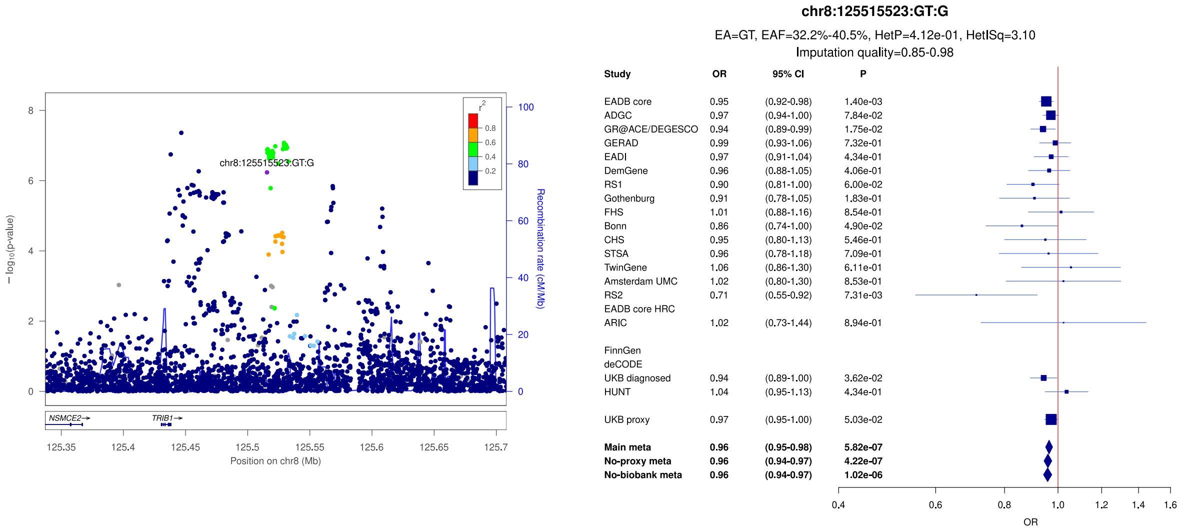


b)

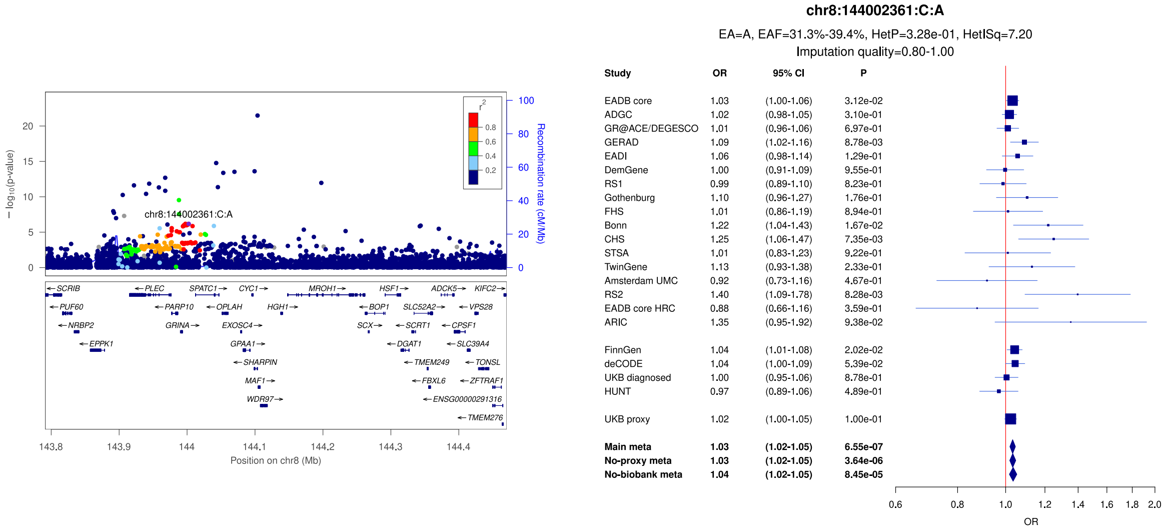


c)

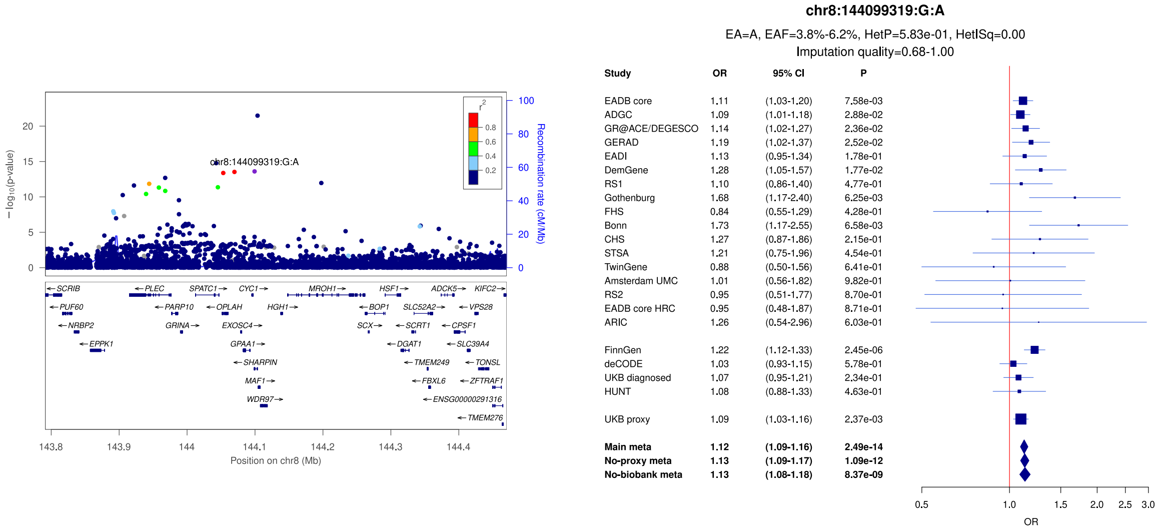

**Supplementary Figure 26:** LocusZoom and forest plots of the main analysis results for (a) *SHARPIN* (lead signal) (b) *ABCA1* (lead signal) and (c) *ECHDC3/USP6NL* (secondary signal). The forest plots also show the results of the no-proxy and no-biobank meta-analyses (where the UKBB diagnosed rather than the UKBB proxy results were used). In the forest plots, data are presented as odds-ratio with 95% confidence interval. P values are two-sided raw P values derived from a fixed-effect meta-analysis. OR: odds ratio, CI: confidence interval, EA: effect allele, EAF: effect allele frequency range across all studies, HetP: heterogeneity P value, HetISq: heterogeneity statistic.

a)

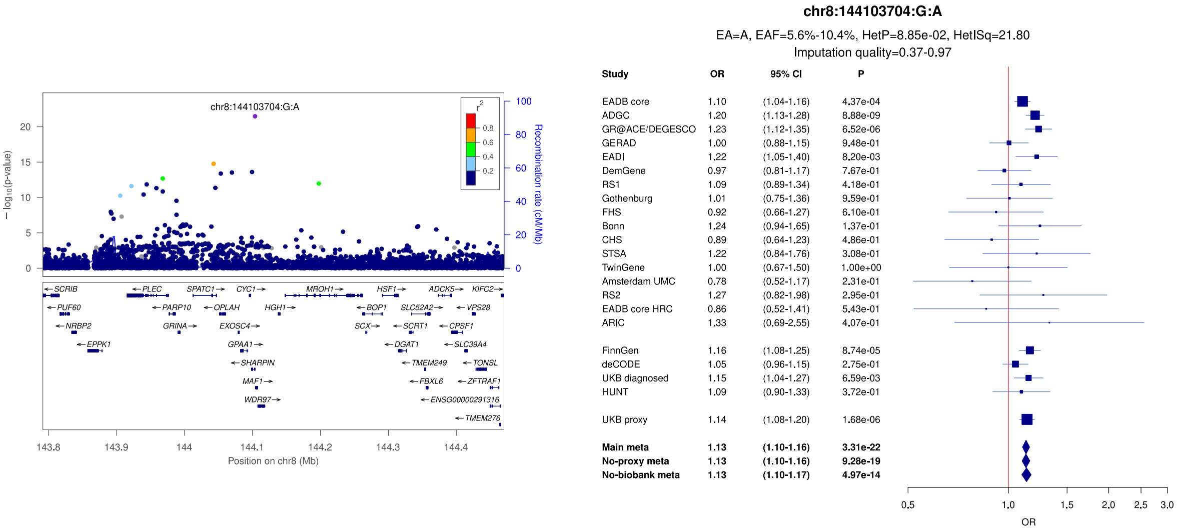


b)

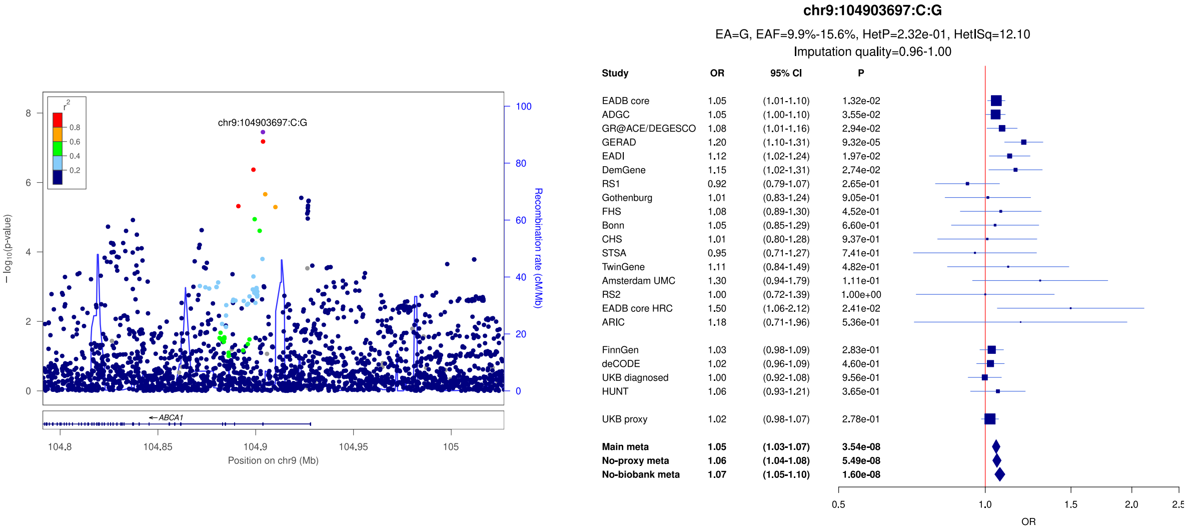


c)

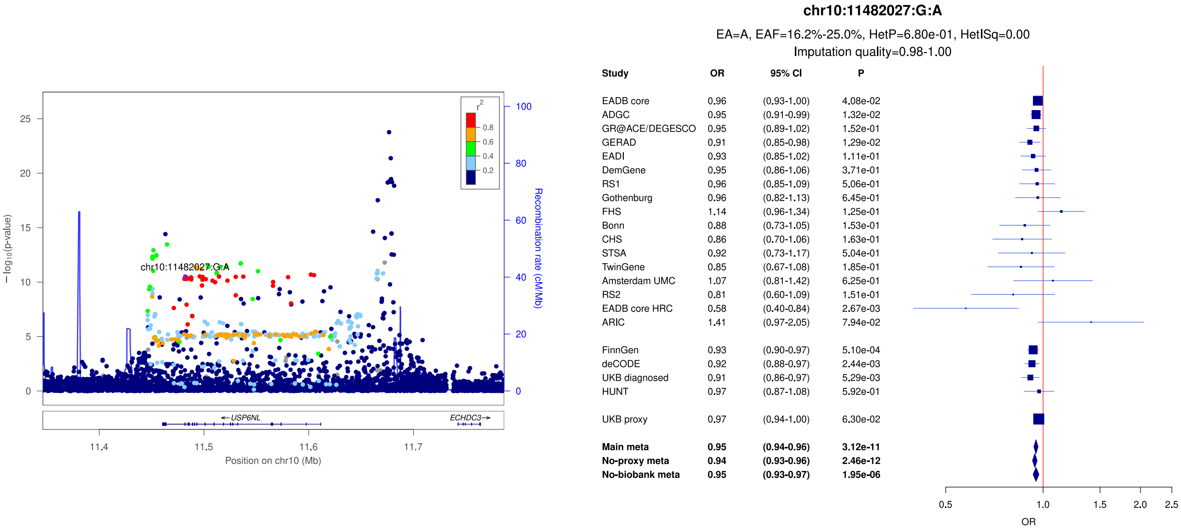

**Supplementary Figure 27:** LocusZoom and forest plots of the main analysis results for (a) *ECHDC3/USP6NL* (lead signal) (b) *IPMK* and (c) *ANK3*. The forest plots also show the results of the no-proxy and no-biobank meta-analyses (where the UKBB diagnosed rather than the UKBB proxy results were used). In the forest plots, data are presented as odds-ratio with 95% confidence interval. P values are two-sided raw P values derived from a fixed-effect meta-analysis. OR: odds ratio, CI: confidence interval, EA: effect allele, EAF: effect allele frequency range across all studies, HetP: heterogeneity P value, HetISq: heterogeneity statistic.

a)

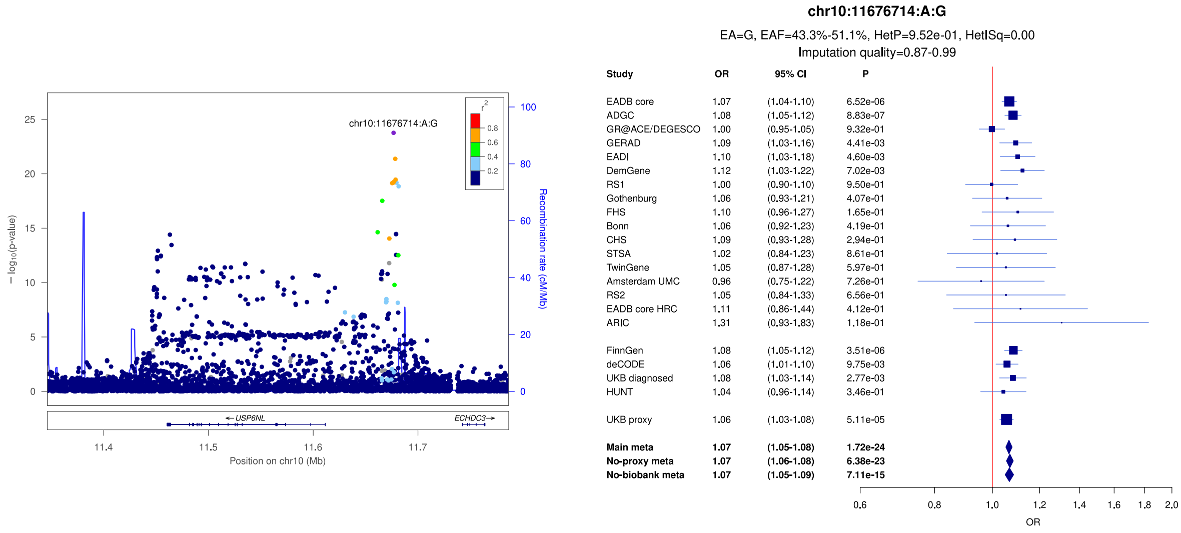


b)

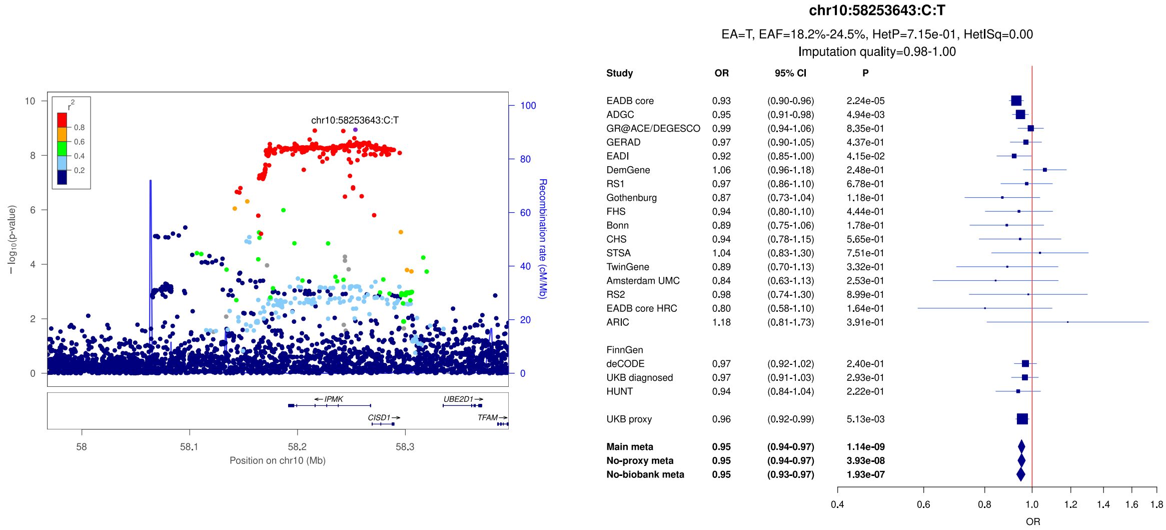


c)

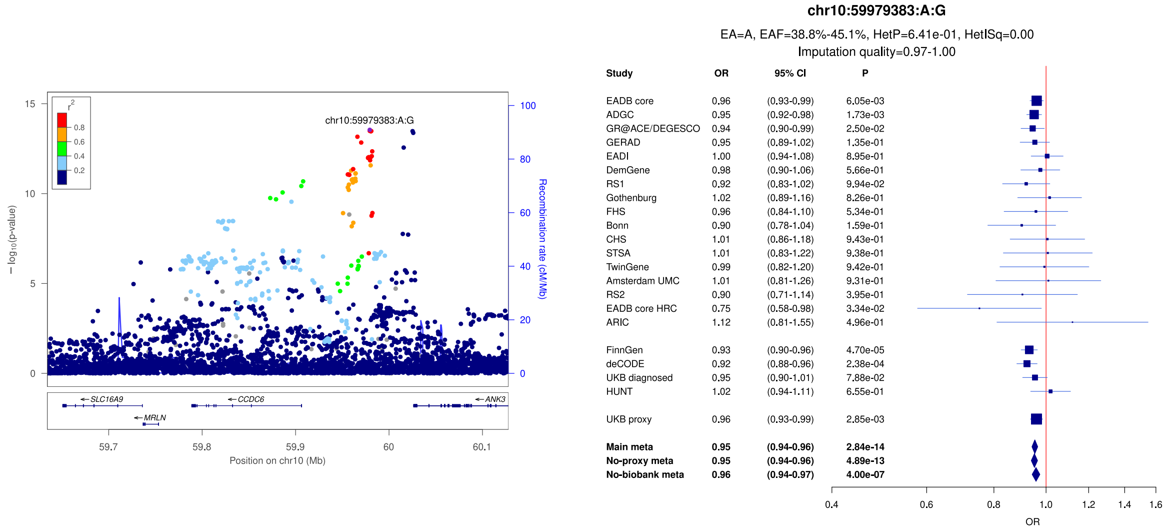

**Supplementary Figure 28:** LocusZoom and forest plots of the main analysis results for (a) *TSPAN14* (b) *BLNK* (secondary signal) and (c) *BLNK* (lead signal). The forest plots also show the results of the no-proxy and no-biobank meta-analyses (where the UKBB diagnosed rather than the UKBB proxy results were used). In the forest plots, data are presented as odds-ratio with 95% confidence interval. P values are two-sided raw P values derived from a fixed-effect meta-analysis. OR: odds ratio, CI: confidence interval, EA: effect allele, EAF: effect allele frequency range across all studies, HetP: heterogeneity P value, HetISq: heterogeneity statistic.

a)

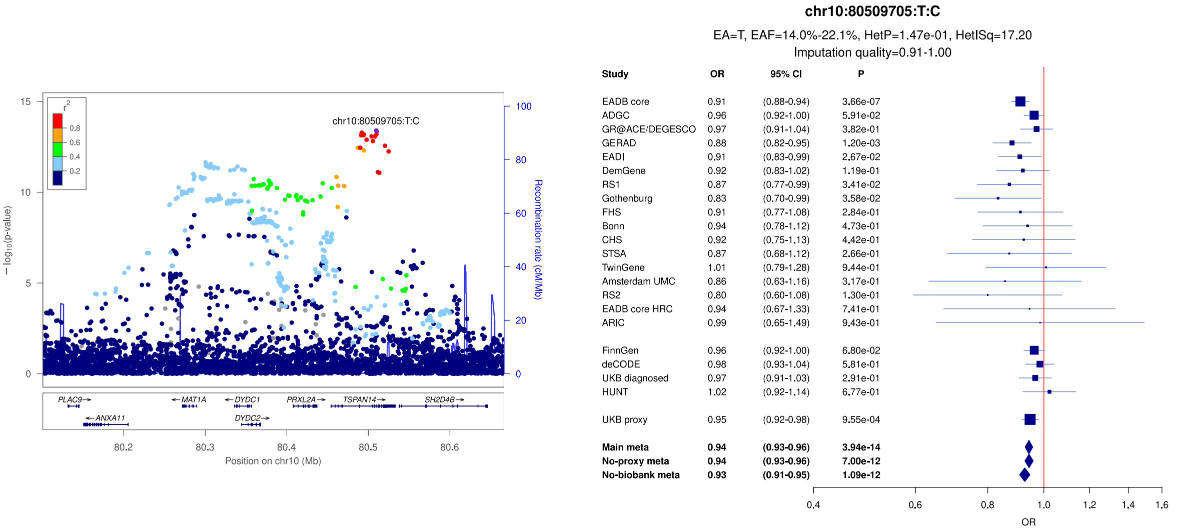


b)

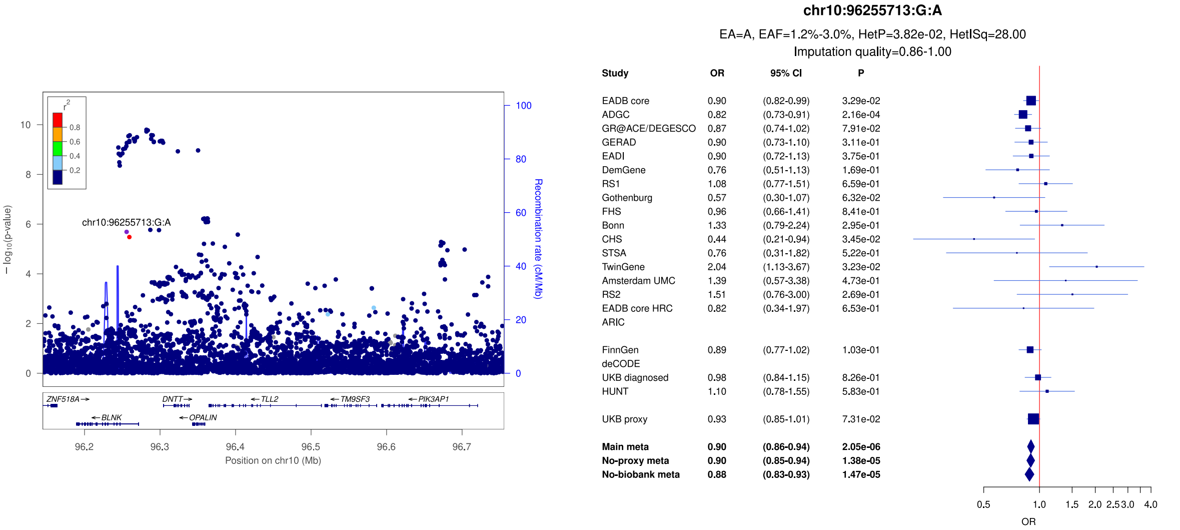


c)

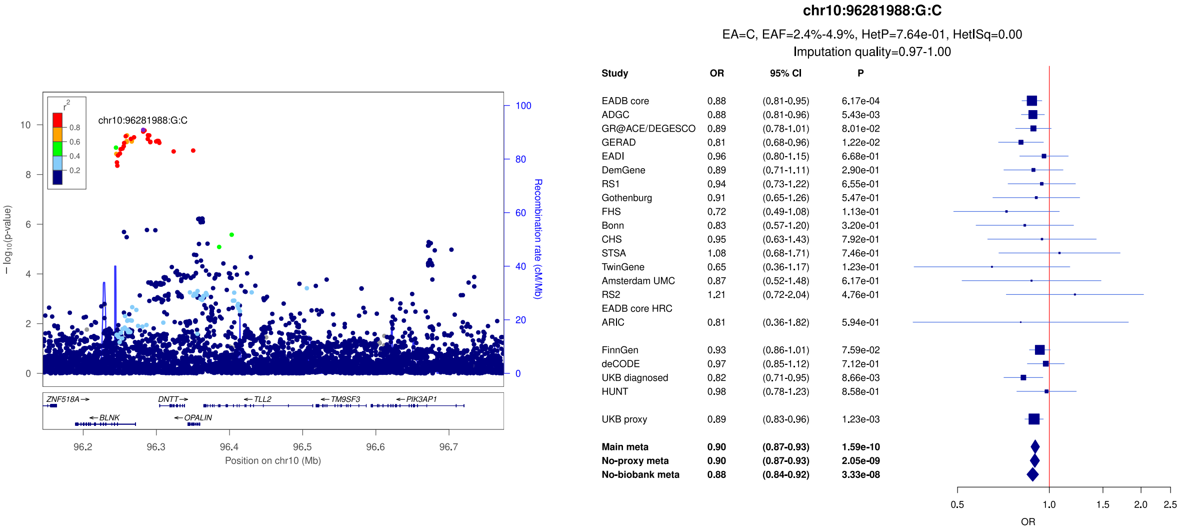

**Supplementary Figure 29:** LocusZoom and forest plots of the main analysis results for (a) *PLEKHA1* (b) *CELF1/SPI1* and (c) *MS4A* (lead signal). The forest plots also show the results of the no-proxy and no-biobank meta-analyses (where the UKBB diagnosed rather than the UKBB proxy results were used). In the forest plots, data are presented as odds-ratio with 95% confidence interval. P values are two-sided raw P values derived from a fixed-effect meta-analysis. OR: odds ratio, CI: confidence interval, EA: effect allele, EAF: effect allele frequency range across all studies, HetP: heterogeneity P value, HetISq: heterogeneity statistic.

a)

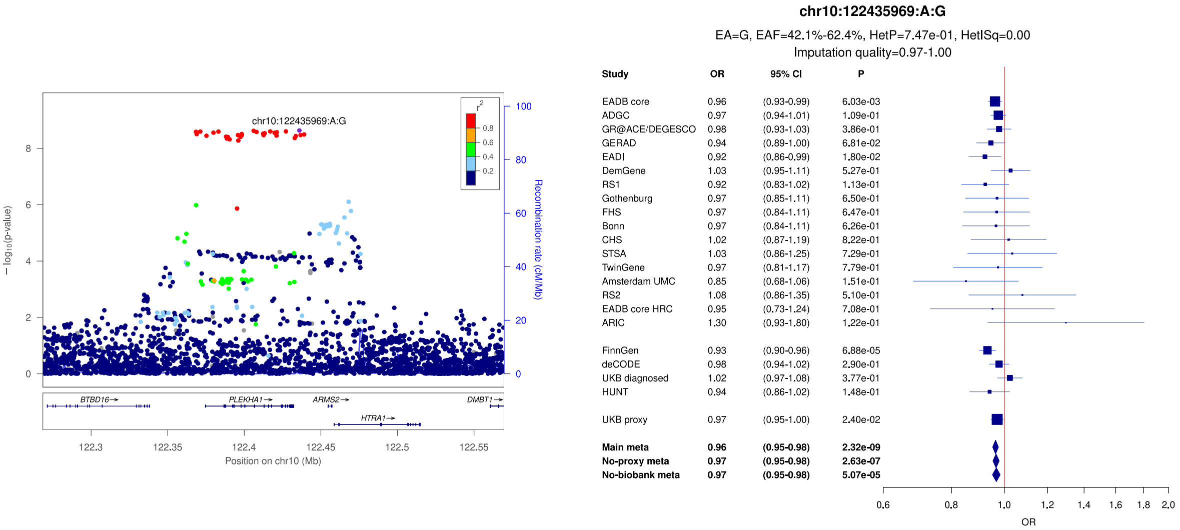


b)

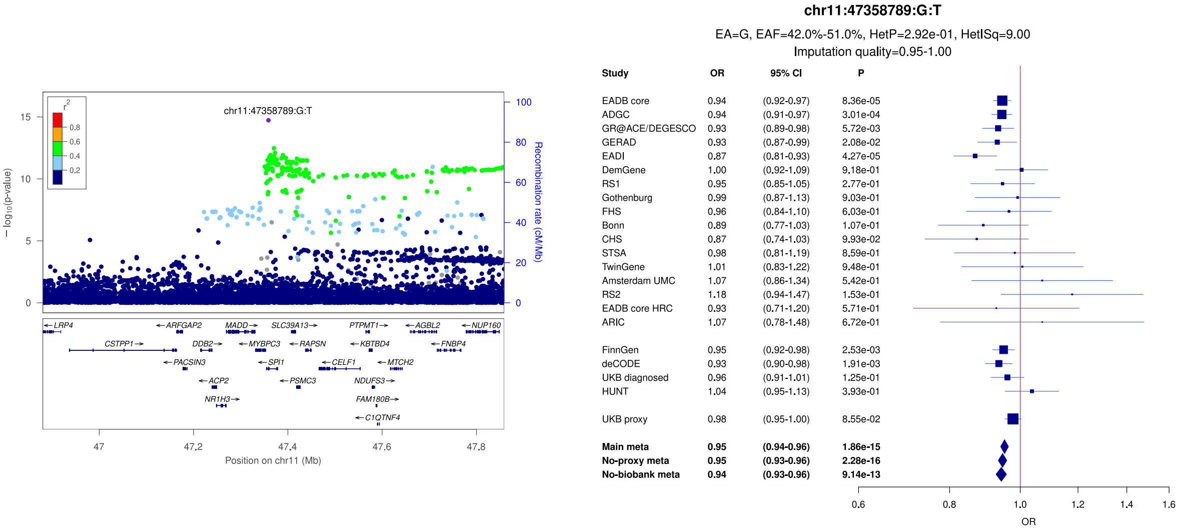


c)

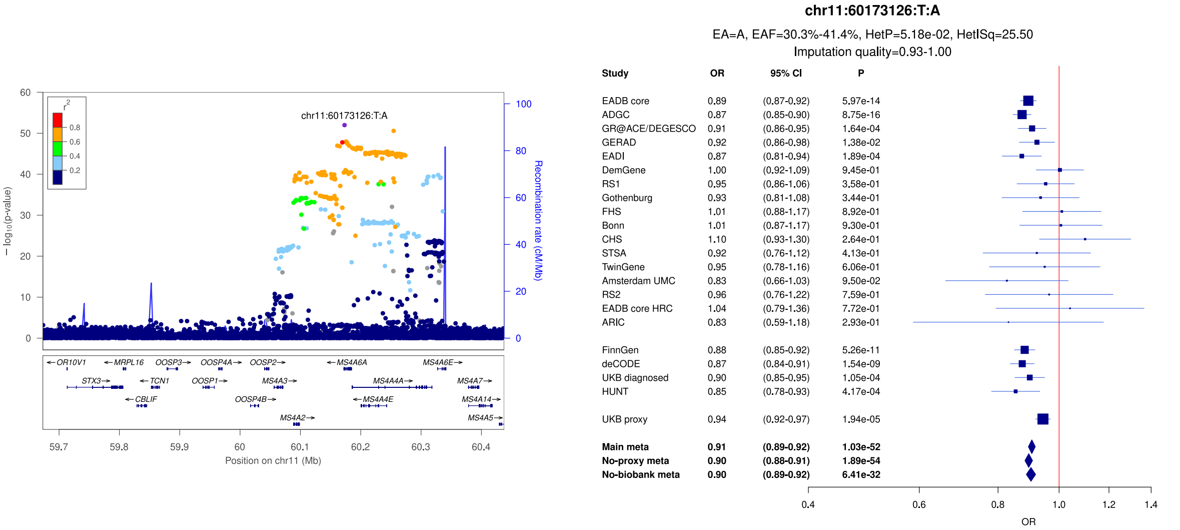

**Supplementary Figure 30:** LocusZoom and forest plots of the main analysis results for (a) *MS4A* (secondary signal) (b) *PICALM* (secondary signal) and (c) *PICALM* (lead signal). The forest plots also show the results of the no-proxy and no-biobank meta-analyses (where the UKBB diagnosed rather than the UKBB proxy results were used). In the forest plots, data are presented as odds-ratio with 95% confidence interval. P values are two-sided raw P values derived from a fixed-effect meta-analysis. OR: odds ratio, CI: confidence interval, EA: effect allele, EAF: effect allele frequency range across all studies, HetP: heterogeneity P value, HetISq: heterogeneity statistic.

a)

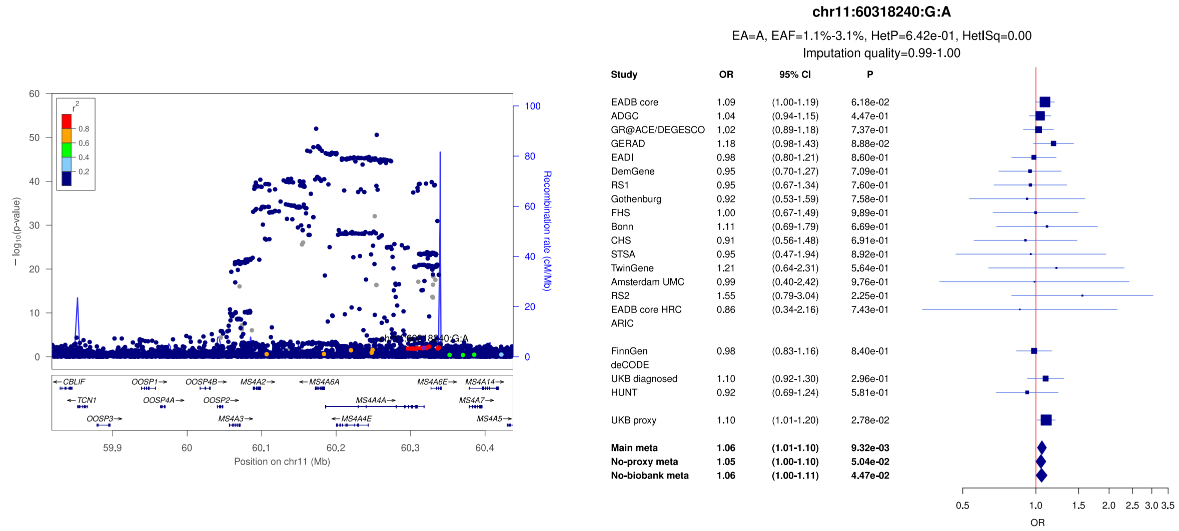


b)

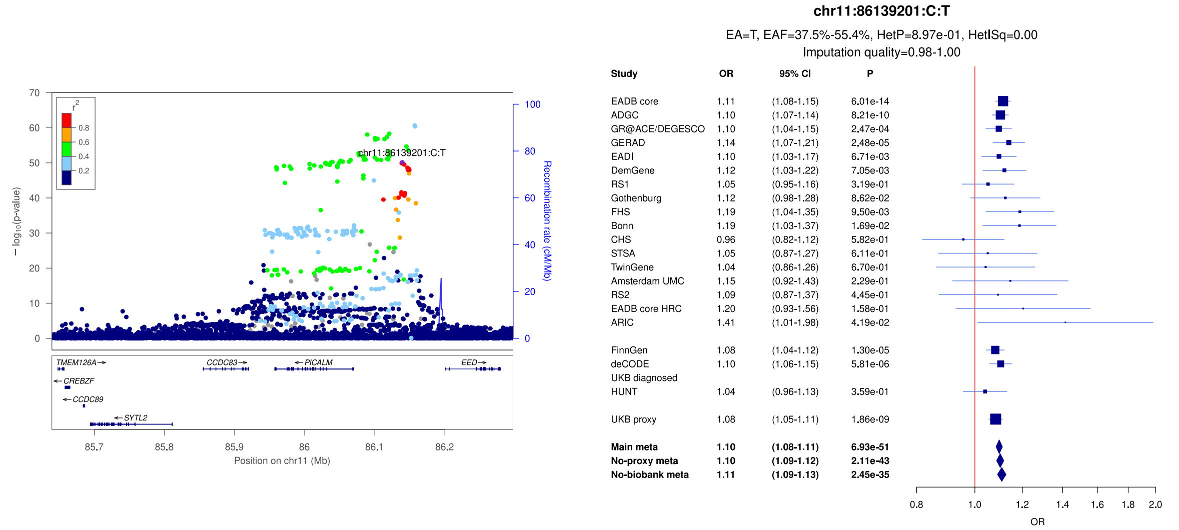


c)

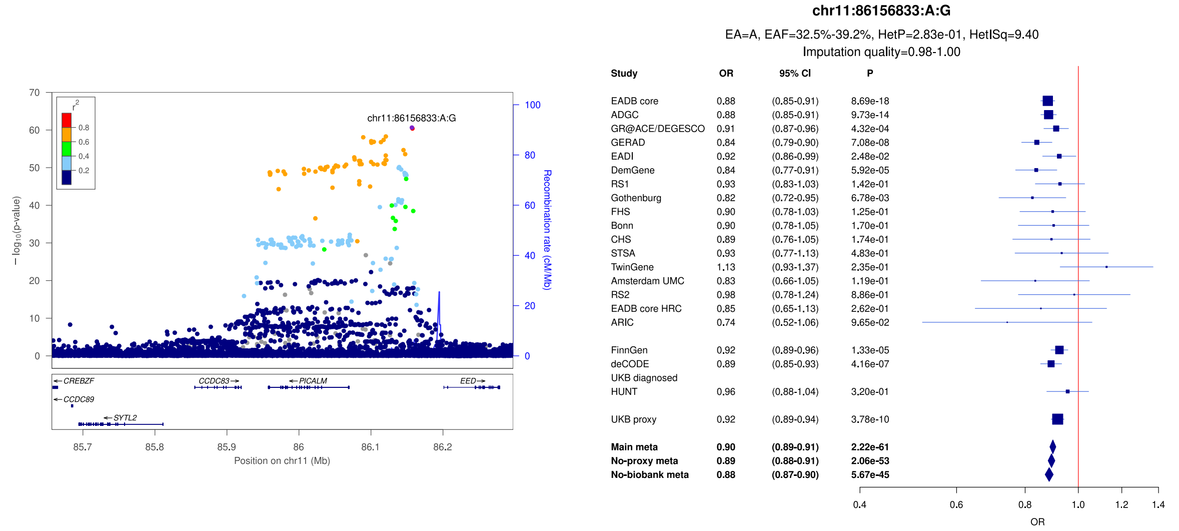

**Supplementary Figure 31:** LocusZoom and forest plots of the main analysis results for (a) *SORL1* (secondary signal) (b) *SORL1* (lead signal) and (c) *TPCN1/RITA1/IQCD*. The forest plots also show the results of the no-proxy and no-biobank meta-analyses (where the UKBB diagnosed rather than the UKBB proxy results were used). In the forest plots, data are presented as odds-ratio with 95% confidence interval. P values are two-sided raw P values derived from a fixed-effect meta-analysis. OR: odds ratio, CI: confidence interval, EA: effect allele, EAF: effect allele frequency range across all studies, HetP: heterogeneity P value, HetISq: heterogeneity statistic.

a)

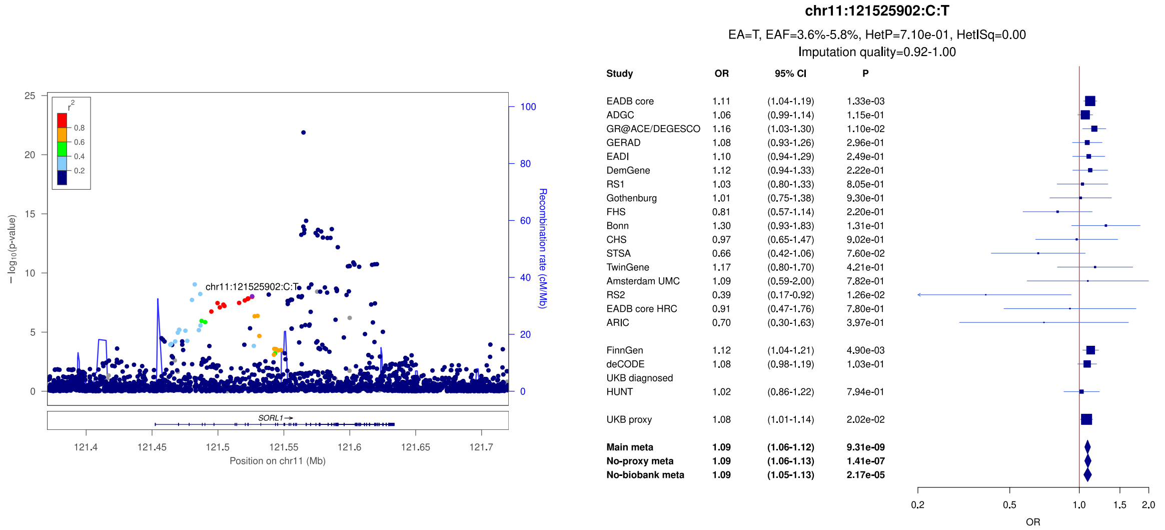


b)

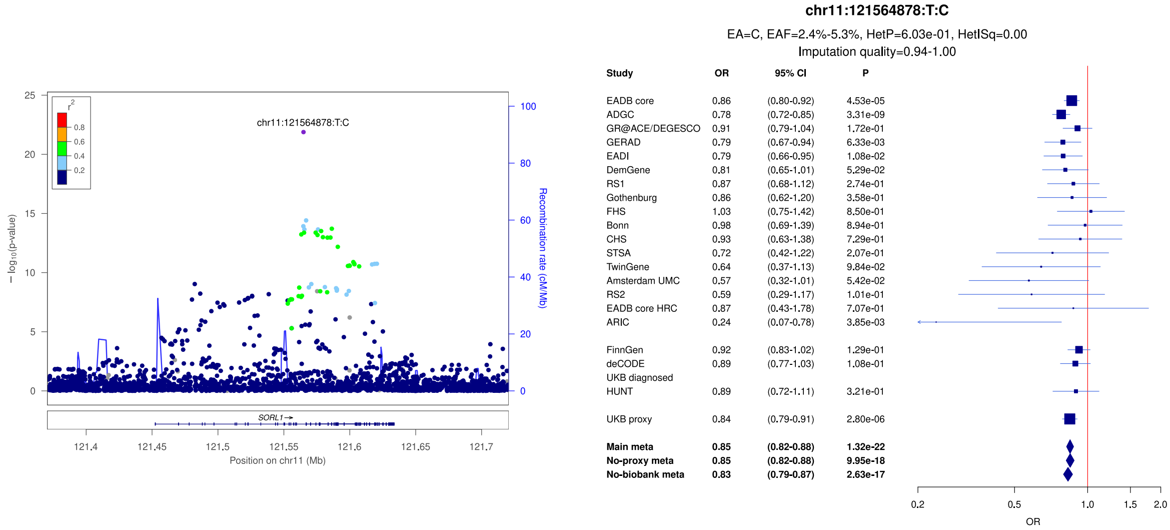


c)

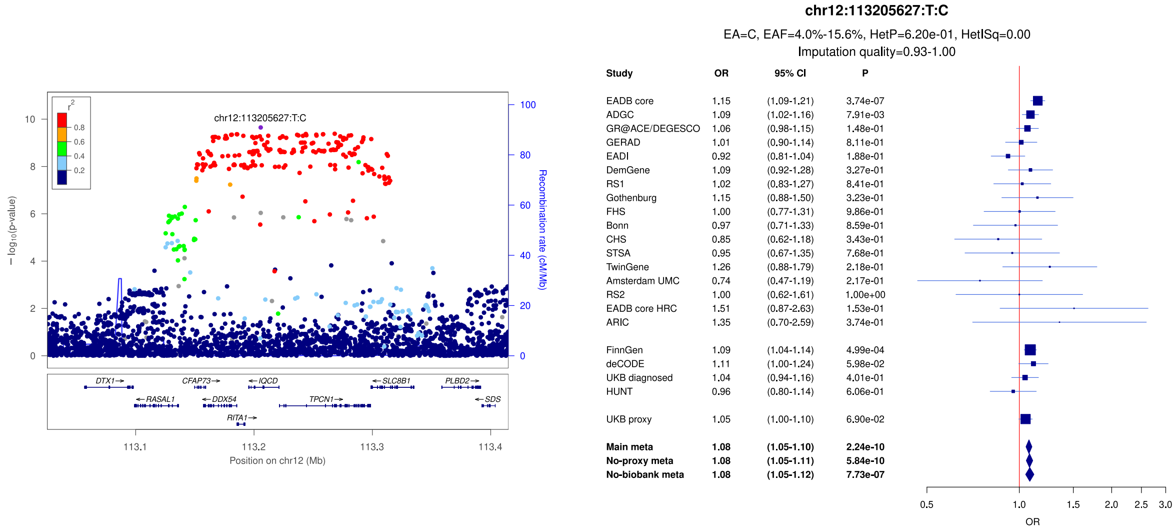

**Supplementary Figure 32:** LocusZoom and forest plots of the main analysis results for (a) *FERMT2* (b) *SLC24A4/RIN3* (secondary signal) and (c) *SLC24A4/RIN3* (lead signal). The forest plots also show the results of the no-proxy and no-biobank meta-analyses (where the UKBB diagnosed rather than the UKBB proxy results were used). In the forest plots, data are presented as odds-ratio with 95% confidence interval. P values are two-sided raw P values derived from a fixed-effect meta-analysis. OR: odds ratio, CI: confidence interval, EA: effect allele, EAF: effect allele frequency range across all studies, HetP: heterogeneity P value, HetISq: heterogeneity statistic.

a)

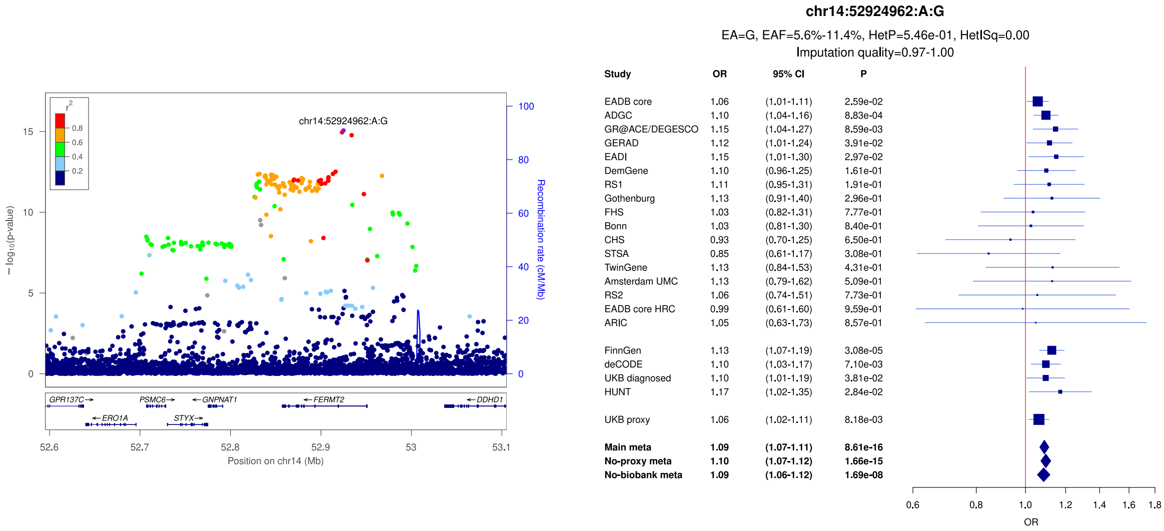


b)

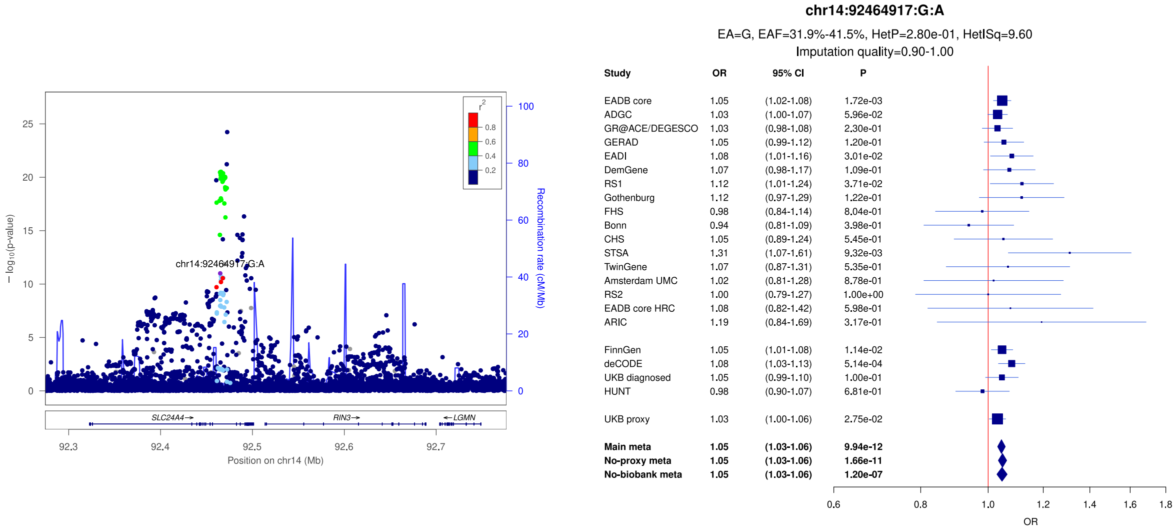


c)

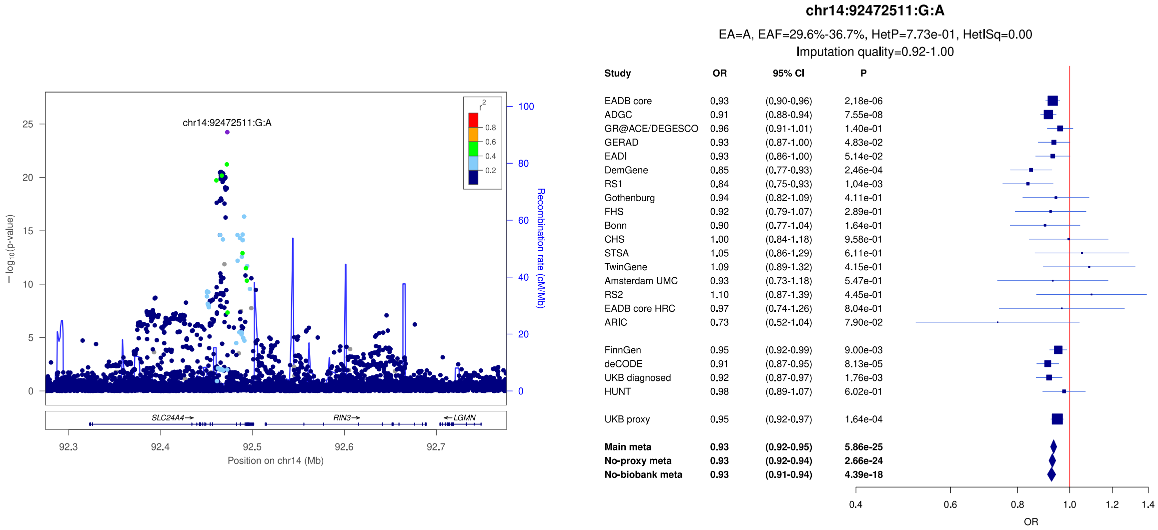

**Supplementary Figure 33:** LocusZoom and forest plots of the main analysis results for secondary signals of *SLC24A4/RIN3* (a, b, c). The forest plots also show the results of the no-proxy and no-biobank meta-analyses (where the UKBB diagnosed rather than the UKBB proxy results were used). In the forest plots, data are presented as odds-ratio with 95% confidence interval. P values are two-sided raw P values derived from a fixed-effect meta-analysis. OR: odds ratio, CI: confidence interval, EA: effect allele, EAF: effect allele frequency range across all studies, HetP: heterogeneity P value, HetISq: heterogeneity statistic.

a)

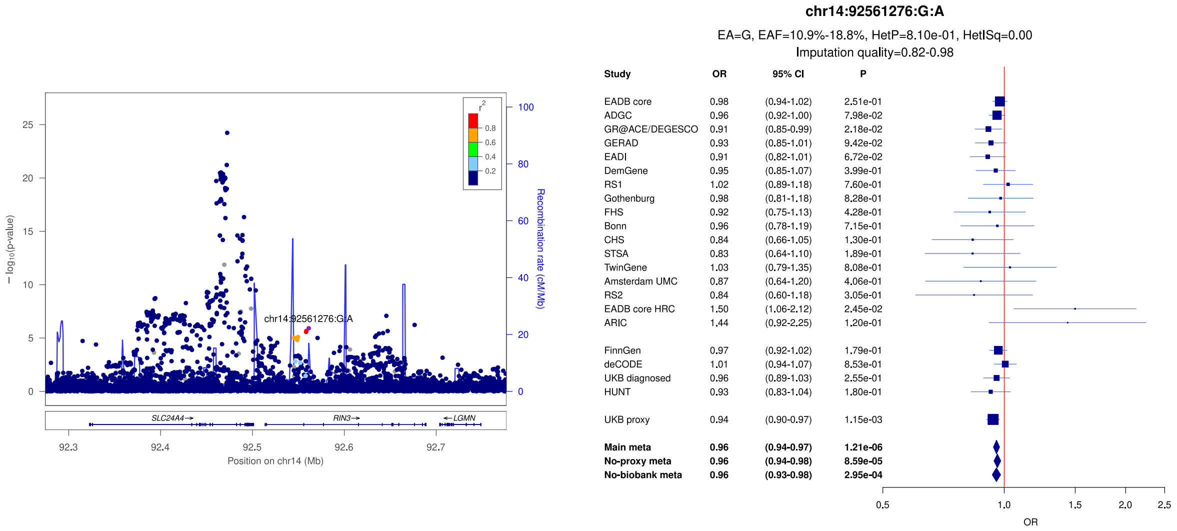


b)

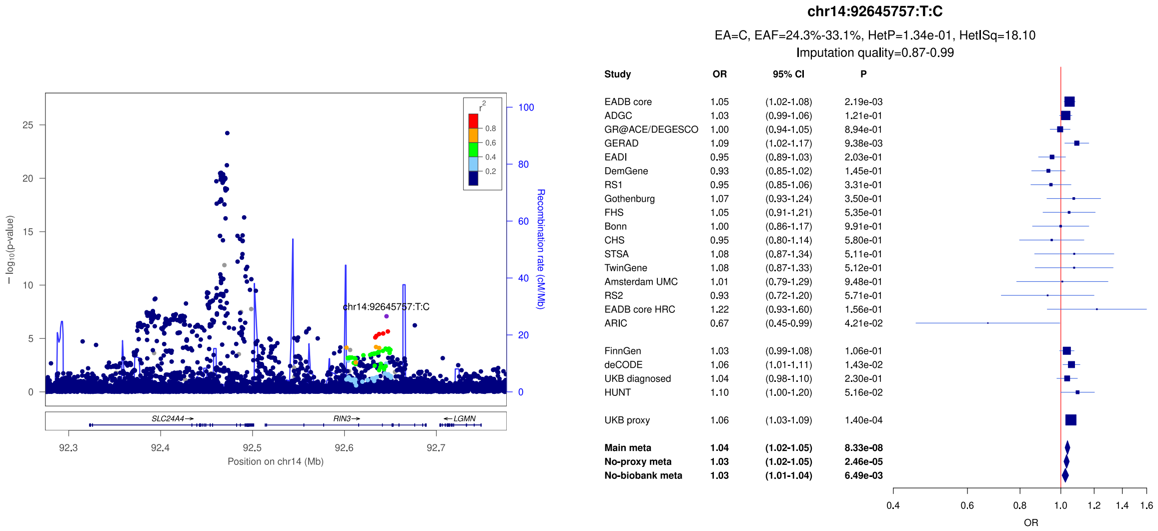


c)

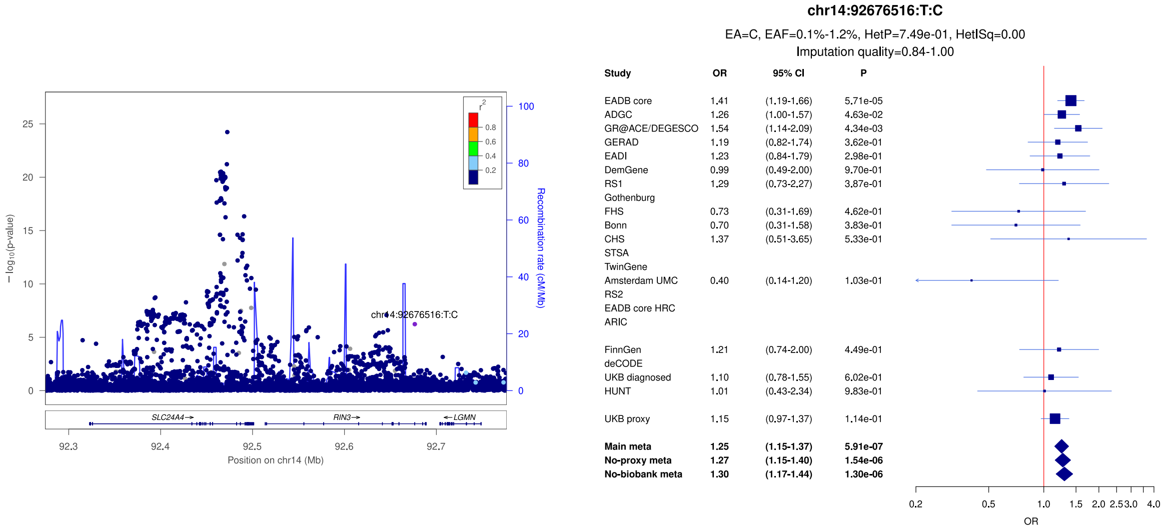

**Supplementary Figure 34:** LocusZoom and forest plots of the main analysis results for (a) *IGH* gene cluster (lead signal) (b) *IGH* gene cluster (lead signal) and (c) *ATP8B4* (lead signal). The forest plots also show the results of the no-proxy and no-biobank meta-analyses (where the UKBB diagnosed rather than the UKBB proxy results were used). In the forest plots, data are presented as odds-ratio with 95% confidence interval. P values are two-sided raw P values derived from a fixed-effect meta-analysis. OR: odds ratio, CI: confidence interval, EA: effect allele, EAF: effect allele frequency range across all studies, HetP: heterogeneity P value, HetISq: heterogeneity statistic.

a)

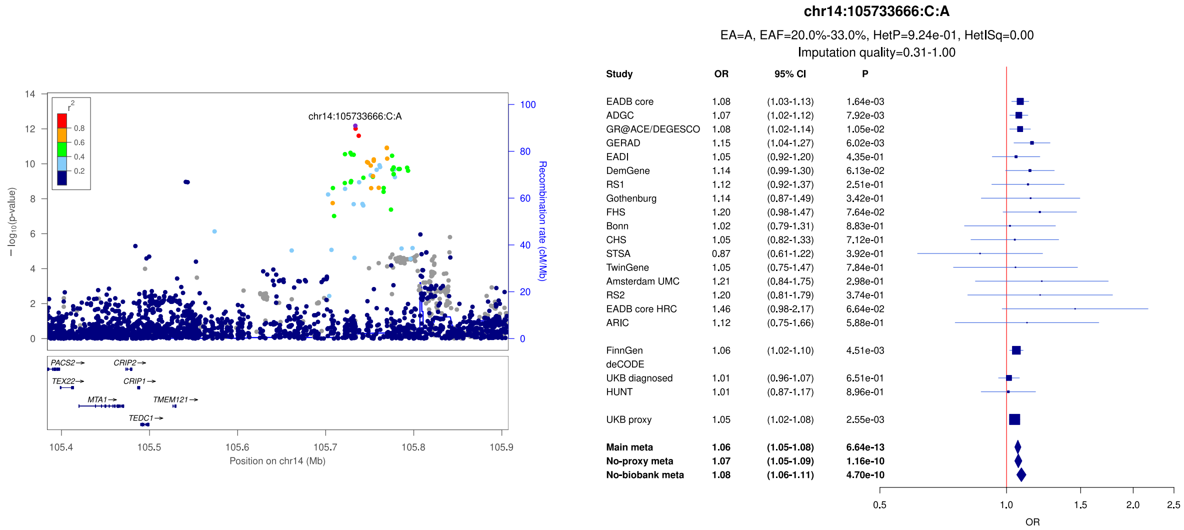


b)

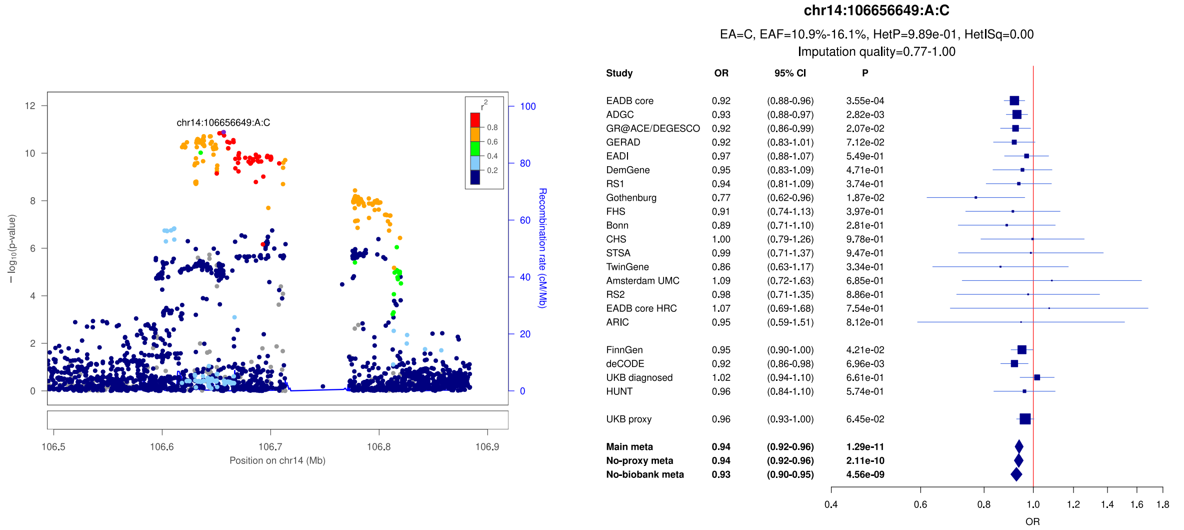


c)

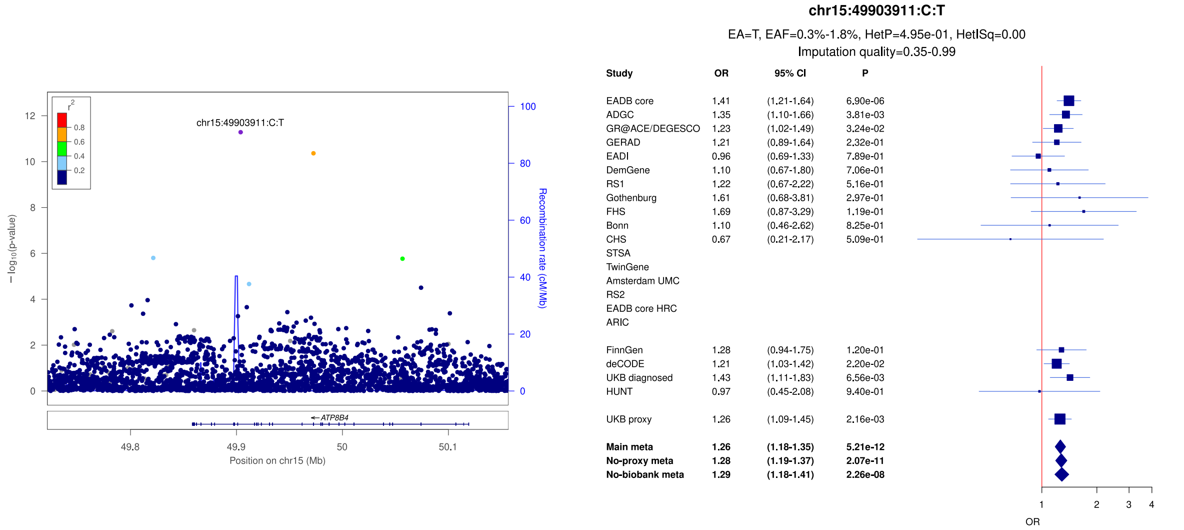

**Supplementary Figure 35:** LocusZoom and forest plots of the main analysis results for (a) *SPPL2A/USP8/USP50* (secondary signal) (b) *SPPL2A/USP8/USP50* (lead signal) and (c) *ADAM10* (secondary signal). The forest plots also show the results of the no-proxy and no-biobank meta-analyses (where the UKBB diagnosed rather than the UKBB proxy results were used). In the forest plots, data are presented as odds-ratio with 95% confidence interval. P values are two-sided raw P values derived from a fixed-effect meta-analysis. OR: odds ratio, CI: confidence interval, EA: effect allele, EAF: effect allele frequency range across all studies, HetP: heterogeneity P value, HetISq: heterogeneity statistic.

a)

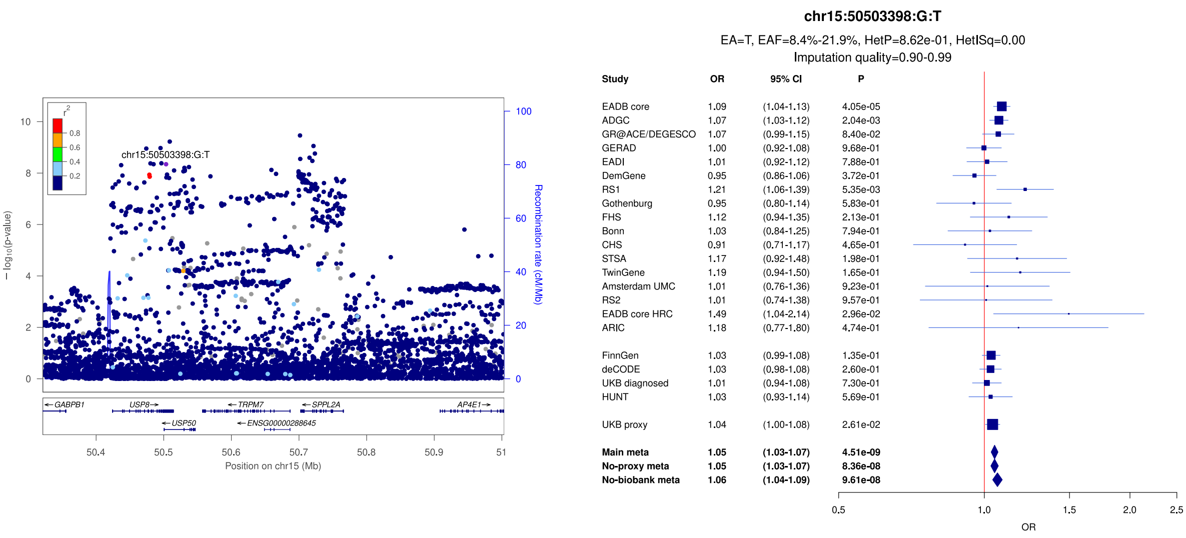


b)

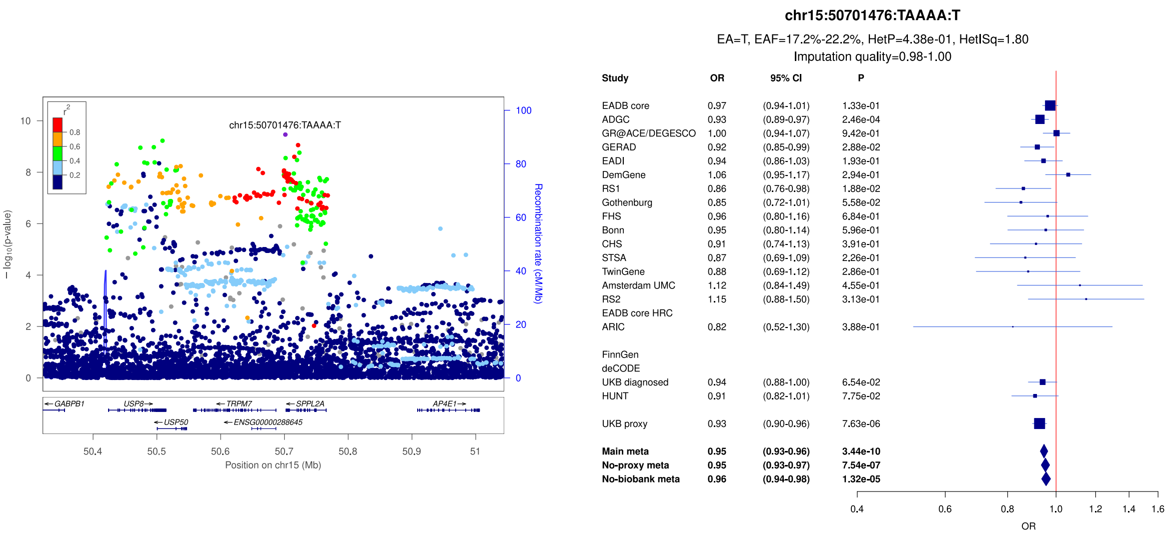


c)

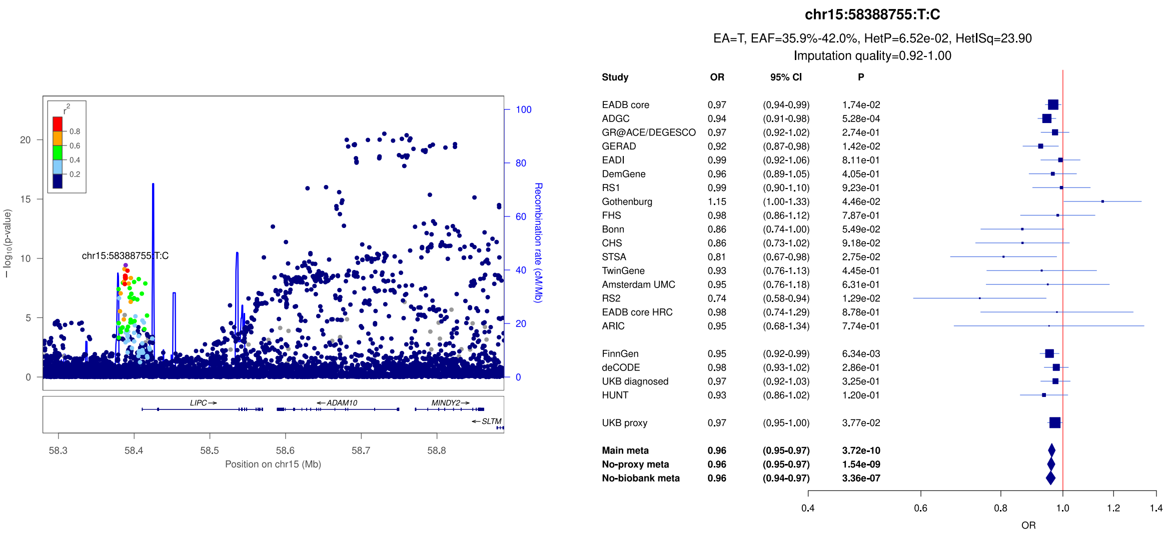

**Supplementary Figure 36:** LocusZoom and forest plots of the main analysis results for (a) *ADAM10* (secondary signal) (b) *ADAM10* (lead signal) and (c) *APH1B*. The forest plots also show the results of the no-proxy and no-biobank meta-analyses (where the UKBB diagnosed rather than the UKBB proxy results were used). In the forest plots, data are presented as odds-ratio with 95% confidence interval. P values are two-sided raw P values derived from a fixed-effect meta-analysis. OR: odds ratio, CI: confidence interval, EA: effect allele, EAF: effect allele frequency range across all studies, HetP: heterogeneity P value, HetISq: heterogeneity statistic.

a)

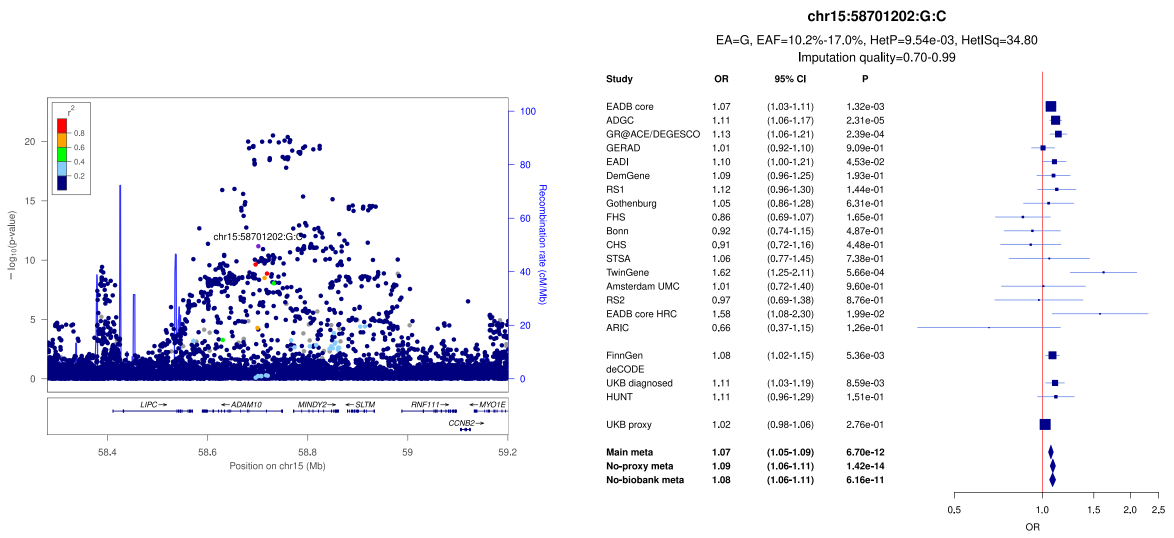


b)

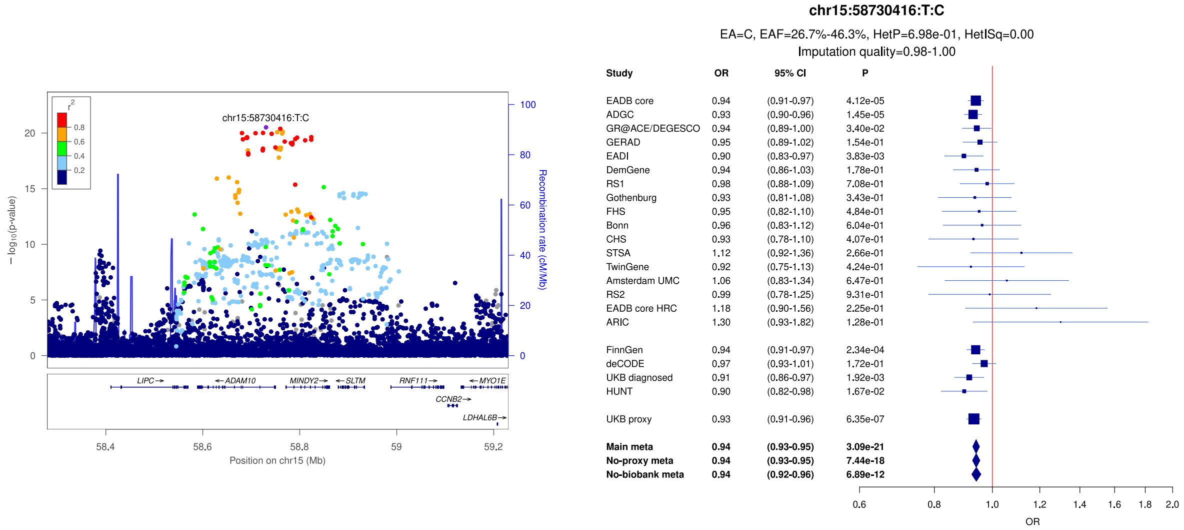


c)

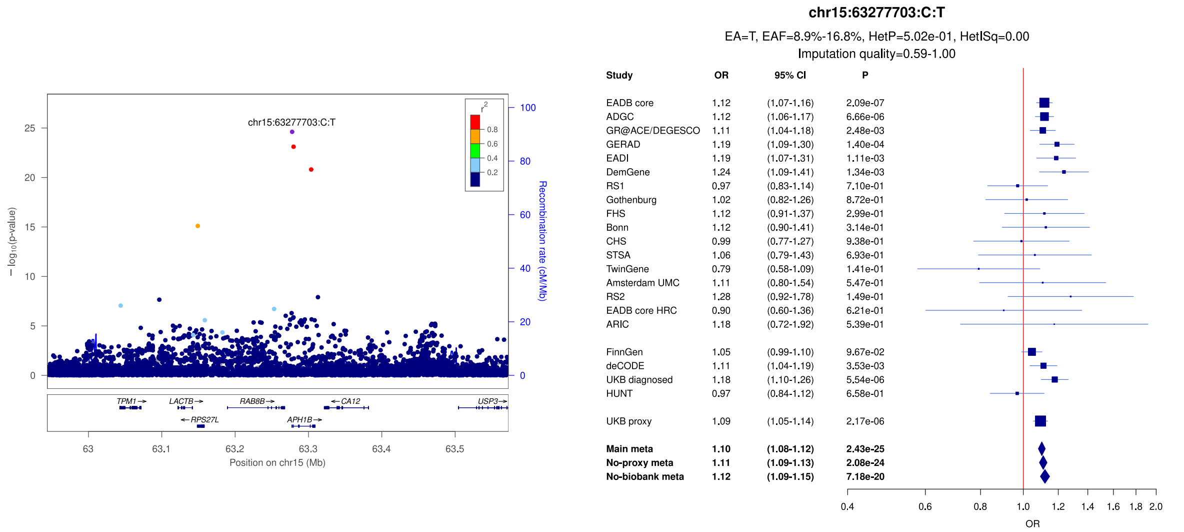

**Supplementary Figure 37:** LocusZoom and forest plots of the main analysis results for (a) *SNX1/CIAO2A* (lead signal) (b) *SNX1/CIAO2A* (secondary signal) and (c) *CTSH*. The forest plots also show the results of the no-proxy and no-biobank meta-analyses (where the UKBB diagnosed rather than the UKBB proxy results were used). In the forest plots, data are presented as odds-ratio with 95% confidence interval. P values are two-sided raw P values derived from a fixed-effect meta-analysis. OR: odds ratio, CI: confidence interval, EA: effect allele, EAF: effect allele frequency range across all studies, HetP: heterogeneity P value, HetISq: heterogeneity statistic.

a)

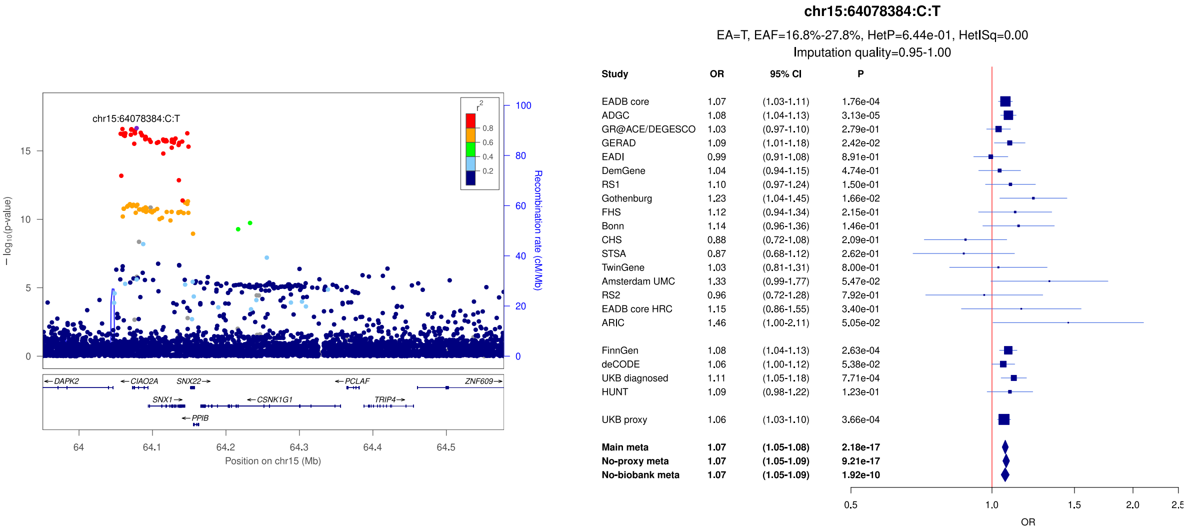


b)

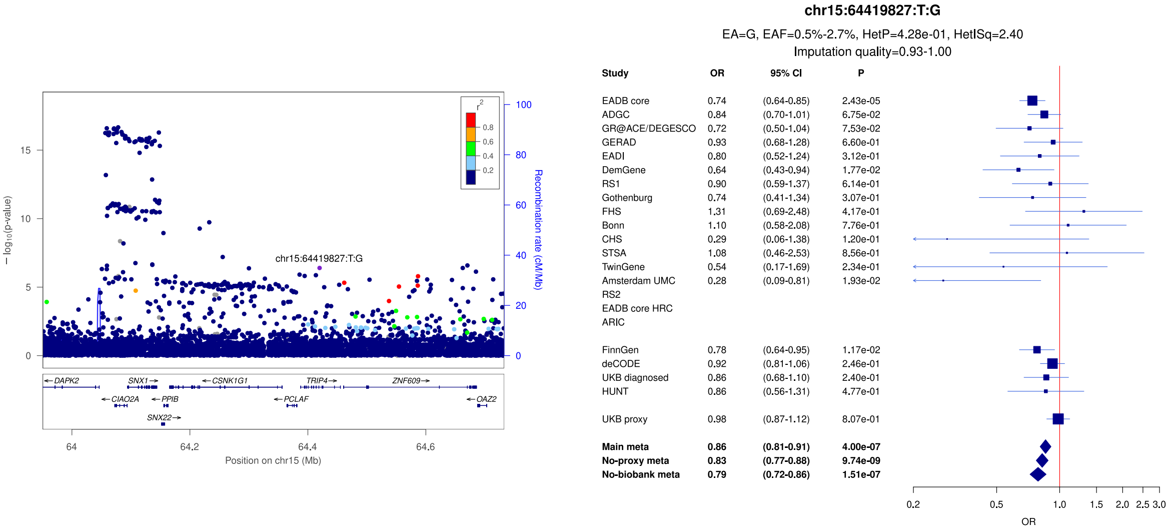


c)

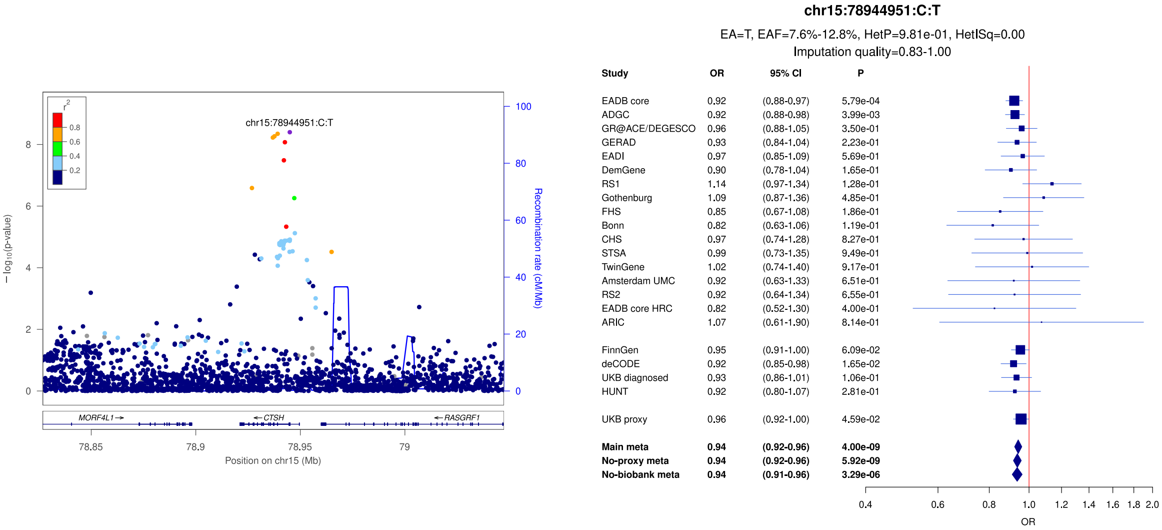

**Supplementary Figure 38:** LocusZoom and forest plots of the main analysis results for (a) *AXIN1* (lead signal) (b) *AXIN1* (secondary signal) and (c) *IQCK*. The forest plots also show the results of the no-proxy and no-biobank meta-analyses (where the UKBB diagnosed rather than the UKBB proxy results were used). In the forest plots, data are presented as odds-ratio with 95% confidence interval. P values are two-sided raw P values derived from a fixed-effect meta-analysis. OR: odds ratio, CI: confidence interval, EA: effect allele, EAF: effect allele frequency range across all studies, HetP: heterogeneity P value, HetISq: heterogeneity statistic.

a)

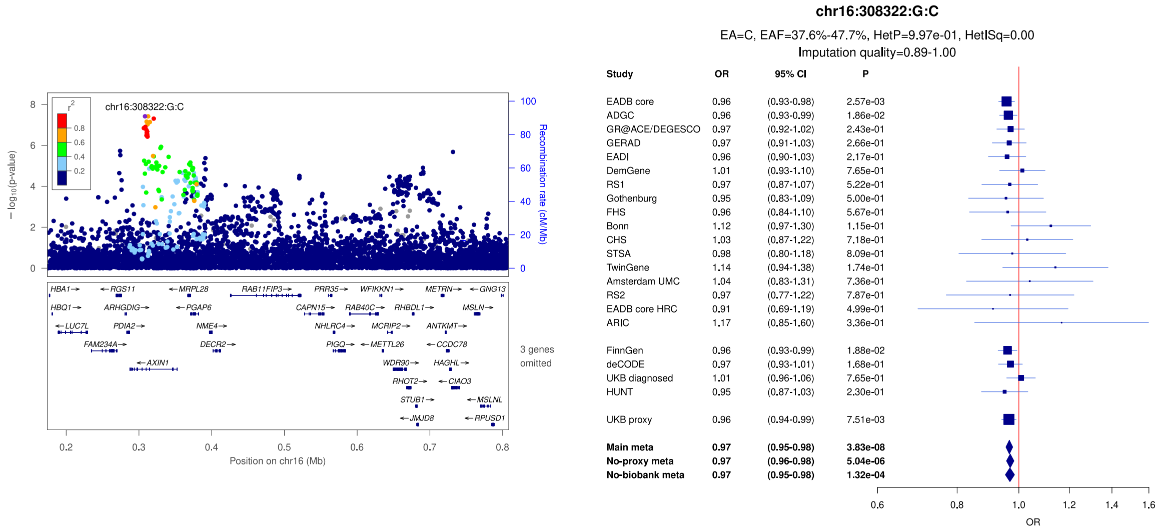


b)

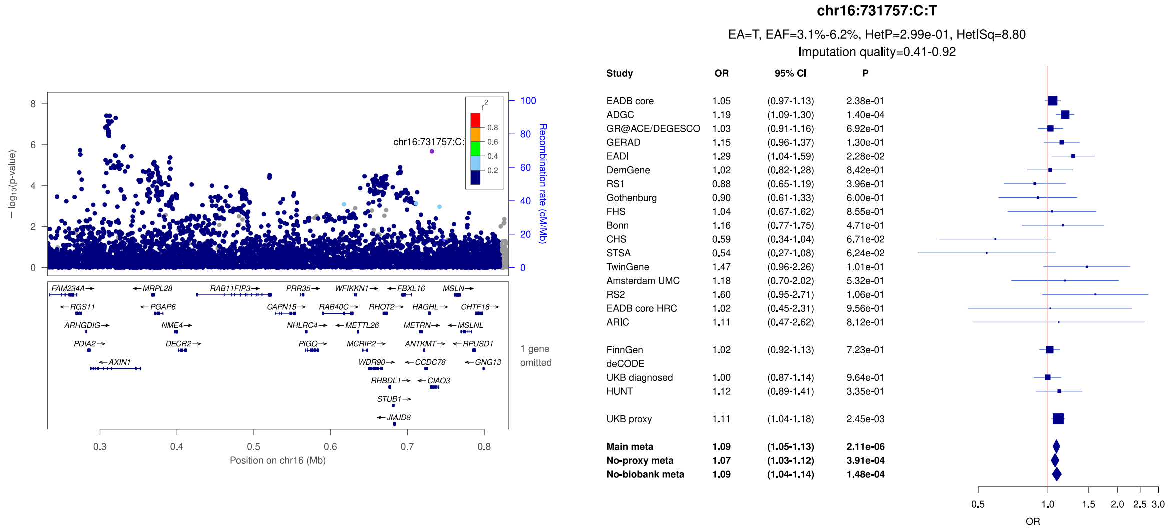


c)

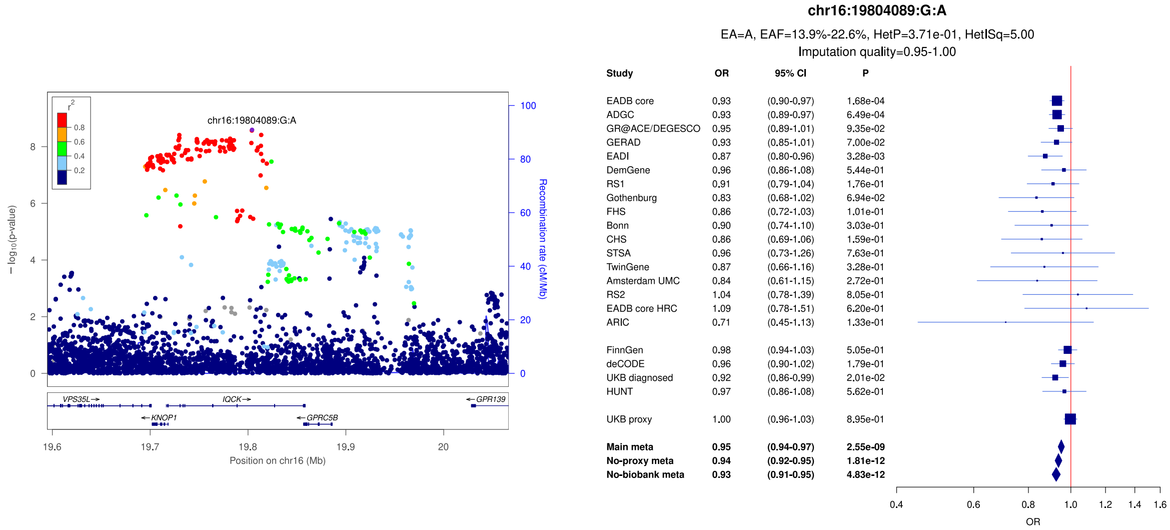

**Supplementary Figure 39:** LocusZoom and forest plots of the main analysis results for (a) *UBFD1* (b) *DOC2A* and (c) *KAT8/BCKDK*. The forest plots also show the results of the no-proxy and no-biobank meta-analyses (where the UKBB diagnosed rather than the UKBB proxy results were used). In the forest plots, data are presented as odds-ratio with 95% confidence interval. P values are two-sided raw P values derived from a fixed-effect meta-analysis. OR: odds ratio, CI: confidence interval, EA: effect allele, EAF: effect allele frequency range across all studies, HetP: heterogeneity P value, HetISq: heterogeneity statistic.

a)

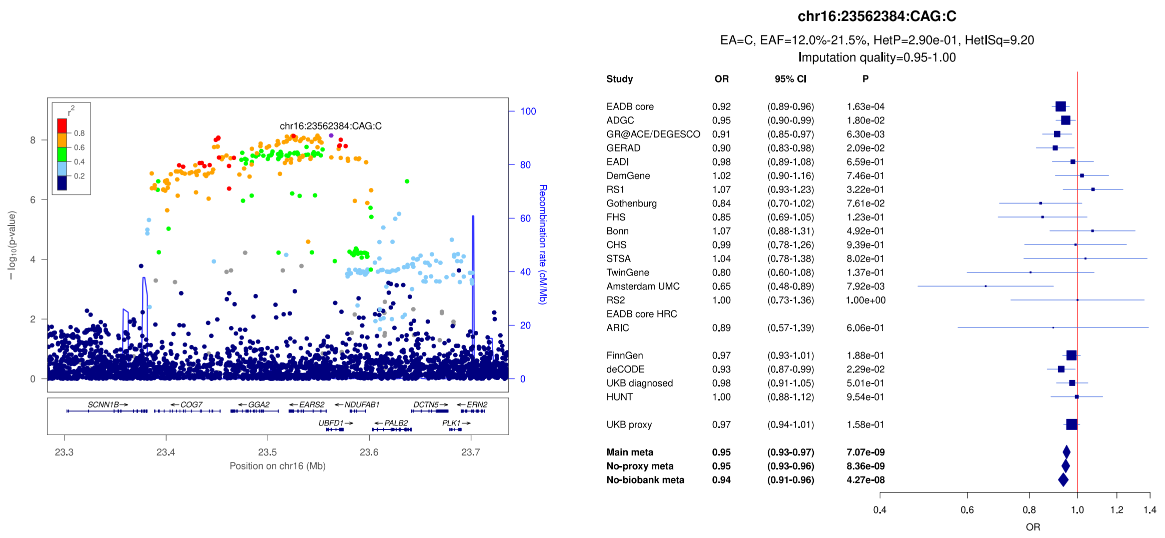


b)

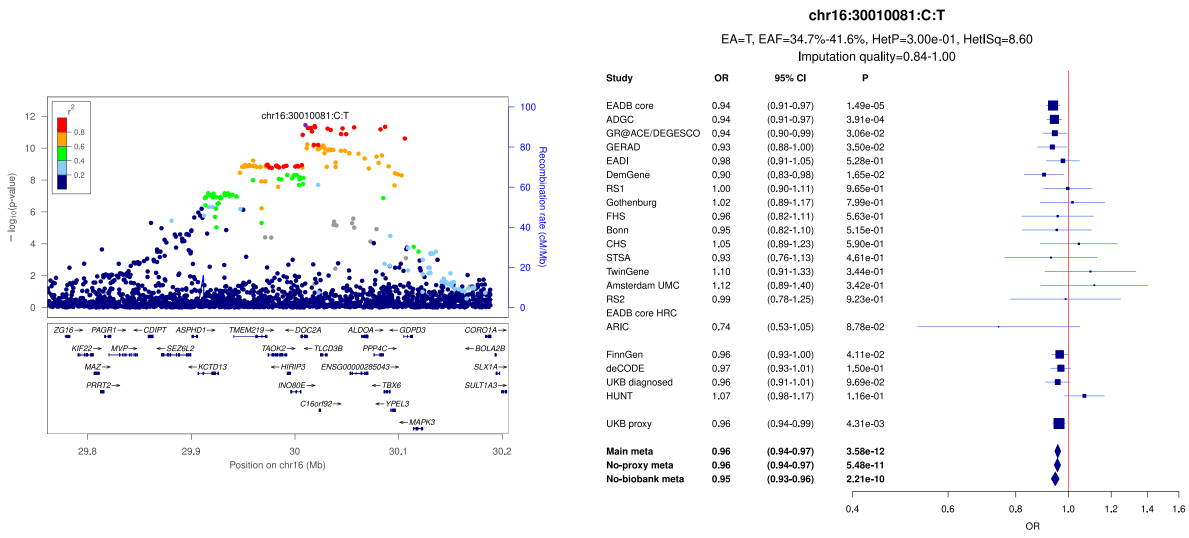


c)

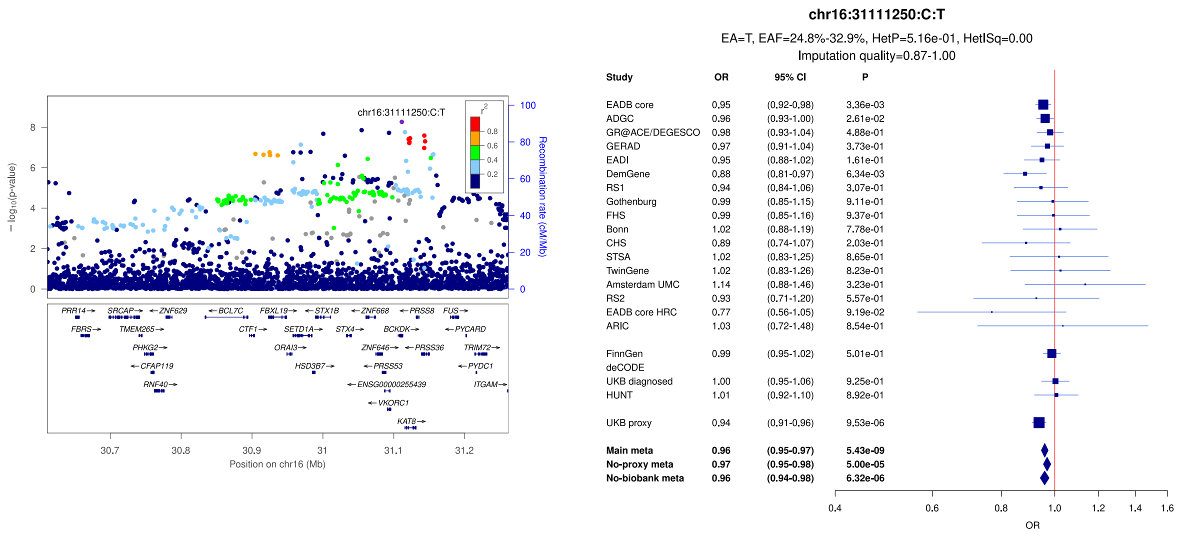

**Supplementary Figure 40:** LocusZoom and forest plots of the main analysis results for (a) *IL34/MTSS2* (b) *MAF* and (c) *PLCG2* (secondary signal). The forest plots also show the results of the no-proxy and no-biobank meta-analyses (where the UKBB diagnosed rather than the UKBB proxy results were used). In the forest plots, data are presented as odds-ratio with 95% confidence interval. P values are two-sided raw P values derived from a fixed-effect meta-analysis. OR: odds ratio, CI: confidence interval, EA: effect allele, EAF: effect allele frequency range across all studies, HetP: heterogeneity P value, HetISq: heterogeneity statistic.

a)

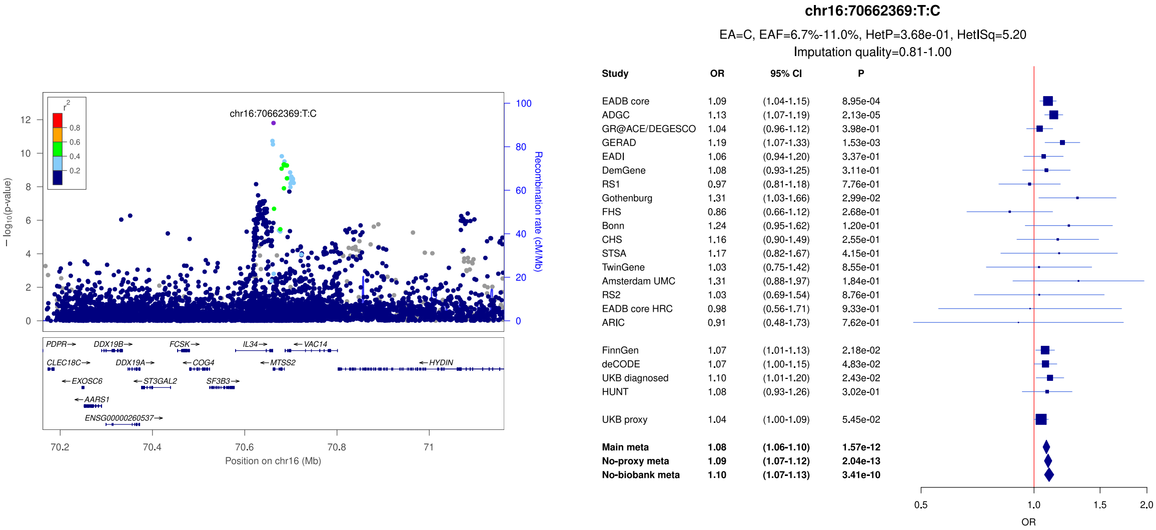


b)

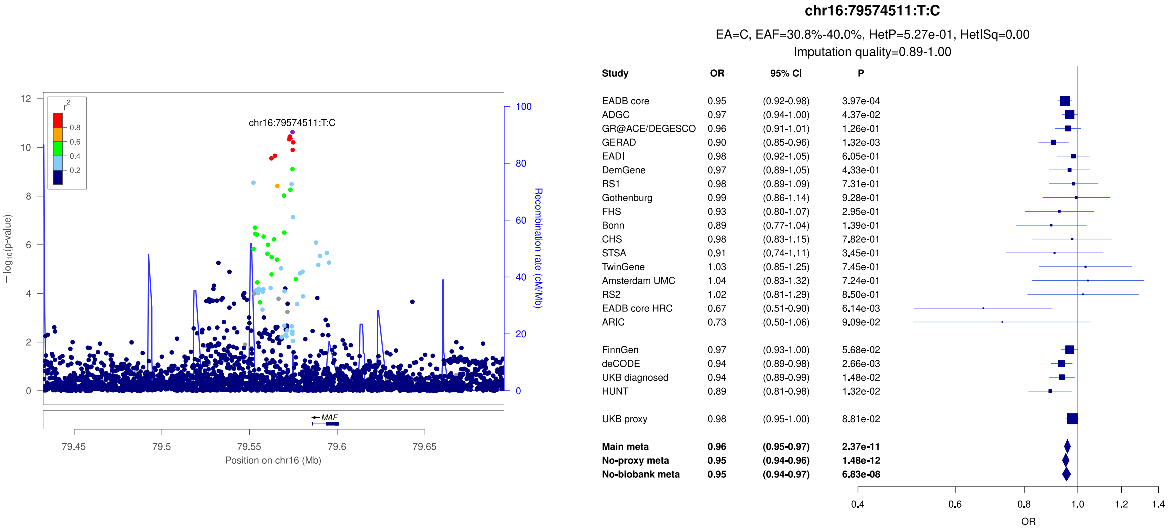


c)

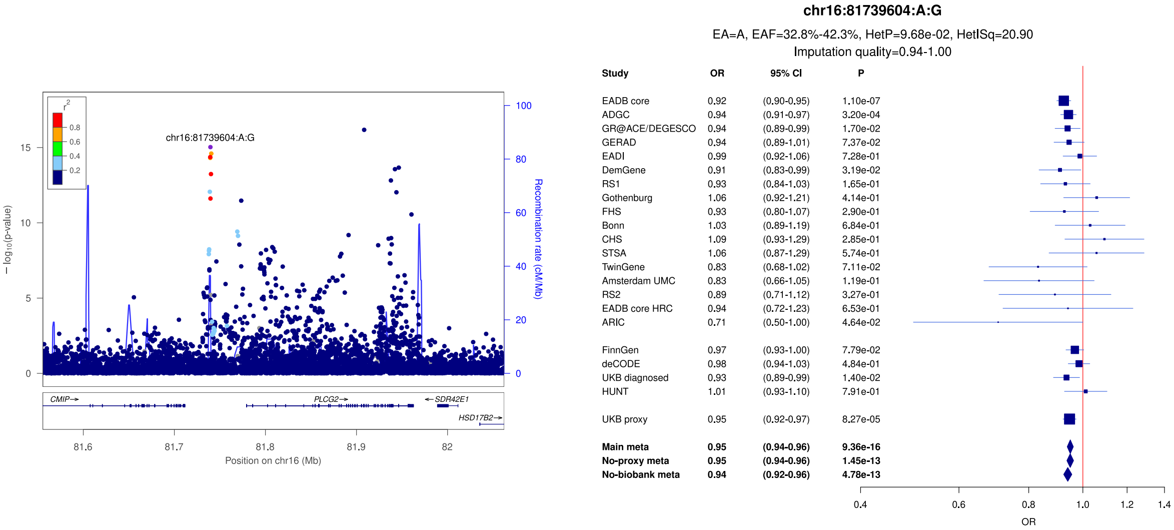

**Supplementary Figure 41:** LocusZoom and forest plots of the main analysis results for secondary signals in *PLCG2* (a, b, c). The forest plots also show the results of the no-proxy and no-biobank meta-analyses (where the UKBB diagnosed rather than the UKBB proxy results were used). In the forest plots, data are presented as odds-ratio with 95% confidence interval. P values are two-sided raw P values derived from a fixed-effect meta-analysis. OR: odds ratio, CI: confidence interval, EA: effect allele, EAF: effect allele frequency range across all studies, HetP: heterogeneity P value, HetISq: heterogeneity statistic.

a)

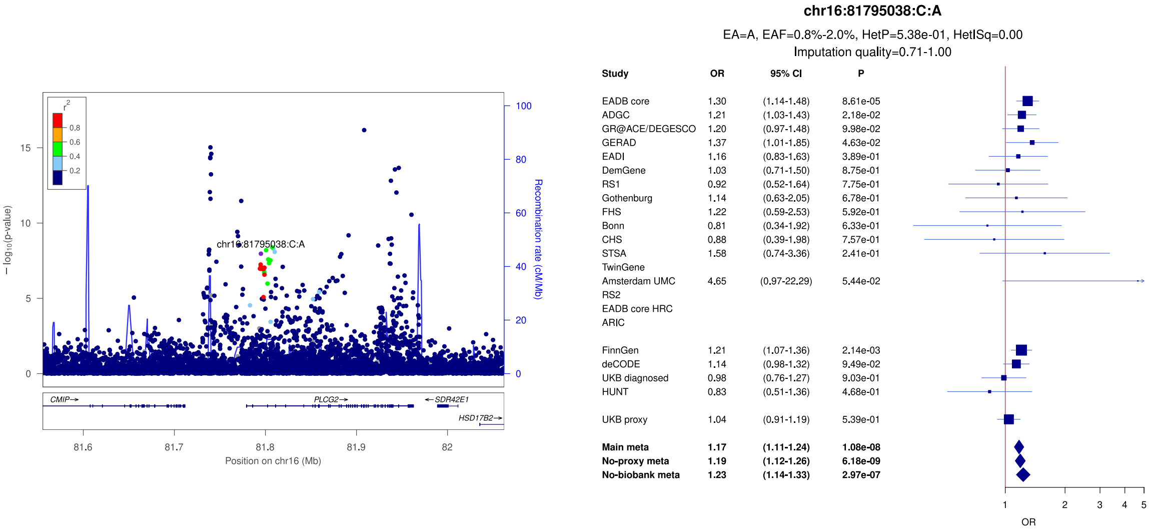


b)

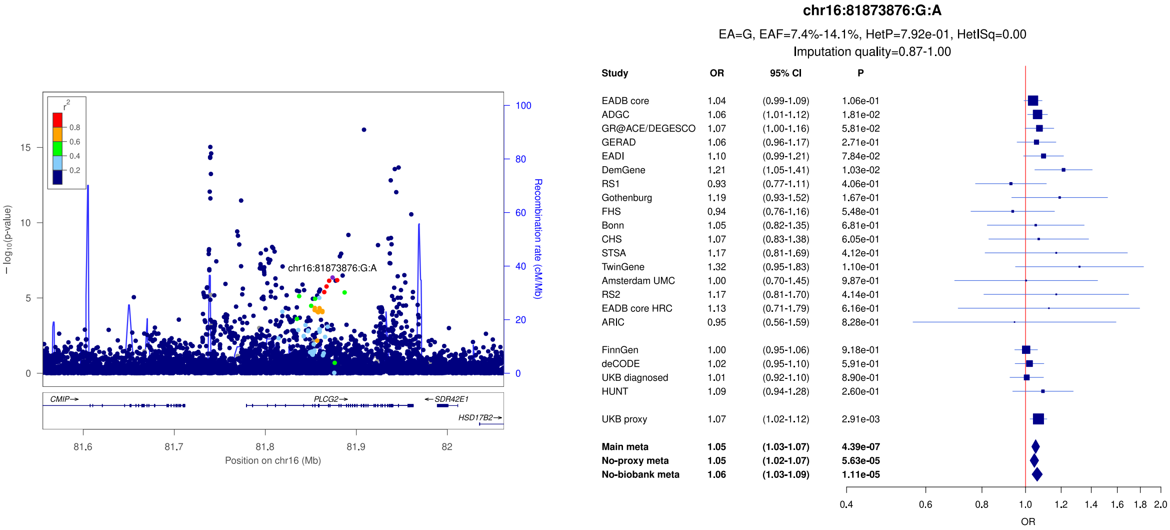


c)

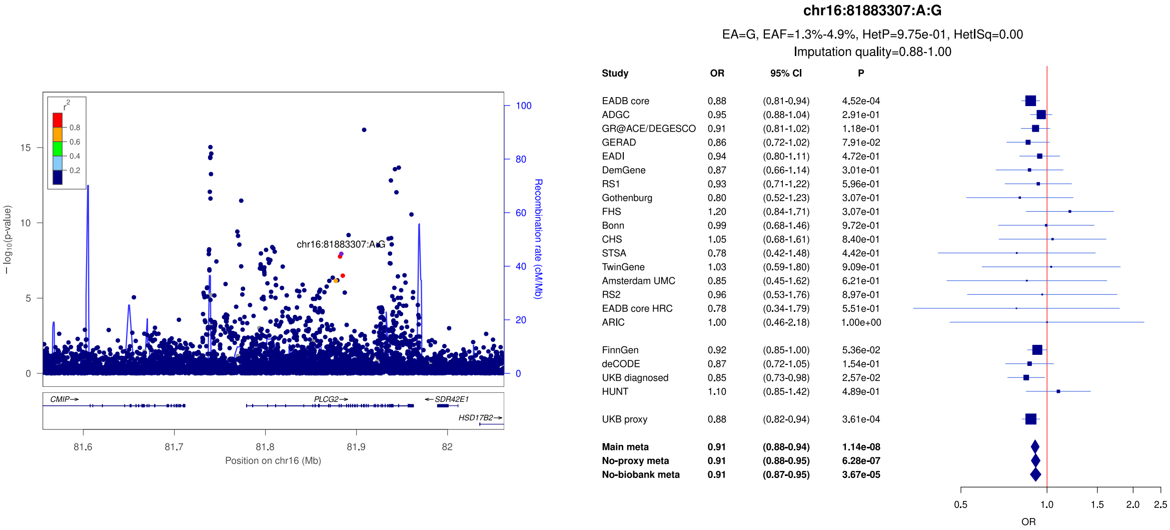

**Supplementary Figure 42:** LocusZoom and forest plots of the main analysis results for (a) *PLCG2* (lead signal) (b) *PLCG2* (secondary signal) and (c) *PRDM7* (secondary signal). The forest plots also show the results of the no-proxy and no-biobank meta-analyses (where the UKBB diagnosed rather than the UKBB proxy results were used). In the forest plots, data are presented as odds-ratio with 95% confidence interval. P values are two-sided raw P values derived from a fixed-effect meta-analysis. OR: odds ratio, CI: confidence interval, EA: effect allele, EAF: effect allele frequency range across all studies, HetP: heterogeneity P value, HetISq: heterogeneity statistic.

a)

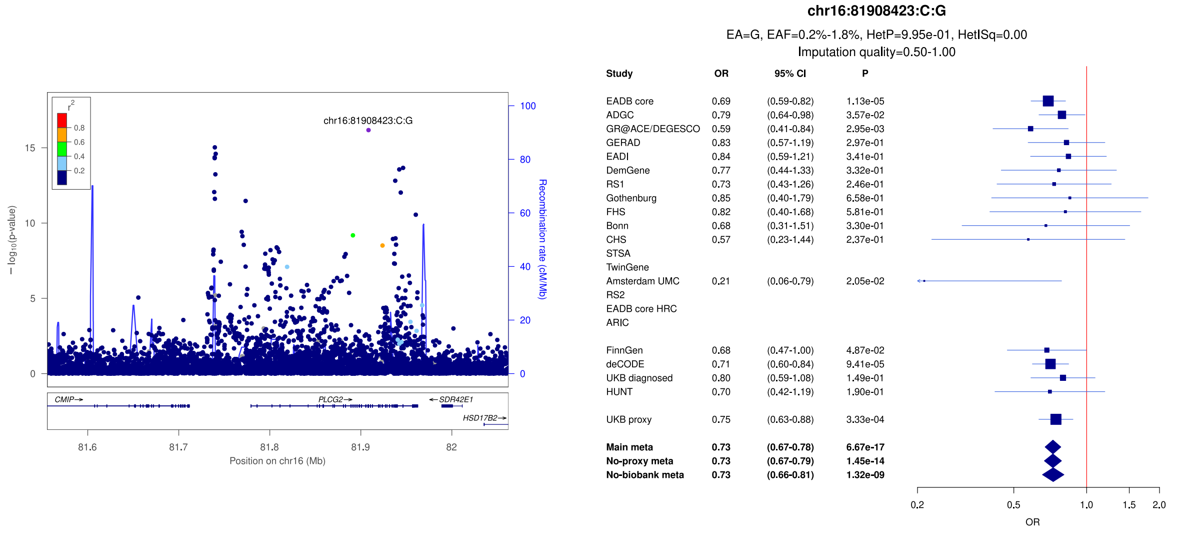


b)

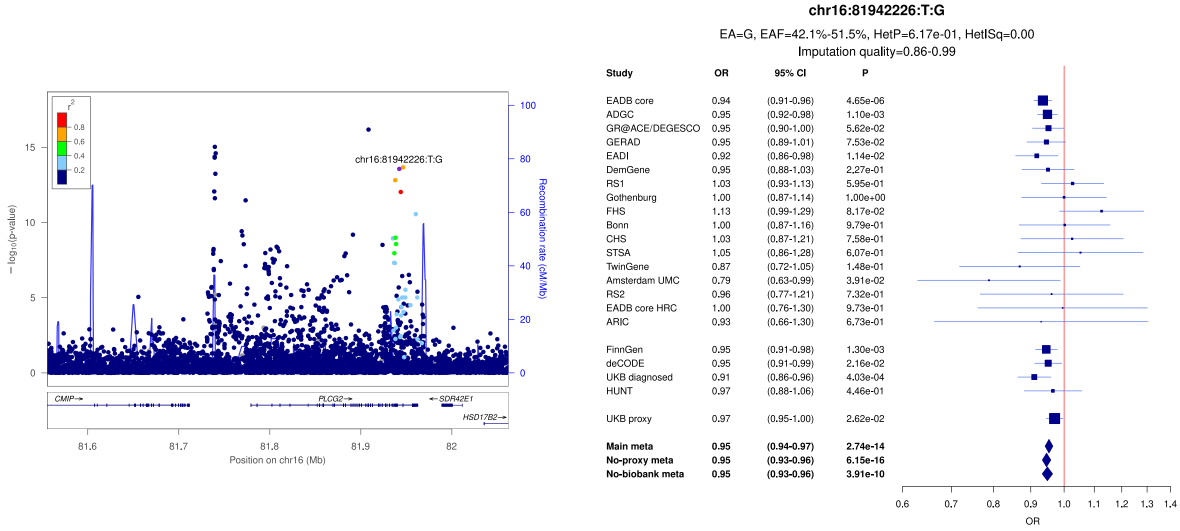


c)

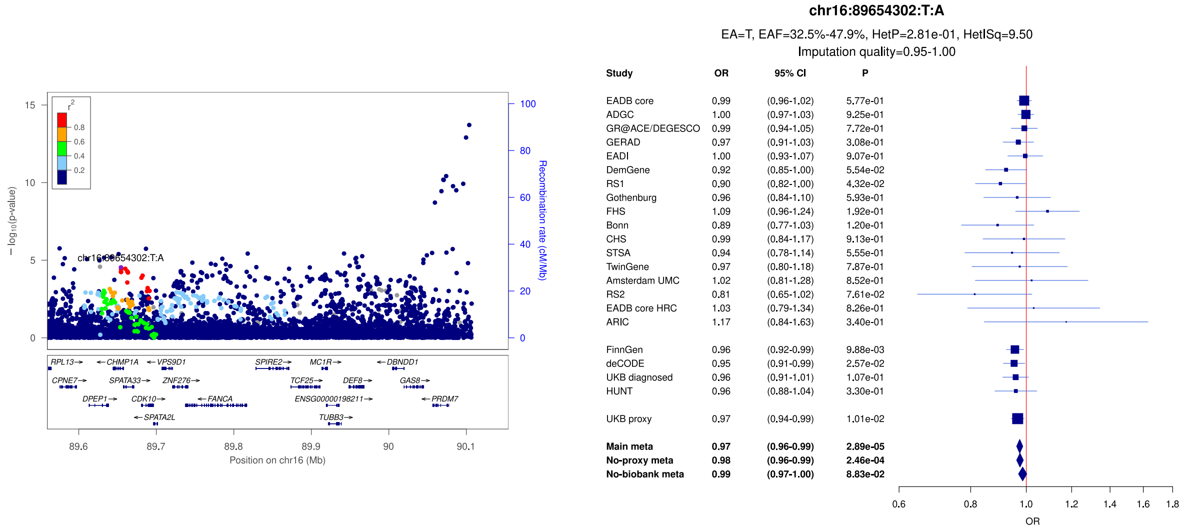

**Supplementary Figure 43:** LocusZoom and forest plots of the main analysis results for (a) *PRDM7* (lead signal) (b) *WDR81/SERPINF2* (secondary signal) and (c) *WDR81/SERPINF2* (lead signal). The forest plots also show the results of the no-proxy and no-biobank meta-analyses (where the UKBB diagnosed rather than the UKBB proxy results were used). In the forest plots, data are presented as odds-ratio with 95% confidence interval. P values are two-sided raw P values derived from a fixed-effect meta-analysis. OR: odds ratio, CI: confidence interval, EA: effect allele, EAF: effect allele frequency range across all studies, HetP: heterogeneity P value, HetISq: heterogeneity statistic.

a)

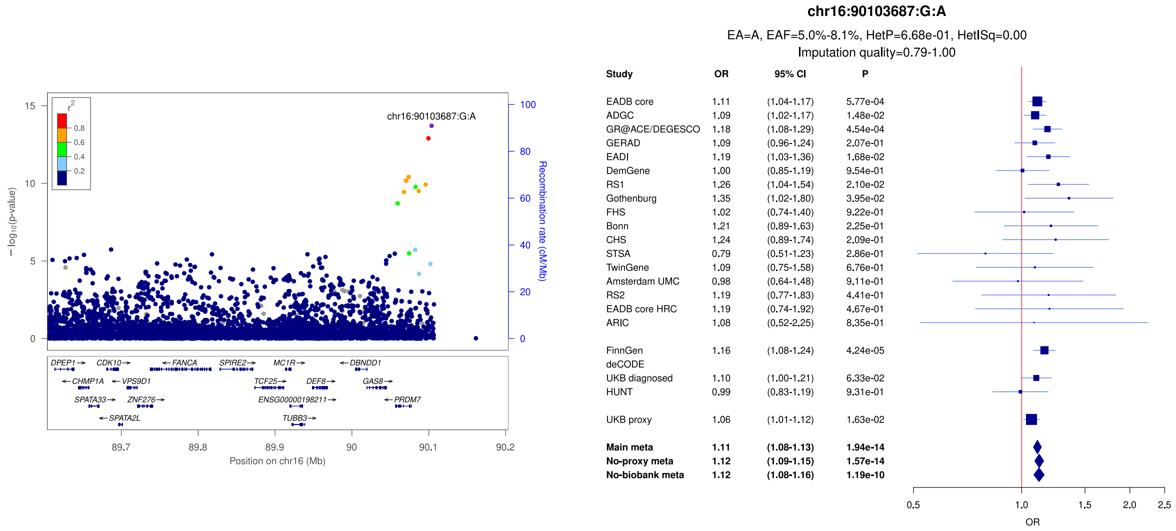


b)

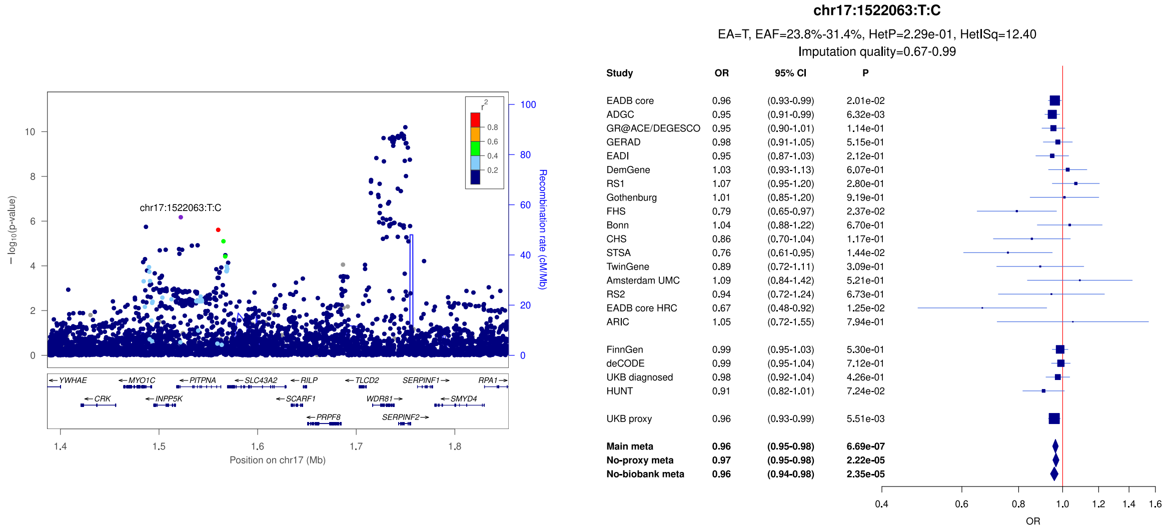


c)

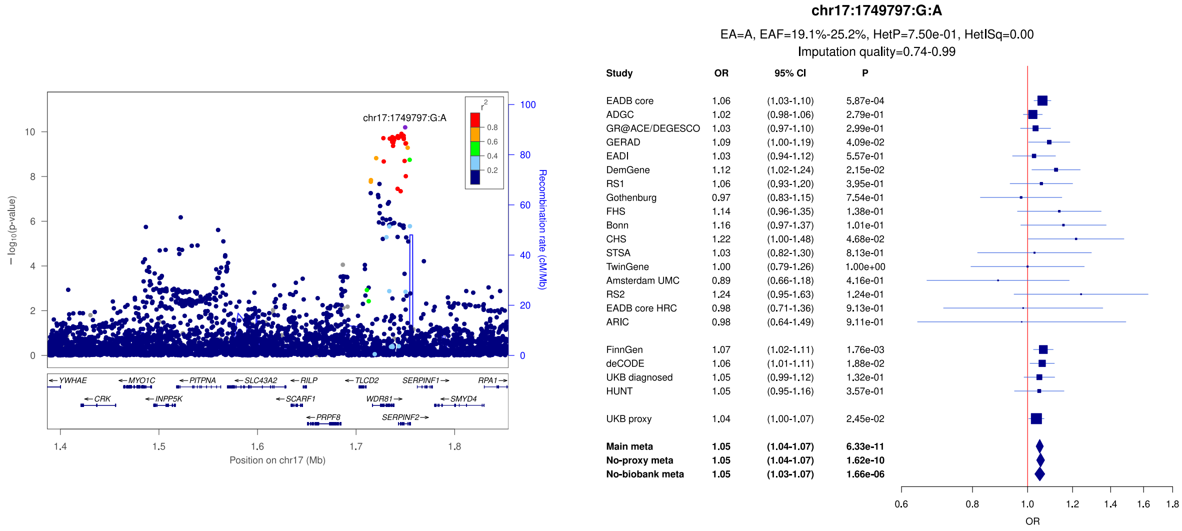

**Supplementary Figure 44:** LocusZoom and forest plots of the main analysis results for (a) *SCIMP/RABEP1* (secondary signal) (b) *SCIMP/RABEP1* (lead signal) and (c) *MYO15A/TOM1L2*. The forest plots also show the results of the no-proxy and no-biobank meta-analyses (where the UKBB diagnosed rather than the UKBB proxy results were used). In the forest plots, data are presented as odds-ratio with 95% confidence interval. P values are two-sided raw P values derived from a fixed-effect meta-analysis. OR: odds ratio, CI: confidence interval, EA: effect allele, EAF: effect allele frequency range across all studies, HetP: heterogeneity P value, HetISq: heterogeneity statistic.

a)

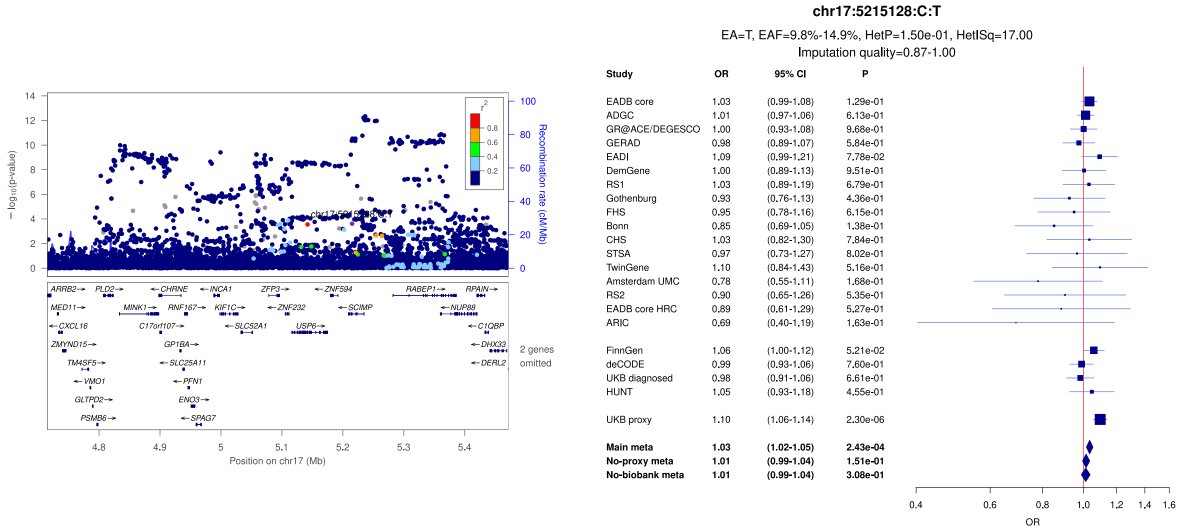


b)

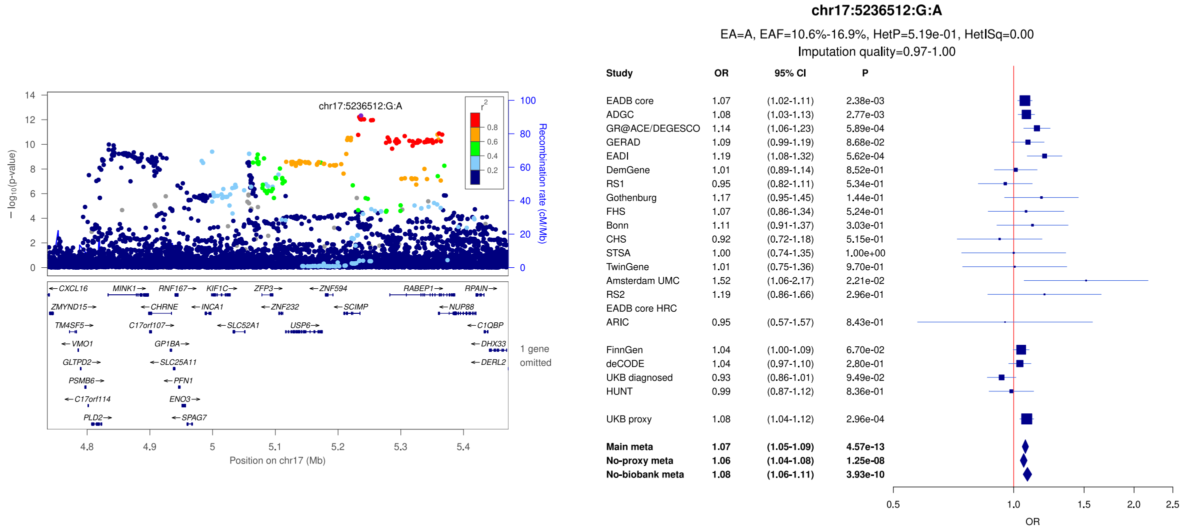


c)

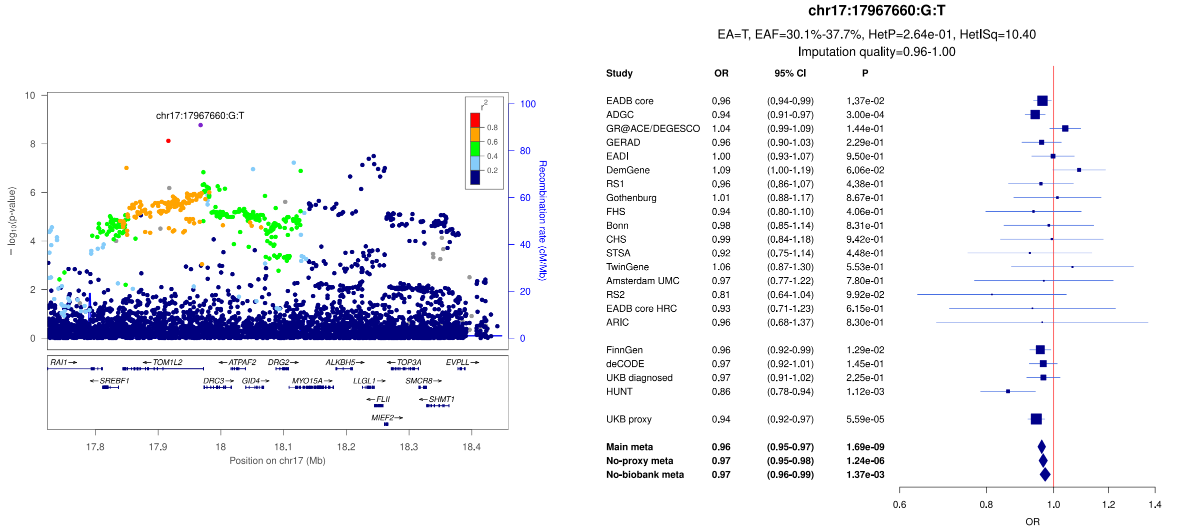

**Supplementary Figure 45:** LocusZoom and forest plots of the main analysis results for (a) *GRN* (b) *MAPT* (secondary signal) and (c) *MAPT* (lead signal). The forest plots also show the results of the no-proxy and no-biobank meta-analyses (where the UKBB diagnosed rather than the UKBB proxy results were used). In the forest plots, data are presented as odds-ratio with 95% confidence interval. P values are two-sided raw P values derived from a fixed-effect meta-analysis. OR: odds ratio, CI: confidence interval, EA: effect allele, EAF: effect allele frequency range across all studies, HetP: heterogeneity P value, HetISq: heterogeneity statistic.

a)

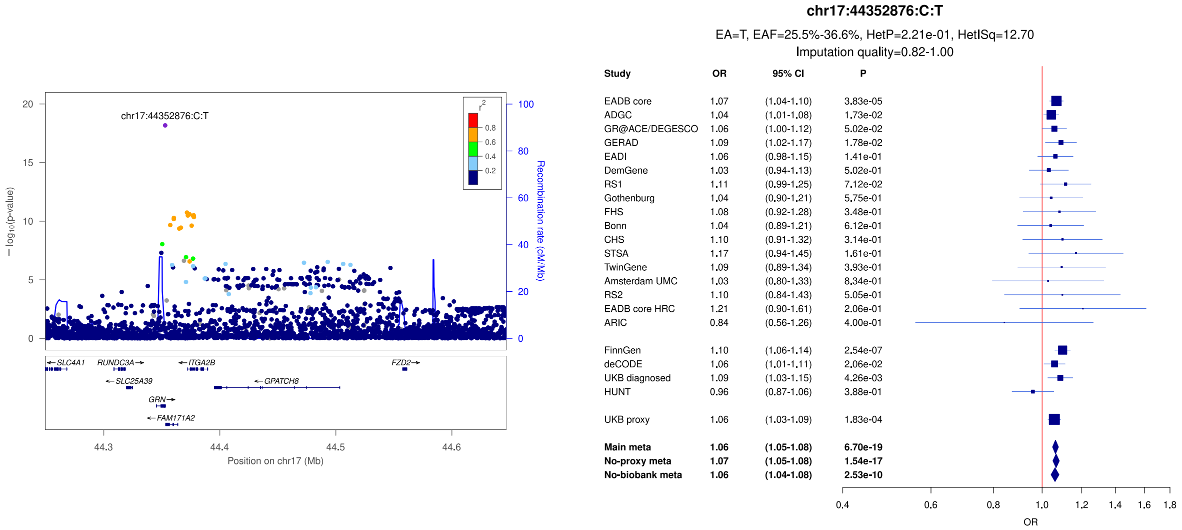


b)

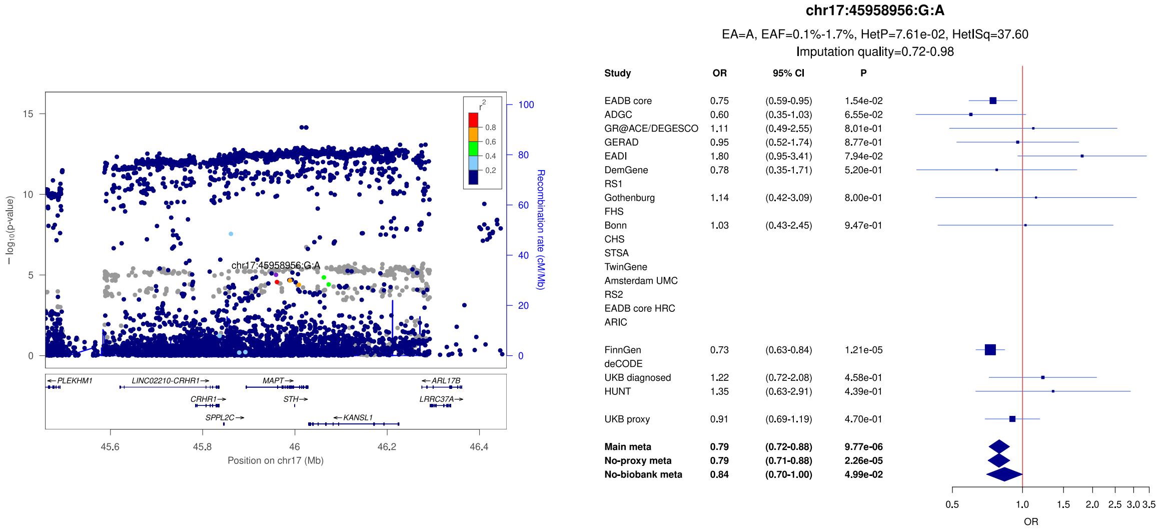


c)

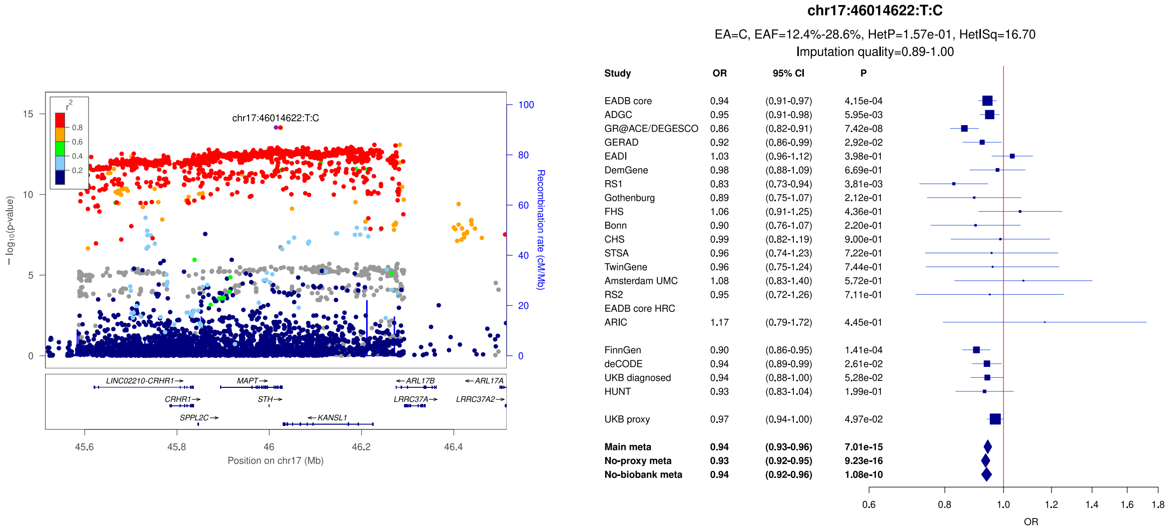

**Supplementary Figure 46:** LocusZoom and forest plots of the main analysis results for (a) *ABI3* (lead signal) (b) *ABI3* (secondary signal) and (c) *TSPOAP1*. The forest plots also show the results of the no-proxy and no-biobank meta-analyses (where the UKBB diagnosed rather than the UKBB proxy results were used). In the forest plots, data are presented as odds-ratio with 95% confidence interval. P values are two-sided raw P values derived from a fixed-effect meta-analysis. OR: odds ratio, CI: confidence interval, EA: effect allele, EAF: effect allele frequency range across all studies, HetP: heterogeneity P value, HetISq: heterogeneity statistic.

a)

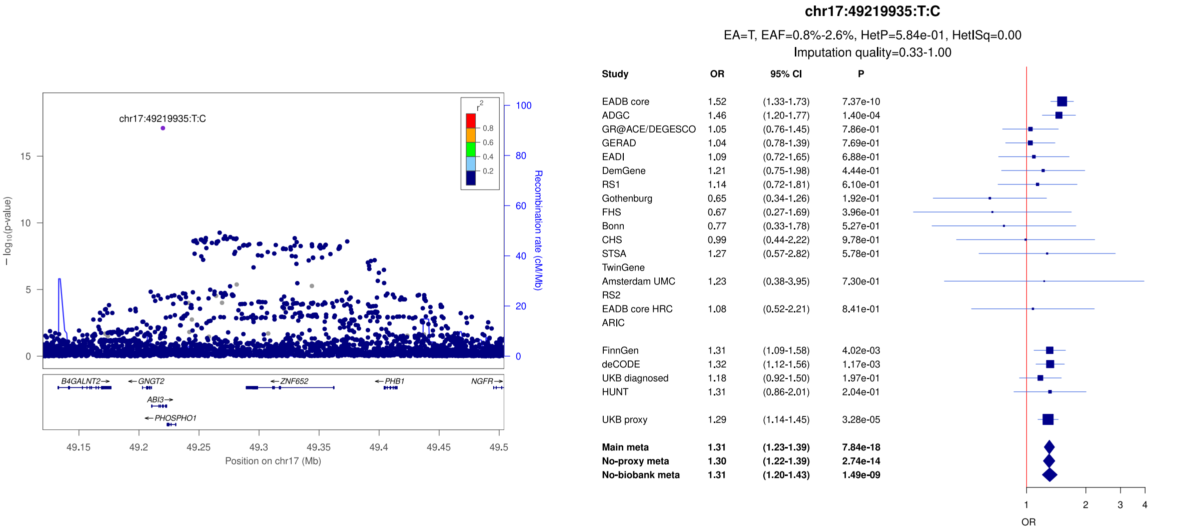


b)

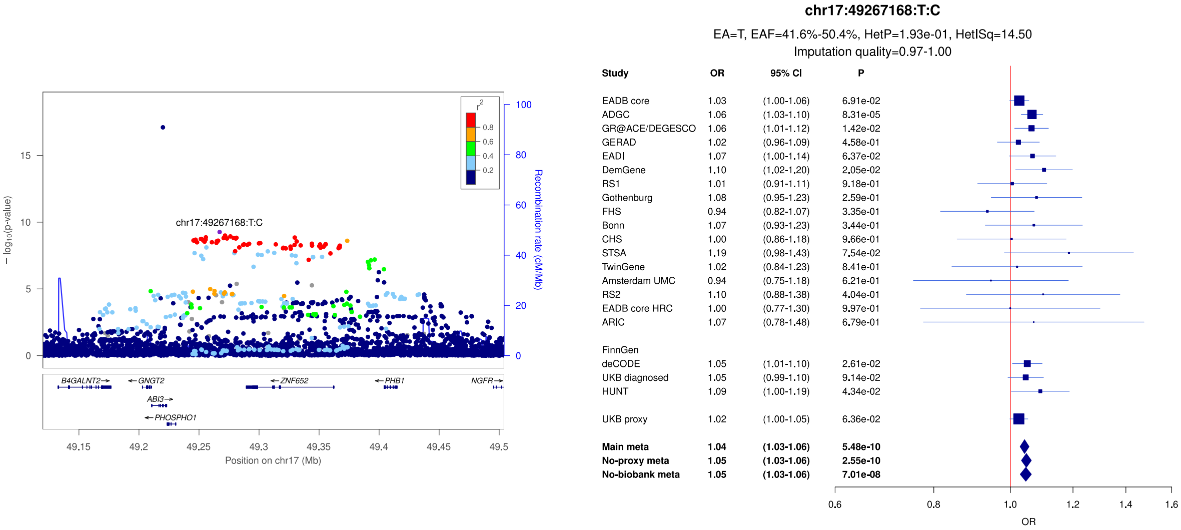


c)

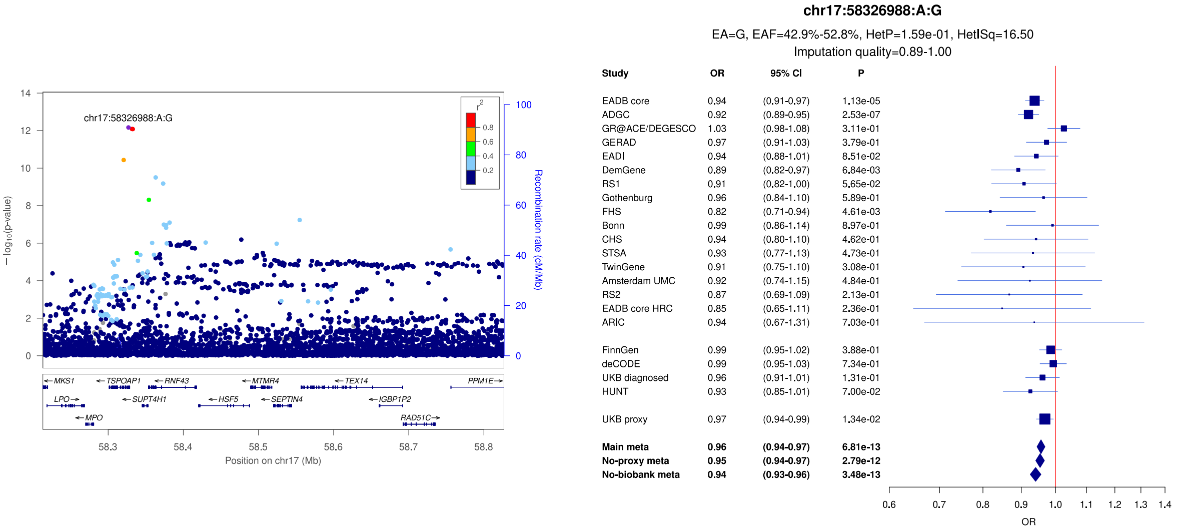

**Supplementary Figure 47:** LocusZoom and forest plots of the main analysis results for (a) *ACE* (secondary signal) (b) *ACE* (lead signal) and (c) *ABCA7* (secondary signal). The forest plots also show the results of the no-proxy and no-biobank meta-analyses (where the UKBB diagnosed rather than the UKBB proxy results were used). In the forest plots, data are presented as odds-ratio with 95% confidence interval. P values are two-sided raw P values derived from a fixed-effect meta-analysis. OR: odds ratio, CI: confidence interval, EA: effect allele, EAF: effect allele frequency range across all studies, HetP: heterogeneity P value, HetISq: heterogeneity statistic.

a)

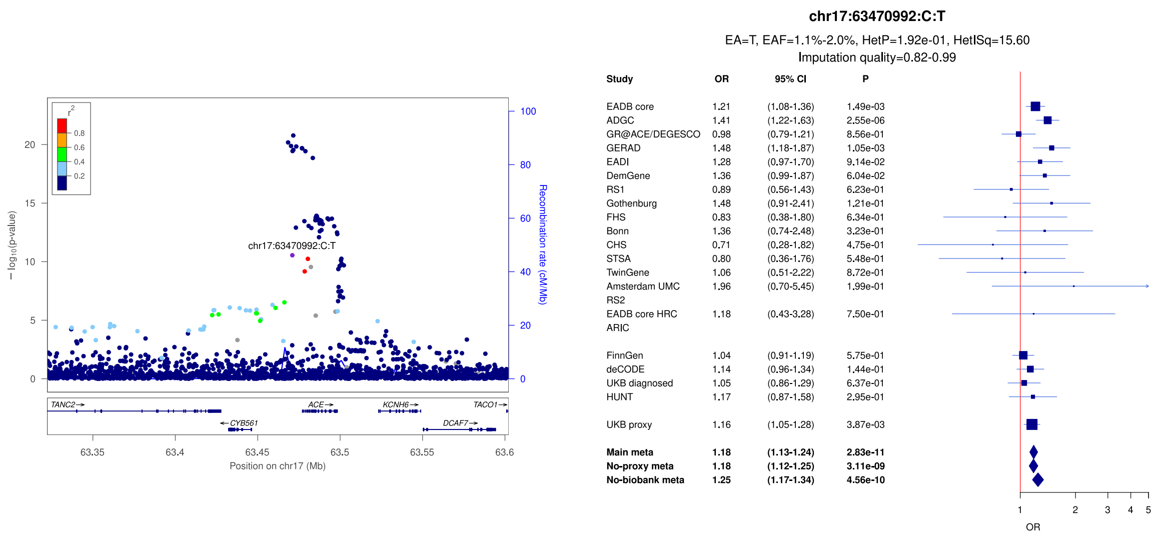


b)

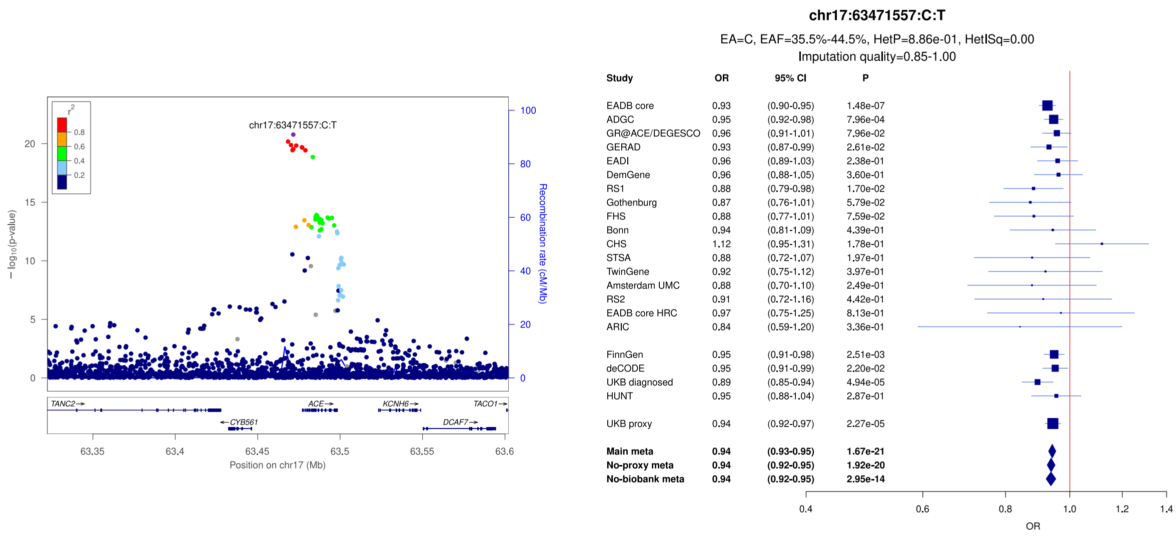


c)

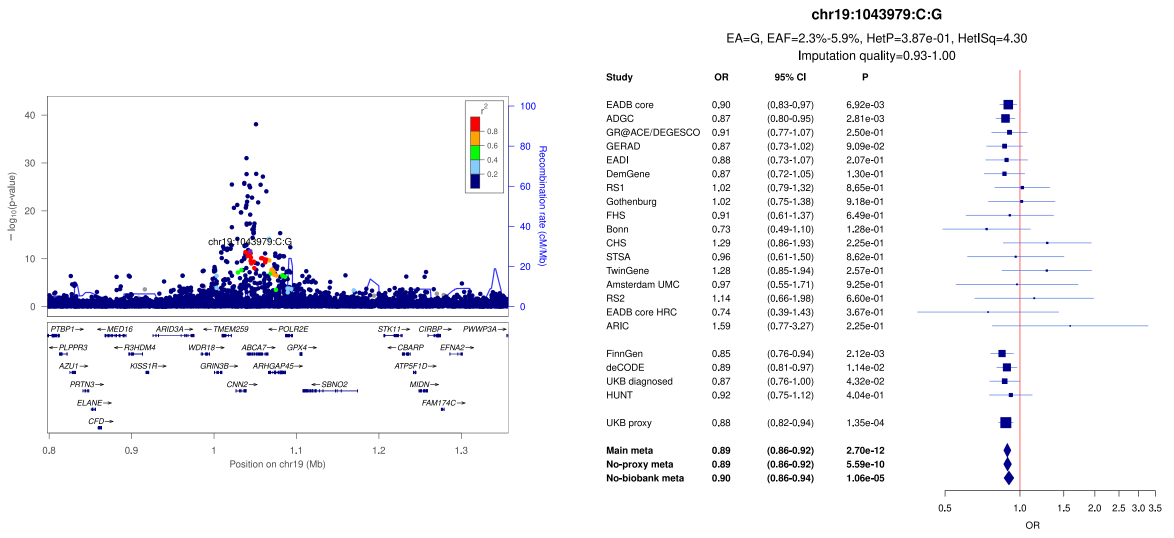

**Supplementary Figure 48:** LocusZoom and forest plots of the main analysis results for ABCA7 (a) lead signal (b) and (c) secondary signals. The forest plots also show the results of the no-proxy and no-biobank meta-analyses (where the UKBB diagnosed rather than the UKBB proxy results were used). In the forest plots, data are presented as odds-ratio with 95% confidence interval. P values are two-sided raw P values derived from a fixed-effect meta-analysis. OR: odds ratio, CI: confidence interval, EA: effect allele, EAF: effect allele frequency range across all studies, HetP: heterogeneity P value, HetISq: heterogeneity statistic.

a)

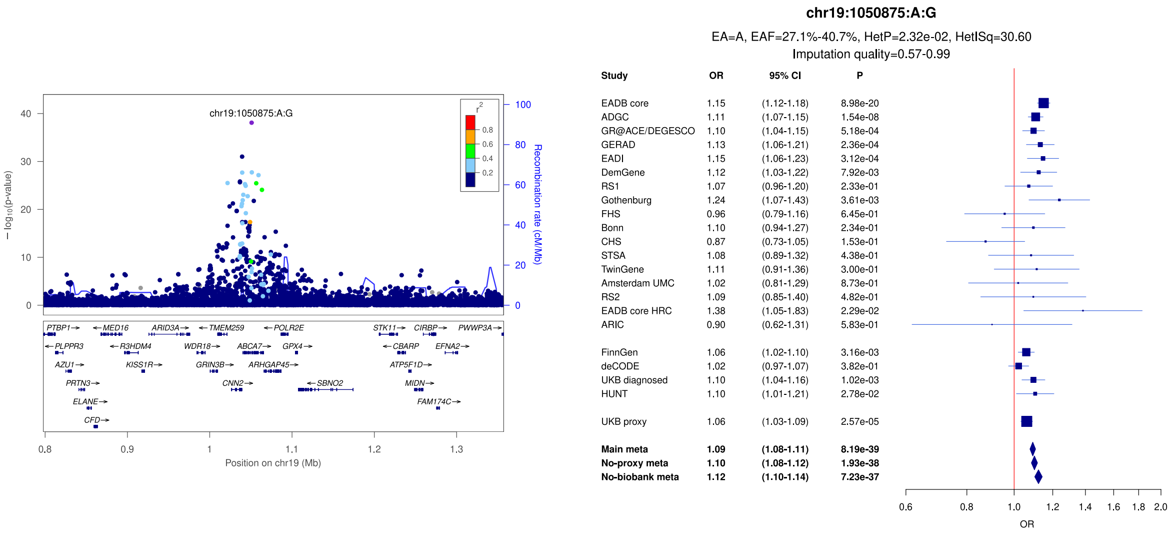


b)

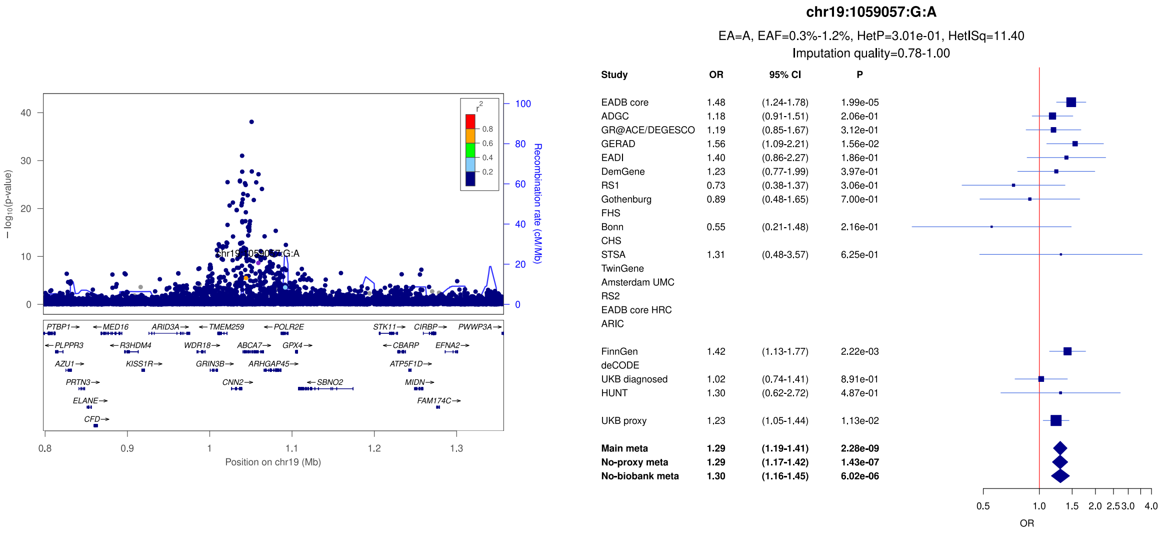


c)

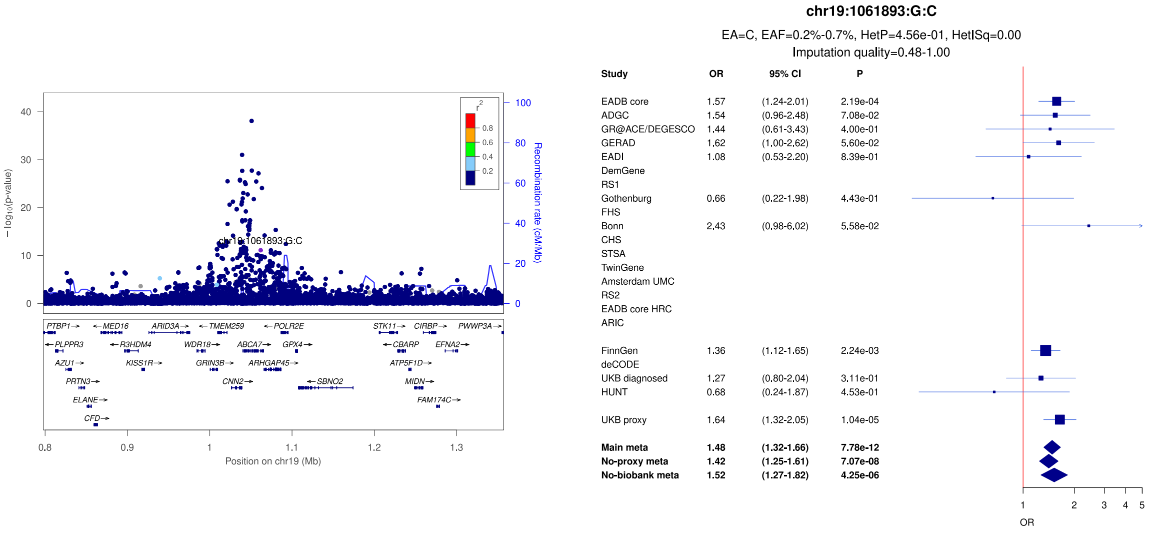

**Supplementary Figure 49:** LocusZoom and forest plots of the main analysis results for (a) *KLF16/REXO1* (b) *VMAC* (secondary signal) and (c) *VMAC* (lead signal). The forest plots also show the results of the no-proxy and no-biobank meta-analyses (where the UKBB diagnosed rather than the UKBB proxy results were used). In the forest plots, data are presented as odds-ratio with 95% confidence interval. P values are two-sided raw P values derived from a fixed-effect meta-analysis. OR: odds ratio, CI: confidence interval, EA: effect allele, EAF: effect allele frequency range across all studies, HetP: heterogeneity P value, HetISq: heterogeneity statistic.

a)

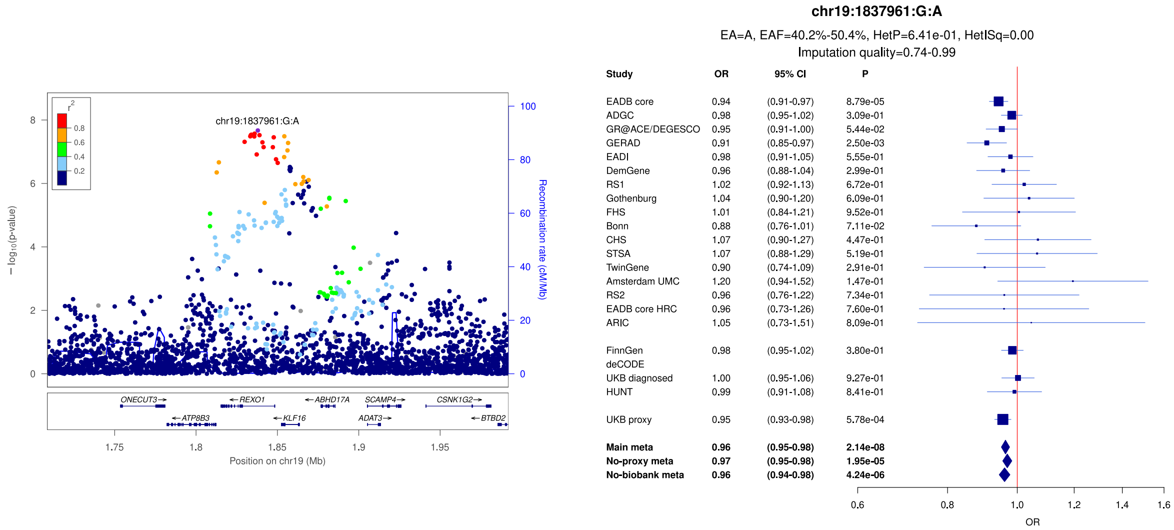


b)

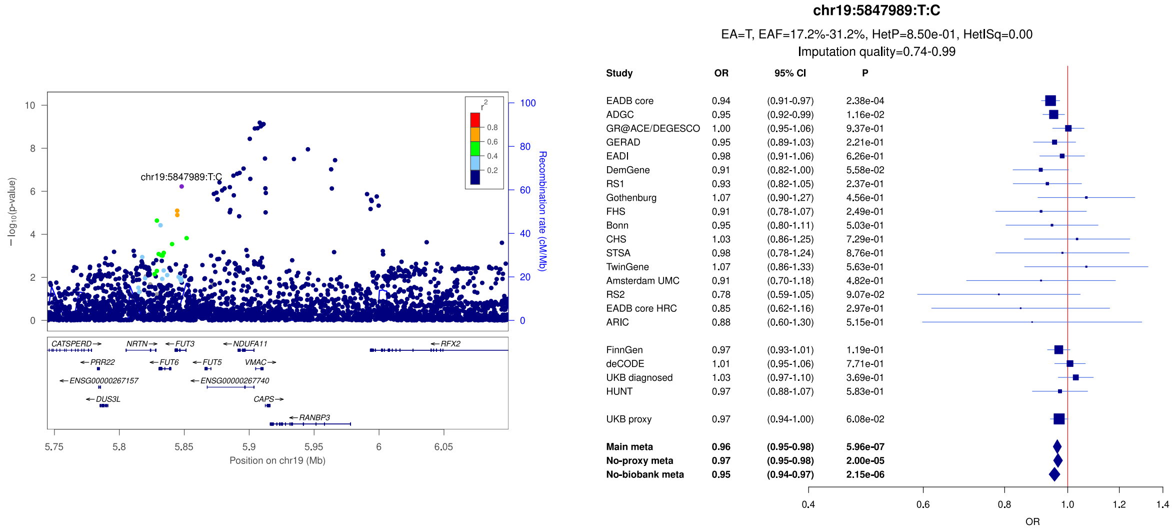


c)

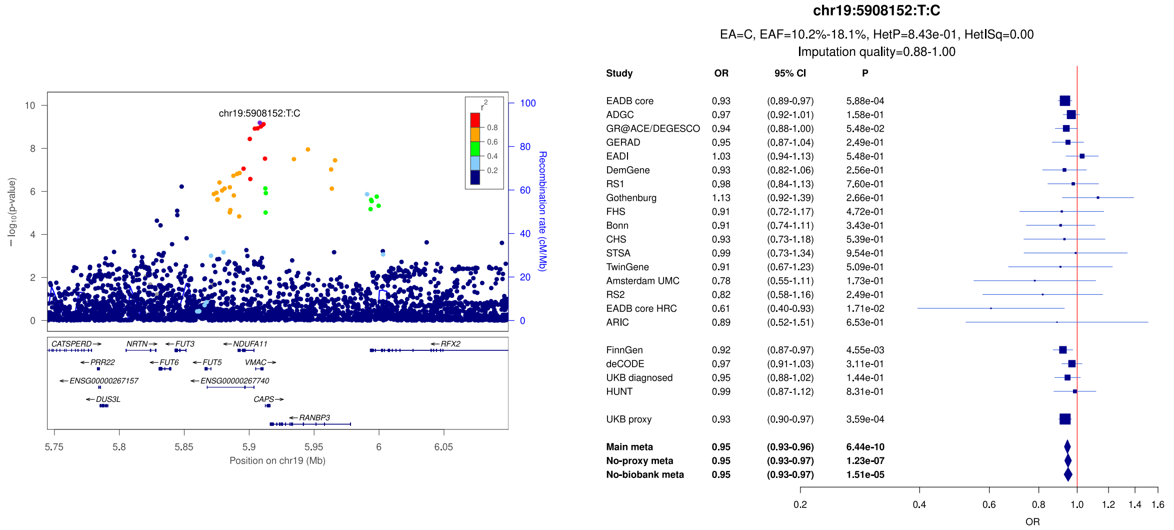

**Supplementary Figure 50:** LocusZoom and forest plots of the main analysis results for (a) *VAV1* (b) *LRRC25* and (c) *CEP89*. The forest plots also show the results of the no-proxy and no-biobank meta-analyses (where the UKBB diagnosed rather than the UKBB proxy results were used). In the forest plots, data are presented as odds-ratio with 95% confidence interval. P values are two-sided raw P values derived from a fixed-effect meta-analysis. OR: odds ratio, CI: confidence interval, EA: effect allele, EAF: effect allele frequency range across all studies, HetP: heterogeneity P value, HetISq: heterogeneity statistic.

a)

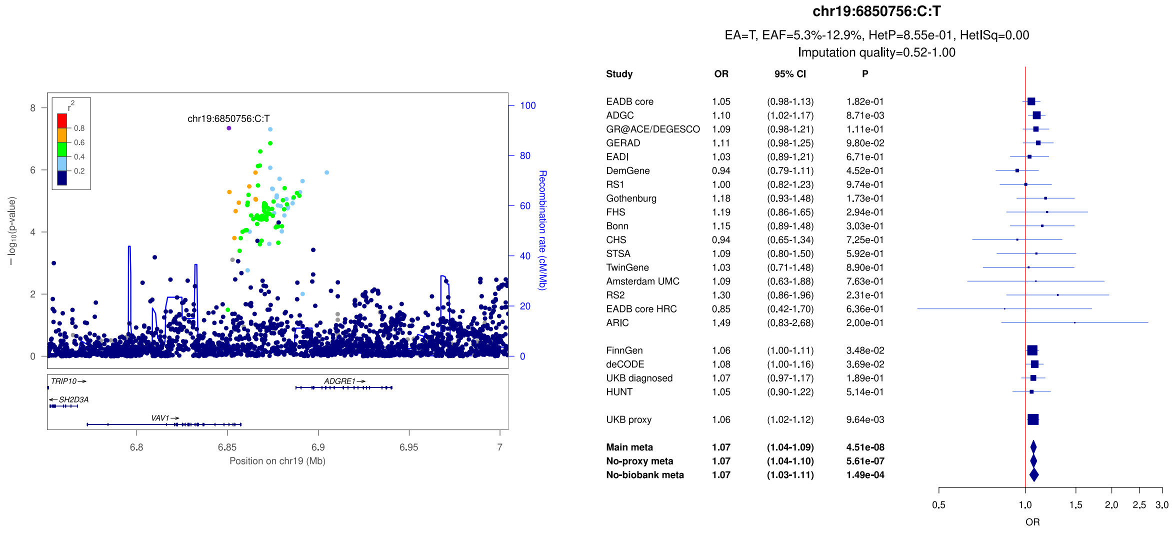


b)

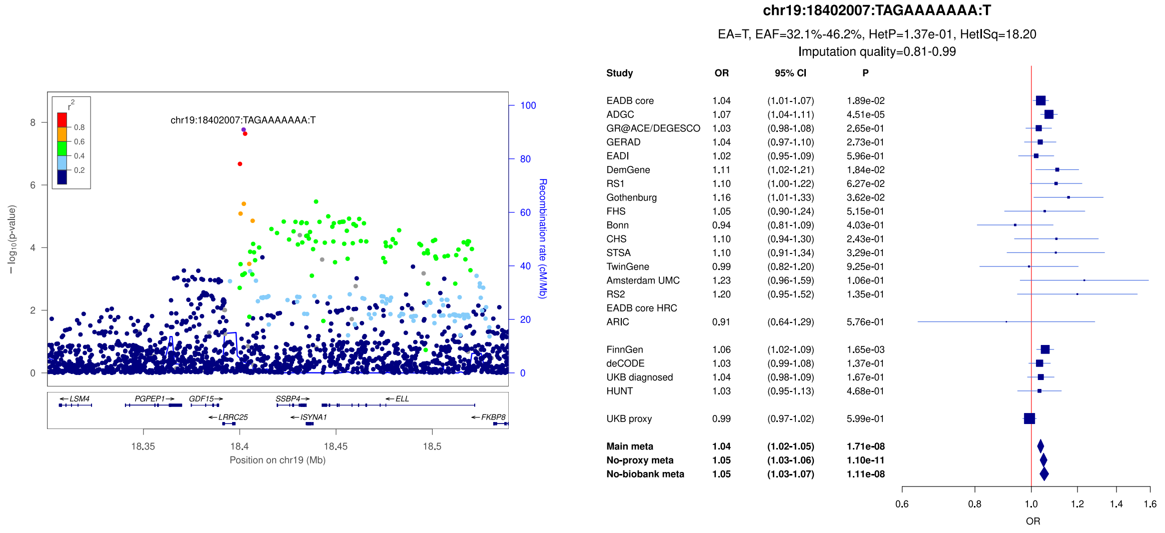


c)

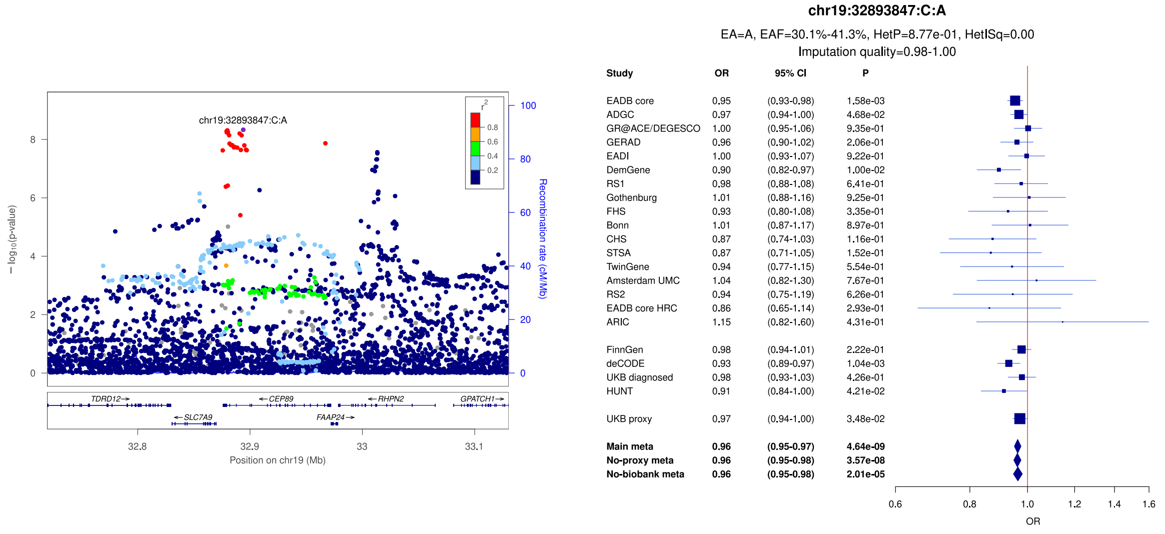

**Supplementary Figure 51:** LocusZoom and forest plots of the main analysis results for (a) *APOE* (P values < $3x10^{-324}$ were set as $3x10^{-324}$) (b) *SIGLEC11* and (c) *CD33* (secondary signal). The forest plots also show the results of the no-proxy and no-biobank meta-analyses (where the UKBB diagnosed rather than the UKBB proxy results were used). In the forest plots, data are presented as odds-ratio with 95% confidence interval. P values are two-sided raw P values derived from a fixed-effect meta-analysis. OR: odds ratio, CI: confidence interval, EA: effect allele, EAF: effect allele frequency range across all studies, HetP: heterogeneity P value, HetISq: heterogeneity statistic.

a)

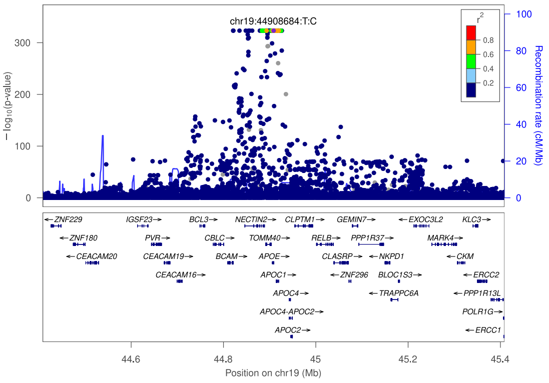


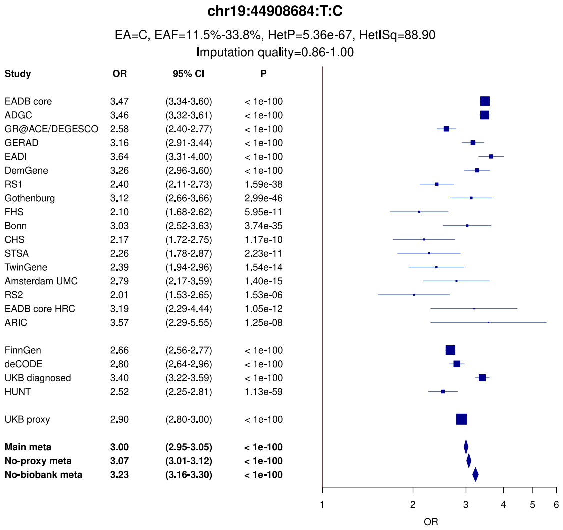


b)

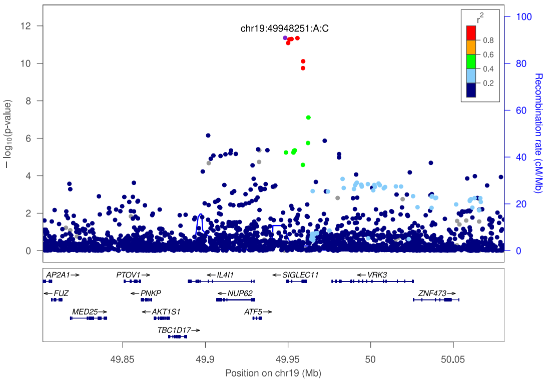


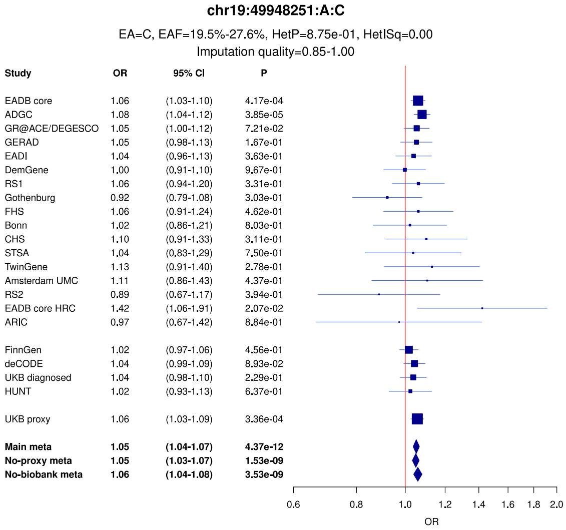


c)

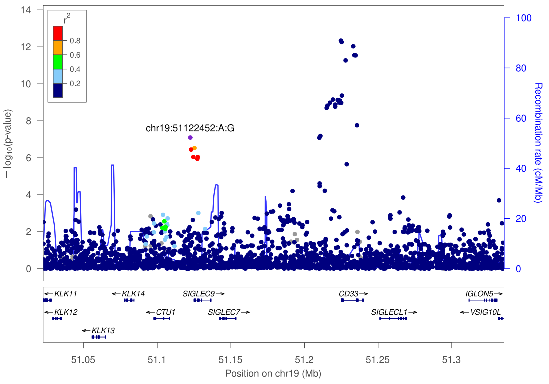


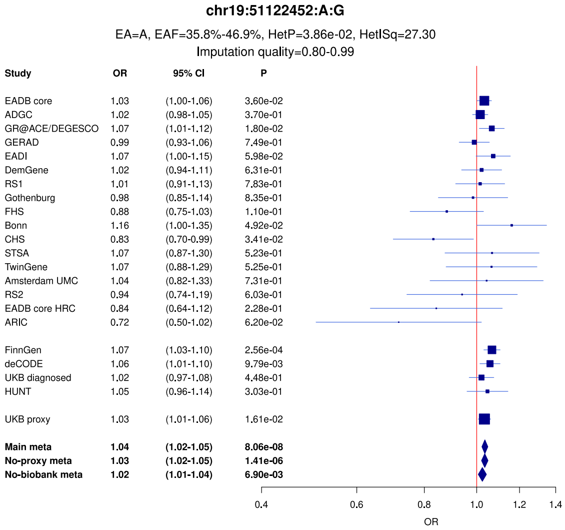

**Supplementary Figure 52:** LocusZoom and forest plots of the main analysis results for (a) *CD33* (b) *LILRB2* (secondary signal) and (c) *LILRB2* (lead signal). The forest plots also show the results of the no-proxy and no-biobank meta-analyses (where the UKBB diagnosed rather than the UKBB proxy results were used). In the forest plots, data are presented as odds-ratio with 95% confidence interval. P values are two-sided raw P values derived from a fixed-effect meta-analysis. OR: odds ratio, CI: confidence interval, EA: effect allele, EAF: effect allele frequency range across all studies, HetP: heterogeneity P value, HetISq: heterogeneity statistic.

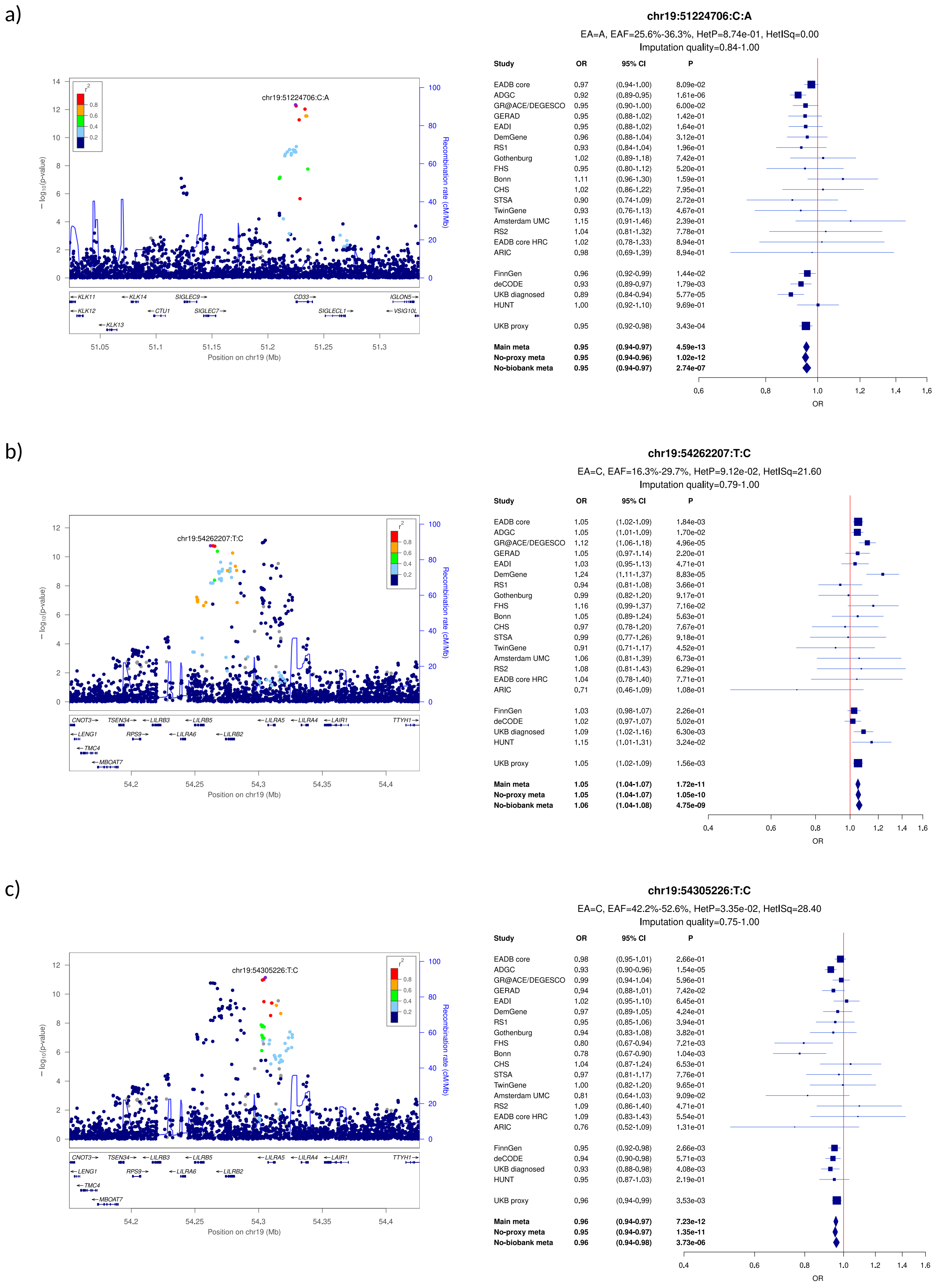

**Supplementary Figure 53:** LocusZoom and forest plots of the main analysis results for (a) *LILRB1/LILRB4* (lead signal) (b) *LILRB1/LILRB4* (secondary signal) and (c) *RBCK1* (secondary signal). The forest plots also show the results of the no-proxy and no-biobank meta-analyses (where the UKBB diagnosed rather than the UKBB proxy results were used). In the forest plots, data are presented as odds-ratio with 95% confidence interval. P values are two-sided raw P values derived from a fixed-effect meta-analysis. OR: odds ratio, CI: confidence interval, EA: effect allele, EAF: effect allele frequency range across all studies, HetP: heterogeneity P value, HetISq: heterogeneity statistic.

a)

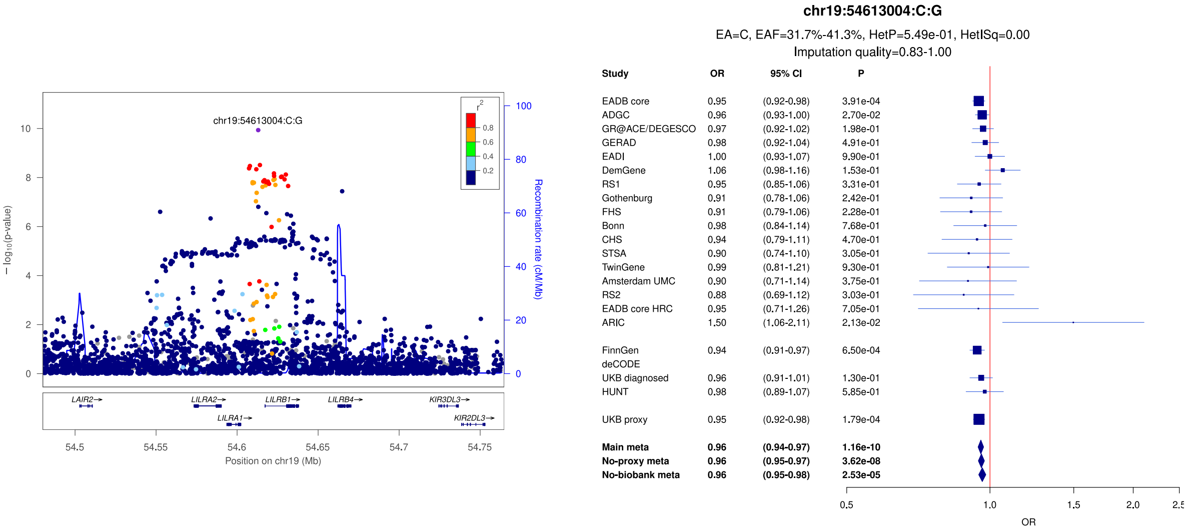


b)

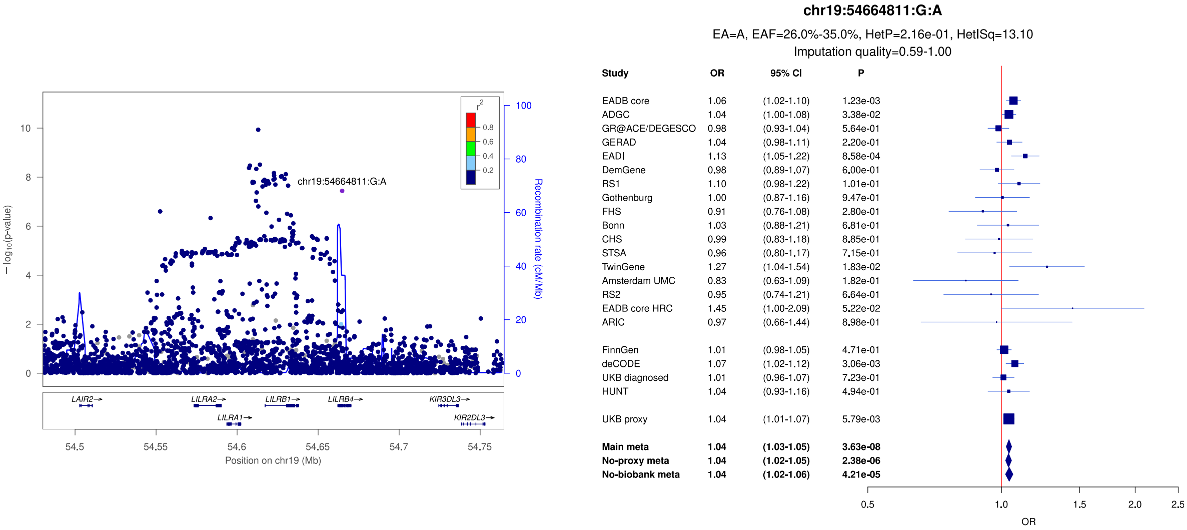


c)

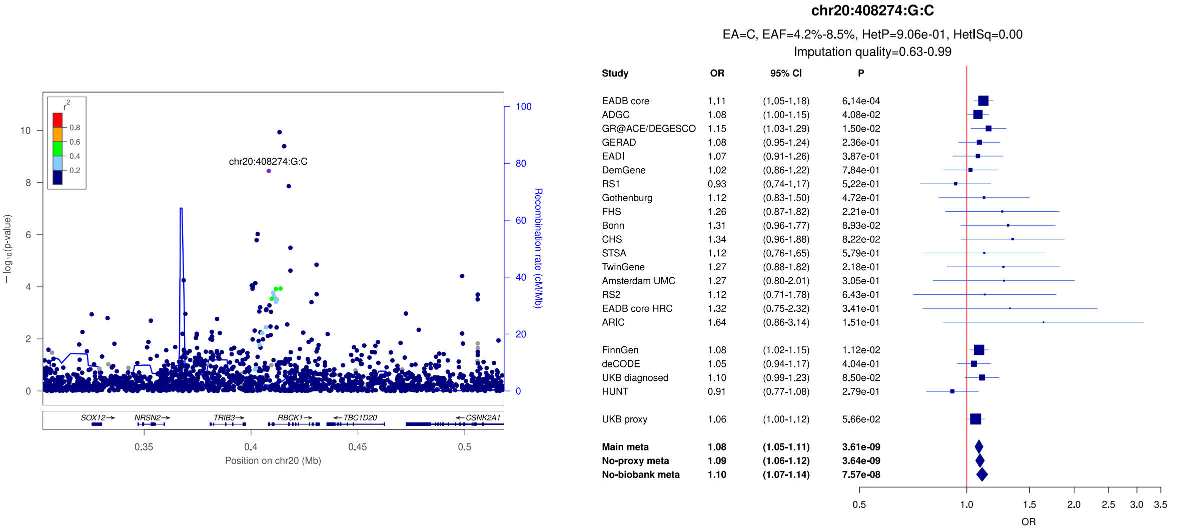

**Supplementary Figure 54:** LocusZoom and forest plots of the main analysis results for (a) *RBCK1* (lead signal) (b) *SRC* and (c) *CASS4*. The forest plots also show the results of the no-proxy and no-biobank meta-analyses (where the UKBB diagnosed rather than the UKBB proxy results were used). In the forest plots, data are presented as odds-ratio with 95% confidence interval. P values are two-sided raw P values derived from a fixed-effect meta-analysis. OR: odds ratio, CI: confidence interval, EA: effect allele, EAF: effect allele frequency range across all studies, HetP: heterogeneity P value, HetISq: heterogeneity statistic.

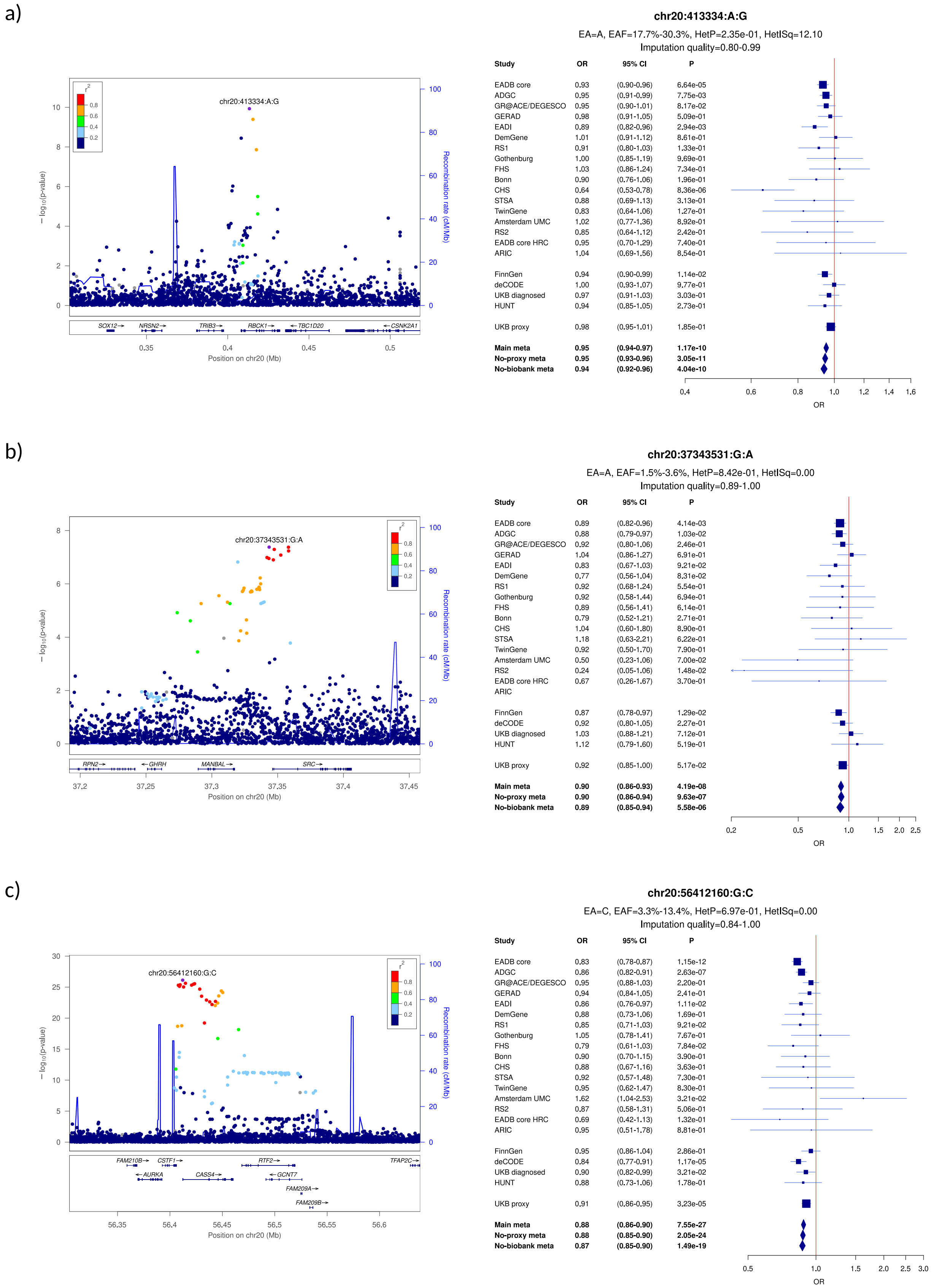

**Supplementary Figure 55:** LocusZoom and forest plots of the main analysis results for (a) *APP* (secondary signal) (b) *APP* (lead signal) and (c) *ADAMTS1*. The forest plots also show the results of the no-proxy and no-biobank meta-analyses (where the UKBB diagnosed rather than the UKBB proxy results were used). In the forest plots, data are presented as odds-ratio with 95% confidence interval. P values are two-sided raw P values derived from a fixed-effect meta-analysis. OR: odds ratio, CI: confidence interval, EA: effect allele, EAF: effect allele frequency range across all studies, HetP: heterogeneity P value, HetISq: heterogeneity statistic.

a)

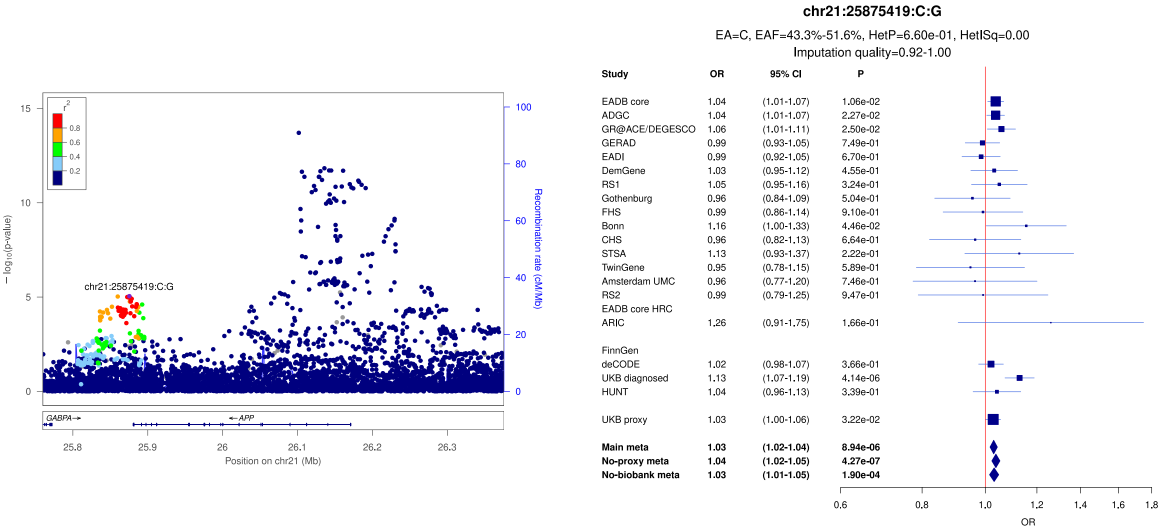


b)

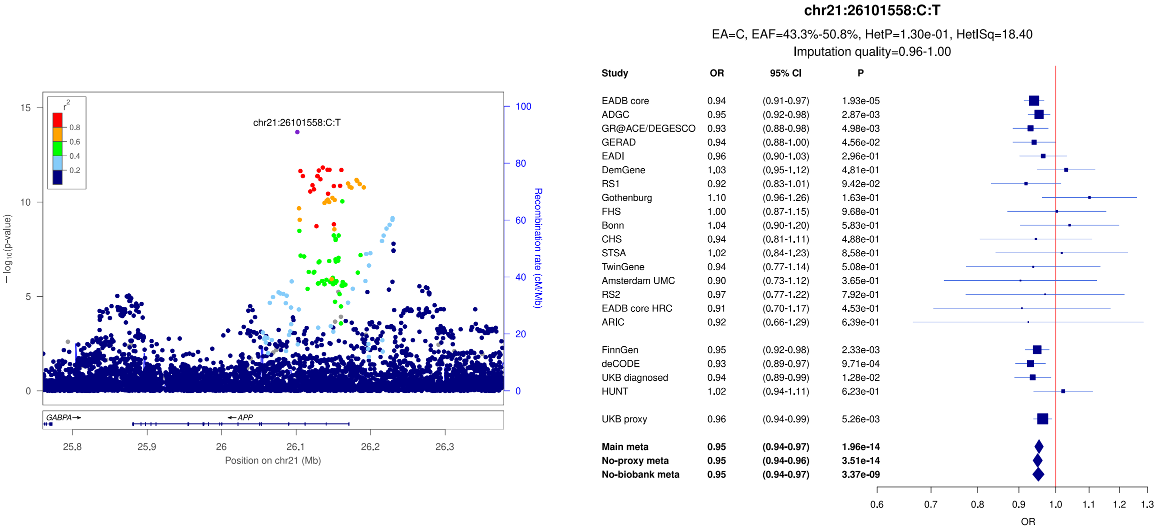


c)

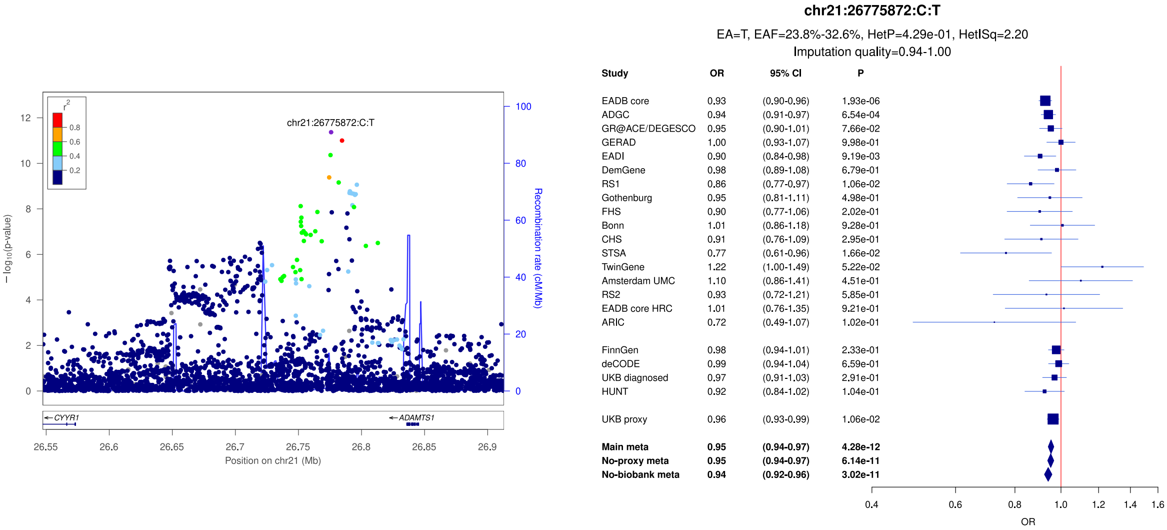

**Supplementary Figure 56:** Forest plots of the main analysis results for index variants of loci from the EADB and PGC2 studies that are not genome-wide significant in the EADB-IGAP-PGC meta-analysis: a) *HAVCR2*; b) *FOXF1*; c) *NTN5*; d) *SLC2A4RG/LIME1*. Index variants are described in Supplementary Table 5. Data are presented as odds-ratio with 95% confidence interval. P values are two-sided raw P values derived from a fixed-effect meta-analysis. OR: odds ratio, CI: confidence interval, EA: effect allele, EAF: effect allele frequency range across all studies, HetP: heterogeneity P value, HetISq: heterogeneity statistic.

a)

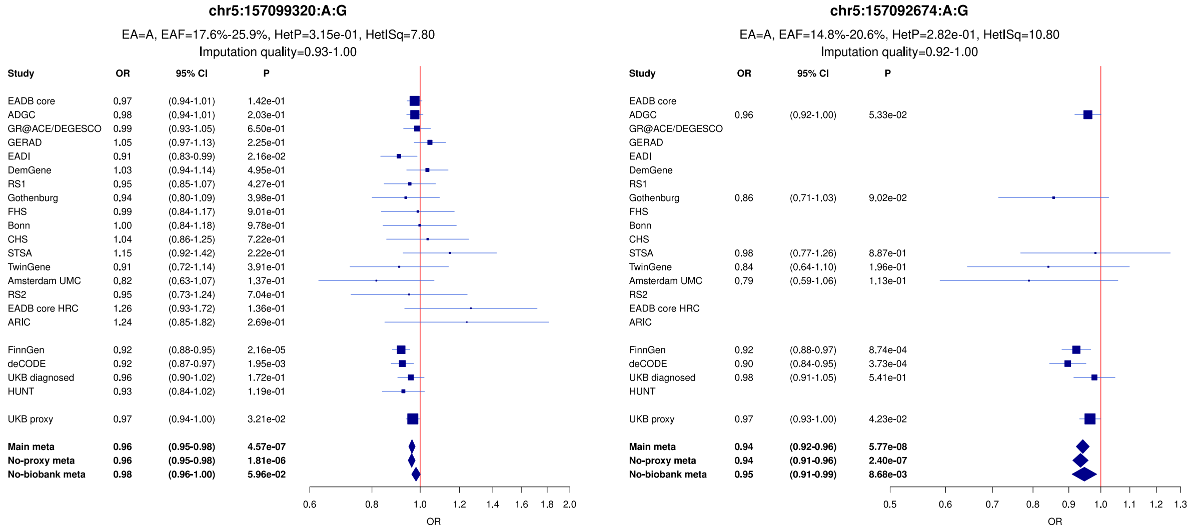


b)

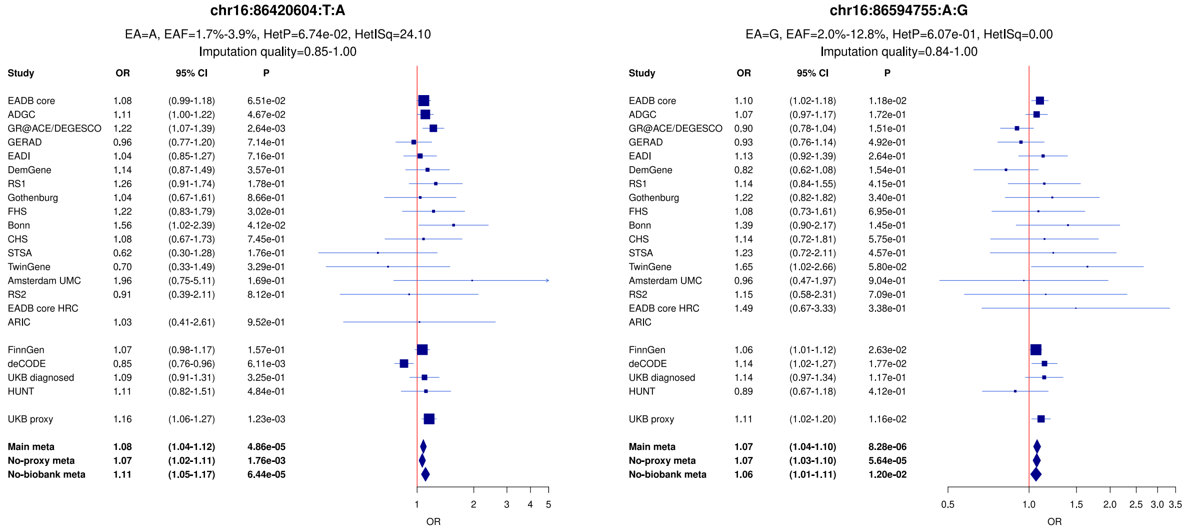


c)

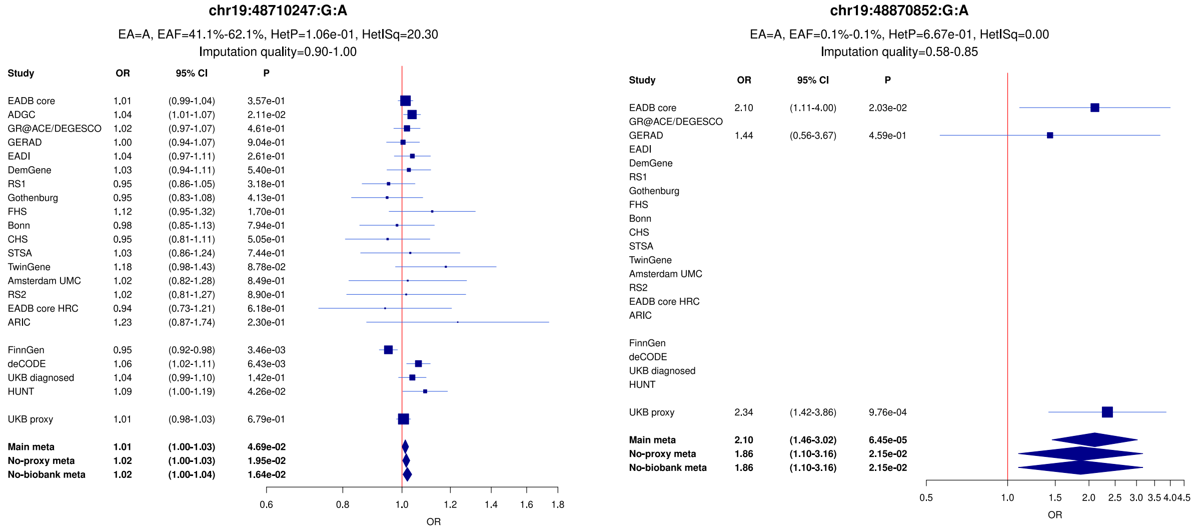

d)

**chr20:63743088:T:C**

EA=T, EAF=18.6%-24.1%, HetP=4.46e-01, HetISq=1.40

Imputation quality=0.64-1.00

| Study | OR | 95% CI | P |
|---|---|---|---|
| EADB core | 0.94 | (0.91-0.98) | 1.17e-03 |
| ADGC | 0.94 | (0.90-0.98) | 3.65e-03 |
| GR@ACE/DEGESCO | 1.00 | (0.94-1.07) | 9.49e-01 |
| GERAD | 0.94 | (0.87-1.02) | 1.39e-01 |
| EADI | 0.95 | (0.87-1.04) | 2.44e-01 |
| DemGene | 0.93 | (0.83-1.04) | 2.23e-01 |
| RS1 | 1.01 | (0.89-1.15) | 9.00e-01 |
| Gothenburg | 0.94 | (0.78-1.13) | 5.14e-01 |
| FHS | 0.95 | (0.80-1.14) | 5.99e-01 |
| Bonn | 0.94 | (0.79-1.13) | 5.27e-01 |
| CHS | 0.87 | (0.70-1.08) | 2.17e-01 |
| STSA | 0.99 | (0.77-1.27) | 9.14e-01 |
| TwinGene | 1.05 | (0.84-1.33) | 6.59e-01 |
| Amsterdam UMC | 1.27 | (0.94-1.72) | 1.14e-01 |
| RS2 | 1.41 | (1.07-1.85) | 1.80e-02 |
| EADB core HRC | 1.03 | (0.74-1.44) | 8.42e-01 |
| ARIC | 1.23 | (0.82-1.83) | 3.17e-01 |
| FinnGen | 0.97 | (0.93-1.01) | 1.92e-01 |
| deCODE | 0.99 | (0.94-1.04) | 6.67e-01 |
| UKB diagnosed | 0.99 | (0.93-1.06) | 7.73e-01 |
| HUNT | 1.05 | (0.93-1.17) | 4.39e-01 |
| UKB proxy | 0.97 | (0.94-1.00) | 9.62e-02 |
| **Main meta** | **0.97** | **(0.95-0.98)** | **1.36e-05** |
| **No-proxy meta** | **0.97** | **(0.95-0.98)** | **7.07e-05** |
| **No-biobank meta** | **0.96** | **(0.94-0.98)** | **3.01e-05** |

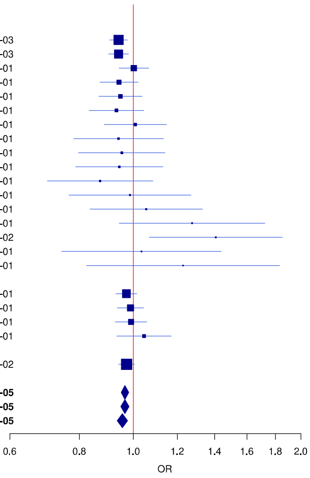

**Supplementary Figure 57:** LocusZoom and forest plots of the no-proxy analysis results for (a) *MFSD10* (b) *ILRUN* and (c) *AKR1D1*. The forest plots also show the results of the main and no-biobank meta-analyses. The UKBB diagnosed rather than the UKBB proxy results were used in the no-proxy and no-biobank meta-analyses. In the forest plots, data are presented as odds-ratio with 95% confidence interval. P values are two-sided raw P values derived from a fixed-effect meta-analysis. OR: odds ratio, CI: confidence interval, EA: effect allele, EAF: effect allele frequency range across all studies, HetP: heterogeneity P value, HetISq: heterogeneity statistic.

a)

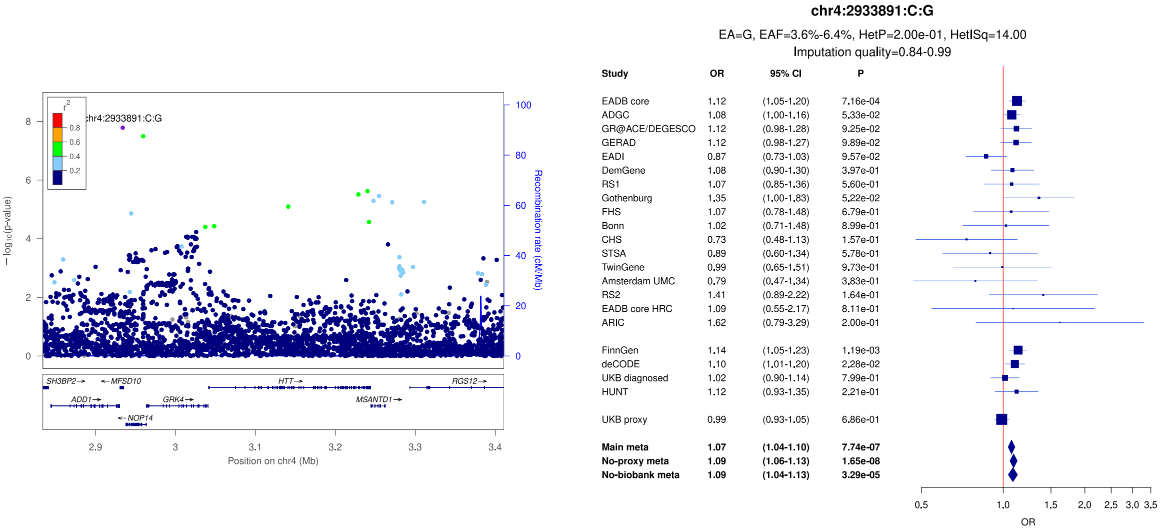


b)

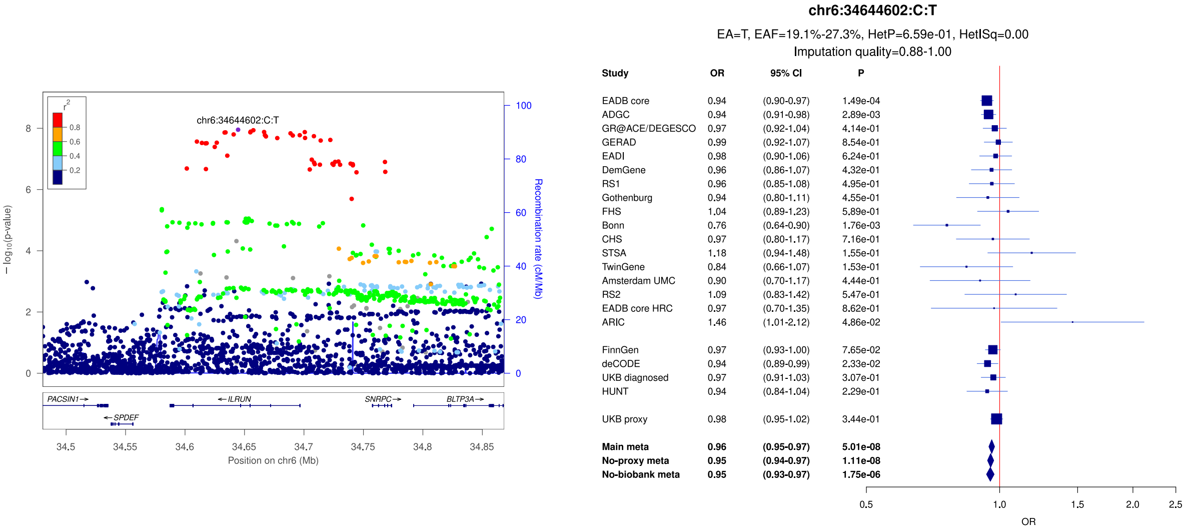


c)

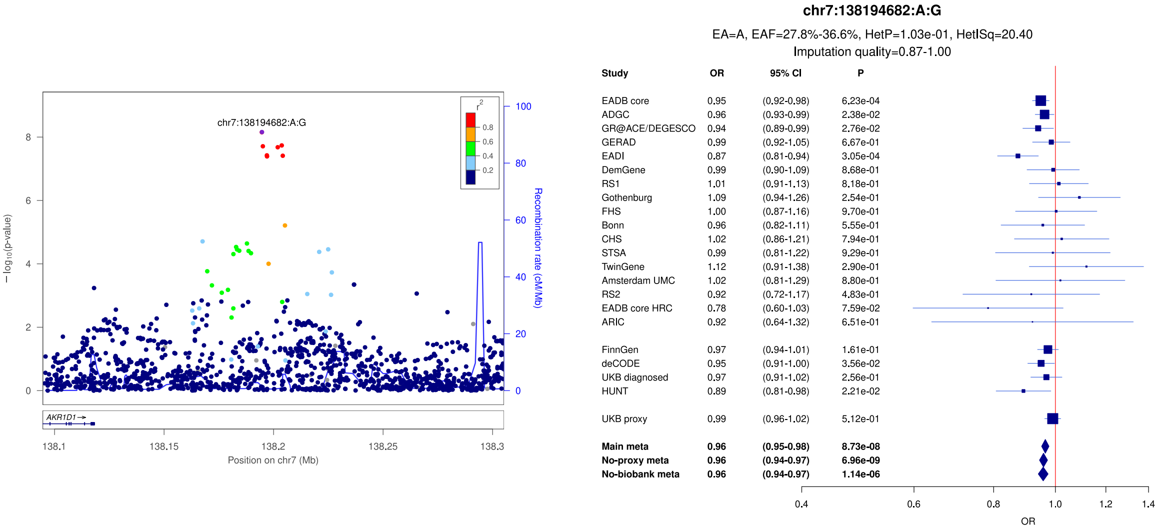

**Supplementary Figure 58:** LocusZoom and forest plots of the no-proxy analysis results for (a) *ANO5* (b) R*IC8B* and (c) *GTF2H3*. The forest plots also show the results of the main and no-biobank meta-analyses. The UKBB diagnosed rather than the UKBB proxy results were used in the no-proxy and no-biobank meta-analyses. In the forest plots, data are presented as odds-ratio with 95% confidence interval. P values are two-sided raw P values derived from a fixed-effect meta-analysis. OR: odds ratio, CI: confidence interval, EA: effect allele, EAF: effect allele frequency range across all studies, HetP: heterogeneity P value, HetISq: heterogeneity statistic.

a)

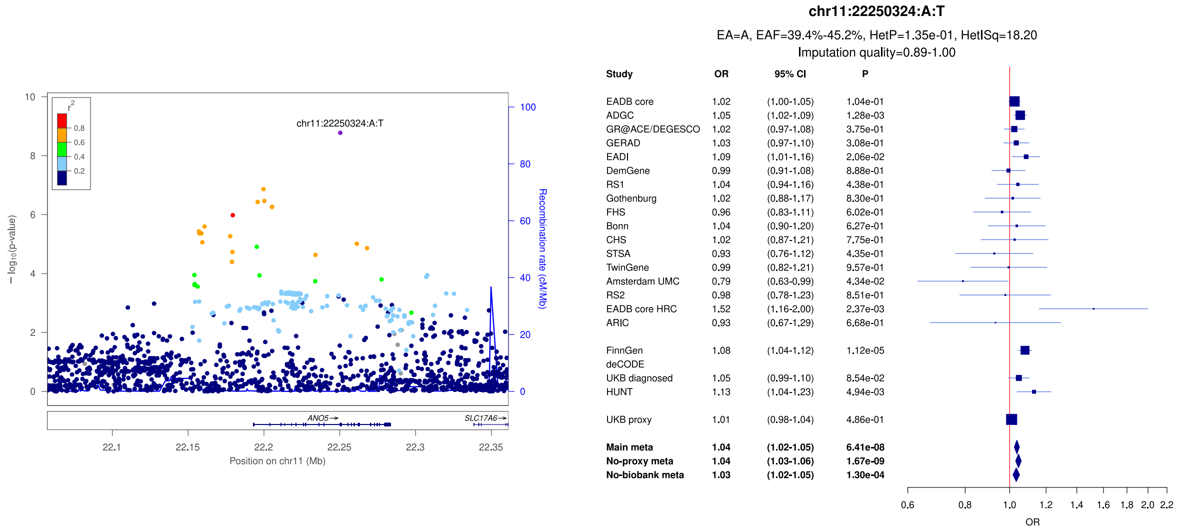


b)

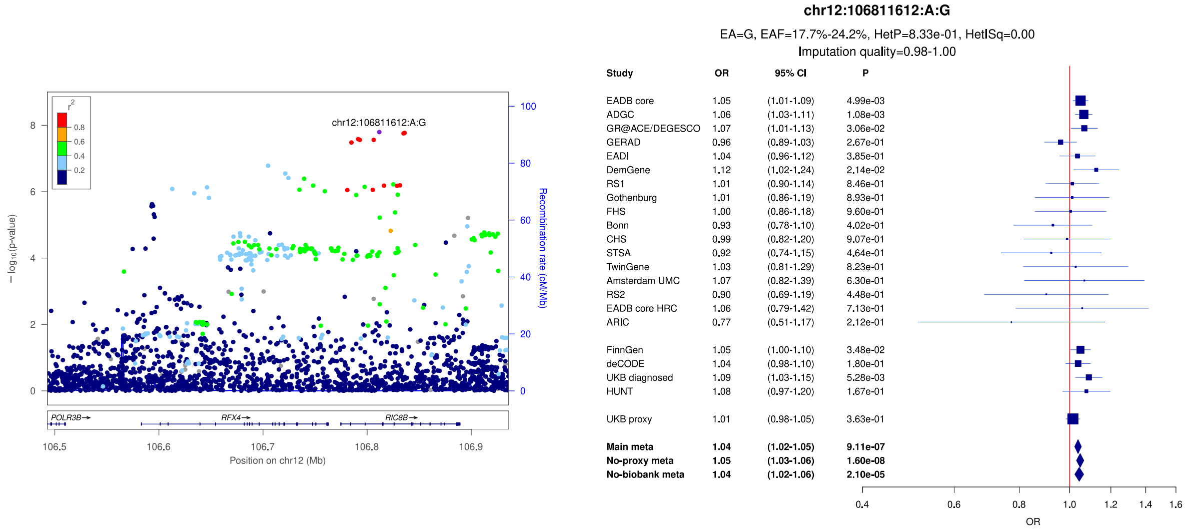


c)

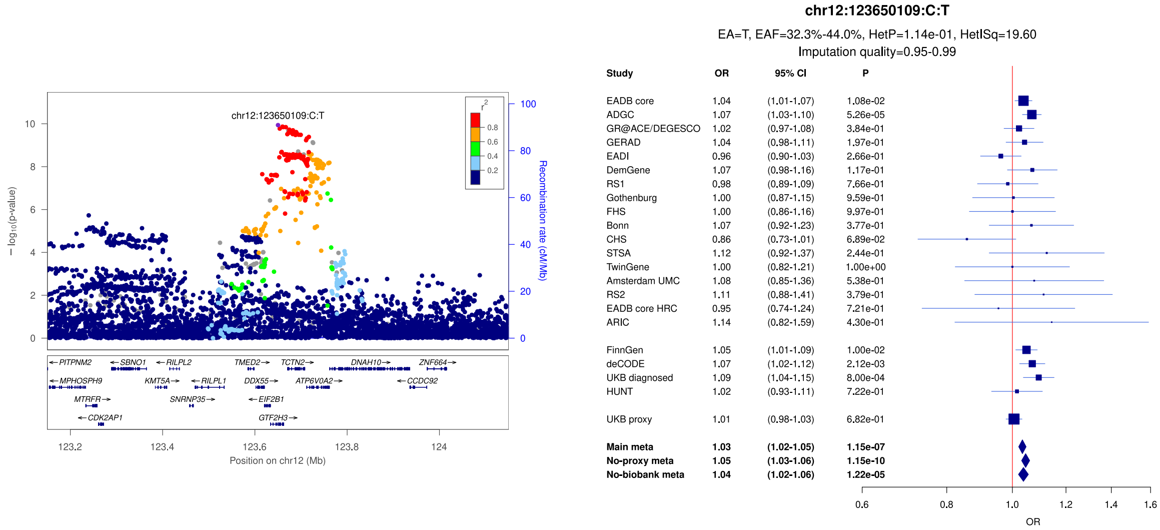

**Supplementary Figure 59:** LocusZoom and forest plots of the no-proxy analysis results for (a) *C16orf95* (b) *SNAI1* and (c) *PSMG1*. The forest plots also show the results of the main and no-biobank meta-analyses. The UKBB diagnosed rather than the UKBB proxy results were used in the no-proxy and no-biobank meta-analyses. In the forest plots, data are presented as odds-ratio with 95% confidence interval. P values are two-sided raw P values derived from a fixed-effect meta-analysis. OR: odds ratio, CI: confidence interval, EA: effect allele, EAF: effect allele frequency range across all studies, HetP: heterogeneity P value, HetISq: heterogeneity statistic.

a)

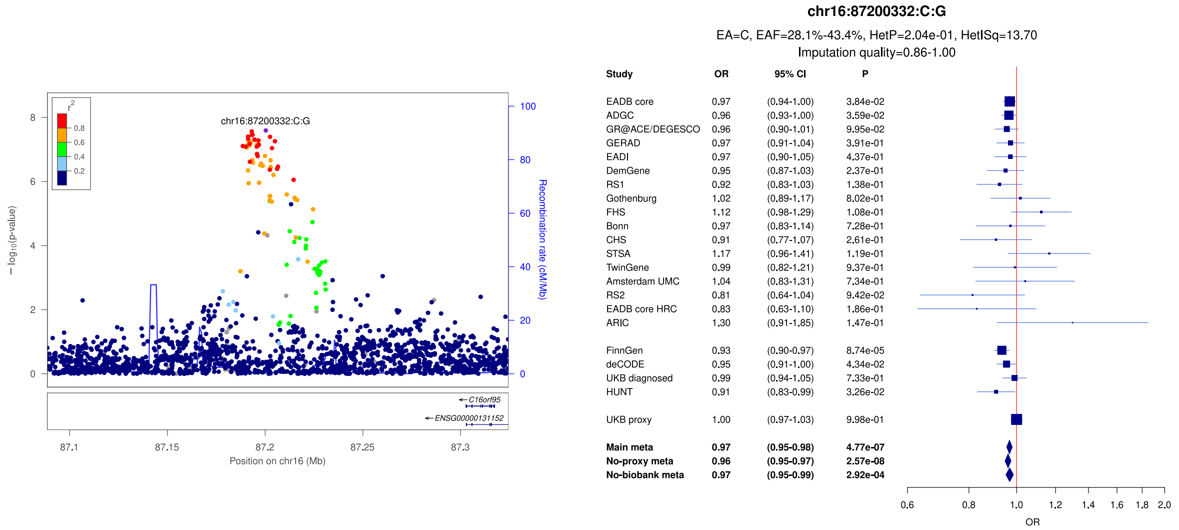


b)

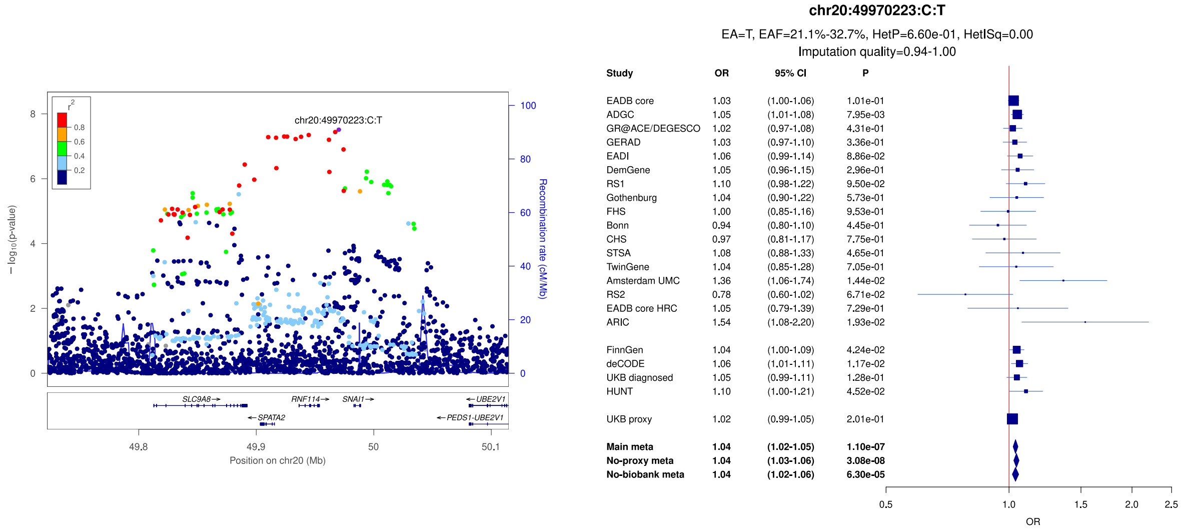


c)

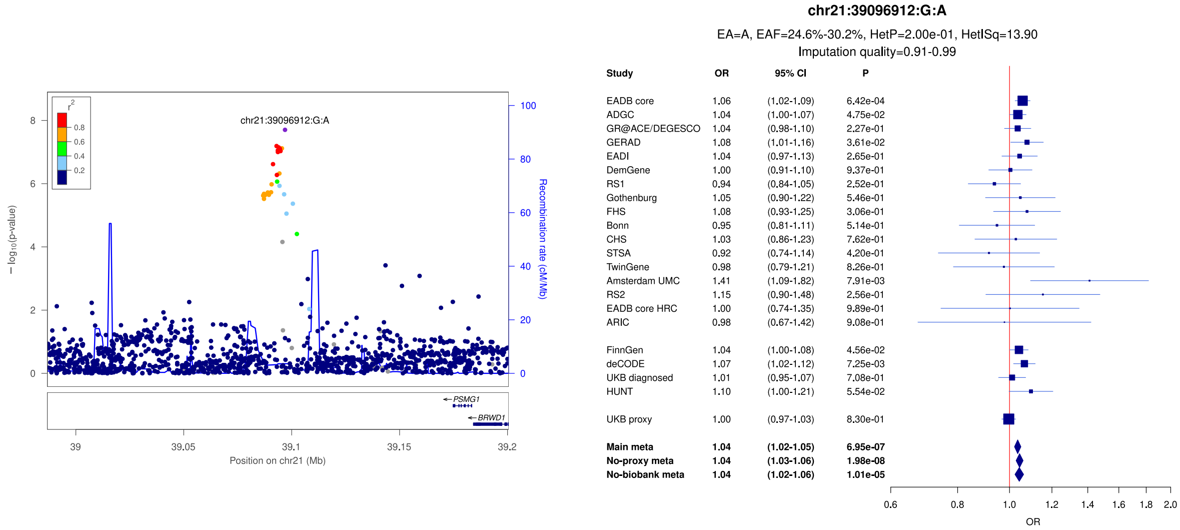

**Supplementary Figure 60:** LocusZoom and forest plots of the no-biobank analysis results for (a) *C1S* (b) *MEFV* and (c) *ZCCHC2*. The forest plots also show the results of the main and no-proxy meta-analyses. The UKBB diagnosed rather than the UKBB proxy results were used in the no-proxy and no-biobank meta-analyses. In the forest plots, data are presented as odds-ratio with 95% confidence interval. P values are two-sided raw P values derived from a fixed-effect meta-analysis. OR: odds ratio, CI: confidence interval, EA: effect allele, EAF: effect allele frequency range across all studies, HetP: heterogeneity P value, HetISq: heterogeneity statistic.

a)

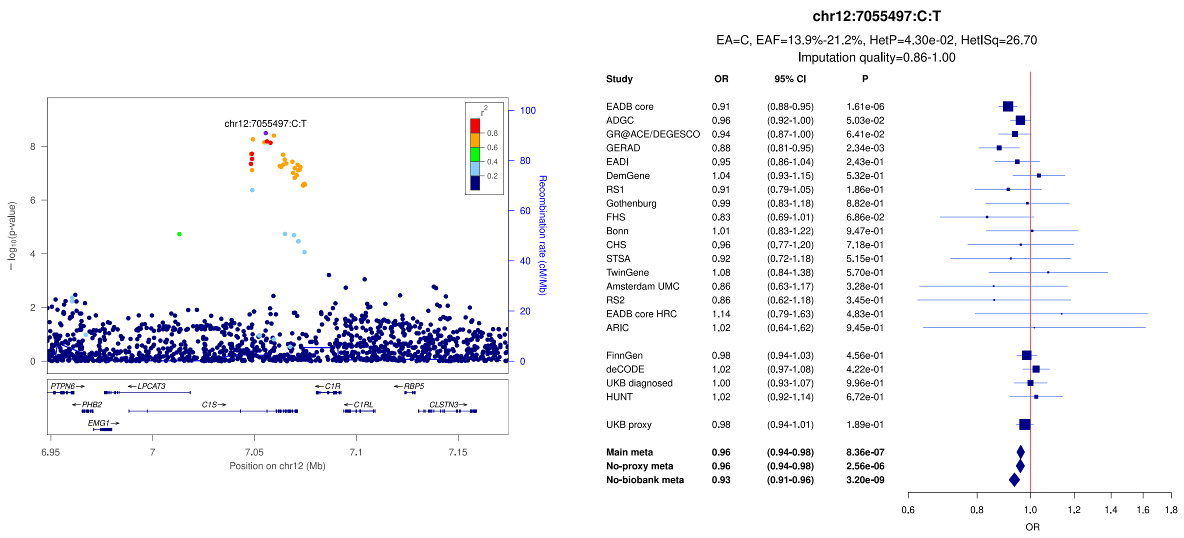


b)

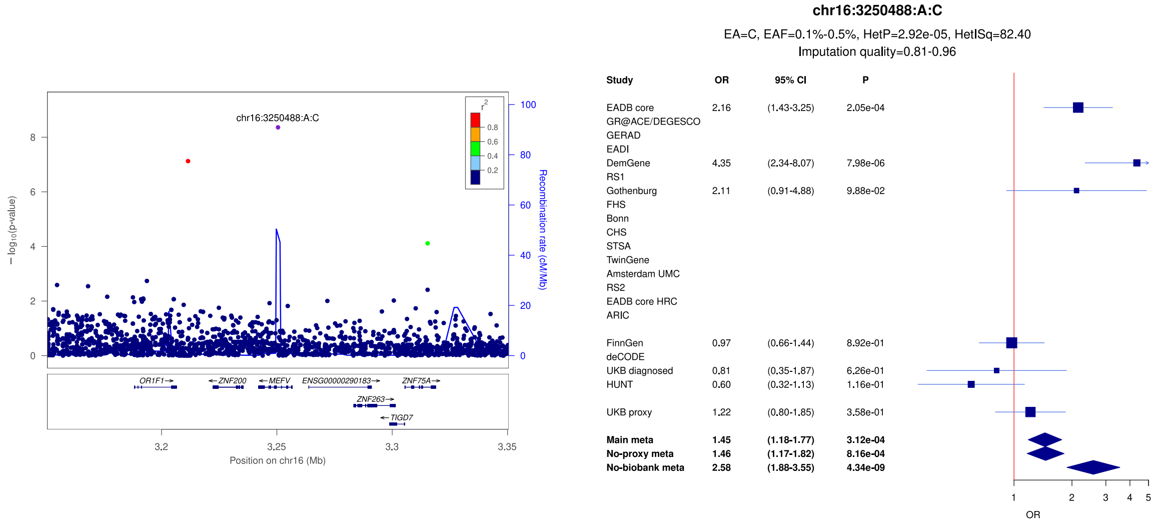


c)

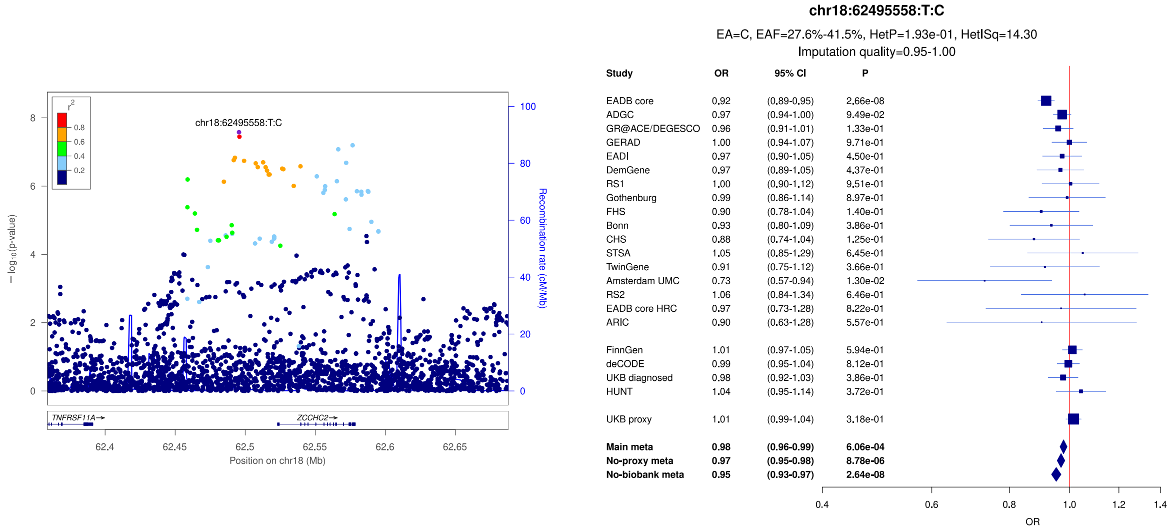

**Supplementary Figure 61:** LocusZoom and forest plots of the no-biobank analysis results for *RIPOR3*. The forest plots also show the results of the main and no-proxy meta-analyses. The UKBB diagnosed rather than the UKBB proxy results were used in the no-proxy and no-biobank meta-analyses. In the forest plots, data are presented as odds-ratio with 95% confidence interval. P values are two-sided raw P values derived from a fixed-effect meta-analysis. OR: odds ratio, CI: confidence interval, EA: effect allele, EAF: effect allele frequency range across all studies, HetP: heterogeneity P value, HetISq: heterogeneity statistic.

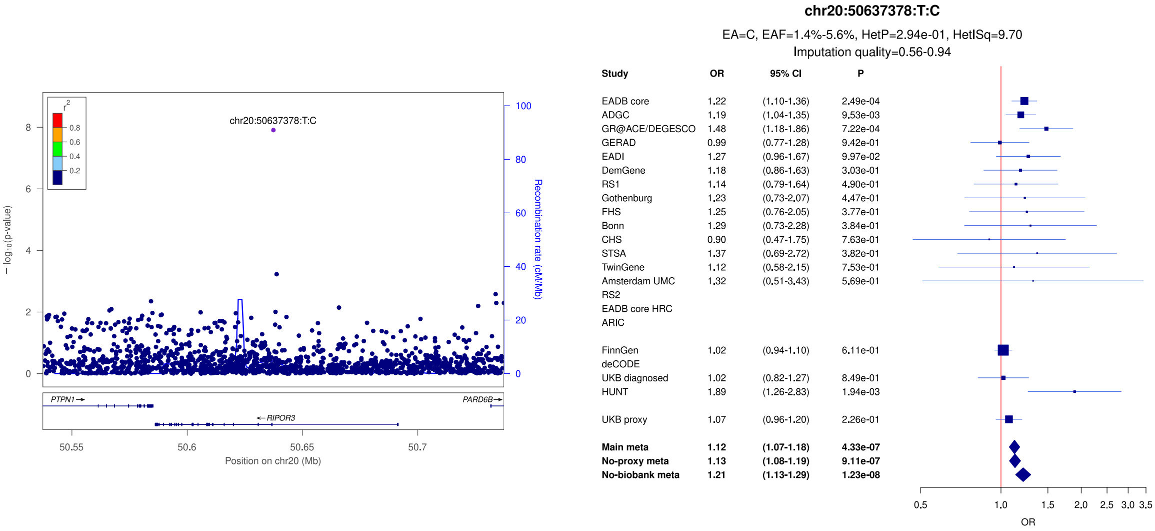

**Supplementary Figure 62:** The step 1 and step 3 results of the cell type association analysis highlight microglia as a cell type where genes enriched with association signal with ADD are overexpressed. a) The step 1 results show which cell types have significant overexpression of genes enriched in ADD association signal after Bonferroni correction for 136 tested cell types. The Y axis represents the -log10(P-value), where P-value is raw and one-sided. The x axis represents the cell type label. The colour of the bars represents the dataset. The horizontal dashed line represents the Bonferroni correction threshold (0.05/136). b) The step 3 results show that the associations across datasets are largely driven by a shared set of genes. The axes text colours represent the dataset and match the legend from a). The colour of the heatmap (independence) represents the proportional significance after the cell type on the y axis is conditioned on the cell type on the x axis (-log10(P_y_conditioned_on_x)/-log10(P_y)). Larger values represent higher independence between cell type associations.

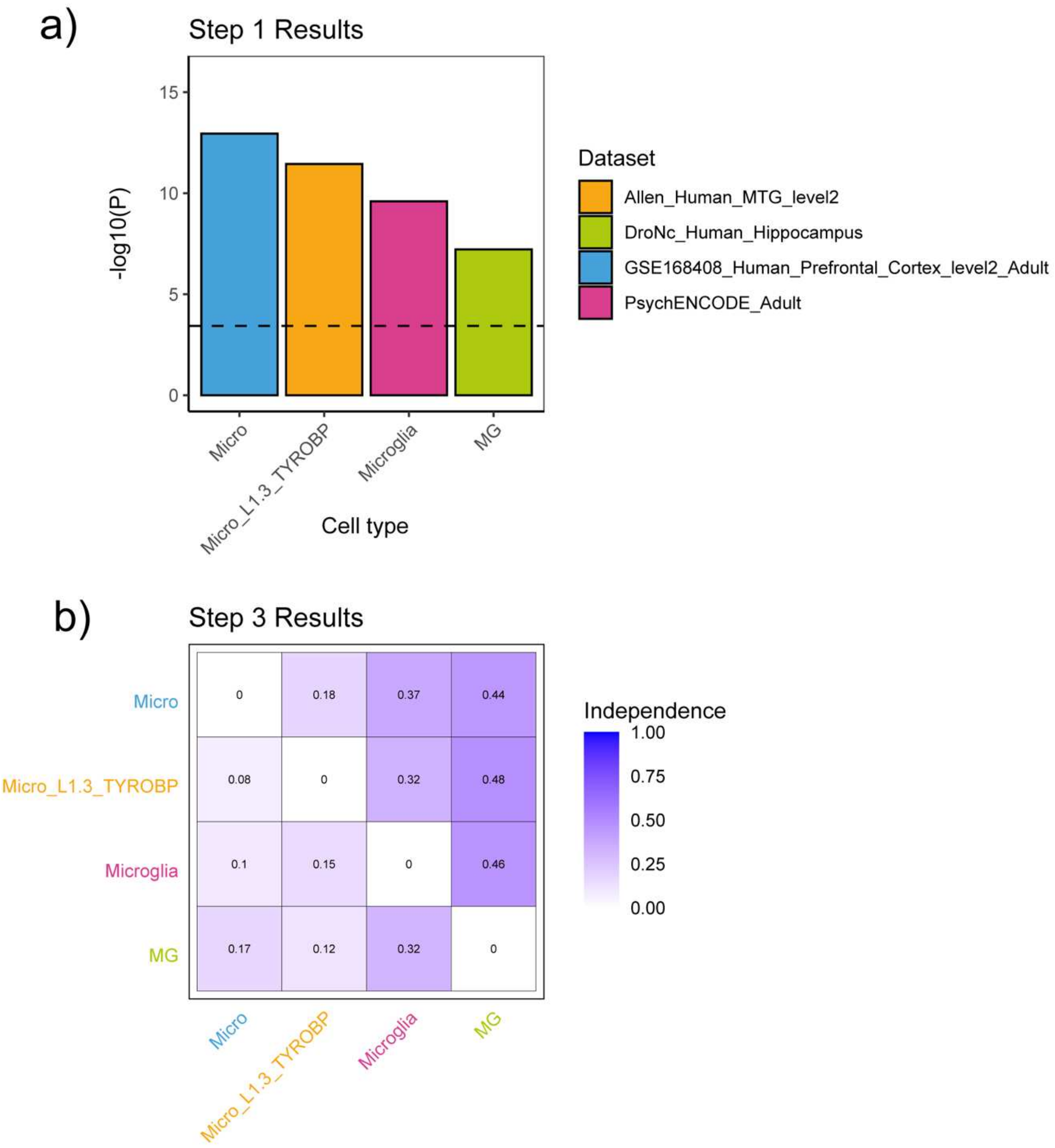

**Supplementary Figure 63:** The primary analysis results from step 1 of the cell type association analysis are robust to all five sensitivity analyses. The Y axis represents the -log10(P-value) of the step 1 cell type associations, where P-value is raw and one-sided, and the x-axis represents the different cell types. The colours represent the analysis models: Primary, exclusion of rare variants (CommonOnly), exclusion of variants in the *APOE* region (NoAPOE), larger variant to gene mapping windows (LargerWindow), exclusion of large biobanks (NoBiobank) or proxy datasets (NoProxy). The dashed line represents the Bonferroni corrected threshold (0.05/136).

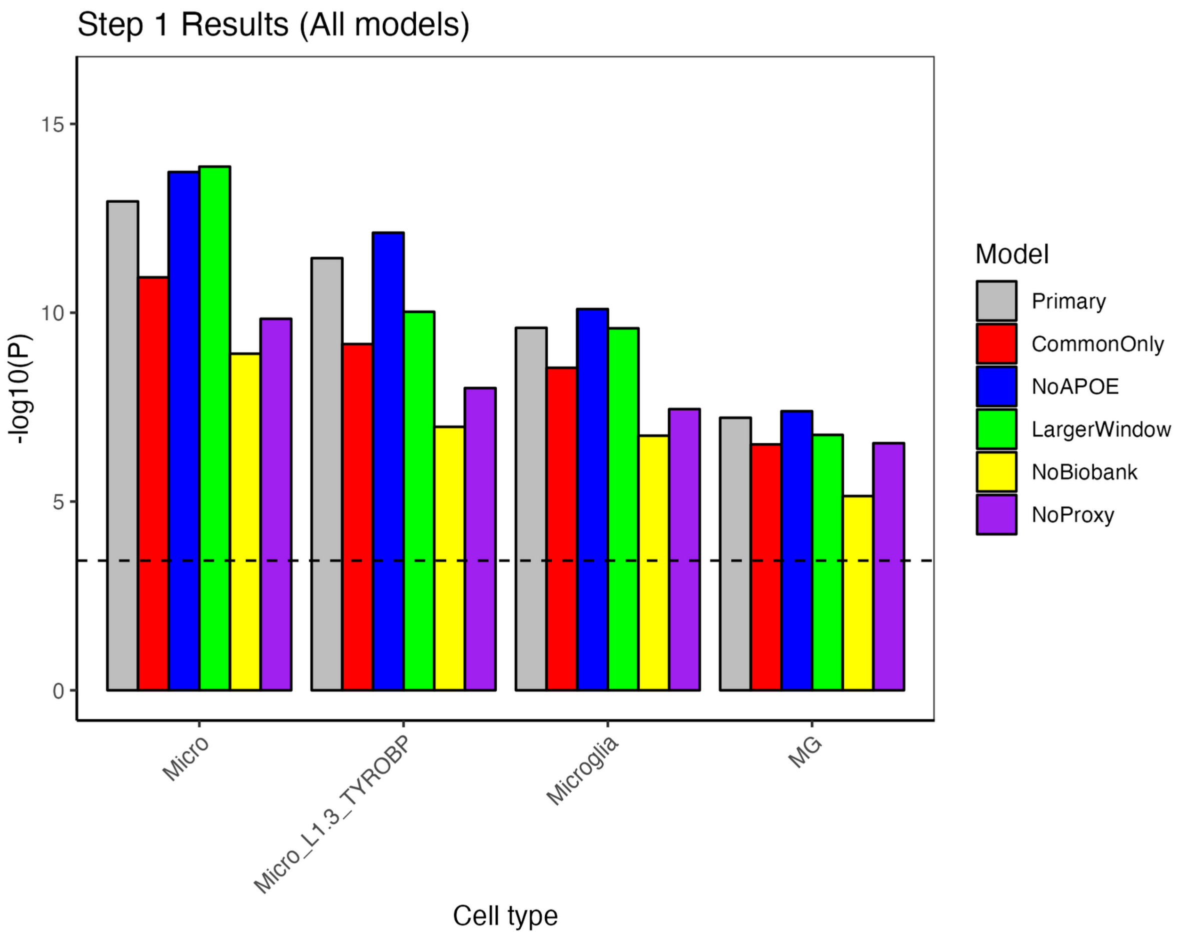

**Supplementary Figure 64:** The step 3 results of the cell type association analysis show that the associations across datasets are largely driven by a shared set of genes and this is not affected by the sensitivity analyses. The axes text colours represent the dataset and match the legend from Supplementary Figure 62 a). The colour of the heatmap (independence) represents the proportional significance after the cell type on the y axis is conditioned on the cell type on the x axis (-log10(P_y_conditioned_on_x)/-log10(P_y)). Larger values represent higher independence between cell type associations. The plot titles represent each analysis model: Primary, exclusion of rare variants (CommonOnly), exclusion of variants in the *APOE* region (NoAPOE), larger variant to gene mapping windows (LargerWindow), exclusion of large biobanks (NoBiobank) or proxy datasets (NoProxy).

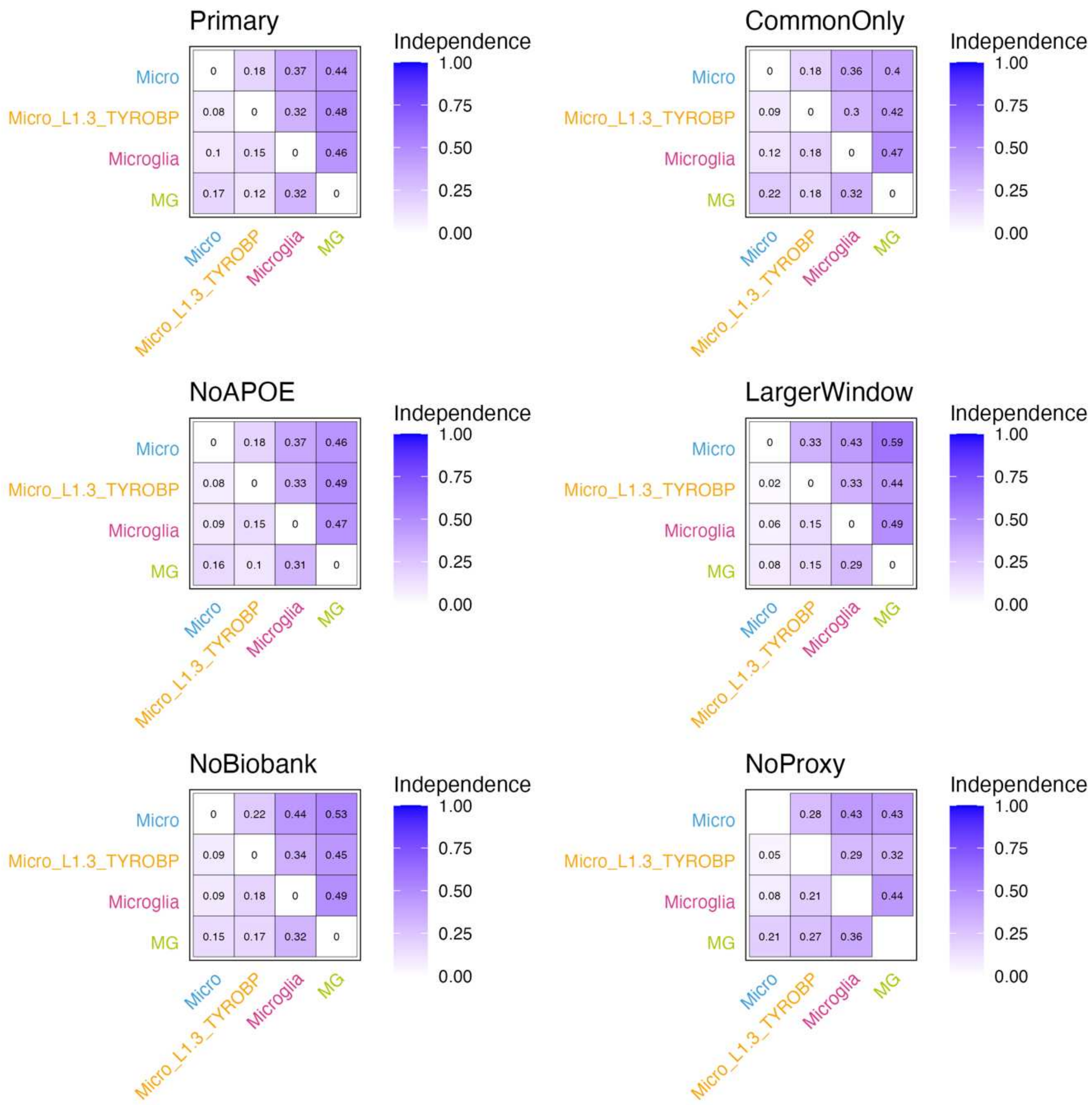

**Supplementary Figure 65:** Association of ADRD polygenic scores (PGS) with the neuropathology endophenotypes (NPE). a) After minimal adjustment on age at death, sex, the number of APOE ε4 and ε2 alleles, principal components and centers. b) After additional adjustment on AD diagnosis. OR: odds-ratio; NFT: neurofibrillary tangles; LATE-NC: limbic-predominant age-related TDP-43 encephalopathy neuropathological change. Bars indicate 95% confidence intervals

a)

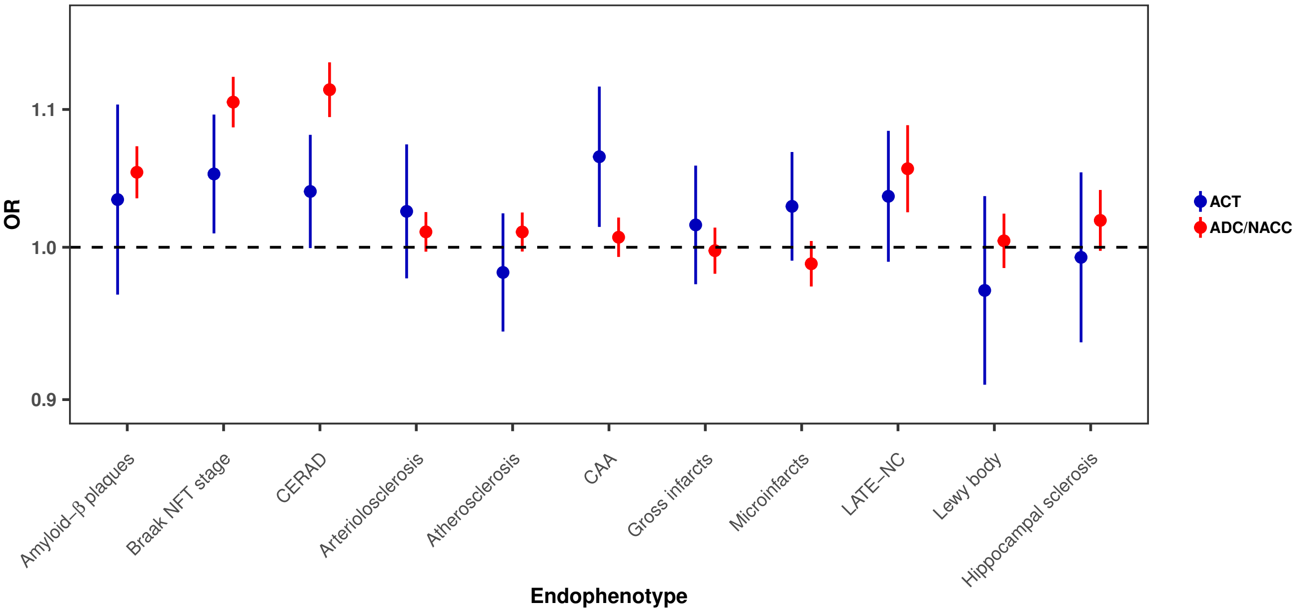


b)

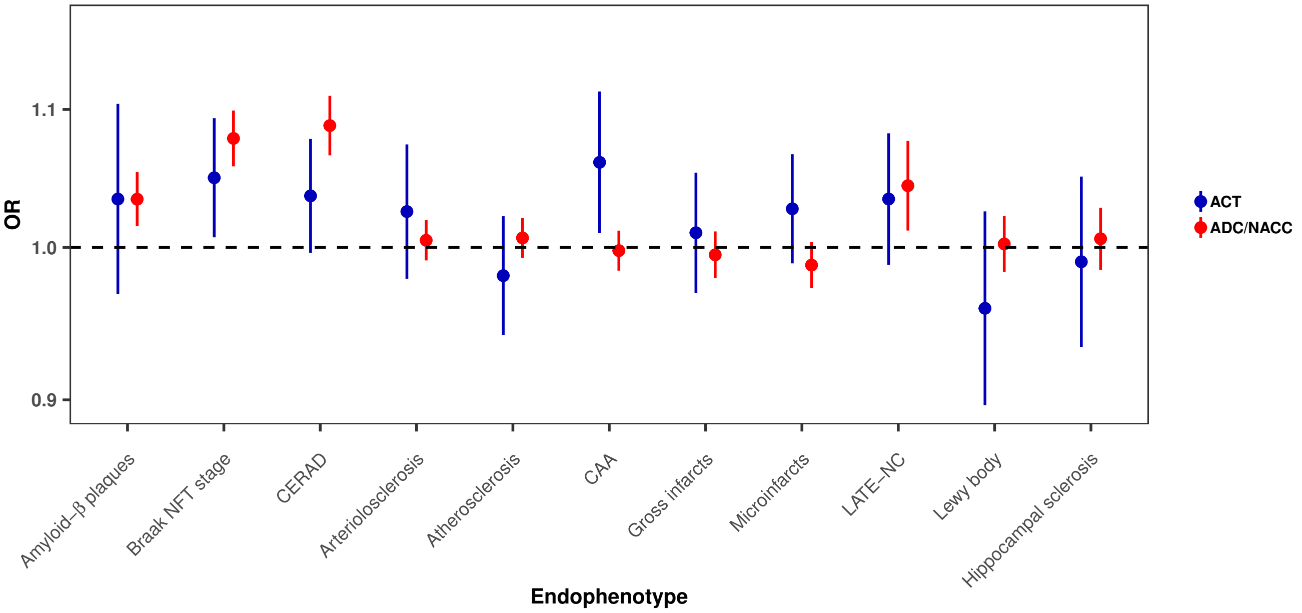

**Supplementary Figure 66:** Association of the main, no-proxy and no-biobank polygenic scores (PGS) with the neuropathology endophenotypes (NPE) in ADC/NACC and ACT, after minimal adjustment on age at death, sex, the number of APOE ε4 and ε2 alleles, principal components and centers. OR: odds-ratio; NFT: neurofibrillary tangles; LATE-NC: limbic-predominant age-related TDP-43 encephalopathy neuropathological change. Bars indicate 95% confidence intervals

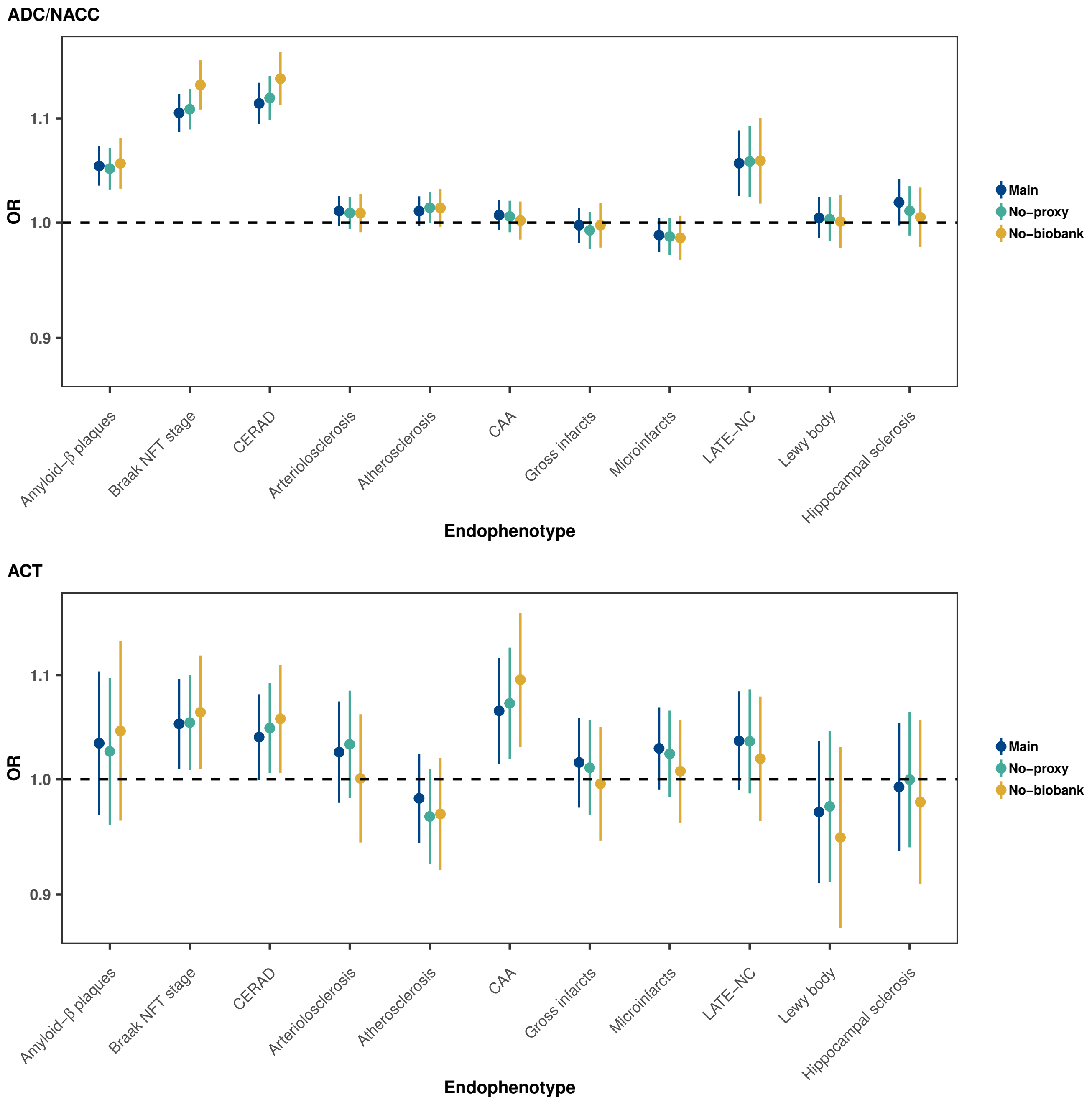

**Supplementary Figure 67:** Association of the main, no-proxy and no-biobank polygenic scores (PGS) quintiles with Braak stage and CERAD in ADC/NACC (panel a) and ACT (panel b) after minimal adjustment on age at death, sex, the number of APOE ε4 and ε2 alleles, principal components and centers. OR: odds-ratio; NFT: neurofibrillary tangles; LATE-NC: limbic-predominant age-related TDP-43 encephalopathy neuropathological change. Bars indicate 95% confidence intervals

a)

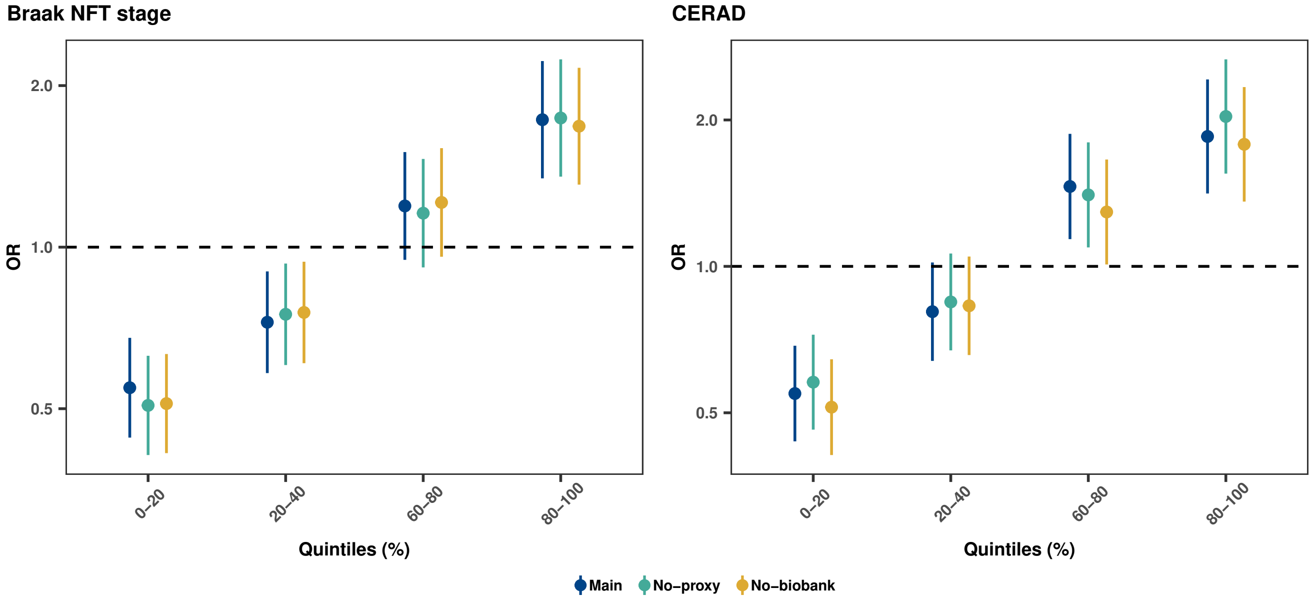


b)

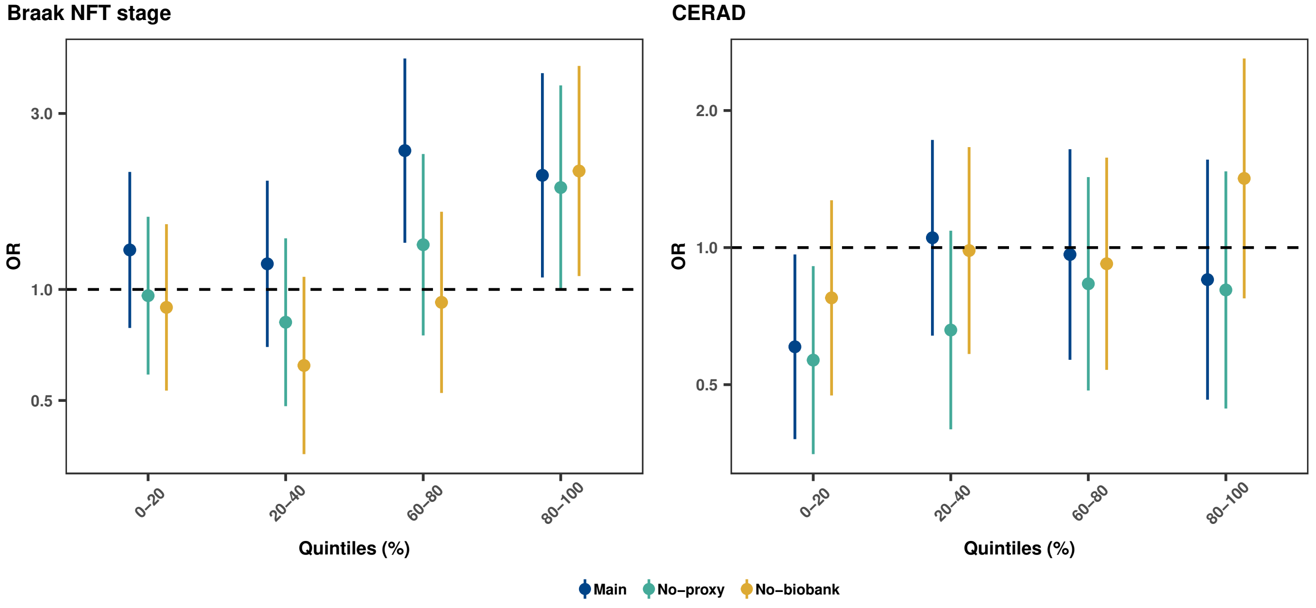
